# Effects of saffron and crocin on glycemic control: an exploratory systematic review and meta-analysis of randomized trials

**DOI:** 10.64898/2026.10.30.26366889

**Authors:** Juno Kim, Michele Schaffer, Grace Kim, Bryan Anh Nguyen, Prisha Batra, Alexander Tai Nguyen, Pranav Potluri, Kitai Kim

**Affiliations:** Johns Hopkins University, Baltimore, Maryland, USA; Virginia University of Integrative Medicine, Virginia, USA; Santa Monica College, Santa Monica, California, USA; University of California, Los Angeles, California, USA; University of Richmond, Richmond, Virginia, USA

**Author notes:** Corresponding author: Kitai Kim.

**Keywords:** saffron, *Crocus sativus*, crocin, glycated hemoglobin, fasting glucose, insulin resistance, meta-analysis

## Abstract

**Background:** Saffron and its constituent crocin have been investigated as adjuncts for metabolic dysfunction. Their effects on longer-term glycemic exposure and fasting indices may differ.

**Objectives:** To assess the effects of saffron and its constituents on glycated hemoglobin (HbA1c), fasting glucose, and insulin resistance.

**Search methods:** We searched PubMed Central on June 11, 2025, and PubMed on October 5, 2025.

**Selection criteria:** Randomized controlled trials in humans comparing saffron or its constituents with a control for at least four weeks and reporting a relevant glycemic outcome were eligible. Comparisons were selected to isolate the effect of saffron or crocin.

**Data collection and analysis:** We synthesized continuous outcomes as mean differences (MDs) with 95% confidence intervals (CIs), using inverse-variance random-effects models. HbA1c was the primary outcome. Fasting glucose and the homeostatic model assessment of insulin resistance (HOMA-IR) were secondary outcomes.

**Main results:** Twelve trials contributed 570 participants to the analyzed comparisons. Populations included adults with type 1 or type 2 diabetes, metabolic syndrome or coronary artery disease, and adolescents with obesity and prediabetes. Seven trials (391 participants) reported HbA1c: the pooled MD was *−*0.25 percentage points (95% CI *−*0.58 to 0.08; *I*^2^ = 61%). Fasting glucose was lower with saffron or crocin across 12 trials (570 participants; MD *−*16.16 mg/dL, 95% CI *−*27.02 to *−*5.31; *I*^2^ = 74%). Five trials (212 participants) contributed to HOMA-IR (MD *−*1.19, 95% CI *−*2.25 to *−*0.12; *I*^2^ = 80%). Fixed-effect sensitivity analyses gave smaller estimates across the three outcomes.

**Authors’ conclusions:** Saffron and crocin were associated with lower fasting glucose and HOMA-IR, whereas the HbA1c estimate remained compatible with both a reduction and little difference. Variation in populations, preparations, and trial sizes limits interpretation of the average effects. Larger trials with standardized preparations and adequate follow-up are needed to establish the persistence and clinical relevance of these findings.

**Funding:** None.

**Plain language summary:** Saffron and crocin, a constituent of saffron, have been studied as supplements for people with metabolic conditions. We combined randomized trials that compared these interventions with a control. Twelve trials contributed data from 570 participants in the analyzed groups. Fasting glucose was lower on average in the intervention groups. An index of insulin resistance also decreased, although results varied considerably among trials. The results for HbA1c, which reflects glucose exposure over several months, did not establish a clear reduction. The trials used different preparations and included people with different metabolic conditions. These findings do not establish that saffron or crocin can replace established diabetes treatment, or that the observed changes prevent diabetes complications.

## 1 Background

### 1.1 Description of the condition

Glycemic control can be assessed over different time scales. HbA1c reflects integrated glucose exposure, whereas fasting glucose measures glycemia at a particular time. In the A1c-Derived Average Glucose study, HbA1c measured at the end of three months was strongly associated with average glucose measured during the preceding three months.[1] These outcomes therefore provide complementary information when evaluating a glucose-lowering intervention. HOMA-IR is a model-based estimate derived from fasting glucose and insulin; it provides additional information about fasting insulin resistance but is not a direct measurement of insulin sensitivity by a glucose clamp.[2]

### 1.2 Description of the intervention

Saffron is obtained from the stigmas of *Crocus sativus* L. Preparations investigated for metabolic effects include saffron powder, aqueous or hydroalcoholic extracts, and isolated constituents. Differences in composition, constituent concentration, bioavailability, and manufacturing mean that these preparations cannot be assumed to deliver equivalent exposure or biological effects. Background treatment and baseline glycemia are also relevant to the interpretation of clinical comparisons.[3, 4] Traditional medicines can provide starting points for drug discovery. Metformin’s development has historical roots in *Galega officinalis*, also known as French lilac or goat’s rue, a traditional European herbal remedy whose guanidine-related chemistry informed the development of synthetic biguanides.[5] This precedent motivates the evaluation of traditional remedies using chemically defined constituents, reproducible preparations, and rigorous randomized comparisons.

### 1.3 How the intervention might work

Experimental evidence provides a rationale for investigating effects on glucose uptake and cellular energy regulation. Dehghan 2016 found that, in cultured rat muscle cells, saffron increased glucose uptake and GLUT4 and AMPK*α* protein expression; a parallel rat experiment reported improvements in glycemic and metabolic measures.[6] These expression changes do not by themselves demonstrate AMPK activation. Luo 2019 found that, in diabetic *db/db* mice, crocin increased hepatic AMPK phosphorylation, reduced lipogenic gene expression, and increased expression of genes involved in fatty-acid oxidation; downregulation of AMPK attenuated these effects.[7] These observations can be mapped to the human AMPK signaling pathway, including AMPK, GLUT4, acetyl-CoA carboxylase, and downstream lipid-metabolism branches.[8]

Protein abundance, phosphorylation, and loss of an experimental effect after pathway knockdown address different mechanistic questions. The cited preclinical experiments support downstream responses and pathway involvement rather than direct compound–protein binding. Extrapolation also depends on tissue, dose, preparation, and species. The relevant KEGG branches therefore provide a biological hypothesis for testing rather than proof that the same pathway mediates a human treatment effect.[6–8]

Figure 2 presents a detailed selected pathway and a summarized view of the same proposed route. Expected direction icons refer to activity, localization, metabolite pools, or physiologic measurements as labeled; they are not claims that every protein is transcriptionally up- or down-regulated. The clinical-translation region includes blood plasma glucose before hemoglobin glycation and HbA1c. PubChem two-dimensional structures, molecular weights, molecular formulas, and systematic IUPAC names for crocin, crocetin, safranal, and picrocrocin are shown in Figure 3. Crocetin is included as the aglycone of crocin for chemical context.[9–12]

**Figure 1:**
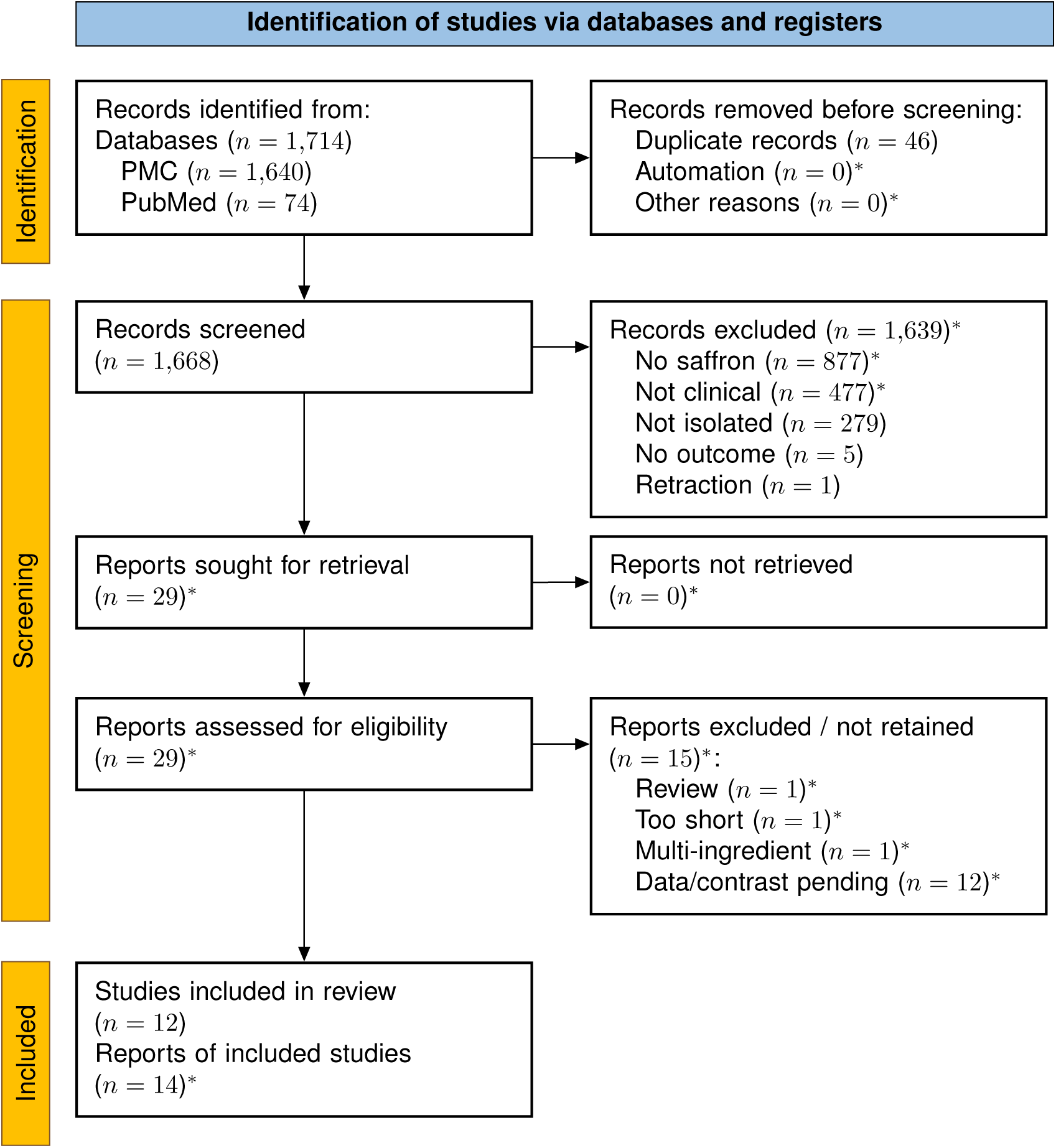
PRISMA 2020 study-selection flowchart. Adapted from the PRISMA 2020 database/register template. *^∗^*Estimated or assumed for reconstruction and subject to verification in the electronic screening audit. Five unfinished screening records are assigned likely categories; all 29 candidates are assumed retrieved and assessed. Fourteen reports are tentatively linked to 12 retained studies. The 12 data/contrast-unresolved reports are outside the current synthesis but are not established to be ineligible. Recorded decisions and inferred dispositions are preserved separately.

**Figure 2:**
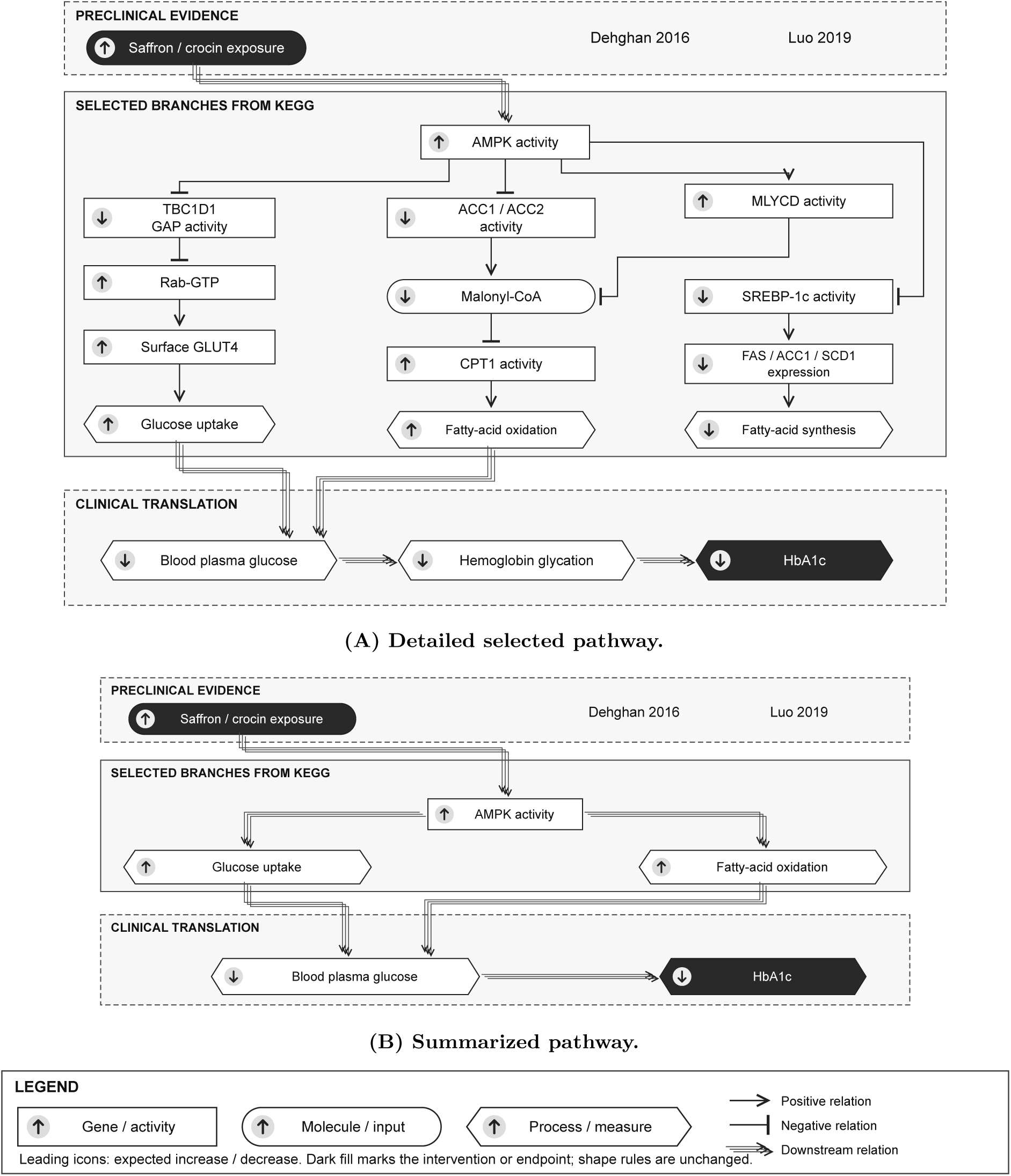
Proposed saffron and crocin pathway. The pathway is redrawn from relevant KEGG AMPK branches. A retains selected intermediates; B summarizes the same route. Rectangles denote gene products or activities, rounded nodes denote molecular inputs, and hexagons denote processes or physiologic measurements. Leading icons show expected increases or decreases in the labeled quantity. Dark fill identifies the intervention and endpoint using the same shape rules as the legend. Single arrows and bars indicate positive and negative relationships; stacked offset arrows indicate downstream effects or inference, including the preclinical intervention link and clinical translation. Expected directions are mechanistic hypotheses, not demonstrated human mediation or clinical efficacy.

**Figure 3:**
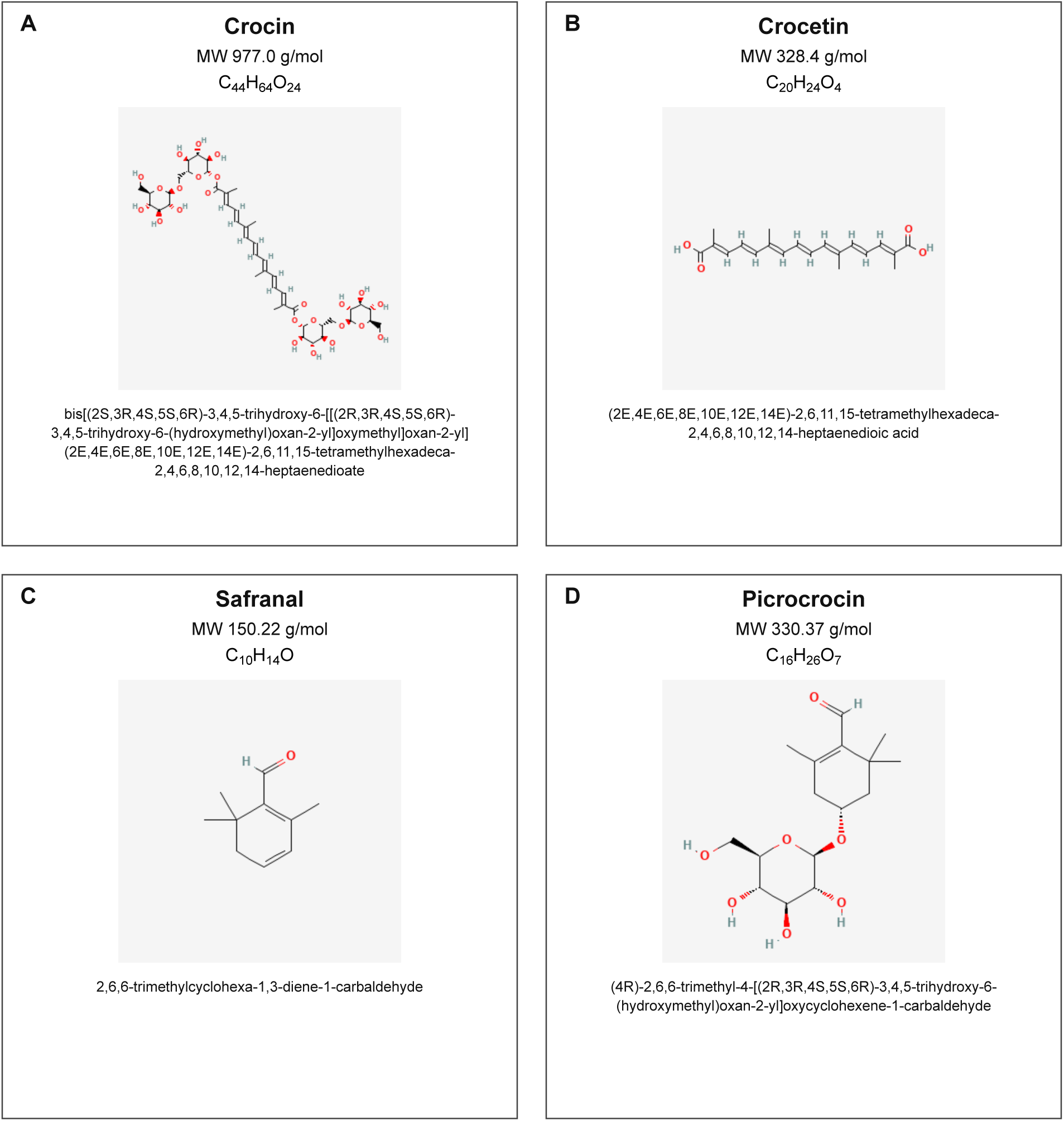
Molecular representations of saffron constituents. PubChem depictions: A, crocin; B, crocetin; C, safranal; D, picrocrocin. Each panel gives molecular weight, a molecular formula with subscripts, and the systematic IUPAC name. Whole-saffron preparations contain a mixture of constituents; these depictions identify specific molecular records.

### 1.4 Why it is important to do this review

The available trials cover different clinical populations, preparations, and treatment durations. Some report fasting glucose without HbA1c, while others provide insulin-resistance measures. A synthesis that preserves the distinction between these outcomes can help determine whether any signal is consistent across measures of glycemic control. It can also identify where apparent benefits depend on heterogeneous populations or a small number of participants. Previous reviews have already addressed saffron and crocin, with differing populations, preparations, and outcome selections (Table 4). Giannoulaki 2020 also summarized diabetes and metabolic-syndrome trials.[13] The present exploratory synthesis should be interpreted in that existing literature rather than as the first meta-analysis of this intervention.

## 2 Objectives

To assess the effects of saffron and its constituents, compared with a control, on HbA1c in humans, and to assess their effects on fasting glucose and HOMA-IR as secondary measures of glycemic regulation.

## 3 Methods

The review was informed by an Open Science Framework preregistration entitled *Effect of Saffron on Diabetes Clinical Markers*. The manuscript structure follows the Cochrane review framework, with reporting guided by PRISMA 2020.[14, 15]

### 3.1 Criteria for considering studies for this review

#### 3.1.1 Types of studies

We considered parallel-group randomized controlled trials with an intervention duration of at least four weeks. Non-randomized studies, observational studies, case reports, animal experiments, and in vitro studies were excluded from the clinical synthesis.

#### 3.1.2 Types of participants

Human participants were eligible without restrictions on age, sex, geographic location, or publication language. Eligibility was not restricted to a diagnosis of diabetes. Clinical populations were described separately to preserve distinctions among diabetes, prediabetes, metabolic syndrome, and other conditions.

#### 3.1.3 Types of interventions

Eligible interventions were saffron (*Crocus sativus*) or its constituents, including crocin, safranal, and picrocrocin, compared with placebo or another control. The intervention had to be the differing treatment between the analyzed groups. Identical background interventions were allowed in both groups. A multi-ingredient product or factorial design was not excluded solely because additional components or trial arms were present: eligibility depended on whether the selected comparison isolated saffron or its constituent. Background treatment had to be matched between the selected groups.

#### 3.1.4 Types of outcome measures

The primary outcome was HbA1c, expressed on the percentage scale. Effects on HbA1c are reported as absolute percentage-point differences, not relative percentage changes. Secondary outcomes addressed in the synthesis were fasting glucose, expressed in mg/dL, and HOMA-IR. The preregistered outcome hierarchy also encompassed random glucose, fasting insulin, postprandial glucose, metabolic measures, and adverse events. We summarized safety information descriptively when available.

### 3.2 Search methods for identification of studies

#### 3.2.1 Electronic searches

The searches contributing to this analysis were PubMed Central (PMC; June 11, 2025) and PubMed (October 5, 2025). PMC was used as a full-text resource and PubMed as a bibliographic resource. Search dates are reported separately because the searches were performed at different times.

The search combined intervention terms (*Crocus sativus*, saffron, crocin, safranal, and picrocrocin) with glycemic outcome terms, including HbA1c and related terminology, fasting or random glucose, fasting insulin, HOMA-IR, and postprandial glucose. No publication-language restriction was specified. The recorded search expression is reproduced in Appendix A.

### 3.3 Data collection and analysis

#### 3.3.1 Selection of studies

Reviewers recorded screening decisions in individual spreadsheets. Screening considered the intervention, human randomized trial design, whether the intervention effect could be isolated, relevant outcomes, endpoint measurement, and treatment duration. Any report marked for further analysis by a reviewer was reassessed for inclusion. The final synthesis used the trial comparisons retained after reassessment. Records were linked using bibliographic identifiers and titles; repeated copies in reviewer workbooks were treated as assessments of the same report.

#### 3.3.2 Data extraction and management

We extracted study characteristics, intervention and comparator descriptions, treatment duration, outcome values, measures of dispersion, and analyzed sample sizes from reports and study tables. For quantitative synthesis, we used arm-level endpoint means, standard deviations (SDs), and sample sizes. Report-level information was linked to the corresponding trial to avoid treating companion publications as separate participants. When multiple reports described a trial, information was considered at trial level and the appropriate report was used for each outcome.

Where fasting glucose was reported as a median and interquartile range, the central value was used as an approximation to the mean and the SD was approximated as the interquartile range divided by 1.35. Such estimates were interpreted cautiously because the conversion assumes a distribution that may not be well represented by the published summaries.

Trial-level recruitment, allocation, follow-up, and analysis counts were extracted where recoverable and displayed in a review-level adaptation of the CONSORT 2025 flow-diagram template. Unknown source-stage counts were retained as logical bounds in the participant-flow audit, with an open upper bound when the total number assessed could not be established. The diagram used mutually exclusive allocation-to-analysis differences for its postrandomization side branches; discontinuation and follow-up loss were not added as independent exclusions. Clinical trial stages were kept distinct from review-level screening decisions.[16, 17]

#### 3.3.3 Measures of treatment effect

We calculated between-group MDs with 95% CIs for continuous outcomes. Negative values indicate lower HbA1c, fasting glucose, or HOMA-IR in the intervention group. We synthesized endpoint values rather than interpreting within-group changes as treatment effects. An endpoint MD does not itself adjust for baseline differences between randomized groups.

#### 3.3.4 Unit of analysis issues

Each trial contributed a single non-overlapping comparison to an outcome synthesis. Participants were counted within the selected intervention and control groups. Other arms of multi-arm trials were not added to these denominators. The same participants could contribute to more than one outcome; sample sizes across outcome analyses were therefore not summed.

#### 3.3.5 Dealing with missing data

The quantitative syntheses used available analyzed sample sizes and outcome summaries. These denominators were distinguished from the total numbers randomized when studies included withdrawals or additional intervention arms. No participant-level missing outcomes were imputed for this review.

#### 3.3.6 Assessment of heterogeneity

We examined clinical variation in participants, preparations, doses, and treatment durations. Statistical heterogeneity was described using the Chi-squared statistic, *I*^2^, and the between-study variance (*τ* ^2^). We interpreted these statistics alongside the number and size of trials rather than using a single threshold to determine the model.

#### 3.3.7 Data synthesis

We used inverse-variance random-effects meta-analysis, consistent with the preregistered analysis plan. Analyses used the DerSimonian–Laird estimator of *τ* ^2^ and normal-approximation CIs. For each study, the variance of the endpoint MD was calculated as *SD_T_*^2^ */n_T_* + *SD_C_*^2^ */n_C_*. Analyses were prepared in RevMan; arithmetic checks, endpoint verification, and fixed-effect sensitivity analyses were performed using Python 3.12. Statistical tests were two-sided. We interpreted secondary-outcome findings in the context of multiple outcomes and heterogeneity.

#### 3.3.8 Sensitivity analysis

For each of the three outcomes, we repeated the synthesis using a fixed-effect inverse-variance model to examine dependence on the weighting model. We compared the direction, magnitude, and precision of estimates rather than interpreting statistical significance alone.

## 4 Results

### 4.1 Study selection and noteworthy comparisons

The PMC and PubMed exports contained 1,640 and 74 records, respectively. Linking duplicate bibliographic records removed 46 repeated entries, leaving 1,668 distinct reports. Screening sheets contained a substantive decision for 1,663 reports; five reports had no completed decision. Twenty-nine reports had been marked for further analysis by at least one reviewer. Twelve trial comparisons were represented in the quantitative syntheses. The available record-level selection accounting is shown in Figure 1; retrieval and full-text-stage totals were not separately recorded. Figure 1 therefore marks reconstructed stage counts as estimates: 29 reports sought and assessed, none assumed not retrieved, 15 provisionally outside the synthesis, and 14 reports tentatively linked to the 12 studies. Supplementary Table S2 provides the recorded reviewer assessments retained in the PDF screening appendix; the complete electronic screening table accompanies the review. The 12 unresolved data/contrast dispositions do not establish ineligibility.

Some comparator structures require distinction between a genuinely unequal co-intervention and matched background treatment. Giannoulaki 2024 compared otherwise matched probiotic, prebiotic, and magnesium formulations with or without saffron extract. The selected Rajabi 2022 and Rajabi 2024 comparisons used saffron-only and placebo-only groups, excluding the exercise arms from those contrasts.[18–20] The additional metformin arm in Kotanidou 2023 was not part of the selected saffron–placebo comparison.[21]

Several reports in the current searches warrant reassessment of recorded labels or quantitative availability (Table 2; Supplementary Table S1). Azimi 2014 used saffron with black tea against black tea alone, and Saghafi 2025 compared crocin with placebo on a metformin background in both groups; a shared background does not by itself violate the isolated-intervention criterion.[22, 23] Sepahi 2018 was a randomized placebo-controlled diabetic maculopathy trial despite a screening label indicating that it was not a clinical trial.[24] These reports were not represented in the current quantitative set; their recorded labels should not be interpreted as independently validated reasons for exclusion.

**Table 1:**
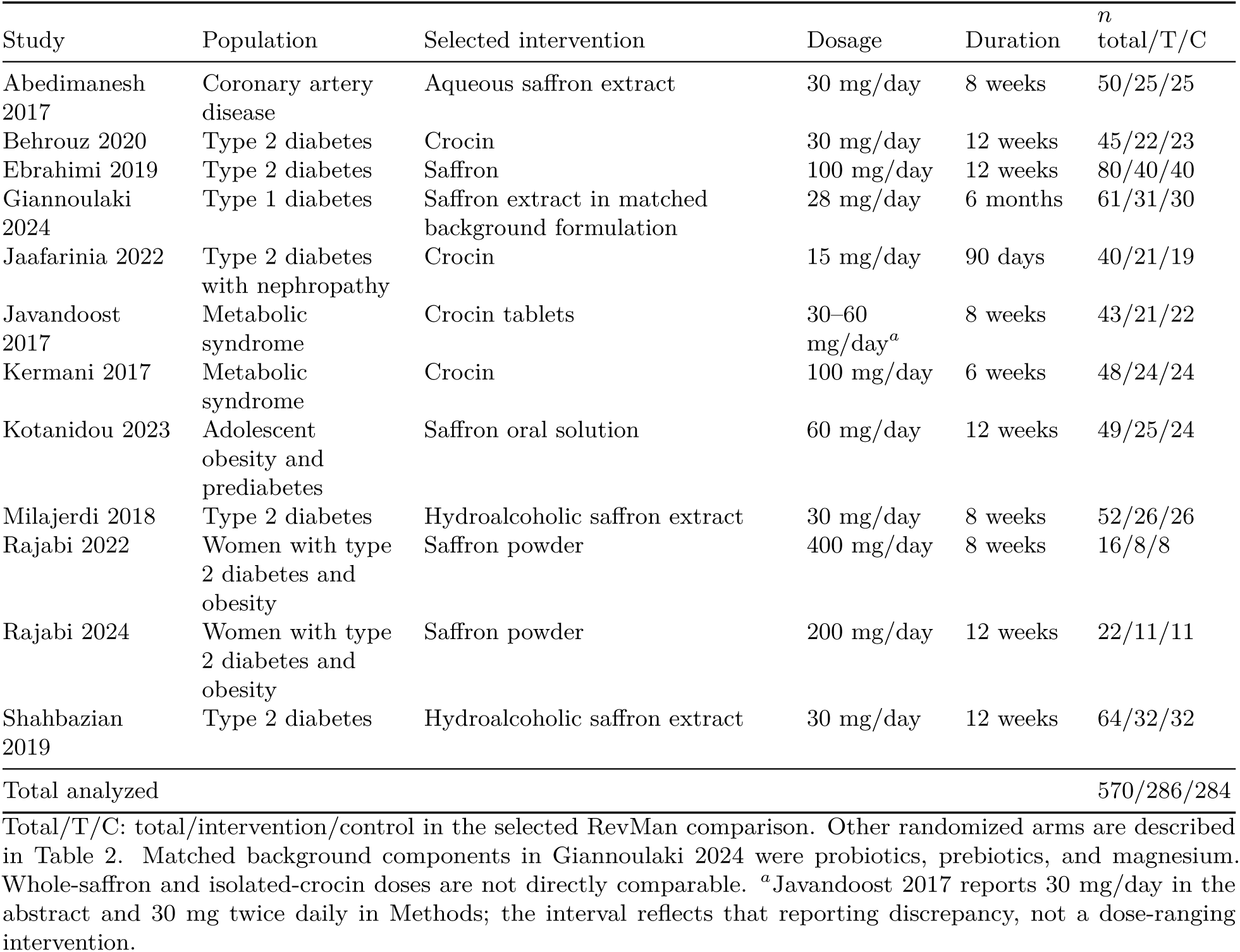
Characteristics of the trial comparisons included in the quantitative syntheses.

| Study | Population | Selected intervention | Dosage | Duration | *n*<br>total/T/C |
|---|---|---|---|---|---|
| Abedimanesh 2017 | Coronary artery disease | Aqueous saffron extract | 30 mg/day | 8 weeks | 50/25/25 |
| Behrouz 2020 | Type 2 diabetes | Crocin | 30 mg/day | 12 weeks | 45/22/23 |
| Ebrahimi 2019 | Type 2 diabetes | Saffron | 100 mg/day | 12 weeks | 80/40/40 |
| Giannoulaki 2024 | Type 1 diabetes | Saffron extract in matched background formulation | 28 mg/day | 6 months | 61/31/30 |
| Jaafarinia 2022 | Type 2 diabetes with nephropathy | Crocin | 15 mg/day | 90 days | 40/21/19 |
| Javandoost 2017 | Metabolic syndrome | Crocin tablets | 30–60 mg/day<sup>*a*</sup> | 8 weeks | 43/21/22 |
| Kermani 2017 | Metabolic syndrome | Crocin | 100 mg/day | 6 weeks | 48/24/24 |
| Kotanidou 2023 | Adolescent obesity and prediabetes | Saffron oral solution | 60 mg/day | 12 weeks | 49/25/24 |
| Milajerdi 2018 | Type 2 diabetes | Hydroalcoholic saffron extract | 30 mg/day | 8 weeks | 52/26/26 |
| Rajabi 2022 | Women with type 2 diabetes and obesity | Saffron powder | 400 mg/day | 8 weeks | 16/8/8 |
| Rajabi 2024 | Women with type 2 diabetes and obesity | Saffron powder | 200 mg/day | 12 weeks | 22/11/11 |
| Shahbazian 2019 | Type 2 diabetes | Hydroalcoholic saffron extract | 30 mg/day | 12 weeks | 64/32/32 |
| Total analyzed |  |  |  |  | 570/286/284 |

Total/T/C: total/intervention/control in the selected RevMan comparison. Other randomized arms are described in Table 2. Matched background components in Giannoulaki 2024 were probiotics, prebiotics, and magnesium. Whole-saffron and isolated-crocin doses are not directly comparable. <sup>*a*</sup>Javandoost 2017 reports 30 mg/day in the abstract and 30 mg twice daily in Methods; the interval reflects that reporting discrepancy, not a dose-ranging intervention.

**Table 2:**
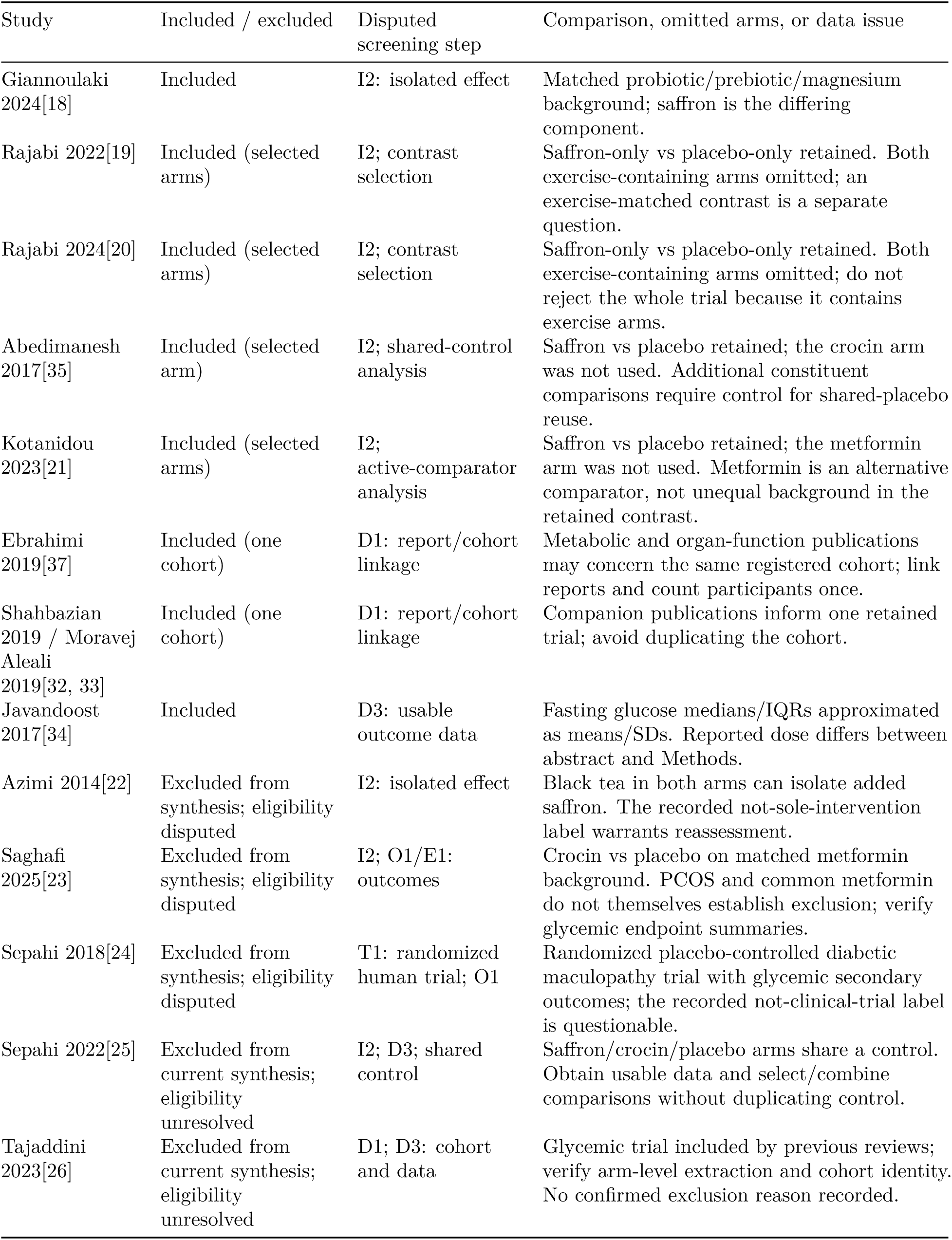
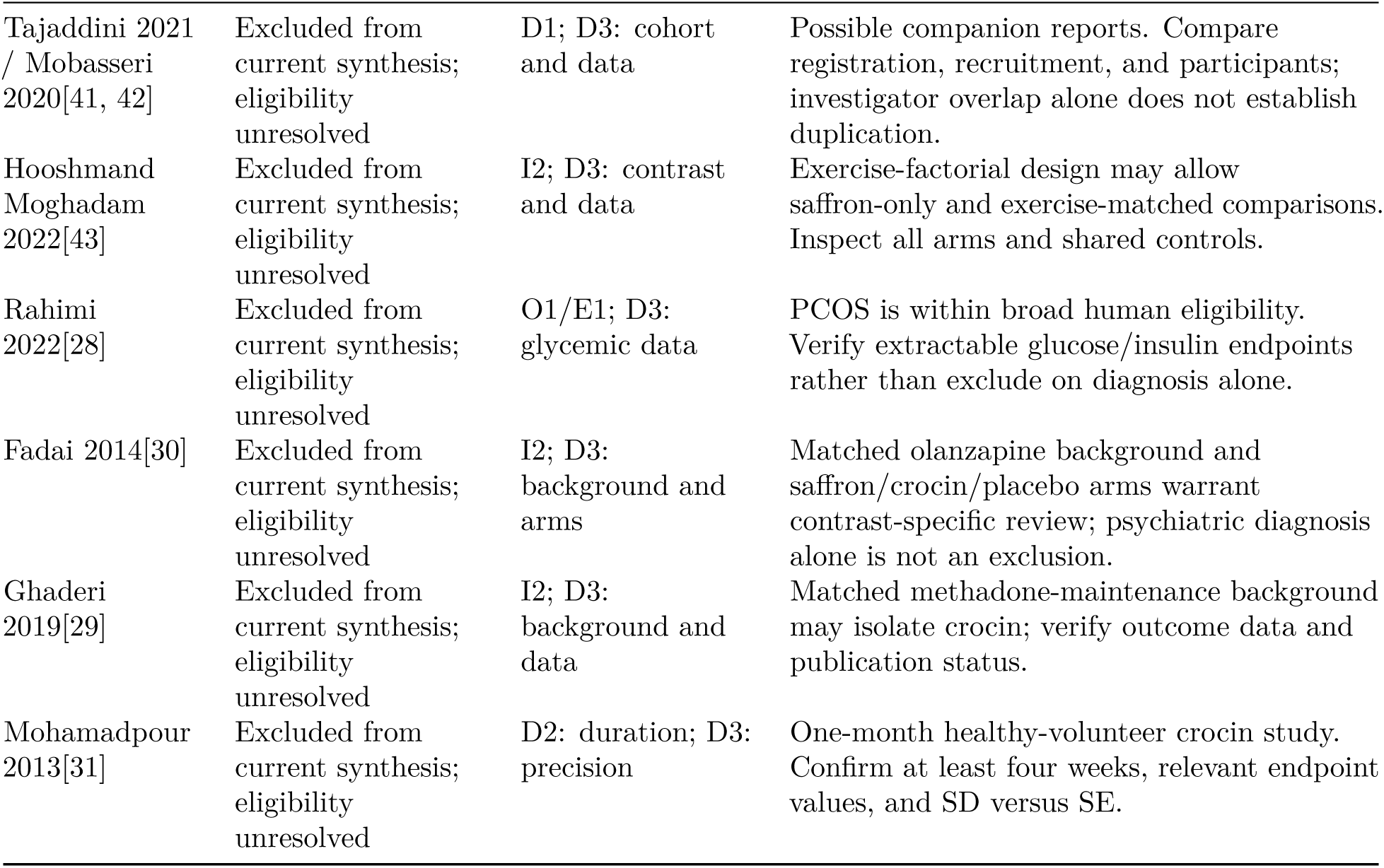
Notable study and comparison decisions in the current synthesis.

**Table 3:**
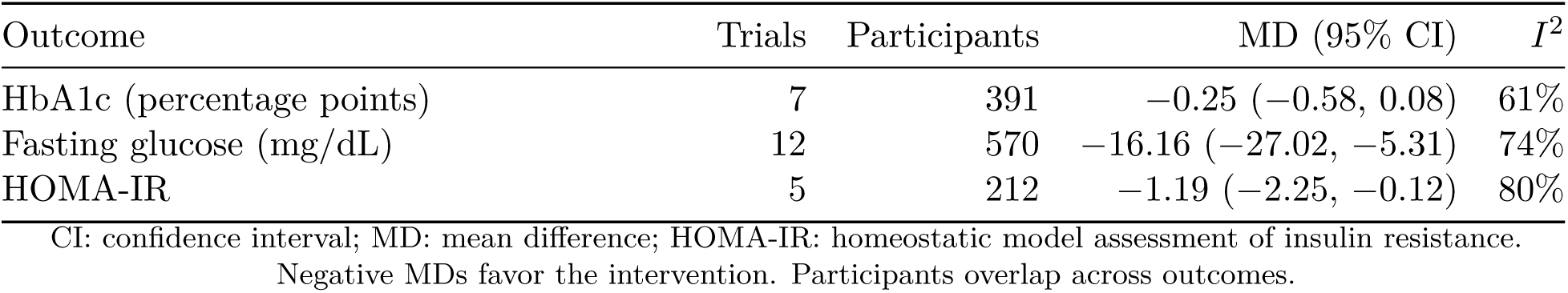
Random-effects meta-analysis of glycemic outcomes.

| Outcome | Trials | Participants | MD (95% CI) | *I*<sup>2</sup> |
|---|---|---|---|---|
| HbA1c (percentage points) | 7 | 391 | −0.25 (−0.58, 0.08) | 61% |
| Fasting glucose (mg/dL) | 12 | 570 | −16.16 (−27.02, −5.31) | 74% |
| HOMA-IR | 5 | 212 | −1.19 (−2.25, −0.12) | 80% |

CI: confidence interval; MD: mean difference; HOMA-IR: homeostatic model assessment of insulin resistance.
Negative MDs favor the intervention. Participants overlap across outcomes.

Sepahi 2022 and Tajaddini 2023 were included in previous glycemic reviews but were not represented in the supplied analyses.[3, 25–27] Their absence requires verification of usable outcome data, shared controls, and possible companion reports; their absence from the current synthesis is not a confirmed eligibility exclusion. Other flagged reports involve PCOS, methadone maintenance, olanzapine background treatment, or safety monitoring in healthy volunteers; these populations and matched treatments are not automatically excluded under the broad participant criteria.[28–31]

For retained studies, Shahbazian 2019 and Moravej Aleali 2019 were linked to the relevant trial rather than counted as independent participants.[32, 33] The fasting-glucose synthesis used an approximate SD derived from the reported interquartile range in Javandoost 2017.[34] The current Abedimanesh comparison selected saffron versus placebo from a trial also containing a crocin arm; additional eligible contrasts require care to avoid duplicate use of a shared control.[35]

### 4.2 Description of studies

#### 4.2.1 Included studies

Twelve randomized trials contributed data to the quantitative syntheses (Table 1). The analyzed comparisons comprised 570 participants: 286 assigned to saffron or crocin and 284 assigned to control. These numbers describe the selected comparisons rather than all arms randomized in every source trial.

Seven trials involved adults with type 2 diabetes, including one trial in diabetic nephropathy; one involved adults with type 1 diabetes; two involved metabolic syndrome; one involved coronary artery disease; and one involved adolescents with obesity and prediabetes.[18–21, 32, 34–40] Interventions included saffron powder or extracts and isolated crocin. Treatment periods ranged from six weeks to six months. All selected comparisons used a placebo or a matched control formulation.

Included/excluded refers to representation in the current quantitative synthesis; it does not convert unresolved reports into confirmed eligibility exclusions. D1: duplicate/report linkage; I2: isolated intervention effect; T1: randomized human trial; O1: relevant outcome; E1: endpoint measurement; D2: minimum duration; D3: usable outcome data. Contrast selection and shared-control handling are analysis decisions following screening. The full decision workflow is in Supplementary Figure S10. No additional comparison is added to the pooled results by this table.

The adapted participant-flow diagram (Figure 4) retains trial order across every stage and distinguishes all randomized arms from the selected review comparisons. Of 720 randomized participants, 93 were allocated to other arms and 627 to the selected comparisons. The current analyses represent 286 of 314 intervention participants and 284 of 313 controls; the respective allocation-to-analysis differences are 28 and 29. These differences do not establish that all such participants were lost to follow-up. Source qualifications and stage-specific bounds are given in Supplementary Table S3. Behrouz 2020 reports analyzed arm counts of 23 and 22, whereas the supplied RevMan data use 22 and 23; the diagram follows the current RevMan denominators, with the combined trial count remaining 45. The current pooled inputs are unchanged.

**Figure 4:**
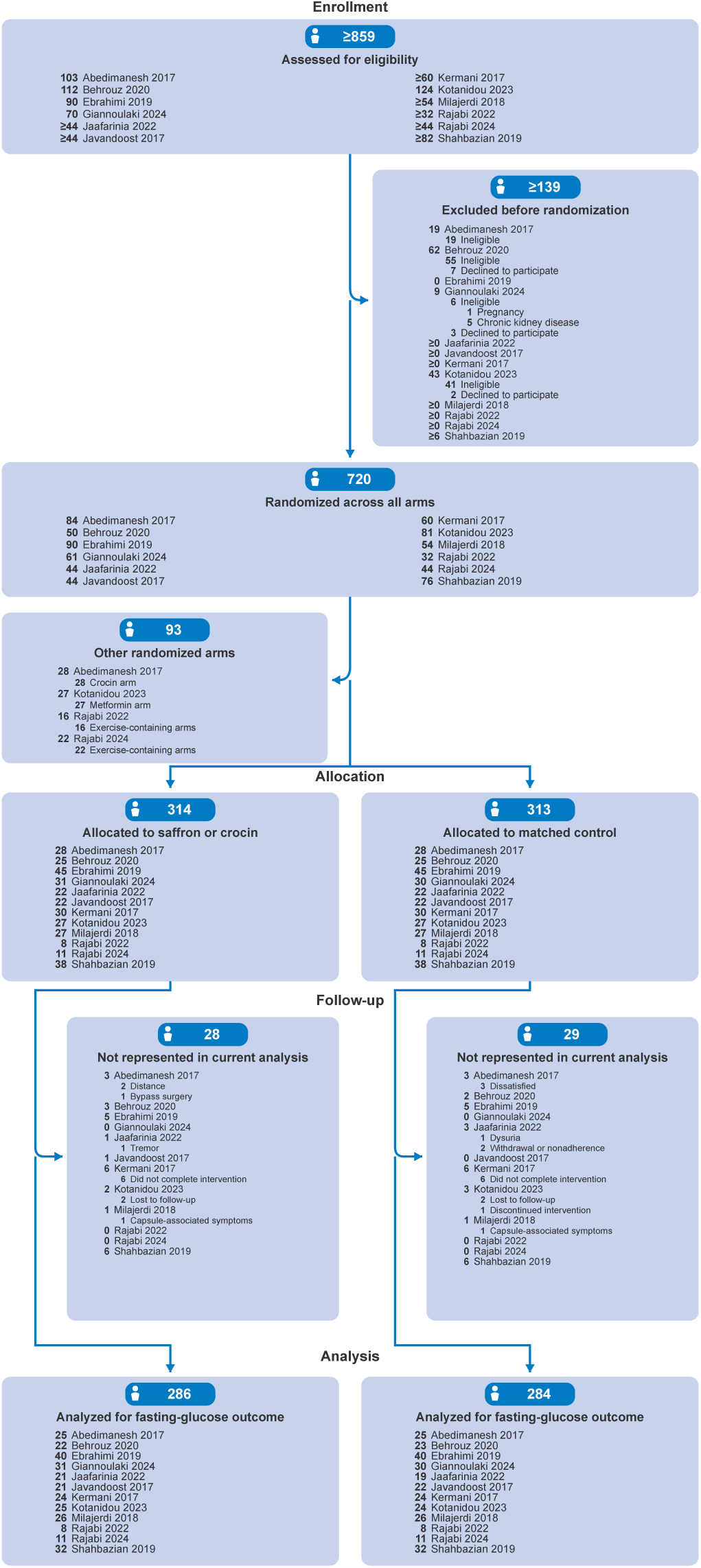
Participant flow across the included trials. Trials follow the RevMan fasting-glucose order. Blue badges give summed counts or bounds; bold, right-aligned trial counts precede regular descriptions. Only reported subcategories are indented. Ranges are logical bounds, and *≥* denotes an open upper bound. Side branches are mutually exclusive differences between allocation and analysis; reported reasons are nested where available. Analysis counts use the current RevMan inputs: 286 intervention participants and 284 control participants, totaling 570. Table S3 provides the source qualifications.

### 4.3 Effects of interventions

#### 4.3.1 Glycated hemoglobin

Seven trials with 391 participants (197 intervention and 194 control) contributed HbA1c data. The pooled MD was *−*0.25 percentage points (95% CI *−*0.58 to 0.08; *P* = 0.14; Figure 5). There was heterogeneity between trial estimates (*τ* ^2^ = 0.10; *χ*^2^ = 15.53, 6 degrees of freedom, *P* = 0.02; *I*^2^ = 61%). The CI was compatible with a reduction in HbA1c as well as little difference between groups.

**Figure 5:**
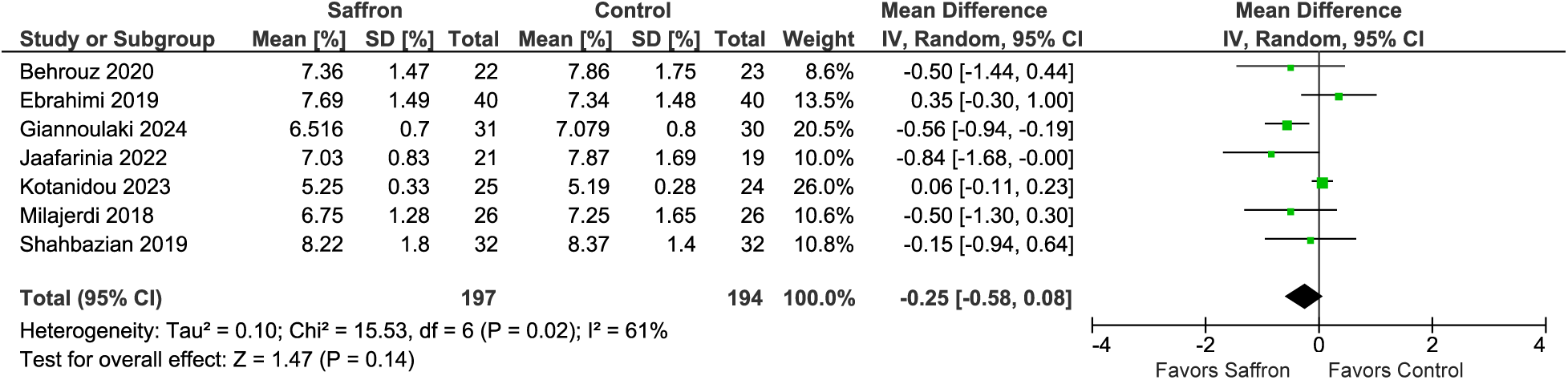
Effect of saffron or crocin on HbA1c. Values are endpoint means on the percentage scale. The summary estimate is a mean difference in percentage points. IV: inverse variance.

#### 4.3.2 Fasting glucose

All 12 trials contributed fasting-glucose data from 570 participants. Saffron or crocin was associated with lower fasting glucose compared with control (MD *−*16.16 mg/dL, 95% CI *−*27.02 to *−*5.31; *P* = 0.004; Figure 6). Heterogeneity was substantial (*τ* ^2^ = 226.14; *χ*^2^ = 42.53, 11 degrees of freedom, *P <* 0.001; *I*^2^ = 74%). The average estimate therefore does not describe a uniform response across populations and preparations.

**Figure 6:**
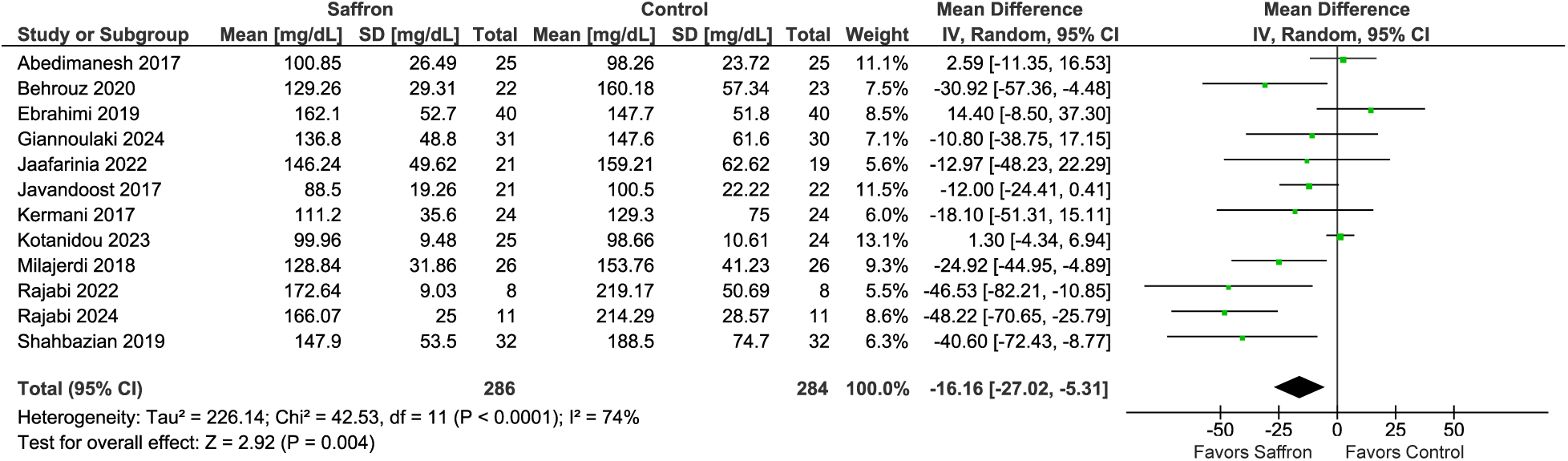
Effect of saffron or crocin on fasting glucose. Mean differences are expressed in mg/dL. The intervention column includes both saffron preparations and isolated crocin.

#### 4.3.3 Insulin resistance

Five trials with 212 participants (106 per group) contributed HOMA-IR data. The pooled MD was *−*1.19 (95% CI *−*2.25 to *−*0.12; *P* = 0.029; Figure 7). There was considerable heterogeneity (*τ* ^2^ = 1.12; *χ*^2^ = 20.02, 4 degrees of freedom, *P* = 0.0005; *I*^2^ = 80%). The small number of trials, heterogeneity, and proximity of the upper CI to zero limit interpretation of the pooled result.

**Figure 7:**
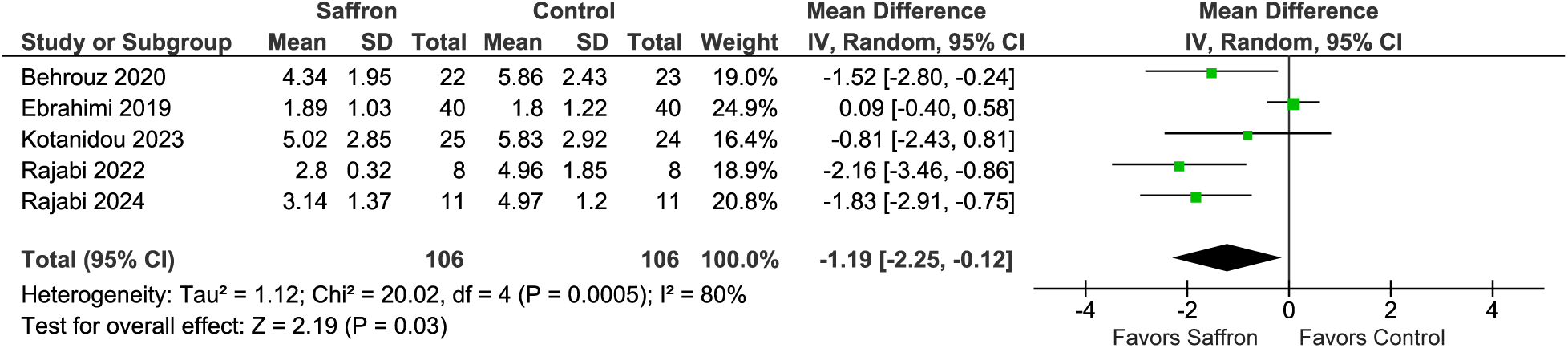
Effect of saffron or crocin on HOMA-IR. Negative mean differences indicate lower estimated fasting insulin resistance in the intervention group.

#### 4.3.4 Sensitivity analyses

Under the fixed-effect model, the HbA1c estimate was −0.08 percentage points (95% CI −0.22 to 0.06; *P* = 0.283). The fasting-glucose estimate was −5.67 mg/dL (95% CI −9.91 to −1.43; *P* = 0.009), and the HOMA-IR estimate was −0.57 (95% CI −0.96 to −0.18; *P* = 0.004). These estimates were smaller in magnitude than their random-effects counterparts. The direction of the fasting-glucose and HOMA-IR results was retained, but their magnitude depended on the weighting model. The adolescent trial, which had relatively small outcome SDs, contributed approximately 69% of the fixed-effect HbA1c weight and 56% of the fixed-effect fasting-glucose weight. The corresponding RevMan fixed-effect forest plots are presented in Supplementary Figures S6–S8.

#### 4.3.5 Adverse events

Safety information was descriptive. Behrouz 2020 reported no serious adverse events during the intervention.[36] In Jaafarinia 2022, mild tremor was reported in one participant assigned to crocin and dysuria in one participant assigned to placebo.[38] In Milajerdi 2018, one participant in each group withdrew after symptoms described as complications of the capsules, particularly headache.[40] These observations do not provide a precise comparative estimate of harms.

## 5 Discussion

### 5.1 Summary of main results

Across 12 randomized trials, saffron or crocin was associated with lower fasting glucose. The pooled HOMA-IR estimate also favored intervention, although its CI narrowly excluded zero and heterogeneity was considerable. The primary HbA1c analysis did not establish a clear reduction: its CI included both a potentially relevant decrease and little difference. These findings support distinguishing an effect on fasting measures from a sustained effect on overall glycemic exposure.

The fasting-glucose finding was consistent in direction under random-effects and fixed-effect models, but the estimated magnitude changed appreciably. The HbA1c estimate was also closer to zero under fixed-effect weighting. This difference reflects the balance between smaller, more variable trials and a trial with small SDs, rather than providing an independent replication of an effect. It argues against treating a single pooled MD as the expected response for every clinical population.

### 5.2 Overall completeness and applicability of evidence

The trials addressed several distinct populations. Adults receiving treatment for type 2 diabetes, adults with type 1 diabetes, people with metabolic syndrome or coronary artery disease, and adolescents with obesity and prediabetes differ in baseline glycemia, physiology, and background care. The pooled effects cannot establish that these groups benefit equally. In particular, a trial-level average cannot substitute for a subgroup interaction test or a participant-level assessment of treatment-effect modification.

The intervention category was similarly broad. Whole saffron, hydroalcoholic or aqueous extracts, and isolated crocin differ in constituent concentrations and pharmacological exposure. The dose of a whole extract cannot be equated with the dose of isolated crocin. Variation in preparation may contribute to heterogeneity and prevents selection of an optimal regimen from the average estimates. The analyzed comparisons generally evaluated supplementation alongside background care; they do not support replacing established glucose-lowering treatment.

Treatment periods of six weeks to six months provide limited information about persistence of response. Shorter follow-up is particularly relevant to interpretation of HbA1c, which integrates glycemic exposure over time.[1] The syntheses do not establish effects on diabetes progression, cardiovascular events, microvascular complications, or mortality. Descriptive adverse-event reports also cannot establish safety during long-term use.

### 5.3 Potential limitations of the review and synthesis

The outcome syntheses included relatively few trials, several with small analyzed groups. Heterogeneity was present for each outcome and was greatest for HOMA-IR. The normal-approximation intervals and estimated between-study variances should therefore be interpreted in the context of limited information. A pooled secondary-outcome result with a CI close to zero is not, by itself, compelling evidence of a reproducible clinical benefit.

We synthesized unadjusted endpoint summaries. Although randomization supports a between-group comparison, chance baseline imbalances can influence an endpoint difference in a small trial. These estimates are not equivalent to baseline-adjusted treatment effects or to differences in change from baseline. Moreover, replacing medians and quartiles with approximate means and SDs can introduce error when an outcome is skewed. This issue is relevant to the fasting-glucose data from Javandoost 2017.[34]

The same participants contributed to multiple outcomes, and the secondary outcomes were correlated. Fasting-glucose and HOMA-IR findings should therefore be interpreted as related signals within a broader outcome set rather than as independent confirmations. The available pooled results also do not identify whether apparent effects are attributable to saffron extracts, isolated crocin, participant characteristics, or background therapy. The documented search scope is limited to PMC and PubMed, and several potentially eligible reports within those results were not represented in the quantitative set. Incomplete report-level dispositions and retrieval-stage accounting limit assessment of the completeness of selection.

### 5.4 Comparison with previous reviews

Previous meta-analyses generally reported reductions in fasting glucose, whereas their HbA1c and insulin-resistance findings were less uniform (Table 4). For example, Liu 2024 reported an HbA1c MD of *−*0.22 percentage points and fasting-glucose MD of *−*8.42 mg/dL; Zhang 2025b, the metabolic-syndrome review, reported corresponding MDs of *−*0.25 percentage points and *−*6.67 mg/dL.[3, 4] Both reviews reported no clear HOMA-IR effect. The current endpoint synthesis differs in the selected reports, populations, preparations, and weighting. These differences, and the potentially eligible reports identified above, prevent interpreting discrepancies as evidence that one pooled estimate is intrinsically more reliable.

**Table 4:**
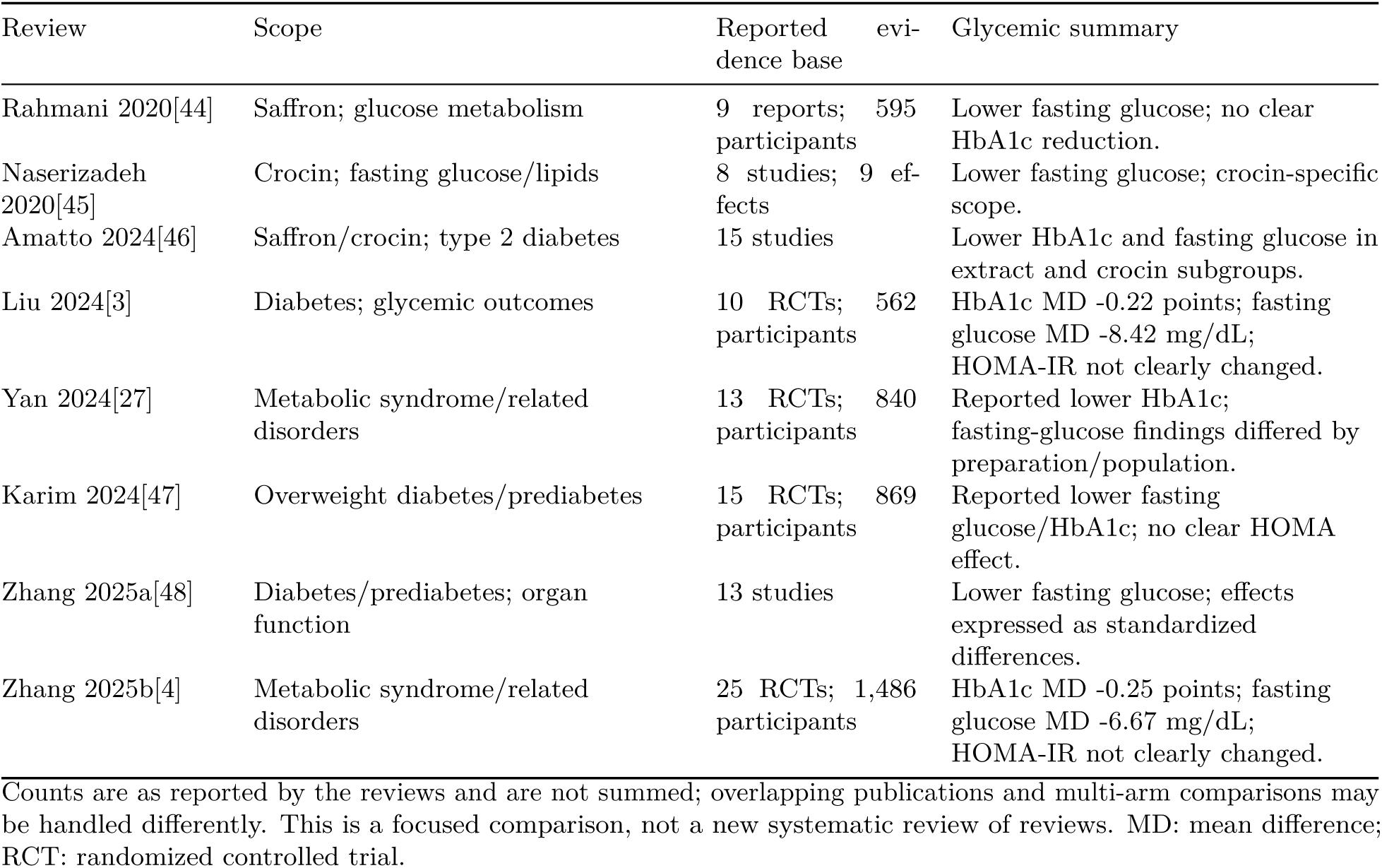
Selected previous meta-analyses of saffron or crocin with glycemic outcomes.

| Review | Scope | Reported evi-dence base | Glycemic summary |
|---|---|---|---|
| Rahmani 2020[44] | Saffron; glucose metabolism | 9 reports; 595 participants | Lower fasting glucose; no clear HbA1c reduction. |
| Naserizadeh 2020[45] | Crocin; fasting glucose/lipids | 8 studies; 9 ef-fects | Lower fasting glucose; crocin-specific scope. |
| Amatto 2024[46] | Saffron/crocin; type 2 diabetes | 15 studies | Lower HbA1c and fasting glucose in extract and crocin subgroups. |
| Liu 2024[3] | Diabetes; glycemic outcomes | 10 RCTs; 562 participants | HbA1c MD -0.22 points; fasting glucose MD -8.42 mg/dL; HOMA-IR not clearly changed. |
| Yan 2024[27] | Metabolic syndrome/related disorders | 13 RCTs; 840 participants | Reported lower HbA1c; fasting-glucose findings differed by preparation/population. |
| Karim 2024[47] | Overweight diabetes/prediabetes | 15 RCTs; 869 participants | Reported lower fasting glucose/HbA1c; no clear HOMA effect. |
| Zhang 2025a[48] | Diabetes/prediabetes; organ function | 13 studies | Lower fasting glucose; effects expressed as standardized differences. |
| Zhang 2025b[4] | Metabolic syndrome/related disorders | 25 RCTs; 1,486 participants | HbA1c MD -0.25 points; fasting glucose MD -6.67 mg/dL; HOMA-IR not clearly changed. |

Counts are as reported by the reviews and are not summed; overlapping publications and multi-arm comparisons may be handled differently. This is a focused comparison, not a new systematic review of reviews. MD: mean difference; RCT: randomized controlled trial.

### 5.5 Biological interpretation

AMPK-related pathways are plausible candidates for further investigation. Saffron-associated changes in GLUT4 and AMPK-related expression in experimental models, and the loss of crocin-associated metabolic effects after hepatic AMPK downregulation in mice, provide more specific mechanistic support than a general antioxidant rationale.[6, 7] The KEGG AMPK map offers a framework for tracing glucose transport and lipid-metabolism branches, including GLUT4, acetyl-CoA carboxylase, and the regulation of fatty-acid oxidation.[8]

However, these experiments do not demonstrate a direct drug–target interaction in humans. In Behrouz 2020, which measured phosphorylated AMPK in peripheral blood mononuclear cells, the intervention did not significantly alter that marker.[36] The clinical findings are compatible with several explanations, including tissue-specific mechanisms not captured by the assay, effects through other pathways, or variation unrelated to AMPK activation. Mechanistic diagrams should preserve the distinction between established signaling relationships and hypothesized intervention effects.

## 6 Authors’ conclusions

### 6.1 Implications for practice

The primary implication of this review is the need for large-scale clinical trials with well-controlled head-to-head comparisons between the different constituents of saffron and standardized whole-saffron preparations. The analyzed trials suggest possible reductions in fasting glucose, with a less stable signal for HOMA-IR. They do not establish a clear reduction in HbA1c or a durable clinical benefit. Differences among populations and preparations, together with heterogeneity and limited safety information, preclude a general treatment recommendation or selection of a preferred product or dose.

### 6.2 Implications for research

Further randomized trials should use preparations with documented constituent content, maintain comparable background therapy, and follow participants long enough to evaluate sustained HbA1c responses. Studies should report endpoint and change values with their dispersion, analyzed and randomized denominators, adherence, withdrawals, and adverse events. Mechanistic substudies should prespecify tissue-relevant measures and distinguish pathway activation from changes in protein abundance. Trials should also report outcomes important to patients rather than relying exclusively on surrogate markers.

## Data Availability

All data produced in the present study are available upon reasonable request to the authors

## A Recorded electronic search expression

The following expression was recorded in the search workbooks and preregistration:

(“Crocus sativus” OR saffron OR crocin OR safranal OR picrocrocin) AND (A1c OR HbA1c OR HbA1 OR “Hb A1” OR A1a OR “A1a-1” OR A1b OR (glycated AND hemoglobin) OR (glycosylated AND hemoglobin) OR (fructated AND hemoglobin) OR glycohemoglobin OR (fasting AND glucose) OR “random glucose” OR (fasting AND insulin) OR “HOMA-IR” OR “Homeostatic Model Assessment for Insulin Resistance” OR “2 hour glucose” OR (postprandial AND glucose)).

