## Supplementary Materials for "Effects of saffron and crocin on glycemic control: an exploratory systematic review and meta-analysis of randomized trials"

Juno Kim and colleagues

This supplement contains the completed item-by-item reporting assessments for PRISMA 2020, PRISMA 2020 for Abstracts, and PRISMA-S; fixed-effect sensitivity plots; a catalog of noteworthy clinical reports in the current search records; an adapted participant-flow diagram; and a separate recorded-screening appendix.[1, 2]

**Checklist notation.** R means reported; P means partly reported; NR means not reported; NA means not applicable to the current analysis. Every checklist item has been filled. These assessments identify reporting gaps and do not imply that unreported methods were performed. Item prompts are paraphrased, with attribution to the original checklists. Main-manuscript locations refer to the separately paginated accompanying manuscript.

**Scope.** The current analysis uses PMC and PubMed searches. CENTRAL, Embase, and ClinicalTrials.gov are not part of this version's search yield or flow diagram. Contextual checks of previous reviews do not add trials to the quantitative set.

- Figures S1–S3: PRISMA 2020 checklist (three panels covering all 42 entries).
- Figure S4: PRISMA 2020 for Abstracts (12 items).
- Figure S5: PRISMA-S (16 items).
- Figures S6–S8: fixed-effect HbA1c, fasting-glucose, and HOMA-IR sensitivity analyses.
- Table S1: noteworthy reports in the current search records.
- Figures S9–S11: selection flow, screening workflow, and CONSORT-style participant accounting.
- Table S2: recorded screening decisions (separate landscape appendix).
- Table S3: participant-flow source audit and bounds.

| Item | Reporting topic | Status | Manuscript location and assessment |
|---|---|---|---|
| 1 | Review title | R | Title identifies a systematic review and meta-analysis. |
| 2 | Abstract | P | Structured abstract; see Supplementary Figure S4 for item-level assessment. |
| 3 | Rationale | R | Main p. 2; existing evidence and review rationale. |
| 4 | Objectives | R | Main p. 7; primary and secondary objectives. |
| 5 | Eligibility | P | Main p. 7; criteria specified, but final dispositions remain incomplete. |
| 6 | Sources and dates | R | Main p. 8; PMC and PubMed with individual search dates. |
| 7 | Search expressions | P | Main p. 29; recorded generic expression; exact platform-specific runs unavailable. |
| 8 | Selection procedure | P | Main p. 8; workflow described; per-stage independent reviewer counts not reported. |
| 9 | Data collection | P | Main p. 8; extracted fields described; independent extraction and author-contact details incomplete. |
| 10a | Outcomes | P | Main p. 7; outcome hierarchy and endpoints; full selection of all compatible results not reported. |
| 10b | Other data fields | P | Main p. 8 and Table 1; core study variables, but not a complete extraction dictionary. |
| 11 | Bias assessment methods | NR | No completed risk-of-bias methods or assessments reported. |
| 12 | Effect measures | R | Main p. 9; endpoint mean differences and outcome units. |
| 13a | Synthesis eligibility | P | Main p. 8 and Main p. 10; selected contrasts and unresolved candidate reports. |

**Figure S1: PRISMA 2020 reporting checklist, part 1 of 3.** Item prompts are paraphrased from PRISMA 2020. R: reported; P: partly reported; NR: not reported; NA: not applicable. Main-manuscript page numbers refer to the accompanying manuscript.

| Item | Reporting topic | Status | Manuscript location and assessment |
| --- | --- | --- | --- |
| 13b | Data preparation | R | Main p. 8; SD approximation and treatment of available endpoint summaries. |
| 13c | Tables and graphics | R | Main Tables 1–4 and forest plots; Supplementary Figures S6–S8. |
| 13d | Pooling methods | R | Main p. 9; estimator, variance, intervals, heterogeneity, and software. |
| 13e | Heterogeneity exploration | P | Clinical differences discussed; no formal subgroup or meta-regression analysis reported. |
| 13f | Sensitivity methods | R | Main p. 9; fixed-effect analyses for all three outcomes. |
| 14 | Missing-results bias methods | NR | No completed reporting-bias assessment methods reported. |
| 15 | Certainty methods | NR | No completed certainty-of-evidence assessment methods reported. |
| 16a | Selection results | P | Main p. 10 and Figure 1; record accounting provided, retrieval/full-text stages incomplete. |
| 16b | Noteworthy exclusions | P | Main p. 10 and Supplementary Table S1; disputed labels identified, final exclusion reasons incomplete. |
| 17 | Study characteristics | R | Main p. 11 and Table 1; selected-comparison populations, interventions, duration, and sample sizes. |
| 18 | Study bias results | NR | No completed trial-level risk-of-bias judgments reported. |
| 19 | Individual estimates | R | Main forest plots show group summaries and individual effects with 95% CIs. |
| 20a | Contributing evidence | P | Main p. 11 and Table 1; characteristics reported, risk-of-bias judgments unavailable. |
| 20b | Pooled estimates | R | Main results and Table 2; outcome-specific counts, estimates, intervals, and heterogeneity. |

**Figure S2: PRISMA 2020 reporting checklist, part 2 of 3.** Item prompts are paraphrased from PRISMA 2020. R: reported; P: partly reported; NR: not reported; NA: not applicable. Main-manuscript page numbers refer to the accompanying manuscript.

| Item | Reporting topic | Status | Manuscript location and assessment |
|---|---|---|---|
| 20c | Heterogeneity findings | NA | No formal subgroup or meta-regression analyses were conducted in this synthesis. |
| 20d | Sensitivity findings | R | Main p. 15; Supplementary Figures S6–S8. |
| 21 | Missing-results bias findings | NR | No completed reporting-bias assessment results reported. |
| 22 | Certainty findings | NR | No completed GRADE or other certainty ratings reported. |
| 23a | Interpretation | R | Main p. 19; primary and secondary findings compared with previous reviews. |
| 23b | Evidence limitations | R | Main p. 19; sample size, variability, approximations, and applicability. |
| 23c | Review-process limitations | R | Main p. 19; limited search scope and incomplete candidate dispositions. |
| 23d | Implications | R | Main p. 20 and Main p. 21. |
| 24a | Registration | P | Main p. 7; OSF protocol title, but no verifiable identifier or registration date. |
| 24b | Protocol access | P | Main p. 7; locally supplied protocol; public access URL not reported. |
| 24c | Protocol amendments | NR | Protocol amendment/deviation reporting is not included. |
| 25 | Support | R | Main p. 21; no funding declared. |
| 26 | Competing interests | R | Main p. 21; no conflicts declared. |
| 27 | Materials availability | P | Main p. 21; on-request data statement; public repository and complete material-access details not reported. |

**Figure S3: PRISMA 2020 reporting checklist, part 3 of 3.** Item prompts are paraphrased from PRISMA 2020. R: reported; P: partly reported; NR: not reported; NA: not applicable. Main-manuscript page numbers refer to the accompanying manuscript.

| Item | Reporting topic | Status | Manuscript location and assessment |
|---|---|---|---|
| 1 | Systematic-review title | R | Title identifies the design. |
| 2 | Review question | R | Objectives paragraph. |
| 3 | Eligibility | R | Selection criteria paragraph. |
| 4 | Sources and last dates | R | Search methods: PMC June 11, 2025; PubMed October 5, 2025. |
| 5 | Risk-of-bias approach | NR | No risk-of-bias assessment method in the abstract. |
| 6 | Synthesis approach | R | Data collection and analysis: inverse-variance random-effects MDs and CIs. |
| 7 | Studies and participants | R | Main results: 12 trials, 570 participants, population summary. |
| 8 | Main outcome results | R | Main results: study/participant counts and MDs/CIs for all three outcomes. |
| 9 | Evidence limitations | R | Conclusions: variation in populations, preparations, and trial sizes. |
| 10 | Interpretation | R | Authors conclusions: direction, uncertainty, and research implications. |
| 11 | Funding | R | Funding: none. |
| 12 | Registration | NR | Register and registration number are not provided in the abstract. |

**Figure S4: PRISMA 2020 for Abstracts checklist.** Every item is assessed against the structured abstract; NR identifies an unreported item, not an affirmative finding.

| Item | Reporting topic | Status | Manuscript location and assessment |
|---|---|---|---|
| 1 | Databases and platforms | R | Main p. 8; PubMed and PMC identified separately. |
| 2 | Joint database searching | NA | No joint multi-database platform search is included in this analysis. |
| 3 | Registries | NA | No registry contributes to the current analysis. |
| 4 | Other online resources | NA | No additional website/repository search contributes to the current search yield. |
| 5 | Citation searching | P | Previous-review study tables were checked for contextual eligibility notes; not an added search yield. |
| 6 | Contacts | NR | Author/expert/manufacturer search contacts are not documented. |
| 7 | Other search methods | NR | No additional case-finding procedure is documented. |
| 8 | Exact search strategies | P | Main Appendix A contains the recorded expression; exact run syntax for each platform is unavailable. |
| 9 | Limits | P | Main p. 8; no language restriction specified; all platform limits are not documented. |
| 10 | Search filters | NR | Use or absence of a published search filter is not documented. |
| 11 | Adapted strategies | NR | Source/adaptation of a prior search strategy is not documented. |
| 12 | Search updates | NA | No later update is incorporated in the current analysis. |
| 13 | Search dates | R | Main p. 8; date provided for each included resource. |
| 14 | Search peer review | NR | No search peer-review process is documented. |
| 15 | Records per source | R | Main Figure 1: PMC 1,640; PubMed 74. |
| 16 | Deduplication | P | Main p. 8 and Figure 1; linkage reconstructed from identifiers/titles, original software workflow incompletely documented. |

**Figure S5: PRISMA-S search-reporting checklist.** Assessment is restricted to the PMC and PubMed searches contributing to the current analysis. Prompts are paraphrased from the 16-item PRISMA-S checklist.

| Study or Subgroup | Saffron Mean [%] | Saffron SD [%] | Saffron Total | Control Mean [%] | Control SD [%] | Control Total | Weight | Mean Difference IV, Fixed, 95% CI |
|---|---|---|---|---|---|---|---|---|
| Behrouz 2020 | 7.36 | 1.47 | 22 | 7.86 | 1.75 | 23 | 2.3% | -0.50 [-1.44, 0.44] |
| Ebrahimi 2019 | 7.69 | 1.49 | 40 | 7.34 | 1.48 | 40 | 4.8% | 0.35 [-0.30, 1.00] |
| Giannoulaki 2024 | 6.516 | 0.7 | 31 | 7.079 | 0.8 | 30 | 14.2% | -0.56 [-0.94, -0.19] |
| Jaafarinia 2022 | 7.03 | 0.83 | 21 | 7.87 | 1.69 | 19 | 2.9% | -0.84 [-1.68, -0.00] |
| Kotanidou 2023 | 5.25 | 0.33 | 25 | 5.19 | 0.28 | 24 | 69.4% | 0.06 [-0.11, 0.23] |
| Milajerdi 2018 | 6.75 | 1.28 | 26 | 7.25 | 1.65 | 26 | 3.2% | -0.50 [-1.30, 0.30] |
| Shahbazian 2019 | 8.22 | 1.8 | 32 | 8.37 | 1.4 | 32 | 3.3% | -0.15 [-0.94, 0.64] |
| **Total (95% CI)** | | | **197** | | | **194** | **100.0%** | **-0.08 [-0.22, 0.06]** |

Heterogeneity: Chi² = 15.53, df = 6 (P = 0.02); I² = 61%
Test for overall effect: Z = 1.07 (P = 0.28)

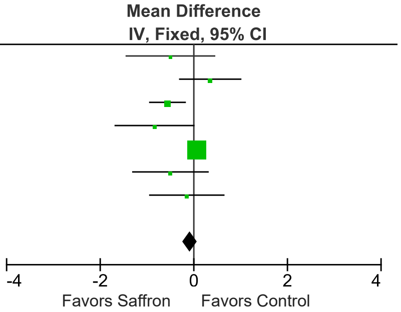


**Figure S6: Fixed-effect sensitivity analysis of HbA1c.** Mean difference −0.08 percentage points (95% CI −0.22 to 0.06).

| | Saffron | | | Control | | | | Mean Difference |
|---|---|---|---|---|---|---|---|---|
| Study or Subgroup | Mean [mg/dL] | SD [mg/dL] | Total | Mean [mg/dL] | SD [mg/dL] | Total | Weight | IV, Fixed, 95% CI |
| Abedimanesh 2017 | 100.85 | 26.49 | 25 | 98.26 | 23.72 | 25 | 9.3% | 2.59 [-11.35, 16.53] |
| Behrouz 2020 | 129.26 | 29.31 | 22 | 160.18 | 57.34 | 23 | 2.6% | -30.92 [-57.36, -4.48] |
| Ebrahimi 2019 | 162.1 | 52.7 | 40 | 147.7 | 51.8 | 40 | 3.4% | 14.40 [-8.50, 37.30] |
| Giannoulaki 2024 | 136.8 | 48.8 | 31 | 147.6 | 61.6 | 30 | 2.3% | -10.80 [-38.75, 17.15] |
| Jaafarinia 2022 | 146.24 | 49.62 | 21 | 159.21 | 62.62 | 19 | 1.4% | -12.97 [-48.23, 22.29] |
| Javandoost 2017 | 88.5 | 19.26 | 21 | 100.5 | 22.22 | 22 | 11.7% | -12.00 [-24.41, 0.41] |
| Kermani 2017 | 111.2 | 35.6 | 24 | 129.3 | 75 | 24 | 1.6% | -18.10 [-51.31, 15.11] |
| Kotanidou 2023 | 99.96 | 9.48 | 25 | 98.66 | 10.61 | 24 | 56.5% | 1.30 [-4.34, 6.94] |
| Milajerdi 2018 | 128.84 | 31.86 | 26 | 153.76 | 41.23 | 26 | 4.5% | -24.92 [-44.95, -4.89] |
| Rajabi 2022 | 172.64 | 9.03 | 8 | 219.17 | 50.69 | 8 | 1.4% | -46.53 [-82.21, -10.85] |
| Rajabi 2024 | 166.07 | 25 | 11 | 214.29 | 28.57 | 11 | 3.6% | -48.22 [-70.65, -25.79] |
| Shahbazian 2019 | 147.9 | 53.5 | 32 | 188.5 | 74.7 | 32 | 1.8% | -40.60 [-72.43, -8.77] |
| **Total (95% CI)** | | | **286** | | | **284** | **100.0%** | **-5.67 [-9.91, -1.43]** |

Heterogeneity: Chi² = 42.53, df = 11 (P < 0.0001); I² = 74%
Test for overall effect: Z = 2.62 (P = 0.009)

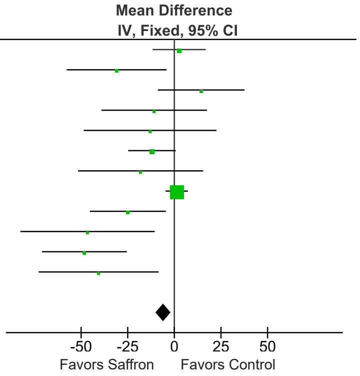


**Figure S7: Fixed-effect sensitivity analysis of fasting glucose.** Mean difference −5.67 mg/dL (95% CI −9.91 to −1.43).

| | Saffron | | | Control | | | | Mean Difference |
|---|---|---|---|---|---|---|---|---|
| Study or Subgroup | Mean | SD | Total | Mean | SD | Total | Weight | IV, Fixed, 95% CI |
| Behrouz 2020 | 4.34 | 1.95 | 22 | 5.86 | 2.43 | 23 | 9.3% | -1.52 [-2.80, -0.24] |
| Ebrahimi 2019 | 1.89 | 1.03 | 40 | 1.8 | 1.22 | 40 | 62.6% | 0.09 [-0.40, 0.58] |
| Kotanidou 2023 | 5.02 | 2.85 | 25 | 5.83 | 2.92 | 24 | 5.9% | -0.81 [-2.43, 0.81] |
| Rajabi 2022 | 2.8 | 0.32 | 8 | 4.96 | 1.85 | 8 | 9.1% | -2.16 [-3.46, -0.86] |
| Rajabi 2024 | 3.14 | 1.37 | 11 | 4.97 | 1.2 | 11 | 13.2% | -1.83 [-2.91, -0.75] |
| **Total (95% CI)** | | | **106** | | | **106** | **100.0%** | **-0.57 [-0.96, -0.18]** |

Heterogeneity: Chi² = 20.02, df = 4 (P = 0.0005); I² = 80%
Test for overall effect: Z = 2.85 (P = 0.004)

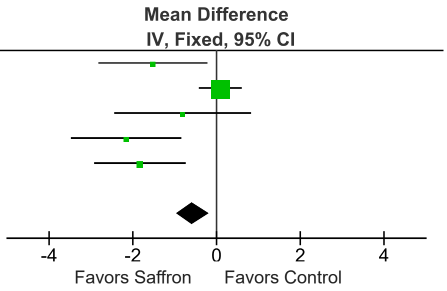


**Figure S8: Fixed-effect sensitivity analysis of HOMA-IR.** Mean difference −0.57 (95% CI −0.96 to −0.18).

**Table S1:** Notable study and comparison decisions in the current synthesis.

| Study | Included / excluded | Disputed screening step | Comparison, omitted arms, or data issue |
|---|---|---|---|
| Giannoulaki 2024[3] | Included | I2: isolated effect | Matched probiotic/prebiotic/magnesium background; saffron is the differing component. |
| Rajabi 2022[4] | Included (selected arms) | I2; contrast selection | Saffron-only vs placebo-only retained. Both exercise-containing arms omitted; an exercise-matched contrast is a separate question. |
| Rajabi 2024[5] | Included (selected arms) | I2; contrast selection | Saffron-only vs placebo-only retained. Both exercise-containing arms omitted; do not reject the whole trial because it contains exercise arms. |
| Abedimanesh 2017[6] | Included (selected arm) | I2; shared-control analysis | Saffron vs placebo retained; the crocin arm was not used. Additional constituent comparisons require control for shared-placebo reuse. |
| Kotanidou 2023[7] | Included (selected arms) | I2; active-comparator analysis | Saffron vs placebo retained; the metformin arm was not used. Metformin is an alternative comparator, not unequal background in the retained contrast. |
| Ebrahimi 2019[8] | Included (one cohort) | D1: report/cohort linkage | Metabolic and organ-function publications may concern the same registered cohort; link reports and count participants once. |
| Shahbazian 2019 / Moravej Aleali 2019[9, 10] | Included (one cohort) | D1: report/cohort linkage | Companion publications inform one retained trial; avoid duplicating the cohort. |
| Javandoost 2017[11] | Included | D3: usable outcome data | Fasting glucose medians/IQRs approximated as means/SDs. Reported dose differs between abstract and Methods. |
| Azimi 2014[12] | Excluded from synthesis; eligibility disputed | I2: isolated effect | Black tea in both arms can isolate added saffron. The recorded not-sole-intervention label warrants reassessment. |
| Saghafi 2025[13] | Excluded from synthesis; eligibility disputed | I2; O1/E1: outcomes | Crocin vs placebo on matched metformin background. PCOS and common metformin do not themselves establish exclusion; verify glycemic endpoint summaries. |
| Sepahi 2018[14] | Excluded from synthesis; eligibility disputed | T1: randomized human trial; O1 | Randomized placebo-controlled diabetic maculopathy trial with glycemic secondary outcomes; the recorded not-clinical-trial label is questionable. |
| Sepahi 2022[15] | Excluded from current synthesis; eligibility unresolved | I2; D3; shared control | Saffron/crocin/placebo arms share a control. Obtain usable data and select/combine comparisons without duplicating control. |
| Tajaddini 2023[16] | Excluded from current synthesis; eligibility unresolved | D1; D3: cohort and data | Glycemic trial included by previous reviews; verify arm-level extraction and cohort identity. No confirmed exclusion reason recorded. |

Notable study decisions, continued

| Study | Included / excluded | Disputed screening step | Comparison, omitted arms, or data issue |
|---|---|---|---|
| Tajaddini 2021 / Mobasseri 2020[17, 18] | Excluded from current synthesis; eligibility unresolved | D1; D3: cohort and data | Possible companion reports. Compare registration, recruitment, and participants; investigator overlap alone does not establish duplication. |
| Hooshmand Moghadam 2022[19] | Excluded from current synthesis; eligibility unresolved | I2; D3: contrast and data | Exercise-factorial design may allow saffron-only and exercise-matched comparisons. Inspect all arms and shared controls. |
| Rahimi 2022[20] | Excluded from current synthesis; eligibility unresolved | O1/E1; D3: glycemic data | PCOS is within broad human eligibility. Verify extractable glucose/insulin endpoints rather than exclude on diagnosis alone. |
| Fadai 2014[21] | Excluded from current synthesis; eligibility unresolved | I2; D3: background and arms | Matched olanzapine background and saffron/crocin/placebo arms warrant contrast-specific review; psychiatric diagnosis alone is not an exclusion. |
| Ghaderi 2019[22] | Excluded from current synthesis; eligibility unresolved | I2; D3: background and data | Matched methadone-maintenance background may isolate crocin; verify outcome data and publication status. |
| Mohamadpour 2013[23] | Excluded from current synthesis; eligibility unresolved | D2: duration; D3: precision | One-month healthy-volunteer crocin study. Confirm at least four weeks, relevant endpoint values, and SD versus SE. |

Included/excluded refers to representation in the current quantitative synthesis; it does not convert unresolved reports into confirmed eligibility exclusions. D1: duplicate/report linkage; I2: isolated intervention effect; T1: randomized human trial; O1: relevant outcome; E1: endpoint measurement; D2: minimum duration; D3: usable outcome data. Contrast selection and shared-control handling are analysis decisions following screening. The full decision workflow is in Supplementary Figure S10. No additional comparison is added to the pooled results by this table.

### Complete screening decisions

Supplementary Table S2 is supplied as a separate landscape appendix, `screening-supplement.pdf`, containing the recorded reviewer assessments retained for PDF presentation. The complete 1,668-paper electronic CSV and workbook/row audit accompany the appendix.

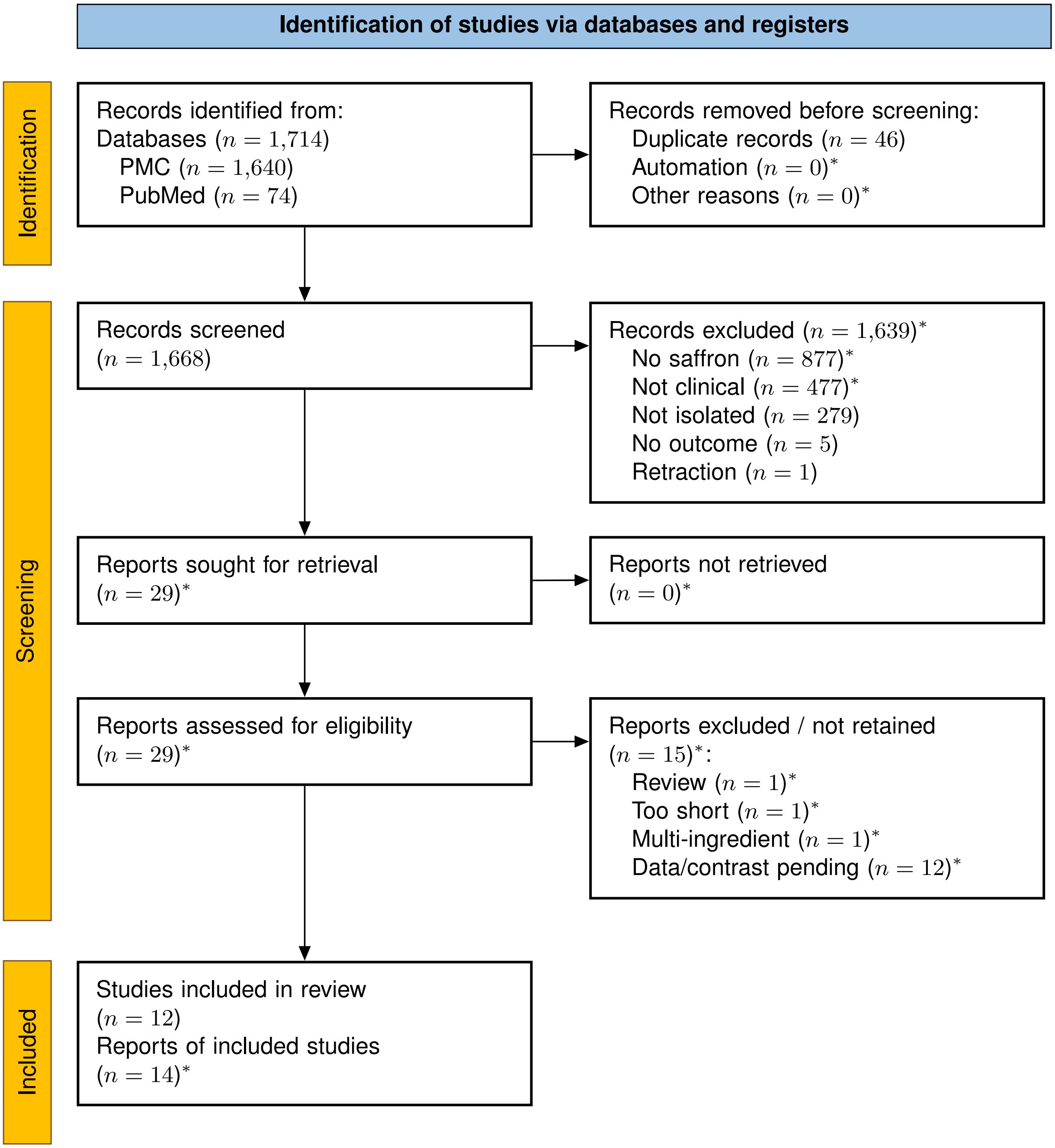


**Figure S9: PRISMA 2020 study-selection flowchart.** Selection flow for the current PMC/PubMed analysis, also shown as main Figure 1. Asterisks identify reconstructed stage counts; the assumptions remain subject to verification.

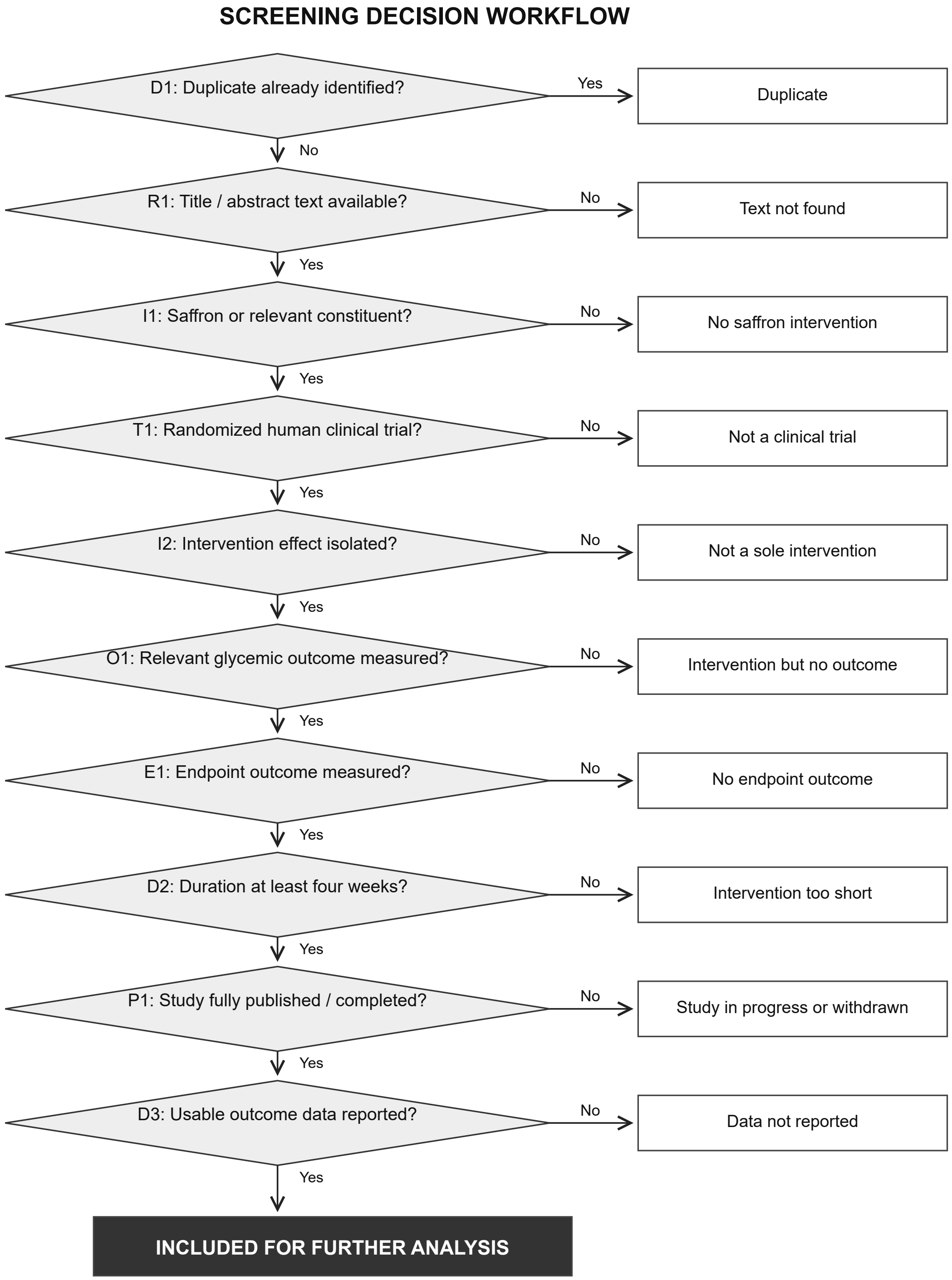


**Figure S10: Screening decision workflow.** Consolidated from the supplied five-page decision tree and adapted to saffron. D1: duplicate/report linkage; R1: text availability; I1: intervention identity; T1: randomized human trial; I2: isolated intervention effect; O1: relevant outcome; E1: endpoint measurement; D2: duration; P1: completed/published study; D3: usable data. Matched background treatment and selection of multi-arm comparisons require interpretation at I2 and at subsequent analysis. The source tree's illustrative percentages are not search results and are omitted.

### Participant accounting

The diagram adapts the official CONSORT 2025 flow template to the included trial reports and selected glycemic contrasts.[24, 25] Source reports are Abedimanesh 2017, Behrouz 2020, Ebrahimi 2019, Giannoulaki 2024, Jaafarinia 2022, Javandoost 2017, Kermani 2017, Kotanidou 2023, Milajerdi 2018, Rajabi 2022, Rajabi 2024, and Shahbazian 2019.[3–9, 11, 26–29]

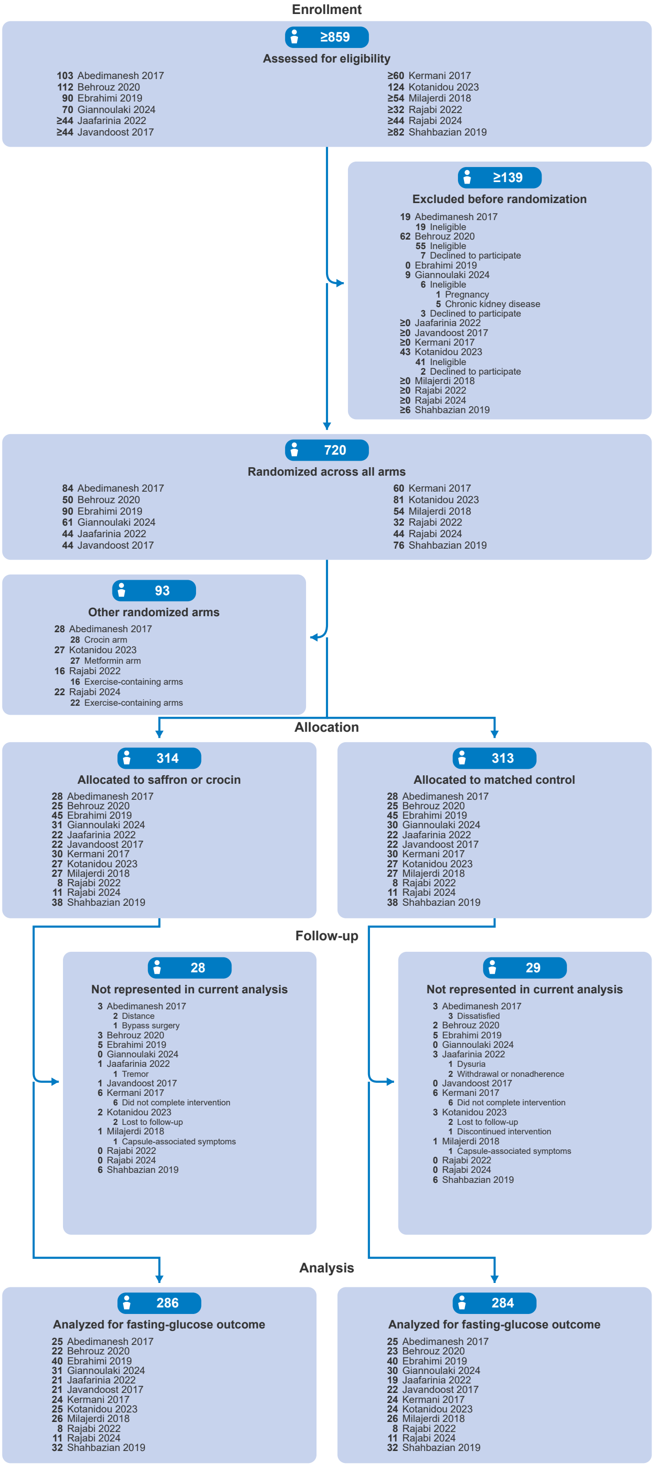


**Figure S11: Participant flow across the included trials.** Trials follow the RevMan fasting-glucose order. Blue badges give summed counts or bounds; bold, right-aligned trial counts precede regular descriptions. Only reported subcategories are indented. Ranges are logical bounds, and $\geq$ denotes an open upper bound. Side branches are mutually exclusive differences between allocation and analysis; reported reasons are nested where available. Analysis counts use the current RevMan inputs: 286 intervention participants and 284 control participants, totaling 570. Table S3 provides the source qualifications.

### Participant-flow sources and bounds

**Table S3:** CONSORT-style cross-trial participant accounting and source qualifications.

| Trial / order | Assessed; randomized | Selected allocated / analyzed (T; C) | Source findings and limits |
|---|---|---|---|
| 1. Abedimanesh 2017 | 103; 84 | 28/28; 25/25 | Assessed 103; 19 ineligible. Saffron and placebo selected (28 each); crocin arm 28 randomized/25 analyzed omitted. Selected discontinuations: saffron 2 distance/1 bypass surgery; placebo 3 dissatisfied. |
| 2. Behrouz 2020 | 112; 50 | 25/25; 22–23/22–23 | Assessed 112; 55 ineligible and 7 declined. Source flow: 25/25 allocated, 2/3 lost, 23/22 analyzed (crocin/control). RevMan: 22/23. Analysis slots show 22-23 bounds, with combined n fixed at 45. No count correction made to meta-analysis. |
| 3. Ebrahimi 2019 | 90; 90 | 45/45; 40/40 | 90 screened/randomized, 45 each; 5 each dropped out. Reasons include personal reasons, insulin initiation, migration, stomach symptoms, without reason-specific counts. All 80 completers reportedly consumed assigned powders. |
| 4. Giannoulaki 2024 | 70; 61 | 31/30; 31/30 | 70 assessed; 6 ineligible (pregnancy 1, CKD 5), 3 refused; 31/30 randomized and analyzed; zero losses/discontinuations explicitly reported. |
| 5. Jaafarinia 2022 | ≥44; 44 | 22/22; 21/19 | 44 randomized 22 each; 1 crocin tremor exclusion, 1 placebo dysuria exclusion and 2 placebo follow-up/noncompliance withdrawals; analyzed 21/19. Total screened not recovered from available text. |
| 6. Javandoost 2017 | ≥44; 44 | 22/22; 21/22 | 44 randomized 22 each; RevMan endpoint n 21/22. Text does not establish the missing treatment participant disposition. Screened total not recovered from text. Abstract 30 mg/day versus Methods 30 mg twice daily. |
| 7. Kermani 2017 | ≥60; 60 | 30/30; 24/24 | 60 randomized; 48 completed (24 each), with 6 per arm not completing the treatment period. Screened total not stated in available text. |
| 8. Kotanidou 2023 | 124; 81 | 27/27; 25/24 | 124 assessed; 41 ineligible, 2 declined; 81 randomized (27 each). Saffron/control receipt 27/27. Saffron 2 lost/0 discontinued; placebo 2 lost/1 discontinued. Metformin arm 27 randomized/25 analyzed omitted. |
| 9. Milajerdi 2018 | ≥54; 54 | 27/27; 26/26 | 54 randomized (27 each); 1 per arm left due capsule-associated symptoms, particularly headache; analyzed 26 each. Screened total not recovered from available text. |
| 10. Rajabi 2022 | ≥32; 32 | 8/8; 8/8 | 32 participants allocated among 4 groups of 8; selected saffron-only/placebo-only groups 8 each. Exercise groups 16 randomized and reported in full trial but omitted from selected contrast. Attrition stages not separately reported. |

Table S3 continued

| Trial / order | Assessed; randomized | Selected allocated / analyzed (T; C) | Source findings and limits |
|---|---|---|---|
| 11. Rajabi 2024 | ≥44; 44 | 11/11; 11/11 | 44 participants allocated among 4 groups of 11; selected saffron-only/placebo-only groups 11 each. Exercise groups 22 omitted from selected contrast. Attrition stages not separately reported. |
| 12. Shahbazian 2019 | ≥82; 76 | 38/38; 32/32 | 82 eligible, 76 randomized 38 each; 64 completed (32 each). Six per arm dropped out for moving, contact loss, allergy, or insulin use, without reason-specific counts. Number assessed may exceed 82 eligible. |

Where actual receipt or discontinuation is not separately recoverable, conservative bounds are zero to allocated. Consumption reported for Ebrahimi completers and explicit Kotanidou receipt narrow the receipt bounds; outcome completion alone is not treated as proof of treatment receipt. Unknown eligibility-screening totals have open upper bounds. Follow-up loss bounds use available outcome denominators, with reported losses retained. The diagram is an adapted review-level accounting display, not a CONSORT-compliance assessment or a report of one combined trial. Counts in different stages can overlap and should not be added as independent reasons. The primary-outcome label in the original template is adapted to fasting glucose because all 12 selected comparisons contribute this outcome; only seven contribute review-primary HbA1c data.

[4] Ali Rajabi, Mojdeh Khajehlandi, Marefat Siahkuhian, Ali Akbarnejad, Kayvan Khoramipour, and Katsuhiko Suzuki. Effect of 8 Weeks Aerobic Training and Saffron Supplementation on

Inflammation and Metabolism in Middle-Aged Obese Women with Type 2 Diabetes Mellitus. *Sports*, 10(11):167, 2022. doi: 10.3390/sports10110167. URL https://pmc.ncbi.nlm.nih.gov/articles/PMC9697862/.

[20] Giti Rahimi, Saeideh Shams, and Mohammad Reza Aslani. Effects of crocin supplementation on inflammatory markers, lipid profiles, insulin and cardioprotective indices in women with PCOS:

A randomized, double-blind, placebo-controlled trial. *Phytotherapy research : PTR*, 36(6):2605–2615, 2022. doi: 10.1002/ptr.7474. URL https://pubmed.ncbi.nlm.nih.gov/35470916/.

[28] Tayyebe Kermani, Toba Kazemi, Somayye Molki, Khadije Ilkhani, Gholamreza Sharifzadeh, and Omid Rajabi. The Efficacy of Crocin of Saffron (Crocus sativus L.) on the Components of Metabolic Syndrome: A Randomized Controlled Clinical Trial. *Journal of Research in*

*Pharmacy Practice*, 6(4):228–232, 2017. doi: 10.4103/jrpp.JRPP_17_26. URL https://pmc.ncbi.nlm.nih.gov/articles/PMC5787909/.

[29] Alireza Milajerdi, Shima Jazayeri, Najmeh Hashemzadeh, Elham Shirzadi, Zhaleh Derakhshan, Abolghassem Djazayeri, and Shahin Akhondzadeh. The effect of saffron (Crocus sativus L.) hydroalcoholic extract on metabolic control in type 2 diabetes mellitus: A triple-blinded randomized clinical trial. *Journal of Research in Medical Sciences : The Official Journal of Isfahan University of Medical Sciences*, 23:16, 2018. doi: 10.4103/jrms.JRMS_286_17. URL https://pmc.ncbi.nlm.nih.gov/articles/PMC5842443/.
