## Supplementary Table S2 for "Effects of saffron and crocin on glycemic control: an exploratory systematic review and meta-analysis of randomized trials"

### Supplementary Table S2: recorded screening decisions

This appendix lists 1,625 papers with recorded reviewer decisions from the current PMC/PubMed materials. The paper IDs retain their numbering in the complete 1,668-paper electronic table. Historical Joseph and Paul workbook labels are displayed as Juno Kim and Bryan Anh Nguyen. Candidate means marked for further analysis, not automatically eligible. Additional actual reviews appear in the final column. Source workbook and row references are supplied in the electronic audit.

Table S2. Recorded reviewer assessments.

| Paper | Reviewer 1 | Reviewer 2 | Additional recorded reviews |
|---|---|---|---|
| 5. Mohamed 2025. Effectiveness of Calorie Restriction for Weight Loss in Type 2 Diabetes Mellitus: A Systematic Review<br>PMID 40034634 | Michele Schaffer: Not clinical / No saffron | Prisha Batra: Not clinical | – |
| 6. Zhang 2025b. Effects of Saffron Supplementation on Glycolipid Metabolism and Blood Pressure in Patients With Metabolic Syndrome and Related Disorders: A Systematic Review and Meta-Analysis of Randomized Controlled Trials<br>PMID 39931766 | Michele Schaffer: Not clinical | Prisha Batra: Not clinical | – |
| 9. Karim 2024. Effects of saffron (Crocus sativus L.) supplementation on cardiometabolic Indices in diabetic and prediabetic overweight patients: a systematic review and meta-analysis of RCTs<br>PMID 39593185 | Michele Schaffer: Not clinical | Bryan Anh Nguyen: Not clinical | – |
| 11. Mamri 2024. Comprehensive analysis of bioactive compounds in Crocus sativus stamens extracts by HPLC-DAD: investigating antidiabetic activity via in vitro, in vivo, and molecular docking simulation<br>PMID 39072261 | Michele Schaffer: Not clinical | Prisha Batra: Not clinical | – |
| 12. Giannoulaki 2024. Effect of Crocus sativus Extract Supplementation in the Metabolic Control of People with Diabetes Mellitus Type 1: A Double-Blind Randomized Placebo-Controlled Trial<br>PMID 38999837 | Michele Schaffer: Not isolated | Prisha Batra: Candidate | – |
| 13. Rajabi 2024. The effect of 12 weeks of aerobic exercise training with or without saffron supplementation on diabetes-specific markers and inflammation in women with type 2 diabetes: A randomized double-blind placebo-controlled trial<br>PMID 38874882 | Michele Schaffer: Not isolated | Prisha Batra: Candidate | – |
| 14. Yan 2024. Effects of Crocus sativus on glycemic control and cardiometabolic parameters among patients with metabolic syndrome and related disorders: a systematic review and meta-analysis of randomized controlled trials<br>PMID 38796446 | Michele Schaffer: Not clinical | Prisha Batra: Not clinical | – |

Table S2 continued

| Paper | Reviewer 1 | Reviewer 2 | Additional recorded reviews |
|---|---|---|---|
| 15. Liu 2024. Effect of saffron supplementation on the glycemic outcomes in diabetes: a systematic review and meta-analysis<br>PMID 38559777 | Michele Schaffer: Not isolated | Prisha Batra: Not clinical | – |
| 16. Garza 2024. Effect of Aromatic Herbs and Spices Present in the Mediterranean Diet on the Glycemic Profile in Type 2 Diabetes Subjects: A Systematic Review and Meta-Analysis<br>PMID 38542668 | Michele Schaffer: Not isolated | Prisha Batra: Not clinical | – |
| 17. Lv 2024. Extract of Gardenia jasminoides Ellis Attenuates High-Fat Diet-Induced Glycolipid Metabolism Disorder in Rats by Targeting Gut Microbiota and TLR4/Myd88/NF-$\kappa$B Pathway<br>PMID 38539827 | Michele Schaffer: Not isolated | Prisha Batra: Not clinical | – |
| 19. Asdaq 2024. Antidiabetic and antioxidant potential of Crocin in high-fat diet plus streptozotocin-induced type-2 diabetic rats<br>PMID 38233742 | Michele Schaffer: Not clinical | Prisha Batra: Not clinical | – |
| 20. Kotanidou 2023. The Effect of Saffron Kozanis (Crocus sativus L.) Supplementation on Weight Management, Glycemic Markers and Lipid Profile in Adolescents with Obesity: A Double-Blinded Randomized Placebo-Controlled Trial<br>PMID 38002905 | Juno Kim: Candidate | Michele Schaffer: Not clinical | Prisha Batra: Candidate |
| 22. Zhang 2023. The effects of medicinal and food homologous substances on blood lipid and blood glucose levels and liver function in patients with nonalcoholic fatty liver disease: a systematic review of randomized controlled trials<br>PMID 37644446 | Bryan Anh Nguyen: Not clinical | Pranav Potluri: No saffron | Alexander Tai Nguyen: No saffron |
| 23. Yazdani 2023. Effects of Aqueous Saffron Extract on Glucoregulation as Well as Hepatic Agt and TNF-$\alpha$ Gene Expression in Rats Subjected to Sub-Chronic Stress<br>PMID 37434936 | Bryan Anh Nguyen: No saffron | Alexander Tai Nguyen: No saffron | – |
| 24. Bellachioma 2023. Insights on the Hypoglycemic Potential of Crocus sativus Tepal Polyphenols: An In Vitro and In Silico Study<br>PMID 37298165 | Bryan Anh Nguyen: No saffron | Alexander Tai Nguyen: Not clinical | – |
| 25. Alipour 2023. The Effect of the Saffron Intervention on NAFLD Status and Related Gene Expression in a Rat Model<br>PMID 37180863 | Bryan Anh Nguyen: No saffron | Alexander Tai Nguyen: Not clinical | – |
| 26. Radmehr 2023. Protective effects of crocin and gallic acid on the liver damage induced by methylglyoxal in male mice: role of inflammatory factors<br>PMID 37070111 | Bryan Anh Nguyen: Not clinical | Alexander Tai Nguyen: Not clinical | – |

Table S2 continued

| Paper | Reviewer 1 | Reviewer 2 | Additional recorded reviews |
|---|---|---|---|
| 28. Diaz 2023. Maximizing Glycemic Benefits of Using Faster Insulin Formulations in Type 1 Diabetes: In Silico Analysis Under Open- and Closed-Loop Conditions PMID 36595379 | Michele Schaffer: No saffron | Prisha Batra: No saffron | – |
| 30. Zamani 2022. The effects of saffron supplementation on cardiovascular risk factors in adults: A systematic review and dose-response meta-analysis PMID 36570145 | Michele Schaffer: Not clinical | Prisha Batra: Not clinical | – |
| 31. Radmehr 2022. Crocin attenuates endoplasmic reticulum stress in methylglyoxal-induced diabetic nephropathy in male mice: MicroRNAs alterations and glyoxalase 1-Nrf2 signaling pathways PMID 36474578 | Michele Schaffer: Not isolated | Prisha Batra: Not clinical | – |
| 32. Rajabi 2022. Effect of 8 Weeks Aerobic Training and Saffron Supplementation on Inflammation and Metabolism in Middle-Aged Obese Women with Type 2 Diabetes Mellitus PMID 36355818 | Michele Schaffer: Not isolated | Bryan Anh Nguyen: Candidate | – |
| 35. Radmehr 2022. Crocin ameliorates MicroRNAs-associated ER stress in type 2 diabetes induced by methylglyoxal PMID 35655590 | Grace Kim: Not clinical | Pranav Potluri: Not clinical | – |
| 44. Behrouz 2020. The effect of crocin supplementation on glycemic control, insulin resistance and active AMPK levels in patients with type 2 diabetes: a pilot study PMID 32670418 | Grace Kim: Candidate | Alexander Tai Nguyen: Candidate | – |
| 45. Qiu 2020. The Hypoglycemic and Renal Protection Properties of Crocin via Oxidative Stress-Regulated NF-$\kappa$B Signaling in db/db Mice PMID 32425787 | Grace Kim: Not clinical | Alexander Tai Nguyen: Not clinical | – |
| 46. Giannoulaki 2020. Impact of Crocus Sativus L. on Metabolic Profile in Patients with Diabetes Mellitus or Metabolic Syndrome: A Systematic Review PMID 32423173 | Grace Kim: Not clinical | Alexander Tai Nguyen: No saffron | – |
| 48. Algandaby 2020. Crocin prevents metabolic syndrome in rats via enhancing PPAR-gamma and AMPK PMID 32346340 | Grace Kim: Not clinical | Alexander Tai Nguyen: Not clinical | – |
| 50. Malekzadeh 2019. Effect of safranal, a constituent of saffron, on olanzapine (an atypical antipsychotic) induced metabolic disorders in rat PMID 32133067 | Grace Kim: Not clinical | Alexander Tai Nguyen: Not clinical | – |

Table S2 continued

| Paper | Reviewer 1 | Reviewer 2 | Additional recorded reviews |
|---|---|---|---|
| 51. Raafat 2019. Phytochemical and anti-neuropathic investigations of Crocus sativus via alleviating inflammation, oxidative stress and pancreatic beta-cells regeneration<br>PMID 36117567 | Bryan Anh Nguyen: Not clinical | Prisha Batra: Not clinical | – |
| 52. Ebrahimi 2019. Effects Of Saffron Supplementation On Inflammation And Metabolic Responses In Type 2 Diabetic Patients: A Randomized, Double-Blind, Placebo-Controlled Trial<br>PMID 31686882 | Grace Kim: Candidate | Alexander Tai Nguyen: Candidate | – |
| 54. Hashemian 2019. Turmeric, Pepper, Cinnamon, and Saffron Consumption and Mortality<br>PMID 37221812 | Grace Kim: Not clinical | Alexander Tai Nguyen: No saffron | – |
| 57. Motamedrad 2019. The regulatory effect of saffron stigma on the gene expression of the glucose metabolism key enzymes and stress proteins in streptozotocin-induced diabetic rats<br>PMID 31160903 | Grace Kim: Not clinical | Alexander Tai Nguyen: Not clinical | – |
| 59. Luo 2019. Crocin ameliorates hepatic steatosis through activation of AMPK signaling in db/db mice<br>PMID 30621686 | Grace Kim: Not clinical | Bryan Anh Nguyen: Not clinical | Alexander Tai Nguyen: Not clinical |
| 63. Milajerdi 2018. The effect of saffron (Crocus sativus L.) hydroalcoholic extract on metabolic control in type 2 diabetes mellitus: A triple-blinded randomized clinical trial<br>PMID 29531568 | Grace Kim: Candidate | Pranav Potluri: Candidate | – |
| 64. Javandoost 2017. Effect of crocin, a carotenoid from saffron, on plasma cholesteryl ester transfer protein and lipid profile in subjects with metabolic syndrome: A double blind randomized clinical trial<br>PMID 29371871 | Michele Schaffer: Candidate | Grace Kim: Candidate | Bryan Anh Nguyen: Candidate |
| 65. Dehghan 2016. Saffron with resistance exercise improves diabetic parameters through the GLUT4/AMPK pathway in-vitro and in-vivo<br>PMID 27122001 | Grace Kim: Not clinical | Bryan Anh Nguyen: No saffron | – |
| 68. Hemmati 2015. Anti-atherogenic potential of jujube, saffron and barberry: anti-diabetic and antioxidant actions<br>PMID 26600752 | Grace Kim: Not clinical | Bryan Anh Nguyen: Not clinical | – |
| 69. Azimi 2014. Effects of Cinnamon, Cardamom, Saffron, and Ginger Consumption on Markers of Glycemic Control, Lipid Profile, Oxidative Stress, and Inflammation in Type 2 Diabetes Patients<br>PMID 26177486 | Grace Kim: Not isolated | Bryan Anh Nguyen: Not isolated | – |

Table S2 continued

| Paper | Reviewer 1 | Reviewer 2 | Additional recorded reviews |
|---|---|---|---|
| 70. Naderi 2014. Effect of Some High Consumption Spices on Hemoglobin Glycation<br>PMID 25593391 | Michele Schaffer: No saffron | Grace Kim: Not clinical | Bryan Anh Nguyen: Not clinical |
| 75. Papada 2025. Invalid XML: Caught SAXException: org.xml.sax.SAXParseException; lineNumber: 1; columnNumber: 2594; XML document structures must start and end within the same entity.<br>PMID 40296787 | Grace Kim: No saffron | Michele Schaffer: Not clinical / No saffron | Bryan Anh Nguyen: Not clinical / No saffron; Prisha Batra: No saffron / Not clinical |
| 76. Chaker 2025. Characterizing Genetic, Epigenetic, Nutritional, and Clinico-Biochemical Profile of Women With Polycystic Ovary Syndrome: A Case–Control Study<br>PMID 40486662 | Michele Schaffer: Not clinical | Prisha Batra: Not clinical | – |
| 77. Grifoni 2025. The Dark Triad of Particulate Matter, Oxidative Stress and Coronary Artery Disease: What About the Antioxidant Therapeutic Potential<br>PMID 40427454 | Michele Schaffer: Not clinical | Prisha Batra: Not clinical | – |
| 78. Lemoni 2025. Kinetic Modeling and Biological Activities of Rosa canina L. Pseudo-Fruit Extracts Obtained via Enzyme-Assisted Extraction<br>PMID 40427440 | Michele Schaffer: Not clinical | Prisha Batra: Not clinical | – |
| 79. Tian 2025. Curcuma Longa (turmeric): from traditional applications to modern plant medicine research hotspots<br>PMID 40437595 | Michele Schaffer: Not clinical | Prisha Batra: Not clinical | – |
| 80. Carboni 2025. A Comprehensive Review of Edible Flowers with a Focus on Microbiological, Nutritional, and Potential Health Aspects<br>PMID 40428498 | Michele Schaffer: Not clinical | Prisha Batra: Not clinical | – |
| 81. Wang 2025. Bioinformatics analysis combined with experimental validation reveals the novel mechanisms of multi-targets of dapagliflozin attenuating diabetic liver injury<br>PMID 40438398 | Michele Schaffer: Not clinical | Prisha Batra: Not clinical | – |
| 82. Ayad 2025. Propolis Stands out as a Multifaceted Natural Product: Meta-Analysis on Its Sources, Bioactivities, Applications, and Future Perspectives<br>PMID 40430191 | Michele Schaffer: Not clinical | Prisha Batra: Not clinical | – |
| 83. Chen 2025. Development and Validation of the China Dietary Inflammatory Index (CHINA-DII)<br>PMID 40431426 | Michele Schaffer: Not clinical | Prisha Batra: Not clinical | – |

Table S2 continued

| Paper | Reviewer 1 | Reviewer 2 | Additional recorded reviews |
|---|---|---|---|
| 84. Centonze 2025. The Antiaging Potential of Dietary Plant-Based Polyphenols: A Review on Their Role in Cellular Senescence Modulation<br>PMID 40431456 | Michele Schaffer: Not clinical | Prisha Batra: Not clinical | – |
| 85. Vázquez-Lorente 2025. Low-grade systemic inflammation biomarkers in sedentary young healthy adults are not significantly affected by a 24-week concurrent training intervention<br>PMID 40219791 | Michele Schaffer: Not clinical | Prisha Batra: No saffron | – |
| 86. Yu 2025. Association between dietary inflammatory index score and cardiovascular-kidney-metabolic syndrome: a cross-sectional study based on NHANES<br>PMID 40416382 | Michele Schaffer: Not clinical | Prisha Batra: Not clinical | – |
| 87. Ahmadi 2025. Impact of jujube fruit on serum lipid profile, glycemic index, and liver function: a systematic review and meta-analysis<br>PMID 40399274 | Michele Schaffer: Not clinical | Prisha Batra: Not clinical | – |
| 88. Wen 2025. The Therapeutic Effect and Mechanism of Traditional Chinese Medicine in Type 2 Diabetes Mellitus and Its Complications<br>PMID 40391051 | Michele Schaffer: Not clinical | Prisha Batra: Not clinical | – |
| 89. Bansal 2025. A Systematic Review of Inflammatory Markers in Polycystic Ovary Syndrome (PCOS) and Meta-Analysis of Interleukin-6 (IL-6) in Case-Control Studies<br>PMID 40385768 | Michele Schaffer: Not clinical | Prisha Batra: Not clinical | – |
| 90. Hossain 2025. Flavor and Well-Being: A Comprehensive Review of Food Choices, Nutrition, and Health Interactions<br>PMID 40384991 | Michele Schaffer: Not clinical | Prisha Batra: Not clinical | – |
| 91. Zhu 2025. Effect of Elaeagnus angustifolia Honey in the Protection Against Ethanol-Induced Chronic Gastric Injury via Counteracting Oxidative Stress, Interfering with Inflammation and Regulating Gut Microbiota in Mice<br>PMID 40361682 | Michele Schaffer: Not clinical | Prisha Batra: Not clinical | – |
| 92. Perrone 2025. Editorial for a New Section: Nutrition and Neuro Sciences<br>PMID 40362708 | Michele Schaffer: Not clinical | Prisha Batra: Not clinical | – |
| 93. Liu 2025. Genetic Variants, Bioactive Compounds, and PCSK9 Inhibitors in Hyper-LDL-Cholesterolemia: A GWAS and In Silico Study on Cardiovascular Disease Risk<br>PMID 40362720 | Michele Schaffer: Not clinical | Prisha Batra: Not clinical | – |

Table S2 continued

| Paper | Reviewer 1 | Reviewer 2 | Additional recorded reviews |
|---|---|---|---|
| 94. Yaglova 2025. Altering the Hydrogen Isotopic Composition of the Essential Nutrient Water as a Promising Tool for Therapy: Perspectives and Risks<br>PMID 40362685 | Michele Schaffer: Not clinical | Prisha Batra: Not clinical | – |
| 95. Hasan 2025. Efficacy of flaxseed in reducing blood pressure among patients with cardiovascular risk factors: A systematic review and meta-analysis of parallel randomized controlled trials<br>PMID 40365516 | Michele Schaffer: Not clinical | Prisha Batra: Not clinical | – |
| 96. Hanslian 2025. Effects of nutritional counseling on dietary patterns in patients with mild cognitive impairment: insights from the BrainFit-Nutrition study<br>PMID 40357042 | Michele Schaffer: Not clinical | Prisha Batra: Not clinical | – |
| 97. Okita 2025. Acute kaempferol ingestion lowers oxygen uptake during submaximal exercise and improves high-intensity exercise capacity in well-trained male athletes<br>PMID 40346019 | Michele Schaffer: Not clinical | Prisha Batra: No saffron | – |
| 98. Ghasemzadeh Rahbardar 2025. Assessing the efficacy of herbal supplements for managing obesity: A comprehensive review of global clinical trials<br>PMID 40343290 | Michele Schaffer: Not clinical | Prisha Batra: Not clinical | – |
| 99. Huang 2025. Reduced composite dietary antioxidant index increases the risk of Parkinson's disease and all-cause mortality in Parkinson's disease patients: evidence from the NHANES database<br>PMID 40330594 | Michele Schaffer: Not clinical | Prisha Batra: Not clinical | – |
| 100. Irandoost 2025. The effects of an anti-inflammatory diet alone or in combination with acupuncture on mental health, anthropometric indices, and metabolic status in diabetic patients with depression: a randomized, controlled clinical trial<br>PMID 40316554 | Michele Schaffer: Not isolated | Prisha Batra: No saffron | – |
| 101. Conjard-Duplany 2025. Muscle mTOR controls iron homeostasis and ferritinophagy via NRF2, HIFs and AKT/PKB signaling pathways<br>PMID 40293459 | Michele Schaffer: Not clinical | Prisha Batra: Not clinical | – |
| 102. Hu 2025. Explore potential immune-related targets of leeches in the treatment of type 2 diabetes based on network pharmacology and machine learning<br>PMID 40296871 | Michele Schaffer: Not clinical | Prisha Batra: Not clinical | – |

Table S2 continued

| Paper | Reviewer 1 | Reviewer 2 | Additional recorded reviews |
|---|---|---|---|
| 103. Mijajlović 2025. Secondary stroke prevention beyond antiplatelets: The role of colchicine and GLP-1RA - an ounce of prevention is worth a pound of cure<br>PMID 40291758 | Michele Schaffer: Not clinical | Prisha Batra: Not clinical | – |
| 104. Petkova-Parlapanska 2025. Systematic Inflammation and Oxidative Stress Elevation in Diabetic Retinopathy and Diabetic Patients with Macular Edema<br>PMID 40332476 | Michele Schaffer: Not clinical | Prisha Batra: Not clinical | – |
| 105. Afsar 2024. The impact of sodium-glucose cotransporter inhibitors on gut microbiota: a scoping review<br>PMID 38932911 | Michele Schaffer: Not clinical | Prisha Batra: Not clinical | – |
| 106. Bouzana 2025. Phytochemical Profiling and Biological Evaluation of Dianthus sylvestris subsp. aristidis: A Chromatographic and Mass Spectrometry Approach to Uncovering Bioactive Metabolites for Dermatological and Metabolic Disorder Management<br>PMID 40284013 | Michele Schaffer: Not clinical | Prisha Batra: Not clinical | – |
| 107. Salehsari 2024. Investigating the effect of zeaxanthin on olanzapine-induced metabolic disorders in rats<br>PMID 40259963 | Michele Schaffer: Not clinical | Prisha Batra: Not clinical | – |
| 108. Zhang 2025. A systematical review on traditional Chinese medicine treating chronic diseases via regulating ferroptosis from the perspective of experimental evidence and clinical application<br>PMID 40256717 | Michele Schaffer: Not clinical | Prisha Batra: Not clinical | – |
| 109. Kakoti 2024. AMPK pathway: an emerging target to control diabetes mellitus and its related complications<br>PMID 38932895 | Michele Schaffer: Not clinical | Prisha Batra: Not clinical | – |
| 110. Mirhosseini 2025. Tiryāq in traditional Persian medicine: a survey of antidotal plants and their modern pharmacological potential<br>PMID 40242443 | Michele Schaffer: Not clinical | Prisha Batra: Not clinical | – |
| 111. Marrone 2025. Sustainability and role of plant-based diets in chronic kidney disease prevention and treatment<br>PMID 40230686 | Michele Schaffer: Not clinical | Prisha Batra: Not clinical | – |
| 112. Menon 2025. Beyond Genetics: Exploring Lifestyle, Microbiome, and Social Determinants in Oral Cancer Development<br>PMID 40227635 | Michele Schaffer: Not clinical | Prisha Batra: Not clinical | – |

Table S2 continued

| Paper | Reviewer 1 | Reviewer 2 | Additional recorded reviews |
|---|---|---|---|
| 113. Vallazhath 2025. A comprehensive review on the implications of Yogic/Sattvic diet in reducing inflammation in type 2 diabetes<br>PMID 40216734 | Michele Schaffer: Not clinical | Prisha Batra: Not clinical | – |
| 114. Zhao 2025. Association between dietary inflammatory index and cardiovascular-kidney-metabolic syndrome risk: a cross-sectional study<br>PMID 40221720 | Michele Schaffer: Not clinical | Prisha Batra: Not clinical | – |
| 115. Mercurio 2025. Mitochondria at the Crossroads: Linking the Mediterranean Diet to Metabolic Health and Non-Pharmacological Approaches to NAFLD<br>PMID 40218971 | Michele Schaffer: Not clinical | Prisha Batra: Not clinical | – |
| 116. Tufail 2025. Role of Phytonutrients in the Prevention and Treatment of Chronic Diseases: A Concrete Review<br>PMID 40224418 | Michele Schaffer: Not clinical | Prisha Batra: Not clinical | – |
| 117. Poursamimi 2025. A Review of the Prospective Effects of Methadone on Peripheral Neuropathic Pain in Diabetic Patients<br>PMID 40225355 | Michele Schaffer: Not clinical | Prisha Batra: Not clinical | – |
| 118. Fujisaka 2024. Identification of herbal drug extracts that promote growth of Akkermansia muciniphila in high-fat diet fed mice<br>PMID 39101187 | Michele Schaffer: Not clinical | Prisha Batra: Not clinical | – |
| 119. Abou-El-Naga 2025. Restorative effects of Momordica charantia extract on cerebellar GFAP and NGF expression in pregnant diabetic rats and their offspring<br>PMID 40184394 | Michele Schaffer: Not clinical | Prisha Batra: Not clinical | – |
| 120. Ma 2025. Traditional Chinese Medicine-derived formulations and extracts modulating the PI3K/AKT pathway in Alzheimer's disease<br>PMID 40166467 | Michele Schaffer: Not clinical | Prisha Batra: Not clinical | – |
| 121. Zheng 2025. PPARs in atherosclerosis: The spatial and temporal features from mechanism to druggable targets<br>PMID 38555000 | Michele Schaffer: Not clinical | Prisha Batra: Not clinical | – |
| 122. Xin 2025. The circadian rhythm: A new target of natural products that can protect against diseases of the metabolic system, cardiovascular system, and nervous system<br>PMID 38631431 | Michele Schaffer: Not clinical | Prisha Batra: Not clinical | – |
| 123. Kazemi 2025. Recycling Agricultural Waste: Sustainable Solutions for Enhancing Livestock Nutrition<br>PMID 40159453 | Michele Schaffer: Not clinical | Prisha Batra: Not clinical | – |

Table S2 continued

| Paper | Reviewer 1 | Reviewer 2 | Additional recorded reviews |
|---|---|---|---|
| 124. Chen 2025. Role of oxidative balance score in staging and mortality risk of cardiovascular-kidney-metabolic syndrome: Insights from traditional and machine learning approaches PMID 40073760 | Michele Schaffer: Not clinical | Prisha Batra: Not clinical | – |
| 125. Radwan 2024. Chitosan and Grifola Frondosa nanoparticles insulate liver dysfunction in EAC-bearing mice PMID 38559757 | Michele Schaffer: Not clinical | Prisha Batra: Not clinical | – |
| 126. Moccia 2025. Pharmacological Interventions for Negative Symptoms in Schizophrenia: A Systematic Review of Randomised Control Trials PMID 40149518 | Michele Schaffer: Not clinical | Prisha Batra: Not clinical | – |
| 127. Golpasandi 2025. The Effect of Vitamin D3 Injection Combined With High-Intensity Interval Training on Excessive Autophagy in the Heart Tissue of Type 2 Diabetes-Induced Rats: An Analysis of the mTOR-Beclin-1-Fyco-1-Cathepsin D Pathway PMID 40144137 | Michele Schaffer: Not clinical | Prisha Batra: Not clinical | – |
| 128. Ekwudo 2025. The microbiota-gut-brain axis in Huntington's disease: pathogenic mechanisms and therapeutic targets PMID 38426291 | Michele Schaffer: Not clinical | Prisha Batra: Not clinical | – |
| 129. Macdonald 2024. Molecular analysis of the extracellular microenvironment: from form to function PMID 38509768 | Michele Schaffer: Not isolated | Prisha Batra: Not clinical | – |
| 130. Hossain 2025. Dietary Phytochemicals in Health and Disease: Mechanisms, Clinical Evidence, and Applications-A Comprehensive Review PMID 40115248 | Michele Schaffer: Not isolated | Prisha Batra: Not clinical | – |
| 131. Anwar 2025. Herbal remedies for oral and dental health: a comprehensive review of their multifaceted mechanisms including antimicrobial, anti-inflammatory, and antioxidant pathways PMID 39907951 | Michele Schaffer: Not isolated | Prisha Batra: Not clinical | – |
| 132. Li 2025. Exploring Mechanisms of Lang Qing Ata in Non-Alcoholic Steatohepatitis Based on Metabolomics, Network Pharmacological Analysis, and Experimental Validation PMID 40098906 | Michele Schaffer: Not clinical | Prisha Batra: Not clinical | – |
| 133. Qian 2025. Crosstalk between lipid metabolism and macrophages in atherosclerosis: therapeutic potential of natural products PMID 40099271 | Michele Schaffer: No saffron | Prisha Batra: Not clinical | – |

Table S2 continued

| Paper | Reviewer 1 | Reviewer 2 | Additional recorded reviews |
|---|---|---|---|
| 134. Liu 2025. Gene-Lifestyle Interactions in Renal Dysfunction: Polygenic Risk Modulation via Plant-Based Diets, Coffee Intake, and Bioactive Compound Interactions PMID 40077791 | Michele Schaffer: No saffron | Prisha Batra: Not clinical | – |
| 135. Wu 2025. Exploration of the role of drug resistance-associated anoikis-related genes in HER2-Negative breast cancer through bioinformatics analysis PMID 40065765 | Michele Schaffer: Not isolated | Prisha Batra: Not clinical | – |
| 136. Alharbi 2025. Antioxidative and ameliorative properties of probiotic-enriched fermented and unfermented turmeric-camel milk in streptozotocin-induced diabetes and oxidative stress in rats PMID 40046795 | Michele Schaffer: Not clinical | Prisha Batra: Not clinical | – |
| 137. D'Angelo 2025. The Role of Diet and Oral Supplementation for the Management of Diabetic Retinopathy and Diabetic Macular Edema: A Narrative Review PMID 40041571 | Michele Schaffer: Not isolated | Prisha Batra: Not clinical | – |
| 138. Zhao 2025. Clinical features and prognosis of drug-induced liver injury in patients with non-alcoholic fatty liver PMID 40027577 | Michele Schaffer: No saffron | Prisha Batra: Not clinical | – |
| 139. Wu 2025. Ultrasound-Assisted Extraction of Polysaccharides from Lyophyllum decastes: Structural Analysis and Bioactivity Assessment PMID 40005271 | Michele Schaffer: No saffron | Prisha Batra: Not clinical | – |
| 140. Abo-Elghiet 2025. Bridging Gaps in Migraine Management: A Comprehensive Review of Conventional Treatments, Natural Supplements, Complementary Therapies, and Lifestyle Modifications PMID 40005953 | Michele Schaffer: No saffron | Prisha Batra: Not clinical | – |
| 141. Zhao 2025. Mechanism of Formononetin in Improving Energy Metabolism and Alleviating Neuronal Injury in CIRI Based on Nontargeted Metabolomics Research PMID 39993962 | Michele Schaffer: Not isolated | Prisha Batra: Not clinical | – |
| 142. Xiong 2025. Spicy food intake and overweight/obesity in rural Southwest China: findings from a cross-sectional study PMID 39963664 | Michele Schaffer: No saffron | Prisha Batra: Not clinical | – |
| 143. Amin 2025. Antidepressive and anxiolytic effects of a combination of Saffron and Chamomile in rats and their relationship with serotonin using methods PMID 39957158 | Michele Schaffer: Not isolated | Prisha Batra: Not clinical | – |

Table S2 continued

| Paper | Reviewer 1 | Reviewer 2 | Additional recorded reviews |
|---|---|---|---|
| 144. Yan 2024. Promotion of chronic wound healing by plant-derived active ingredients and research progress and potential of plant polysaccharide hydrogels<br>PMID 39949811 | Michele Schaffer: Not isolated | Prisha Batra: Not clinical | – |
| 145. Hoang 2025. Improving cognitive impairment through chronic consumption of natural compounds/extracts: a systematic review and meta-analysis of randomized controlled trials<br>PMID 39949865 | Michele Schaffer: Not isolated | Prisha Batra: Not clinical | – |
| 146. Handu 2025. Evaluating Bioactive-Substance-Based Interventions for Adults with MASLD: Results from a Systematic Scoping Review<br>PMID 39940310 | Michele Schaffer: Not isolated | Prisha Batra: Not clinical | – |
| 147. Nersezashvili 2025. Seseli foliosum (Somm. et Levier) Manden.-A Comprehensive Phytochemical and Biological Evaluation<br>PMID 39942828 | Michele Schaffer: No saffron | Prisha Batra: Not clinical | – |
| 148. Amiri 2025. Relationship between dietary inflammatory index and metabolic dysfunction associated steatotic liver disease in children<br>PMID 39934274 | Michele Schaffer: No saffron | Prisha Batra: Not clinical | – |
| 149. Ildarabadi 2025. Associations between spice or pepper (Capsicum annuum) consumption and diabetes or metabolic syndrome incidence<br>PMID 39932911 | Michele Schaffer: Not isolated | Prisha Batra: Not clinical | – |
| 150. Zhang 2025. Mitochondria and myocardial ischemia/reperfusion injury: Effects of Chinese herbal medicine and the underlying mechanisms<br>PMID 39931135 | Michele Schaffer: No saffron | Prisha Batra: Not clinical | – |
| 151. Chan 2025. Methodological insights into intervention and outcomes in randomised controlled trials of herbal medicine for obesity: a scoping review<br>PMID 39917276 | Michele Schaffer: Not isolated | Prisha Batra: Not clinical | – |
| 152. Guerbette 2025. Bioenergetic adaptations of small intestinal epithelial cells reduce cell differentiation enhancing intestinal permeability in obese mice<br>PMID 39814101 | Michele Schaffer: No saffron | Prisha Batra: Not clinical | – |
| 153. Payandeh 2025. The association between the dietary inflammatory index during pregnancy and risk of gestational diabetes: a prospective cohort study and a meta-analysis<br>PMID 39901152 | Michele Schaffer: No saffron | Prisha Batra: Not clinical | – |

Table S2 continued

| Paper | Reviewer 1 | Reviewer 2 | Additional recorded reviews |
|---|---|---|---|
| 154. Arar 2025. Protein aggregation and neurodegenerative disease: Structural outlook for the novel therapeutics<br>PMID 37530227 | Michele Schaffer: No saffron | Prisha Batra: Not clinical | – |
| 155. EFSA Scientific Committee 2025. Risks to human and animal health from the presence of bromide in food and feed<br>PMID 39877303 | Michele Schaffer: Duplicate | Prisha Batra: Not clinical | – |
| 156. Lopes 2025. Grape Pomace: A Review of Its Bioactive Phenolic Compounds, Health Benefits, and Applications<br>PMID 39860231 | Michele Schaffer: No saffron | Prisha Batra: Not clinical | – |
| 157. Khattab 2025. Critical Appraisal of Pharmaceutical Therapy in Diabetic Cardiomyopathy-Challenges and Prospectives<br>PMID 39861195 | Michele Schaffer: No saffron | Prisha Batra: Not clinical | – |
| 158. Hashim 2024. Plant-Derived Anti-Cancer Therapeutics and Biopharmaceuticals<br>PMID 39851281 | Michele Schaffer: No saffron | Prisha Batra: Not clinical | – |
| 159. Albadawi 2025. Structural and functional changes in the hippocampus induced by environmental exposures<br>PMID 39800422 | Michele Schaffer: No saffron | Prisha Batra: Not clinical | – |
| 160. Aboutaleb 2025. Circulating visfatin concentrations in patients with chronic obstructive pulmonary disease: systematic review and meta-analysis<br>PMID 39845836 | Michele Schaffer: Not isolated | Prisha Batra: Not clinical | – |
| 161. Masyita 2025. Natural pigments: innovative extraction technologies and their potential application in health and food industries<br>PMID 39845791 | Michele Schaffer: Not isolated | Prisha Batra: Not clinical | – |
| 162. Dastenaie 2025. Common Dietary Patterns Among Female Employees Participating in the Persian Cohort Study (Mashhad) and Their Relationship With Metabolic Syndrome<br>PMID 39846042 | Michele Schaffer: Not isolated | Prisha Batra: Not clinical | – |
| 163. Suschana 2025. The role of anti-inflammatory diets and supplementation in metabolic syndrome and symptom remission in adults with schizophrenia: a systematic review<br>PMID 39839138 | Michele Schaffer: Not isolated | Prisha Batra: Not clinical | – |
| 164. Weerarathna 2024. Compatibility of Whole Wheat-Based Composite Flour in the Development of Functional Foods<br>PMID 39830876 | Michele Schaffer: No saffron | Prisha Batra: Not clinical | – |
| 165. Crous 2024. Fungal Planet description sheets: 1697-1780<br>PMID 39830292 | Michele Schaffer: No saffron | Prisha Batra: Not clinical | – |

Table S2 continued

| Paper | Reviewer 1 | Reviewer 2 | Additional recorded reviews |
|---|---|---|---|
| 166. Liu 2025. Investigating misclassification of type 1 diabetes in a population-based cohort of British Pakistanis and Bangladeshis using polygenic risk scores<br>PMID 39805939 | Michele Schaffer: Not clinical | Prisha Batra: Not clinical | – |
| 167. Ayuso 2024. Broccoli, Artichoke, Carob and Apple By-Products as a Source of Soluble Fiber: How It Can Be Affected by Enzymatic Treatment with Pectinex(®) Ultra SP-L, Viscozyme(®) L and Celluclast(®) 1.5 L<br>PMID 39796300 | Michele Schaffer: No saffron | Prisha Batra: Not clinical | – |
| 168. Stoleru 2024. Multiple nanotechnological approaches using natural compounds for diabetes management<br>PMID 38932892 | Michele Schaffer: Not isolated | Prisha Batra: Not clinical | – |
| 169. Modder 2024. Liver-targeted Angptl4 silencing by antisense oligonucleotide treatment attenuates hyperlipidaemia and atherosclerosis development in APOE*3-Leiden.CETP mice<br>PMID 39259836 | Michele Schaffer: No saffron | Prisha Batra: Not clinical | – |
| 171. Siam 2024. Diabetes Mellitus and Cardiovascular Disease: Exploring Epidemiology, Pathophysiology, and Treatment Strategies<br>PMID 39742220 | Michele Schaffer: Not isolated | Bryan Anh Nguyen: No saffron | Prisha Batra: Not clinical |
| 172. Safiri 2024. Alzheimer's disease: a comprehensive review of epidemiology, risk factors, symptoms diagnosis, management, caregiving, advanced treatments and associated challenges<br>PMID 39736972 | Michele Schaffer: No saffron | Bryan Anh Nguyen: No saffron | Prisha Batra: Not clinical |
| 173. Vaghar Seyedin 2024. Determining effective dietary and ruminal fermentation parameters on live weight gain in diets formulated with selected agro-industrial by-products and their validation in performance fattening lambs<br>PMID 39736982 | Michele Schaffer: No saffron | Bryan Anh Nguyen: No saffron | Prisha Batra: Not clinical |
| 174. Le Jan 2024. Omega-3 Fatty Acid and Vitamin D Supplementations Partially Reversed Metabolic Disorders and Restored Gut Microbiota in Obese Wistar Rats<br>PMID 39765737 | Michele Schaffer: No saffron | Bryan Anh Nguyen: No saffron | Prisha Batra: Not clinical |
| 175. Sri Harsha 2024. Grape Pomace as a Source of Phenolics for the Inhibition of Starch Digestion Enzymes: A Comparative Study and Standardization of the Efficacy<br>PMID 39767045 | Michele Schaffer: Not isolated | Bryan Anh Nguyen: No saffron | Prisha Batra: Not clinical |
| 176. Torghabeh 2023. Dietary anethole: a systematic review of its protective effects against metabolic syndrome<br>PMID 38932801 | Michele Schaffer: No saffron | Bryan Anh Nguyen: No saffron | Prisha Batra: Not clinical |

Table S2 continued

| Paper | Reviewer 1 | Reviewer 2 | Additional recorded reviews |
|---|---|---|---|
| 177. Medoro 2024. Palmitoylethanolamide in Postmenopausal Metabolic Syndrome: Current Evidence and Clinical Perspectives<br>PMID 39770936 | Michele Schaffer: Not isolated | Bryan Anh Nguyen: No saffron | Prisha Batra: Not clinical |
| 178. Rezazadeh 2024. Investigating the resilience of kidneys in rats exposed to chronic partial sleep deprivation and circadian rhythm disruption as disruptive interventions<br>PMID 39720585 | Michele Schaffer: No saffron | Bryan Anh Nguyen: No saffron | Prisha Batra: Not clinical |
| 179. Wang 2024. Oviduct Glycoprotein 1 (OVGP1) Diagnoses Polycystic Ovary Syndrome (PCOS) Based on Machine Learning Algorithms<br>PMID 39713694 | Michele Schaffer: No saffron | Bryan Anh Nguyen: No saffron | Prisha Batra: Not clinical |
| 180. Firooznia 2024. Dietary antioxidant index and hypertension in the Iranian population: a nested case-control study within the Fasa adults cohort study<br>PMID 39713776 | Michele Schaffer: Not isolated | Bryan Anh Nguyen: No saffron | Prisha Batra: Not clinical |
| 181. Vafadar 2024. Phytochemical-mediated efferocytosis and autophagy in inflammation control<br>PMID 39695119 | Michele Schaffer: Not isolated | Bryan Anh Nguyen: No saffron | Prisha Batra: Not clinical |
| 182. Ortega 2024. Understanding immune system dysfunction and its context in mood disorders: psychoneuroimmunoendocrinology and clinical interventions<br>PMID 39681901 | Michele Schaffer: No saffron | Bryan Anh Nguyen: No saffron | Prisha Batra: Not clinical |
| 183. Ho 2024. Exploring Bioactive Phytomedicines for Advancing Pulmonary Infection Management: Insights and Future Prospects<br>PMID 39385504 | Michele Schaffer: No saffron | Bryan Anh Nguyen: No saffron | Pranav Potluri: No saffron |
| 184. Zhao 2024. Cognitive Impairment Mechanisms in High-Altitude Exposure: Proteomic and Metabolomic Insights<br>PMID 39566908 | Michele Schaffer: No saffron | Bryan Anh Nguyen: No saffron | Pranav Potluri: No saffron |
| 185. Ross 2024. Optimizing Testosterone Levels and Fertility Outcomes: A Case Series on the Impact of Lepidium peruvianum (Maca-OG(TM)) in Andropause and Sperm Dysfunction<br>PMID 39651557 | Michele Schaffer: No saffron | Bryan Anh Nguyen: No saffron | Pranav Potluri: No saffron |
| 186. Wang 2024. A high-fat plus high-sucrose diet induces age-related macular degeneration in an experimental rabbit model<br>PMID 39463155 | Michele Schaffer: No saffron | Bryan Anh Nguyen: No saffron | – |

Table S2 continued

| Paper | Reviewer 1 | Reviewer 2 | Additional recorded reviews |
|---|---|---|---|
| 187. Farias 2024. Diet and Immune Effects Trial (DIET)- a randomized, double-blinded dietary intervention study in patients with melanoma receiving immunotherapy PMID 39633321 | Michele Schaffer: No saffron | Bryan Anh Nguyen: No saffron | – |
| 188. Narra 2024. Impact of thermal processing on polyphenols, carotenoids, glucosinolates, and ascorbic acid in fruit and vegetables and their cardiovascular benefits PMID 39169551 | Michele Schaffer: Not isolated | Bryan Anh Nguyen: No saffron | – |
| 189. Sung 2023. Dietary Inflammatory Potential and the Risk of Serrated and Adenomatous Colorectal Polyps PMID 37791878 | Michele Schaffer: No saffron | Bryan Anh Nguyen: No saffron | – |
| 190. Rajabi 2025. Comparative Effects of Crocin and Losartan on RAGE, TGF-$\beta$, TNF-$\alpha$ Gene Expression and Histopathological Changes of the Liver Tissue in Rats With Diabetes PMID 39607899 | Michele Schaffer: Not isolated | Bryan Anh Nguyen: No saffron | – |
| 191. Kazemi 2022. The effects of high intensity interval training induced H(2)O(2), Nrf2 changes on antioxidants factors in type 2 diabetes PMID 39610477 | Michele Schaffer: Not isolated | Bryan Anh Nguyen: Not isolated | – |
| 192. Lazaridis 2024. Fruits, Spices and Honey Phenolic Compounds: A Comprehensive Review on Their Origin, Methods of Extraction and Beneficial Health Properties PMID 39594476 | Michele Schaffer: Not isolated | Bryan Anh Nguyen: No saffron | – |
| 193. Liu 2024. Inhibition of Glycolysis Alleviates Chronic Unpredictable Mild Stress Induced Neuroinflammation and Depression-like Behavior PMID 39595861 | Michele Schaffer: No saffron | Bryan Anh Nguyen: No saffron | – |
| 194. Wang 2024. Spectroscopic and Molecular Docking Studies on the Influence of Inulin on the Interaction of Sophoricoside with Whey Protein Concentrate PMID 39594016 | Michele Schaffer: No saffron | Bryan Anh Nguyen: No saffron | – |
| 195. Jia 2024. The effect of green tea on patients with type 2 diabetes mellitus: A meta-analysis PMID 39809182 | Michele Schaffer: No saffron | Bryan Anh Nguyen: No saffron | – |
| 196. Bulhões 2024. The Action of Colchicine in Patients with Metabolic Syndrome and Obesity: Perspectives and Challenges PMID 39590865 | Michele Schaffer: No saffron | Bryan Anh Nguyen: No saffron | – |
| 197. Law 2024. From Microcirculation to Aging-Related Diseases: A Focus on Endothelial SIRT1 PMID 39598406 | Michele Schaffer: Not isolated | Bryan Anh Nguyen: No saffron | – |

Table S2 continued

| Paper | Reviewer 1 | Reviewer 2 | Additional recorded reviews |
|---|---|---|---|
| 198. Opazo 2024. Prolonged Photobiomodulation with Deep Red Light Mitigates Incipient Retinal Deterioration in a Mouse Model of Type 2 Diabetes<br>PMID 39596197 | Michele Schaffer: No saffron | Bryan Anh Nguyen: No saffron | – |
| 199. Ng 2024. Managing Type 2 Diabetes Mellitus via the Regulation of Gut Microbiota: A Chinese Medicine Perspective<br>PMID 39599721 | Michele Schaffer: No saffron | Bryan Anh Nguyen: No saffron | – |
| 200. Sumalla-Cano 2024. Carotenoids Intake and Cardiovascular Prevention: A Systematic Review<br>PMID 39599645 | Michele Schaffer: Not isolated | Bryan Anh Nguyen: No saffron | – |
| 201. Inia 2024. Therapeutic effects of FGF21 mimetic bFKB1 on MASH and atherosclerosis in Ldlr-/-.Leiden mice<br>PMID 39463193 | Michele Schaffer: No saffron | Bryan Anh Nguyen: No saffron | – |
| 202. Chen 2024. Association between serum carotenoids and hyperuricemia: a cross-sectional study based on NHANES 2001-2006<br>PMID 39574526 | Michele Schaffer: No saffron | Bryan Anh Nguyen: No saffron | – |
| 203. Yu 2024. Overview of anti-inflammatory diets and their promising effects on non-communicable diseases<br>PMID 39411832 | Michele Schaffer: No saffron | Bryan Anh Nguyen: No saffron | – |
| 204. Mirzavi 2024. The potential protective role of carotenoids from saffron: A focus on endoplasmic reticulum stress-related organ damage<br>PMID 39554322 | Michele Schaffer: Not clinical | Bryan Anh Nguyen: No saffron | – |
| 205. Rahman 2024. Diosmin: A promising phytochemical for functional foods, nutraceuticals and cancer therapy<br>PMID 39554345 | Michele Schaffer: No saffron | Bryan Anh Nguyen: No saffron | – |
| 206. Yao 2024. Effects of inulin-type oligosaccharides (JSO) from Cichorium intybus L. on behavioral deficits induced by chronic restraint stress in mice and associated molecular alterations<br>PMID 39555096 | Michele Schaffer: No saffron | Bryan Anh Nguyen: No saffron | – |
| 207. Fatimah 2024. Mechanism of streptozotocin to induce cardiac fibrosis through TNFa and Bcl2 pathways in in silico and in vivo study<br>PMID 39553747 | Michele Schaffer: No saffron | Bryan Anh Nguyen: No saffron | – |
| 208. Gholami 2024. Preventive Effects of Crocin, a Key Carotenoid Component in Saffron, Against Nicotine-Triggered Neurodegeneration in Rat Hippocampus: Possible Role of Autophagy and Apoptosis<br>PMID 39539579 | Michele Schaffer: Not clinical | Bryan Anh Nguyen: No saffron | – |

Table S2 continued

| Paper | Reviewer 1 | Reviewer 2 | Additional recorded reviews |
|---|---|---|---|
| 209. Guo 2024. Exploring the Potential of Anthocyanins for Repairing Photoaged Skin: A Comprehensive Review<br>PMID 39517290 | Michele Schaffer: No saffron | Bryan Anh Nguyen: No saffron | – |
| 210. Giurranna 2024. Dietary Antioxidants and Natural Compounds in Preventing Thrombosis and Cardiovascular Disease<br>PMID 39519009 | Michele Schaffer: Not isolated | Bryan Anh Nguyen: No saffron | – |
| 211. Kacar 2024. Morphological analysis and cytotoxicity of acrylamide on SPC212 human mesothelioma cells: Do low doses induce proliferation, while high doses cause toxicity?<br>PMID 39516185 | Michele Schaffer: No saffron | Bryan Anh Nguyen: No saffron | – |
| 212. Ansari 2024. Plant-Based Diets and Phytochemicals in the Management of Diabetes Mellitus and Prevention of Its Complications: A Review<br>PMID 39519546 | Michele Schaffer: Not isolated | Bryan Anh Nguyen: No saffron | – |
| 213. Shamabadi 2024. Crocus sativus (saffron) and age-related macular degeneration<br>PMID 39507811 | Michele Schaffer: Not clinical | Bryan Anh Nguyen: No saffron | – |
| 214. Zuercher 2024. Energy-Adjusted Dietary Inflammatory Index and Diabetes Risk in Postmenopausal Hispanic Women<br>PMID 37544374 | Michele Schaffer: Not isolated | Bryan Anh Nguyen: No saffron | – |
| 215. Magni 2013. Fluoxetine versus other types of pharmacotherapy for depression<br>PMID 24353997 | Grace Kim: No saffron | Michele Schaffer: Not isolated | Bryan Anh Nguyen: No saffron |
| 216. Pashaei 2024. Comparison of diet quality indices for predicting metabolic syndrome in Iran: cross-sectional findings from the persian cohort study<br>PMID 39465430 | Michele Schaffer: Not isolated | Bryan Anh Nguyen: No saffron | – |
| 217. Xiang 2024. DII modulates the relationship between SVD3 and NAFLD prevalence, rather than liver fibrosis severity, in hospitalized T2DM population<br>PMID 39462138 | Michele Schaffer: No saffron | Bryan Anh Nguyen: No saffron | – |
| 218. Jiménez-Jiménez 2024. Antioxidant Therapies in the Treatment of Multiple Sclerosis<br>PMID 39456199 | Michele Schaffer: Not isolated | Bryan Anh Nguyen: No saffron | – |
| 219. Boutaj 2024. A Comprehensive Review of Moroccan Medicinal Plants for Diabetes Management<br>PMID 39452489 | Michele Schaffer: Not isolated | Bryan Anh Nguyen: No saffron | – |

Table S2 continued

| Paper | Reviewer 1 | Reviewer 2 | Additional recorded reviews |
| --- | --- | --- | --- |
| 220. Son 2024. Impact of the anti-inflammatory diet on serum high-sensitivity C-Reactive protein and new-onset frailty in community-dwelling older adults: A 7-year follow-up of the Kashiwa cohort study PMID 38126695 | Michele Schaffer: Not isolated | Bryan Anh Nguyen: No saffron | – |
| 221. Neagu 2024. Double-Edged Sword Effect of Diet and Nutrition on Carcinogenic Molecular Pathways in Breast Cancer PMID 39456858 | Michele Schaffer: Not clinical | Bryan Anh Nguyen: No saffron | – |
| 222. Merino Del Portillo 2024. Nutritional Modulation of the Gut-Brain Axis: A Comprehensive Review of Dietary Interventions in Depression and Anxiety Management PMID 39452930 | Michele Schaffer: Not clinical | Bryan Anh Nguyen: No saffron | – |
| 223. Tauil 2024. Metabolic-Associated Fatty Liver Disease: The Influence of Oxidative Stress, Inflammation, Mitochondrial Dysfunctions, and the Role of Polyphenols PMID 39458995 | Michele Schaffer: Not isolated | Bryan Anh Nguyen: No saffron | – |
| 224. Moise 2024. Plants' Impact on the Human Brain-Exploring the Neuroprotective and Neurotoxic Potential of Plants PMID 39458980 | Michele Schaffer: Not isolated | Bryan Anh Nguyen: No saffron | – |
| 225. Samimi 2024. Evaluation of Antioxidant Effects of Coenzyme Q10 against Hyperglycemia-Mediated Oxidative Stress by Focusing on Nrf2/Keap1/HO-1 Signaling Pathway in the Liver of Diabetic Rats PMID 39449772 | Michele Schaffer: Not isolated | Bryan Anh Nguyen: No saffron | – |
| 226. Guo 2024. Spermidine attenuates chondrocyte inflammation and cellular pyroptosis through the AhR/NF-$\kappa$B axis and the NLRP3/caspase-1/GSDMD pathway PMID 39416781 | Michele Schaffer: No saffron | Bryan Anh Nguyen: No saffron | – |
| 227. Koufan 2024. Structure-Function Relationships and Health-Promoting Properties of the Main Nutraceuticals of the Cactus Pear (Opuntia spp.) Cladodes: A Review PMID 39407660 | Michele Schaffer: No saffron | Bryan Anh Nguyen: No saffron | – |
| 228. Liu 2024. High Polygenic Risk Scores Positively Associated with Gastric Cancer Risk Interact with Coffee and Polyphenol Intake and Smoking Status in Korean Adults PMID 39408230 | Michele Schaffer: No saffron | Bryan Anh Nguyen: No saffron | – |
| 229. Li 2024. Glutathione: A Key Regulator of Extracellular Matrix and Cell Death in Intervertebral Disc Degeneration PMID 39403548 | Michele Schaffer: Not isolated | Bryan Anh Nguyen: No saffron | – |

Table S2 continued

| Paper | Reviewer 1 | Reviewer 2 | Additional recorded reviews |
|---|---|---|---|
| 230. Zhu 2024. NLRP3 inhibitor combined with Yimusake improves erectile dysfunction in rats with diabetes mellitus through the attenuation of pyroptosis<br>PMID 39391494 | Michele Schaffer: Not isolated | Bryan Anh Nguyen: No saffron | – |
| 231. Moreira 2024. Suppression of the postprandial hyperglycemia in patients with type 2 diabetes by a raw medicinal herb powder is weakened when consumed in ordinary hard gelatin capsules: A randomized crossover clinical trial<br>PMID 39383145 | Michele Schaffer: Not isolated | Bryan Anh Nguyen: No saffron | – |
| 232. Yang 2024. The major biogenic amine metabolites in mood disorders<br>PMID 39381610 | Michele Schaffer: Not isolated | Bryan Anh Nguyen: No saffron | – |
| 233. Abujrais 2024. Untargeted Metabolomics and Quantitative Analysis of Tryptophan Metabolites in Myalgic Encephalomyelitis Patients and Healthy Volunteers: A Comparative Study Using High-Resolution Mass Spectrometry<br>PMID 39302151 | Michele Schaffer: No saffron | Bryan Anh Nguyen: No saffron | – |
| 234. Spurgeon 2024. Proceedings of the 2022 "Lifestyle Intervention for Epilepsy (LIFE)" symposium hosted by Cleveland Clinic<br>PMID 39177045 | Michele Schaffer: No saffron | Bryan Anh Nguyen: No saffron | – |
| 235. Yang 2024. The impact of dietary interventions on polycystic ovary syndrome patients with a BMI $\geq$25 kg/m(2): A systematic review and meta-analysis of randomized controlled trials<br>PMID 39351128 | Michele Schaffer: No saffron | Bryan Anh Nguyen: No saffron | – |
| 236. Chauhan 2024. Exploring the Potential of Saffron as a Therapeutic Agent in Depression Treatment: A Comparative Review<br>PMID 39351321 | Michele Schaffer: Not isolated | Bryan Anh Nguyen: No saffron | – |
| 237. Bano 2024. Deciphering the Potentials of Cardamom in Cancer Prevention and Therapy: From Kitchen to Clinic<br>PMID 39334932 | Michele Schaffer: No saffron | Bryan Anh Nguyen: No saffron | – |
| 238. Suzauddula 2024. Bioengineered Anthocyanin-Enriched Tomatoes: A Novel Approach to Colorectal Cancer Prevention<br>PMID 39335919 | Michele Schaffer: No saffron | Bryan Anh Nguyen: No saffron | – |
| 239. Martínez-García 2024. The Molecular Comorbidity Network of Periodontal Disease<br>PMID 39337647 | Michele Schaffer: No saffron | Bryan Anh Nguyen: No saffron | – |

Table S2 continued

| Paper | Reviewer 1 | Reviewer 2 | Additional recorded reviews |
|---|---|---|---|
| 240. Cuchillo-Hilario 2024. Animal Food Products to Support Human Nutrition and to Boost Human Health: The Potential of Feedstuffs Resources and Their Metabolites as Health-Promoters PMID 39330503 | Michele Schaffer: Not isolated | Bryan Anh Nguyen: No saffron | – |
| 241. Sbai 2024. Effect of the Mediterranean Diet (MeDi) on the Progression of Retinal Disease: A Narrative Review PMID 39339769 | Michele Schaffer: Not isolated | Bryan Anh Nguyen: No saffron | – |
| 242. Villaverde 2024. Dietary Inflammatory Index and Blood Pressure Levels in Mexican Adults PMID 39339652 | Michele Schaffer: Not isolated | Bryan Anh Nguyen: No saffron | – |
| 243. Ahmed 2024. Characterising the Metabolomic Diversity and Biological Potentials of Extracts from Different Parts of Two Cistus Species Using UHPLC-MS/MS and In Vitro Techniques PMID 39338986 | Michele Schaffer: No saffron | Bryan Anh Nguyen: No saffron | – |
| 244. Islam 2024. Impact of dietary ingredients on radioprotection and radiosensitization: a comprehensive review PMID 39320122 | Michele Schaffer: No saffron | Bryan Anh Nguyen: No saffron | – |
| 245. Neagu 2024. Diabetic neuropathy: A NRF2 disease? PMID 38158644 | Michele Schaffer: No saffron | Bryan Anh Nguyen: No saffron | – |
| 246. Vajdi 2024. Therapeutic effect of fenugreek supplementation on type 2 diabetes mellitus: A systematic review and meta-analysis of clinical trials PMID 39286181 | Michele Schaffer: Not isolated | Bryan Anh Nguyen: No saffron | – |
| 247. Netala 2024. A Comprehensive Review of Cardiovascular Disease Management: Cardiac Biomarkers, Imaging Modalities, Pharmacotherapy, Surgical Interventions, and Herbal Remedies PMID 39273041 | Michele Schaffer: Not isolated | Bryan Anh Nguyen: No saffron | – |
| 248. Mattioli 2024. From Waste to Health: Olive Mill Wastewater for Cardiovascular Disease Prevention PMID 39275301 | Michele Schaffer: No saffron | Bryan Anh Nguyen: No saffron | – |
| 249. Li 2024. Capsaicin: a spicy way in liver disease PMID 39281271 | Michele Schaffer: No saffron | Bryan Anh Nguyen: No saffron | – |
| 251. Farag 2024. Metabolites profiling of Sapota fruit pulp via a multiplex approach of gas and ultra performance liquid chromatography/mass spectroscopy in relation to its lipase inhibition effect PMID 39221269 | Michele Schaffer: No saffron | Bryan Anh Nguyen: No saffron | – |

Table S2 continued

| Paper | Reviewer 1 | Reviewer 2 | Additional recorded reviews |
|---|---|---|---|
| 252. Pezzino 2024. Innovative treatments for obesity and NAFLD: A bibliometric study on antioxidants, herbs, phytochemicals, and natural compounds<br>PMID 39220898 | Michele Schaffer: Not clinical | Bryan Anh Nguyen: No saffron | – |
| 253. Mulligan 2024. The Dietary Inflammatory Index and Its Associations with Biomarkers of Nutrients with Antioxidant Potential, a Biomarker of Inflammation and Multiple Long-Term Conditions<br>PMID 39199208 | Michele Schaffer: No saffron | Bryan Anh Nguyen: No saffron | – |
| 254. Nunes 2024. Curcumin: A Golden Approach to Healthy Aging: A Systematic Review of the Evidence<br>PMID 39203857 | Michele Schaffer: Not isolated | Bryan Anh Nguyen: No saffron | – |
| 255. Wang 2024. Natural products derived from traditional Chinese medicines targeting ER stress for the treatment of kidney diseases<br>PMID 39192602 | Michele Schaffer: Not isolated | Bryan Anh Nguyen: No saffron | – |
| 256. Turkkahraman 2024. Sclerostin antibody corrects periodontal disease in type 2 diabetic mice<br>PMID 39171525 | Michele Schaffer: No saffron | Bryan Anh Nguyen: No saffron | – |
| 257. Soheilifar 2024. In vitro and in vivo evaluation of the diabetic wound healing properties of Saffron (Crocus Sativus L.) petals<br>PMID 39169073 | Michele Schaffer: Not clinical | Bryan Anh Nguyen: Not clinical | – |
| 258. Hao 2024. The role of calcium channels in osteoporosis and their therapeutic potential<br>PMID 39170742 | Michele Schaffer: No saffron | Bryan Anh Nguyen: No saffron | – |
| 259. Khoramipour 2024. The effect of high-intensity interval training on type 2 diabetic muscle: A metabolomics-based study<br>PMID 39170342 | Michele Schaffer: Not isolated | Bryan Anh Nguyen: No saffron | – |
| 260. Motallaei 2024. A cross- sectional study on the association between lifestyle factors and coronary artery stenosis severity among adults living in central Iran: A protocol for the Iranian- CARDIO study<br>PMID 39165856 | Michele Schaffer: Not isolated | Bryan Anh Nguyen: No saffron | – |
| 261. Jacob 2024. Soybean oil-based HFD induces gut dysbiosis that leads to steatosis, hepatic inflammation and insulin resistance in mice<br>PMID 39165573 | Michele Schaffer: No saffron | Bryan Anh Nguyen: No saffron | – |

Table S2 continued

| Paper | Reviewer 1 | Reviewer 2 | Additional recorded reviews |
|---|---|---|---|
| 262. Liu 2024. Association between oxidative balance score and diabetic kidney disease, low estimated glomerular filtration rate and albuminuria in type 2 diabetes mellitus patients: a cross-sectional study<br>PMID 39145317 | Michele Schaffer: Not isolated | Bryan Anh Nguyen: No saffron | – |
| 263. Etebarian 2024. Propolis as a functional food and promising agent for oral health and microbiota balance: A review study<br>PMID 39139934 | Michele Schaffer: Not isolated | Bryan Anh Nguyen: No saffron | – |
| 264. Yang 2024. Causal association of dietary factors with five common cancers: univariate and multivariate Mendelian randomization studies<br>PMID 39135550 | Michele Schaffer: Not isolated | Bryan Anh Nguyen: No saffron | – |
| 265. Pyrzynska 2024. Plant Extracts for Production of Functionalized Selenium Nanoparticles<br>PMID 39124412 | Michele Schaffer: No saffron | Bryan Anh Nguyen: No saffron | – |
| 266. Vella 2024. The Mediterranean Species Calendula officinalis and Foeniculum vulgare as Valuable Source of Bioactive Compounds<br>PMID 39124999 | Michele Schaffer: No saffron | Bryan Anh Nguyen: No saffron | – |
| 267. Bolat 2024. Polyphenols: Secondary Metabolites with a Biological Impression<br>PMID 39125431 | Michele Schaffer: No saffron | Bryan Anh Nguyen: No saffron | – |
| 268. Yaikwawong 2024. Curcumin Reduces Depression in Obese Patients with Type 2 Diabetes: A Randomized Controlled Trial<br>PMID 39125295 | Michele Schaffer: No saffron | Prisha Batra: No saffron | – |
| 269. Bing 2024. Dysregulated lipid metabolism and intervertebral disc degeneration: the important role of ox-LDL/LOX-1 in endplate chondrocyte senescence and calcification<br>PMID 39123116 | Michele Schaffer: No saffron | Prisha Batra: No saffron | – |
| 270. Li 2024. Regulated anthocyanin release through novel pH-responsive peptide hydrogels in simulated digestive environment<br>PMID 39113736 | Michele Schaffer: No saffron | Prisha Batra: No saffron | – |
| 271. Zheng 2024. Roles of distinct nuclear receptors in diabetic cardiomyopathy<br>PMID 39114353 | Michele Schaffer: Not isolated | Prisha Batra: Not clinical | – |

Table S2 continued

| Paper | Reviewer 1 | Reviewer 2 | Additional recorded reviews |
|---|---|---|---|
| 272. Benhaddou 2024. Effects of gastric bypass on the digestibility and postprandial metabolic fate of 15N dietary protein in rats<br>PMID 39102389 | Michele Schaffer: No saffron | Prisha Batra: No saffron | – |
| 273. Jung 2024. Association of the dietary inflammatory index with sarcopenic obesity and frailty in older adults<br>PMID 39097690 | Michele Schaffer: No saffron | Prisha Batra: Not clinical | – |
| 274. Elkanawati 2024. Impact of Lipids on Insulin Resistance: Insights from Human and Animal Studies<br>PMID 39100221 | Michele Schaffer: Not isolated | Prisha Batra: Not clinical | – |
| 275. Fatima 2024. Biosynthesis and Pharmacological Activities of the Bioactive Compounds of White Mulberry (Morus alba): Current Paradigms and Future Challenges<br>PMID 39056699 | Michele Schaffer: Not isolated | Prisha Batra: No saffron | – |
| 276. Pan 2024. Ghrelin/GHSR System in Depressive Disorder: Pathologic Roles and Therapeutic Implications<br>PMID 39057075 | Michele Schaffer: Not isolated | Prisha Batra: Not clinical | – |
| 277. Gomes 2024. Secondary Bioactive Metabolites from Foods of Plant Origin as Theravention Agents against Neurodegenerative Disorders<br>PMID 39063373 | Michele Schaffer: Not isolated | Prisha Batra: Not clinical | – |
| 278. Ki 2024. Natural Compounds for Preventing Age-Related Diseases and Cancers<br>PMID 39062777 | Michele Schaffer: Not isolated | Prisha Batra: Not clinical | – |
| 279. Cai 2024. Functional Nutrients and Jujube-Based Processed Products in Ziziphus jujuba<br>PMID 39065014 | Michele Schaffer: Not isolated | Prisha Batra: No saffron | – |
| 280. Chen 2024. Optimization of Extraction and Purification of Flavonoids from Stigmaless Floral Residues of Crocus sativus L. and Their Stimulatory Effect on Glucose Uptake In Vitro<br>PMID 39064849 | Michele Schaffer: Not clinical | Prisha Batra: Not clinical | – |
| 281. Rashid 2024. Saffron as a Promising Therapy for Inflammatory Bowel Disease<br>PMID 39064796 | Michele Schaffer: Not clinical | Prisha Batra: Not clinical | – |
| 282. Marrone 2024. Saffron (Crocus sativus L.) and Its By-Products: Healthy Effects in Internal Medicine<br>PMID 39064764 | Michele Schaffer: Not clinical | Prisha Batra: Not clinical | – |
| 283. Xue 2024. Dilemmas in Elderly Diabetes and Clinical Practice Involving Traditional Chinese Medicine<br>PMID 39065801 | Michele Schaffer: Not clinical | Prisha Batra: Not clinical | – |

Table S2 continued

| Paper | Reviewer 1 | Reviewer 2 | Additional recorded reviews |
|---|---|---|---|
| 284. Dissinger 2024. Chronic social isolation affects feeding behavior of juvenile zebrafish (Danio rerio)<br>PMID 39058733 | Michele Schaffer: No saffron | Prisha Batra: No saffron | – |
| 285. Siliman Misha 2024. Preventive effects of a nutraceutical mixture of berberine, citrus and apple extracts on metabolic disturbances in Zucker fatty rats<br>PMID 39058681 | Michele Schaffer: No saffron | Prisha Batra: No saffron | – |
| 286. Sabouni 2024. Stimulating effect of nanocurcumin and crocin on proliferation and pluripotency of bone marrow-derived mesenchymal stem cells<br>PMID 39055876 | Michele Schaffer: Not isolated | Prisha Batra: Not clinical | – |
| 287. Li 2024. Mechanisms of regulation of glycolipid metabolism by natural compounds in plants: effects on short-chain fatty acids<br>PMID 39026248 | Michele Schaffer: Not isolated | Prisha Batra: No saffron | – |
| 288. Farhadnejad 2024. Herbal Products as Complementary or Alternative Medicine for the Management of Hyperglycemia and Dyslipidemia in Patients with Type 2 Diabetes: Current Evidence Based on Findings of Interventional Studies<br>PMID 39021815 | Michele Schaffer: Not isolated | Prisha Batra: Not clinical | – |
| 289. Darmishonnejad 2024. Effects of Acute Exposure to Methylglyoxal or/and A Diet Rich in Advanced Glycation End Products on Sperm Parameters in Mice<br>PMID 38973280 | Michele Schaffer: No saffron | Prisha Batra: No saffron | – |
| 290. #N/A #N/A. ISEV2024 Abstract Book<br>PMID #N/A | Michele Schaffer: No saffron | Prisha Batra: Not clinical | – |
| 291. Catalano 2024. Aloe vera-An Extensive Review Focused on Recent Studies<br>PMID 38998660 | Michele Schaffer: No saffron | Prisha Batra: No saffron | – |
| 292. Del Campo 2024. Food Neophobias in Spanish Adults with Overweight or Obesity by Sex: Their Association with Sociodemographic Factors and the Most Prevalent Chronic Diseases<br>PMID 38998536 | Michele Schaffer: No saffron | Prisha Batra: No saffron | – |
| 293. Zeng 2024. Physiological Mechanisms by Which the Functional Ingredients in Beer Impact Human Health<br>PMID 38999065 | Michele Schaffer: Not isolated | Prisha Batra: Not clinical | – |
| 294. Palus 2024. Dietary Exposure to Acrylamide Has Negative Effects on the Gastrointestinal Tract: A Review<br>PMID 38999779 | Michele Schaffer: Not isolated | Prisha Batra: Not clinical | – |

Table S2 continued

| Paper | Reviewer 1 | Reviewer 2 | Additional recorded reviews |
|---|---|---|---|
| 295. Bauer 2024. A comprehensive in-vitro/in-vivo screening toolbox for the elucidation of glucose homeostasis modulating properties of plant extracts (from roots) and its bioactives PMID 38989154 | Michele Schaffer: No saffron | Prisha Batra: Not clinical | – |
| 296. Yong 2024. The effect and mechanism of palmar ginseng in type 2 diabetic cognitive impairment PMID 38988538 | Michele Schaffer: Not isolated | Prisha Batra: No saffron | – |
| 297. Mao 2024. Clinical characteristics and treatment compounds of obesity-related kidney injury PMID 38983811 | Michele Schaffer: Not isolated | Prisha Batra: No saffron | – |
| 298. Demirel 2024. Vasorelaxant effects of biochemical constituents of various medicinal plants and their benefits in diabetes PMID 38983824 | Michele Schaffer: Not isolated | Prisha Batra: Not clinical | – |
| 299. McBenedict 2024. The Role of Alternative Medicine in Managing Type 2 Diabetes: A Comprehensive Review PMID 38978922 | Michele Schaffer: Not isolated | Prisha Batra: Not clinical | – |
| 300. Mohammad Zaki 2024. The cardioprotective properties of Persicaria maculosa and Citrus sinensis extracts against doxorubicin-induced cardiotoxicity in mice PMID 38952773 | Michele Schaffer: Not isolated | Prisha Batra: Not clinical | – |
| 301. Arammi 2024. The effects of swimming training at different temperatures along with cinnamon supplementation on liver enzymes and thyroid hormones in diabetic rats PMID 38948171 | Michele Schaffer: Not isolated | Prisha Batra: Not clinical | – |
| 302. #N/A 2024. Posters PMID #N/A | Michele Schaffer: No saffron | Prisha Batra: Not clinical | – |
| 303. AkbariRad 2024. Prevalence, characteristics, and risk factors of non-alcoholic fatty liver disease in North East of Iran: a population-based study PMID 38926664 | Michele Schaffer: Not isolated | Prisha Batra: No saffron | – |
| 304. Mantadaki 2024. Benefits of Quercetin on Glycated Hemoglobin, Blood Pressure, PiKo-6 Readings, Night-Time Sleep, Anxiety, and Quality of Life in Patients with Type 2 Diabetes Mellitus: A Randomized Controlled Trial PMID 38930033 | Michele Schaffer: Not isolated | Prisha Batra: No saffron | – |

Table S2 continued

| Paper | Reviewer 1 | Reviewer 2 | Additional recorded reviews |
|---|---|---|---|
| 305. Martemucci 2024. Comprehensive Strategies for Metabolic Syndrome: How Nutrition, Dietary Polyphenols, Physical Activity, and Lifestyle Modifications Address Diabesity, Cardiovascular Diseases, and Neurodegenerative Conditions PMID 38921462 | Michele Schaffer: Not isolated | Prisha Batra: Not clinical | – |
| 306. Kochumon 2024. Gut Dysbiosis Shaped by Cocoa Butter-Based Sucrose-Free HFD Leads to Steatohepatitis, and Insulin Resistance in Mice PMID 38931284 | Michele Schaffer: No saffron | Prisha Batra: No saffron | – |
| 307. Widowati 2024. Antidiabetic, antidyslipidemia, and renoprotector potency of butterfly pea flower extract (Clitorea ternatea L.) in diabetes mellitus and dyslipidemia rats model PMID 38938424 | Michele Schaffer: No saffron | Prisha Batra: No saffron | – |
| 308. Ortega-Regules 2024. Recent Advances in the Therapeutic Potential of Carotenoids in Preventing and Managing Metabolic Disorders PMID 38931016 | Michele Schaffer: Not isolated | Prisha Batra: Not clinical | – |
| 309. Nyulas 2024. Cardiovascular Effects of Herbal Products and Their Interaction with Antihypertensive Drugs-Comprehensive Review PMID 38928095 | Michele Schaffer: Not isolated | Prisha Batra: Not clinical | – |
| 310. Alqarni 2024. Do polyphenols affect body fat and/or glucose metabolism? PMID 38919387 | Michele Schaffer: Not isolated | Prisha Batra: No saffron | – |
| 311. Báez 2023. Non-Chinese herbal medicines for functional dyspepsia PMID 37323050 | Michele Schaffer: Not isolated | Prisha Batra: Not clinical | – |
| 312. Shafe 2024. Lycopene: A Potent Antioxidant with Multiple Health Benefits PMID 38883868 | Michele Schaffer: Not isolated | Prisha Batra: Not clinical | – |
| 313. Munir 2024. Unveiling the Chemistry of Citrus Peel: Insights into Nutraceutical Potential and Therapeutic Applications PMID 38890908 | Michele Schaffer: Not isolated | Prisha Batra: No saffron | – |
| 314. Wan 2024. The associations between dietary flavonoid intake and hyperlipidemia: data from the national health and nutrition examination survey 2007-2010 and 2017-2018 PMID 38883860 | Michele Schaffer: Not isolated | Prisha Batra: Not clinical | – |

Table S2 continued

| Paper | Reviewer 1 | Reviewer 2 | Additional recorded reviews |
|---|---|---|---|
| 315. Vicente-Zurdo 2024. A Comprehensive Analytical Review of Polyphenols: Evaluating Neuroprotection in Alzheimer's Disease<br>PMID 38892094 | Michele Schaffer: No saffron | Prisha Batra: Not clinical | – |
| 316. Song 2024. A Cross Talking between the Gut Microbiota and Metabolites of Participants in a Confined Environment<br>PMID 38892694 | Michele Schaffer: Not isolated | Prisha Batra: No outcome | – |
| 317. Lee 2024. Is Curcumin Intake Really Effective for Chronic Inflammatory Metabolic Disease? A Review of Meta-Analyses of Randomized Controlled Trials<br>PMID 38892660 | Michele Schaffer: Not isolated | Prisha Batra: Not clinical | – |
| 318. Al Dhaheri 2024. The Effect of Therapeutic Doses of Culinary Spices in Metabolic Syndrome: A Randomized Controlled Trial<br>PMID 38892617 | Michele Schaffer: No saffron | Prisha Batra: No saffron | – |
| 319. Mir 2024. Saffron, a Potential Bridge between Nutrition and Disease Therapeutics: Global Health Challenges and Therapeutic Opportunities<br>PMID 38891276 | Michele Schaffer: Not clinical | Prisha Batra: Not clinical | – |
| 320. Lee 2024. Implications of helplessness in depression: diagnosing mild cognitive impairment and analyzing its effects on cognitive decline in older adults<br>PMID 38859876 | Michele Schaffer: Not isolated | Prisha Batra: No saffron | – |
| 321. Althobaiti 2024. Exploring the NRF2/HO-1 and NF-$\kappa$B Pathways: Spirulina Nanoparticles as a Novel Approach to Combat Diabetic Nephropathy<br>PMID 38854532 | Michele Schaffer: Not clinical | Prisha Batra: Not clinical | – |
| 322. Mohammadabadi 2024. Cinnamon: a nutraceutical supplement for the cardiovascular system<br>PMID 38846056 | Michele Schaffer: Not isolated | Prisha Batra: Not clinical | – |
| 323. Golubnitschaja 2024. Mini-encyclopedia of mitochondria-relevant nutraceuticals protecting health in primary and secondary care-clinically relevant 3PM innovation<br>PMID 38841620 | Michele Schaffer: Not isolated | Prisha Batra: Not clinical | – |
| 324. Mahajan 2024. The higher dietary inflammation is associated with a higher burden of multimorbidity of cardio-metabolic and mental health disorders in an urbanizing community of southern India: A cross-sectional analysis for the APCAPS cohort<br>PMID 38828398 | Michele Schaffer: No saffron | Prisha Batra: Not clinical | – |

Table S2 continued

| Paper | Reviewer 1 | Reviewer 2 | Additional recorded reviews |
|---|---|---|---|
| 325. Raslan 2024. Different modalities to manage rheumatoid arthritis: an A to Z story<br>PMID 38827788 | Michele Schaffer: Not clinical | Prisha Batra: Not clinical | – |
| 326. Aziz 2024. Portulaca oleracea L seed extracts counteract diabetic nephropathy through SDF-1/IL10/PPAR$\gamma$-mediated tuning of keap1/Nrf2 and NF-$\kappa$B transcription in Sprague Dawley rats<br>PMID 38811991 | Michele Schaffer: Not isolated | Prisha Batra: No saffron | – |
| 327. Hu 2016. Antidiabetic activity of Helicteres angustifolia root<br>PMID 26866383 | Michele Schaffer: Not clinical | Prisha Batra: No saffron | – |
| 328. Liu 2024. Molecular characterization and structure basis of a malonyltransferase with both substrate promiscuity and catalytic regiospecificity from Cistanche tubulosa<br>PMID 38799633 | Michele Schaffer: Not isolated | Prisha Batra: Not clinical | – |
| 329. Banaszak 2024. The Impact of Curcumin, Resveratrol, and Cinnamon on Modulating Oxidative Stress and Antioxidant Activity in Type 2 Diabetes: Moving beyond an Anti-Hyperglycaemic Evaluation<br>PMID 38790615 | Michele Schaffer: Not isolated | Prisha Batra: Not clinical | – |
| 330. Mao 2024. Biological Functions and Potential Therapeutic Significance of O-GlcNAcylation in Hepatic Cellular Stress and Liver Diseases<br>PMID 38786029 | Michele Schaffer: Not isolated | Prisha Batra: Not clinical | – |
| 331. Koo 2024. Health Outcome Comparison Based on Dietary Inflammatory Levels among Sample of Korean Elderly<br>PMID 38786413 | Michele Schaffer: No saffron | Prisha Batra: No saffron | – |
| 332. Tamas 2024. Metabolic Contrasts: Fatty Acid Oxidation and Ketone Bodies in Healthy Brains vs. Glioblastoma Multiforme<br>PMID 38791520 | Michele Schaffer: Not isolated | Prisha Batra: No saffron | – |
| 333. Zeng 2024. Tanshinone IIA is superior to paricalcitol in ameliorating tubulointerstitial fibrosis through regulation of VDR/Wnt/$\beta$-catenin pathway in rats with diabetic nephropathy<br>PMID 37991543 | Michele Schaffer: Not isolated | Prisha Batra: Not clinical | – |
| 334. Ge 2024. DHCR24 inhibitor SH42 increases desmosterol without preventing atherosclerosis development in mice<br>PMID 38770137 | Michele Schaffer: No saffron | Prisha Batra: Not clinical | – |

Table S2 continued

| Paper | Reviewer 1 | Reviewer 2 | Additional recorded reviews |
|---|---|---|---|
| 335. Portes 2024. Resistance Exercise Training Mitigates Cardiac Remodeling Induced by a High-Fat Diet in Rodents: A Systematic Review<br>PMID 38695409 | Michele Schaffer: Not isolated | Prisha Batra: Not clinical | – |
| 336. Abdulghani 2024. Natural products for managing metabolic syndrome: a scoping review<br>PMID 38746011 | Michele Schaffer: Not isolated | Prisha Batra: Not clinical | – |
| 337. Birkinshaw 2023. Antidepressants for pain management in adults with chronic pain: a network meta-analysis<br>PMID 37160297 | Michele Schaffer: Not isolated | Prisha Batra: Not clinical | – |
| 338. Gao 2024. Sanghuangporus vaninii extract ameliorates hyperlipidemia in rats by mechanisms identified with transcriptome analysis<br>PMID 38726415 | Michele Schaffer: Not isolated | Prisha Batra: No saffron | – |
| 339. Wang 2024. Advancements in magnetic nanoparticle-based biosensors for point-of-care testing<br>PMID 38725992 | Michele Schaffer: Not isolated | Prisha Batra: Not clinical | – |
| 340. Kaddour 2024. Effects of subcutaneous vs. oral nanoparticle-mediated insulin delivery on hemostasis disorders in type 1 diabetes: A rat model study<br>PMID 38711655 | Michele Schaffer: Not isolated | Prisha Batra: Not clinical | – |
| 341. Hasan 2023. Clonal Blumea lacera (Burm. f.) DC. ameliorates diabetic conditions by modulating carbohydrate and lipid hydrolases: a combine in vivo experimental and chemico-biological interaction study<br>PMID 37131966 | Michele Schaffer: No saffron | Prisha Batra: Not clinical | – |
| 342. Goyal 2024. Saffron extract as an emerging novel therapeutic option in reproduction and sexual health: recent advances and future prospectives<br>PMID 38694315 | Michele Schaffer: Not isolated | Prisha Batra: Not clinical | – |
| 343. Remígio 2024. Spray-Drying Microencapsulation of Bauhinia ungulata L. var. obtusifolia Aqueous Extract Containing Phenolic Compounds: A Comparative Study Using Different Wall Materials<br>PMID 38675149 | Michele Schaffer: Not isolated | Prisha Batra: Not clinical | – |
| 344. Mohammadi 2024. Impacts of Supplementation with Silymarin on Cardiovascular Risk Factors: A Systematic Review and Dose-Response Meta-Analysis<br>PMID 38671838 | Michele Schaffer: No saffron | Prisha Batra: Not clinical | – |

Table S2 continued

| Paper | Reviewer 1 | Reviewer 2 | Additional recorded reviews |
|---|---|---|---|
| 345. Ye 2024. Mitochondrial energy metabolism in diabetic cardiomyopathy: Physiological adaption, pathogenesis, and therapeutic targets<br>PMID 38527931 | Michele Schaffer: Not isolated | Prisha Batra: Not clinical | – |
| 346. Fauzi 2024. Hematological, biochemical, and histopathological evaluation of the Morus alba L. leaf extract from Brunei Darussalam: Acute toxicity study in ICR mice<br>PMID 38682142 | Michele Schaffer: No saffron | Prisha Batra: Not clinical | – |
| 347. #N/A 2024. 2024 International Congress on Integrative Medicine & Health Abstracts<br>PMID #N/A | Michele Schaffer: Not isolated | Prisha Batra: Not clinical | – |
| 348. Liu 2024. Targeting autophagy with natural products as a potential therapeutic approach for diabetic microangiopathy<br>PMID 38659578 | Michele Schaffer: Not isolated | Prisha Batra: Not clinical | – |
| 349. Olejarz 2024. Lipoprotein alterations in endocrine disorders - a review of the recent developments in the field<br>PMID 38628593 | Michele Schaffer: Not isolated | Prisha Batra: Not clinical | – |
| 350. Weerawatanakorn 2024. Exploring Phytochemical Mechanisms in the Prevention of Cholesterol Dysregulation: A Review<br>PMID 38517334 | Michele Schaffer: Not isolated | Prisha Batra: Not clinical | – |
| 351. Diab 2024. The Influence of Circadian Rhythm on the Antioxidant Capacity of Saliva in Periodontal Diseases<br>PMID 38618333 | Michele Schaffer: Not clinical | Prisha Batra: Not clinical | – |
| 352. Yaribeygi 2024. Modulating effects of crocin on lipids and lipoproteins: Mechanisms and potential benefits<br>PMID 38617922 | Michele Schaffer: Not clinical | Prisha Batra: Not clinical | – |
| 353. Hajivalizadeh 2024. The Role of Biotechnology in Latest Therapeutic Approaches for Diabetes Mellitus<br>PMID 38618507 | Michele Schaffer: Not isolated | Prisha Batra: No saffron | – |
| 354. Miryan 2024. Dietary inflammatory index in relation to the progression of hepatic steatosis and liver fibrosis: evaluation by elastography/Fibroscan<br>PMID 38589782 | Michele Schaffer: Not isolated | Prisha Batra: Not clinical | – |
| 355. Ma 2024. Jianpi Gushen Huayu decoction ameliorated diabetic nephropathy through modulating metabolites in kidney, and inhibiting TLR4/NF-$\kappa$B/NLRP3 and JNK/P38 pathways<br>PMID 38591083 | Michele Schaffer: Not isolated | Prisha Batra: Not clinical | – |

Table S2 continued

| Paper | Reviewer 1 | Reviewer 2 | Additional recorded reviews |
|---|---|---|---|
| 357. Rahimlou 2024. Association of maternal exposure to endocrine disruptor chemicals with cardio-metabolic risk factors in children during childhood: a systematic review and meta-analysis of cohort studies<br>PMID 38576015 | Michele Schaffer: Not isolated | Prisha Batra: Not clinical | – |
| 358. Shen 2024. Sirtuins in intervertebral disc degeneration: current understanding<br>PMID 38553713 | Michele Schaffer: Not isolated | Prisha Batra: Not clinical | – |
| 359. Zhang 2024. Integration of transcriptomics and metabolomics reveals toxicological mechanisms of ZhuRiHeng drop pill in the 180-day repeated oral toxicity study<br>PMID 38560353 | Michele Schaffer: Not isolated | Prisha Batra: Not clinical | – |
| 360. Sauvé 2024. Glycomacropeptide as an Efficient Agent to Fight Pathophysiological Mechanisms of Metabolic Syndrome<br>PMID 38542783 | Michele Schaffer: No saffron | Prisha Batra: No saffron | – |
| 361. Tonin 2024. Genetic and Transcriptomic Background of Oxidative Stress and Antioxidative Therapies in Late Complications of Type 2 Diabetes Mellitus: A Systematic Review<br>PMID 38539811 | Michele Schaffer: Not isolated | Prisha Batra: Not clinical | – |
| 362. Luo 2024. Unlocking the Power: New Insights into the Anti-Aging Properties of Mushrooms<br>PMID 38535223 | Michele Schaffer: No saffron | Prisha Batra: Not clinical | – |
| 363. Joukar 2024. ATP releasing channels and the ameliorative effects of high intensity interval training on diabetic heart: a multifaceted analysis<br>PMID 38532054 | Michele Schaffer: Not isolated | Prisha Batra: No saffron | – |
| 364. Mohamed 2024. Sourdough bread as nutritional intervention tool for improvement of cognitive dysfunction in diabetic rats<br>PMID 38528644 | Michele Schaffer: Not isolated | Prisha Batra: Not clinical | – |
| 365. Zhang 2024. Crosstalk among mitophagy, pyroptosis, ferroptosis, and necroptosis in central nervous system injuries<br>PMID 38103229 | Michele Schaffer: Not isolated | Prisha Batra: Not clinical | – |
| 366. Burelle 2024. A genetic mouse model of lean-NAFLD unveils sexual dimorphism in the liver-heart axis<br>PMID 38519536 | Michele Schaffer: No saffron | Prisha Batra: Not clinical | – |
| 367. Tong 2024. Integrated chemical characterization, metabolite profiling, and pharmacokinetics analysis of Zhijun Tangshen Decoction by UPLC-Q/TOF-MS<br>PMID 38523634 | Michele Schaffer: Not isolated | Prisha Batra: Not clinical | – |

Table S2 continued

| Paper | Reviewer 1 | Reviewer 2 | Additional recorded reviews |
|---|---|---|---|
| 368. Rahimlou 2024. Polyphenol consumption and Nonalcoholic fatty liver disease risk in adults<br>PMID 38514756 | Michele Schaffer: Not isolated | Prisha Batra: Not clinical | – |
| 369. Netzer 2024. Influence of nutrition and food on sleep-is there evidence?<br>PMID 37740061 | Michele Schaffer: No saffron | Prisha Batra: Not clinical | – |
| 370. Godos 2024. Underrated aspects of a true Mediterranean diet: understanding traditional features for worldwide application of a "Planeterranean" diet<br>PMID 38515140 | Michele Schaffer: No saffron | Prisha Batra: Not clinical | – |
| 371. Saberi 2024. Exercise training improves diabetic renal injury by reducing fetuin-A, oxidative stress and inflammation in type 2 diabetic rats<br>PMID 38510054 | Michele Schaffer: Not isolated | Prisha Batra: Not clinical | – |
| 372. Aghaei-Zarch 2024. Crosstalk between MiRNAs/lncRNAs and PI3K/AKT signaling pathway in diabetes mellitus: Mechanistic and therapeutic perspectives<br>PMID 38511053 | Michele Schaffer: Not isolated | Prisha Batra: Not clinical | – |
| 373. Hasnat 2024. Flavonoids: A treasure house of prospective pharmacological potentials<br>PMID 38496846 | Michele Schaffer: Not isolated | Prisha Batra: Not clinical | – |
| 374. Manojkumar 2024. Phytochemical Screening, In Silico Molecular Docking, ADME Properties, and In Vitro Antioxidant, Anticancer, and Antidiabetic Activity of Marine Halophyte Suaeda maritima (L.) Dumort<br>PMID 38496978 | Michele Schaffer: Not isolated | Prisha Batra: Not clinical | – |
| 375. Rezaie 2024. Melatonin reduces lung injury in type 1 diabetic mice by the modulation of autophagy<br>PMID 38486170 | Michele Schaffer: Not isolated | Prisha Batra: Not clinical | – |
| 376. Papanikolaou 2024. Characterization of the Sideritis scardica Extract SidTea+(TM) and Its Effect on Physiological Profile, Metabolic Health and Redox Biomarkers in Healthy Adults: A Randomized, Double-Blind, Placebo-Controlled Study<br>PMID 38474625 | Michele Schaffer: Not isolated | Prisha Batra: No saffron | – |
| 377. Jin 2024. Exosomes as nanostructures deliver miR-204 in alleviation of mitochondrial dysfunction in diabetic nephropathy through suppressing methyltransferase-like 7A-mediated CIDEC N6-methyladenosine methylation<br>PMID 38334961 | Michele Schaffer: Not isolated | Prisha Batra: Not clinical | – |

Table S2 continued

| Paper | Reviewer 1 | Reviewer 2 | Additional recorded reviews |
|---|---|---|---|
| 378. Lucini Mas 2024. The Effects of Chia Defatted Flour as a Nutritional Supplement in C57BL/6 Mice Fed a Low-Quality Diet<br>PMID 38472791 | Michele Schaffer: Not isolated | Prisha Batra: Not clinical | – |
| 379. Wongmanee 2023. Antihyperglycemic effect of extra virgin sacha inchi oil in type 2 diabetic rats: Mechanisms involved in pancreatic $\beta$-cell function and apoptosis<br>PMID 38481551 | Michele Schaffer: Not isolated | Prisha Batra: Not clinical | – |
| 380. Oladimeji 2023. Antiobesity effect of healthy food crops and functional foods: A systematic review of their mechanisms<br>PMID 38455221 | Michele Schaffer: Not isolated | Prisha Batra: Not clinical | – |
| 381. Kunnumakkara 2023. Role of Turmeric and Curcumin in Prevention and Treatment of Chronic Diseases: Lessons Learned from Clinical Trials<br>PMID 37082752 | Michele Schaffer: Not isolated | Prisha Batra: Not clinical | – |
| 382. Piñón-Simental 2024. Use of Callistemon citrinus as a gastroprotective and anti-inflammatory agent on indomethacin-induced gastric ulcers in obese rats<br>PMID 38435992 | Michele Schaffer: Not isolated | Prisha Batra: Not clinical | – |
| 383. Ademowo 2024. Effects of carotenoids on mitochondrial dysfunction<br>PMID 38385583 | Michele Schaffer: Not isolated | Prisha Batra: Not clinical | – |
| 384. Ursic-Bedoya 2024. FGF19 and its analog Aldafermin cooperate with MYC to induce aggressive hepatocarcinogenesis<br>PMID 38228803 | Michele Schaffer: No saffron | Prisha Batra: Not clinical | – |
| 385. Wang 2024. Crocin protects against endoplasmic reticulum stress-related tubular injury in diabetic nephropathy via the activation of the PI3K/AKT/Nrf2 pathway<br>PMID 38419890 | Michele Schaffer: Not clinical | Prisha Batra: Not clinical | – |
| 386. Saadat 2024. An overview of pharmacological effects of Crocus sativous and its constituents<br>PMID 38419885 | Michele Schaffer: Not clinical | Prisha Batra: Not clinical | – |
| 387. Dong 2024. Metabolic memory: mechanisms and diseases<br>PMID 38413567 | Michele Schaffer: Not isolated | Prisha Batra: Not clinical | – |
| 388. Xing 2024. The Expression of miR-377-3p in Patients with DKD and the Regulatory Mechanism of miR-377-3p on the Inflammatory Response of HK-2 Cells Through TGF-$\beta$<br>PMID 38414866 | Michele Schaffer: Not isolated | Prisha Batra: Not clinical | – |

Table S2 continued

| Paper | Reviewer 1 | Reviewer 2 | Additional recorded reviews |
|---|---|---|---|
| 389. Park 2024. Association of a High Healthy Eating Index Diet with Long-Term Visceral Fat Loss in a Large Longitudinal Study<br>PMID 38398858 | Michele Schaffer: No saffron | Prisha Batra: Not clinical | – |
| 390. Gudović 2024. The Comparative Effects of Myo-Inositol and Metformin Therapy on the Clinical and Biochemical Parameters of Women of Normal Weight Suffering from Polycystic Ovary Syndrome<br>PMID 38397951 | Michele Schaffer: Not isolated | Prisha Batra: No saffron | – |
| 391. Peng 2024. Hesperitin-Copper(II) Complex Regulates the NLRP3 Pathway and Attenuates Hyperuricemia and Renal Inflammation<br>PMID 38397567 | Michele Schaffer: Not isolated | Prisha Batra: Not clinical | – |
| 392. Dibdiakova 2024. Membrane Separation of Chicken Byproduct Hydrolysate for Up-Concentration of Bioactive Peptides<br>PMID 38392655 | Michele Schaffer: No saffron | Prisha Batra: Not clinical | – |
| 393. Chand 2024. Unveiling the potential of linseed mucilage, its health benefits, and applications in food packaging<br>PMID 38385008 | Michele Schaffer: Not isolated | Prisha Batra: Not clinical | – |
| 394. Azarbayjani 2024. Association of dietary inflammatory index with C-reactive protein and interleukin-6 in women with and without polycystic ovarian syndrome<br>PMID 38368454 | Michele Schaffer: Not isolated | Prisha Batra: Not clinical | – |
| 395. Wee 2023. Linking Diabetes to Alzheimer's Disease: Potential Roles of Glucose Metabolism and Alpha-Glucosidase<br>PMID 36372924 | Michele Schaffer: Not isolated | Prisha Batra: Not clinical | – |
| 396. Zhang 2024. Gut microbiota dysbiosis and decreased levels of acetic and propionic acid participate in glucocorticoid-induced glycolipid metabolism disorder<br>PMID 38226811 | Michele Schaffer: Not isolated | Prisha Batra: Not clinical | – |
| 397. Meng 2024. Multi-ancestry genome-wide association study of major depression aids locus discovery, fine mapping, gene prioritization and causal inference<br>PMID 38177345 | Michele Schaffer: Text not found | Prisha Batra: Not clinical | – |
| 398. Shieh 2023. Dietary Inflammatory Index and Fractures in Midlife Women: Study of Women's Health Across the Nation<br>PMID 36780235 | Michele Schaffer: No saffron | Prisha Batra: Not clinical | – |

Table S2 continued

| Paper | Reviewer 1 | Reviewer 2 | Additional recorded reviews |
|---|---|---|---|
| 399. Li 2024. Combined gestational age and serum fucose for early prediction of risk for bronchopulmonary dysplasia in premature infants<br>PMID 38347448 | Michele Schaffer: Not isolated | Prisha Batra: Not clinical | – |
| 400. Uzzan 2024. Chronic Treatment with Nigella sativa Oil Exerts Antimanic Properties and Reduces Brain Inflammation in Rats<br>PMID 38339101 | Michele Schaffer: Not isolated | Prisha Batra: Not clinical | – |
| 401. Bartkowiak-Wieczorek 2024. Natural Products and Health<br>PMID 38337699 | Michele Schaffer: No saffron | Prisha Batra: Not clinical | – |
| 402. Aljuraiban 2024. The Role of Diet in the Prevention of Hypertension and Management of Blood Pressure: An Umbrella Review of Meta-Analyses of Interventional and Observational Studies<br>PMID 37783307 | Michele Schaffer: Not isolated | Prisha Batra: Not clinical | – |
| 403. Chen 2023. Crocin Ameliorates Diabetic Nephropathy through Regulating Metabolism, CYP4A11/PPAR$\gamma$, and TGF-$\beta$/Smad Pathways in Mice<br>PMID 37936469 | Michele Schaffer: Not clinical | Prisha Batra: Not clinical | – |
| 404. Mishra 2023. A Review on the Protective Effects of Probiotics against Alzheimer's Disease<br>PMID 38248439 | Michele Schaffer: Not isolated | Prisha Batra: Not clinical | – |
| 405. Xia 2024. Effect of Different Medium-Chain Triglycerides on Glucose Metabolism in High-Fat-Diet Induced Obese Rats<br>PMID 38254542 | Michele Schaffer: Not isolated | Prisha Batra: Not clinical | – |
| 406. Zhu 2024. A Literature Review of the Pharmacological Effects of Jujube<br>PMID 38254493 | Michele Schaffer: Not isolated | Prisha Batra: Not clinical | – |
| 407. Dhillon 2024. Hydrogen Water: Extra Healthy or a Hoax?-A Systematic Review<br>PMID 38256045 | Michele Schaffer: Not isolated | Prisha Batra: Not clinical | – |
| 408. Geremew 2024. Green Synthesis of Novel Silver Nanoparticles Using Salvia blepharophylla and Salvia greggii: Antioxidant and Antidiabetic Potential and Effect on Foodborne Bacterial Pathogens<br>PMID 38255978 | Michele Schaffer: Not isolated | Prisha Batra: Not clinical | – |

Table S2 continued

| Paper | Reviewer 1 | Reviewer 2 | Additional recorded reviews |
|---|---|---|---|
| 409. Wahid 2024. Exploring the Therapeutic Potential of Natural Products in Polycystic Ovarian Syndrome (PCOS): A Mini-Review of Lipid Profile, Blood Glucose, and Ovarian Histological Improvements<br>PMID 38276279 | Michele Schaffer: Not isolated | Prisha Batra: Not clinical | – |
| 410. #N/A 2023. RESEARCH COMMUNICATIONS OF THE 33<br>PMID 38053473 | Michele Schaffer: No saffron | Prisha Batra: Not clinical | – |
| 411. Aslam 2024. Polyherbal extract improves glycometabolic control in alloxan-induced diabetic rats via down-regulating the MAPK/JNK pathway, modulating Nrf-2/Keap-1 expression, and stimulating insulin signaling<br>PMID 38234664 | Michele Schaffer: Not isolated | Prisha Batra: Not clinical | – |
| 412. Gardin 2024. A functional mini-GDE transgene corrects impairment in models of glycogen storage disease type III<br>PMID 38015640 | Michele Schaffer: No saffron | Prisha Batra: Not clinical | – |
| 413. Khan 2023. Mechanism of Antitumor Effects of Saffron in Human Prostate Cancer Cells<br>PMID 38201944 | Michele Schaffer: Not clinical | Prisha Batra: Not clinical | – |
| 414. Zheng 2023. Diet Quality and Mortality among Chinese Adults: Findings from the China Health and Nutrition Survey<br>PMID 38201925 | Michele Schaffer: No saffron | Prisha Batra: Not clinical | – |
| 415. Bartel 2023. Nutraceutical Properties of Syringic Acid in Civilization Diseases-Review<br>PMID 38201840 | Michele Schaffer: Not isolated | Prisha Batra: Not clinical | – |
| 416. Łanoszka 2023. Natural Sirtuin1 Activators and Atherosclerosis: an Overview<br>PMID 38038821 | Michele Schaffer: Not isolated | Prisha Batra: Not clinical | – |
| 417. Topçu-Tarladaçalışır 2024. Quercetin Attenuates Endoplasmic Reticulum Stress and Apoptosis in TNBS-Induced Colitis by Inhibiting the Glucose Regulatory Protein 78 Activation<br>PMID 38173174 | Michele Schaffer: Not isolated | Prisha Batra: Not clinical | – |
| 418. Mohammed 2023. Enhancing date seed phenolic bioaccessibility in soft cheese through a dehydrated liposome delivery system and its effect on testosterone-induced benign prostatic hyperplasia in rats<br>PMID 38178973 | Michele Schaffer: Not clinical | Prisha Batra: Not clinical | – |

Table S2 continued

| Paper | Reviewer 1 | Reviewer 2 | Additional recorded reviews |
|---|---|---|---|
| 419. Olivares-Ferretti 2023. Exploring the metabolic and antioxidant potential of solergy: Implications for enhanced animal production<br>PMID 38173966 | Michele Schaffer: Not isolated | Prisha Batra: Not clinical | – |
| 420. Appell 2024. Ginger Supplementation Attenuated Mitochondrial Fusion and Improved Skeletal Muscle Size in Type 2 Diabetic Rats<br>PMID 38148056 | Michele Schaffer: Not isolated | Prisha Batra: Not clinical | – |
| 421. Ceci 2024. Impact of Physical Exercise on Melanoma Hallmarks: Current Status of Preclinical and Clinical Research<br>PMID 38164270 | Michele Schaffer: Not isolated | Prisha Batra: Not clinical | – |
| 422. Shabab 2023. Diabetic cardiomyopathy in rats was attenuated by endurance exercise through the inhibition of inflammation and apoptosis<br>PMID 38163155 | Michele Schaffer: Not isolated | Prisha Batra: Not clinical | – |
| 423. Potra Cicalău 2023. Assessing the Antioxidant Benefits of Topical Carvacrol and Magnolol Periodontal Hydrogel Therapy in Periodontitis Associated with Diabetes in Wistar Rats<br>PMID 38132422 | Michele Schaffer: Not isolated | Prisha Batra: Not clinical | – |
| 424. Gostimirovic 2023. Resveratrol and Gut Microbiota Synergy: Preventive and Therapeutic Effects<br>PMID 38139400 | Michele Schaffer: Not isolated | Prisha Batra: Not clinical | – |
| 425. Grant 2023. Diet's Role in Modifying Risk of Alzheimer's Disease: History and Present Understanding<br>PMID 37955087 | Michele Schaffer: Not isolated | Prisha Batra: Not clinical | – |
| 426. Mikhailova 2023. Selenium Nanoparticles: Green Synthesis and Biomedical Application<br>PMID 38138613 | Michele Schaffer: Not isolated | Prisha Batra: Not clinical | – |
| 427. Biolcatti 2023. Pregnancy Protects against Abnormal Gut Permeability Promoted via the Consumption of a High-Fat Diet in Mice<br>PMID 38140300 | Michele Schaffer: Not isolated | Prisha Batra: Not clinical | – |
| 428. Yoon 2023. Aralia continentalis Root Enhances Non-Rapid Eye Movement Sleep by Activating GABA(A) Receptors<br>PMID 38140279 | Michele Schaffer: Not isolated | Prisha Batra: Not clinical | – |
| 429. Esenbuga 2023. Dietary Effects of Some Plant Extracts on Laying Performance, Egg Quality, and Some Blood Parameters in Laying Hens at Different Cage Densities<br>PMID 38136903 | Michele Schaffer: Not isolated | Prisha Batra: Not clinical | – |

Table S2 continued

| Paper | Reviewer 1 | Reviewer 2 | Additional recorded reviews |
|---|---|---|---|
| 430. Cheng 2023. Advances in embedding techniques of anthocyanins: Improving stability, bioactivity and bioavailability<br>PMID 38144721 | Michele Schaffer: Not isolated | Prisha Batra: Not clinical | – |
| 431. Gugsa 2023. Hepatoprotective effect of hydromethanol extract of Otostegia integrifolia benth leaves in isoniazid and rifampicin induced Swiss albino mice<br>PMID 38115863 | Michele Schaffer: Not isolated | Prisha Batra: Not clinical | – |
| 432. Pourmousavi 2023. Effect of crocin and treadmill exercise on oxidative stress and heart damage in diabetic rats<br>PMID 38113243 | Michele Schaffer: Not isolated | Prisha Batra: Not clinical | – |
| 433. Morshedzadeh 2023. A narrative review on the role of hesperidin on metabolic parameters, liver enzymes, and inflammatory markers in nonalcoholic fatty liver disease<br>PMID 38107097 | Michele Schaffer: Not isolated | Prisha Batra: Not clinical | – |
| 434. Khorasanian 2023. Hesperidin reduces depressive symptoms in post-coronary artery bypass graft patients with mild depression<br>PMID 38107143 | Michele Schaffer: Not isolated | Prisha Batra: Not clinical | – |
| 435. Forouzanfar 2023. Protective effects of fruit extract of Rosa canina and quercetin on human umbilical vein endothelial cell injury induced by hydrogen peroxide<br>PMID 38107098 | Michele Schaffer: Not isolated | Prisha Batra: Not clinical | – |
| 436. Bohn 2023. Carotenoids in Health as Studied by Omics-Related Endpoints<br>PMID 37678712 | Michele Schaffer: Not isolated | Prisha Batra: Not clinical | – |
| 437. #N/A #N/A. Medicinal plants and foods with metaphorical concepts in Rumi's "<br>PMID 38106633 | Michele Schaffer: Not isolated | Prisha Batra: Not clinical | – |
| 438. Mackonochie 2023. A Scoping Review of the Clinical Evidence for the Health Benefits of Culinary Doses of Herbs and Spices for the Prevention and Treatment of Metabolic Syndrome<br>PMID 38068725 | Michele Schaffer: Not isolated | Prisha Batra: Not clinical | – |
| 439. Islam 2023. Genus Amorphophallus: A Comprehensive Overview on Phytochemistry, Ethnomedicinal Uses, and Pharmacological Activities<br>PMID 38068582 | Michele Schaffer: Not isolated | Prisha Batra: Not clinical | – |

Table S2 continued

| Paper | Reviewer 1 | Reviewer 2 | Additional recorded reviews |
|---|---|---|---|
| 440. Li 2023. Association between dietary antioxidant indices and glaucoma in the National Health and Nutrition Examination Survey<br>PMID 38075225 | Michele Schaffer: Not isolated | Prisha Batra: Not clinical | – |
| 441. Yan 2023. Gut liver brain axis in diseases: the implications for therapeutic interventions<br>PMID 38057297 | Michele Schaffer: Not isolated | Prisha Batra: Not clinical | – |
| 442. Niu 2023. Treatment of obesity by acupuncture combined with medicine based on pathophysiological mechanism: A review<br>PMID 38050318 | Michele Schaffer: Not isolated | Prisha Batra: Not clinical | – |
| 443. Huang 2023. Exploring global research status and trends in anti-obesity effects of traditional Chinese medicine through intestinal microbiota: a bibliometric study<br>PMID 38045760 | Michele Schaffer: Not isolated | Prisha Batra: Not clinical | – |
| 444. Wang 2023. Combined analysis of inorganic elements and flavonoid metabolites reveals the relationship between flower quality and maturity of Sophora japonica L<br>PMID 38046598 | Michele Schaffer: Not isolated | Prisha Batra: Not clinical | – |
| 445. Bays 2022. Obesity Pillars Roundtable: Obesity and South Asians<br>PMID 37990701 | Michele Schaffer: Not isolated | Prisha Batra: Not clinical | – |
| 446. Vitiello 2023. Herbal and Natural Treatments for the Management of the Glaucoma: An Update<br>PMID 38027044 | Michele Schaffer: Not isolated | Prisha Batra: Not clinical | – |
| 447. Rai 2022. Insights on the role of anti-inflammatory and immunosuppressive agents in the amelioration of diabetes<br>PMID 37090130 | Michele Schaffer: Not isolated | Prisha Batra: Not clinical | – |
| 448. Supriya 2023. Astaxanthin Supplementation Augments the Benefits of CrossFit Workouts on Semaphorin 3C and Other Adipokines in Males with Obesity<br>PMID 38004197 | Michele Schaffer: Not isolated | Prisha Batra: No saffron | – |
| 449. Kolahi Ahari 2023. Serum uric acid to high-density lipoprotein ratio as a novel indicator of inflammation is correlated with the presence and severity of metabolic syndrome: A large-scale study<br>PMID 37605374 | Michele Schaffer: Not isolated | Prisha Batra: Not clinical | – |
| 450. Jafari 2023. The effects of purslane consumption on lipid profile and C-reactive protein: A systematic review and dose-response meta-analysis<br>PMID 37970383 | Michele Schaffer: Not isolated | Prisha Batra: Not clinical | – |

Table S2 continued

| Paper | Reviewer 1 | Reviewer 2 | Additional recorded reviews |
|---|---|---|---|
| 451. Noei Razliqi 2023. Hepatoprotective effects of gentisic acid through its anti-oxidant properties in nicotinamide-streptozotocin induced diabetic mice PMID 37970436 | Michele Schaffer: Not isolated | Prisha Batra: Not clinical | – |
| 452. Mamsharifi 2023. Nano-curcumin effects on nicotine dependence, depression, anxiety and metabolic parameters in smokers: A randomized double-blind clinical study PMID 37954269 | Michele Schaffer: Not isolated | Prisha Batra: No saffron | – |
| 453. Wahab 2023. Potential Pharmacological Applications of Nigella Seeds with a Focus on Nigella sativa and Its Constituents against Chronic Inflammatory Diseases: Progress and Future Opportunities PMID 38005726 | Michele Schaffer: Not isolated | Prisha Batra: Not clinical | – |
| 454. Rodriguez-Amaya 2023. Comprehensive Update on Carotenoid Colorants from Plants and Microalgae: Challenges and Advances from Research Laboratories to Industry PMID 38002140 | Michele Schaffer: Not isolated | Prisha Batra: Not clinical | – |
| 455. Wang 2023. Signaling pathways and targeted therapeutic strategies for polycystic ovary syndrome PMID 37929034 | Michele Schaffer: Not isolated | Prisha Batra: Not clinical | – |
| 456. Sinha 2023. A synergistic blend of Garcinia mangostana fruit rind and Cinnamomum tamala leaf extracts enhances myogenic differentiation and mitochondrial biogenesis in vitro and muscle growth and strength in mice PMID 37920678 | Michele Schaffer: Not isolated | Prisha Batra: Not clinical | – |
| 457. Li 2023. Optimizing diabetic kidney disease animal models: Insights from a meta-analytic approach PMID 37723622 | Michele Schaffer: Not isolated | Prisha Batra: Not clinical | – |
| 458. Naraki 2023. The effect of ellagic acid on the metabolic syndrome: A review article PMID 38027887 | Grace Kim: No saffron | Michele Schaffer: Not isolated | Prisha Batra: Not clinical; Alexander Tai Nguyen: No saffron |
| 459. Hawash 2023. Characterization and Investigation of Novel Benzodioxol Derivatives as Antidiabetic Agents: An In Vitro and In Vivo Study in an Animal Model PMID 37892167 | Grace Kim: No saffron | Michele Schaffer: No saffron | Prisha Batra: Not clinical; Alexander Tai Nguyen: Not clinical |
| 460. Ishigooka 2023. Effects of Angiotensin Receptor Blockers on Streptozotocin-Induced Diabetic Cataracts PMID 37892765 | Grace Kim: No saffron | Michele Schaffer: Not isolated | Prisha Batra: Not clinical; Alexander Tai Nguyen: No saffron |
| 461. Nishida 2023. Astaxanthin: Past, Present, and Future PMID 37888449 | Grace Kim: No saffron | Alexander Tai Nguyen: No saffron | – |

Table S2 continued

| Paper | Reviewer 1 | Reviewer 2 | Additional recorded reviews |
|---|---|---|---|
| 462. Souza 2023. "Does Physical Exercise Promote Health Benefits for Diabetic Patients during the COVID-19 Pandemic?": A Systematic Review<br>PMID 37888519 | Grace Kim: No saffron | Alexander Tai Nguyen: No saffron | – |
| 463. Salem 2023. Effect of Nigella sativa on general health and immune system in young healthy volunteers; a randomized, placebo-controlled, double-blinded clinical trial<br>PMID 37901256 | Grace Kim: No saffron | Alexander Tai Nguyen: No saffron | – |
| 464. Ebrahimnezhad 2023. High-intensity interval training reduced oxidative stress and apoptosis in the hippocampus of male rats with type 2 diabetes: The role of the PGC1$\alpha$-Keap1-Nrf2 signaling pathway<br>PMID 37885999 | Grace Kim: No saffron | Alexander Tai Nguyen: No saffron | – |
| 465. Jung 2023. Effects of natural products on polycystic ovary syndrome: From traditional medicine to modern drug discovery<br>PMID 37867816 | Grace Kim: No saffron | Alexander Tai Nguyen: No saffron | – |
| 466. Hur 2023. Interaction of energy and sulfur microbial diet and smoking status with polygenic variants associated with lipoprotein metabolism<br>PMID 37860035 | Grace Kim: No saffron | Alexander Tai Nguyen: No saffron | – |
| 467. Dadaei 2023. Dietary inflammatory index in relation to severe coronary artery disease in Iranian adults<br>PMID 37841398 | Grace Kim: No saffron | Alexander Tai Nguyen: No saffron | – |
| 468. Piao 2023. Antidepressant-like Effects of Representative Types of Food and Their Possible Mechanisms<br>PMID 37836833 | Grace Kim: No saffron | Alexander Tai Nguyen: No saffron | – |
| 469. Ijinu 2023. Anthocyanins as Immunomodulatory Dietary Supplements: A Nutraceutical Perspective and Micro-/Nano-Strategies for Enhanced Bioavailability<br>PMID 37836436 | Grace Kim: No saffron | Alexander Tai Nguyen: No saffron | – |
| 470. Zhang 2023. Rosa laevigata Michx. Polysaccharide Ameliorates Diabetic Nephropathy in Mice through Inhibiting Ferroptosis and PI3K/AKT Pathway-Mediated Apoptosis and Modulating Tryptophan Metabolism<br>PMID 37840577 | Grace Kim: No saffron | Alexander Tai Nguyen: No saffron | – |
| 471. Hassan 2023. Effect of PENN-DIABEX, a novel polyherbal formulation, in high fat diet streptozotocin-induced diabetic rats<br>PMID 37841663 | Grace Kim: No saffron | Alexander Tai Nguyen: No saffron | – |

Table S2 continued

| Paper | Reviewer 1 | Reviewer 2 | Additional recorded reviews |
|---|---|---|---|
| 472. Bond 2023. Validation of Igls Criteria for Islet Transplant Functional Status Using Person-Reported Outcome Measures in a Cross-Sectional Study<br>PMID 37822448 | Grace Kim: No saffron | Alexander Tai Nguyen: No saffron | – |
| 473. Zahedi 2023. Association of baseline and changes in adiponectin, homocysteine, high-sensitivity C-reactive protein, interleukin-6, and interleukin-10 levels and metabolic syndrome incidence: Tehran lipid and glucose study<br>PMID 37809533 | Grace Kim: No saffron | Alexander Tai Nguyen: No saffron | – |
| 474. Tschiderer 2023. Age at Menopause and the Risk of Stroke: Observational and Mendelian Randomization Analysis in 204 244 Postmenopausal Women<br>PMID 37681566 | Grace Kim: No saffron | Alexander Tai Nguyen: No saffron | – |
| 475. Xu 2023. Oxidative stress signaling in the pathogenesis of diabetic cardiomyopathy and the potential therapeutic role of antioxidant naringenin<br>PMID 37747066 | Grace Kim: No saffron | Alexander Tai Nguyen: No saffron | – |
| 476. Zhao 2023. The Associations between Organophosphate Pesticides (OPs) and Respiratory Disease, Diabetes Mellitus, and Cardiovascular Disease: A Review and Meta-Analysis of Observational Studies<br>PMID 37755752 | Grace Kim: No saffron | Alexander Tai Nguyen: No saffron | – |
| 477. Maiese 2023. The Metabolic Basis for Nervous System Dysfunction in Alzheimer's Disease, Parkinson's Disease, and Huntington's Disease<br>PMID 37488757 | Grace Kim: No saffron | Alexander Tai Nguyen: No saffron | – |
| 478. Sharifan 2023. The effects of low-fat dairy products fortified with 1500 IU vitamin D(3) on serum liver function biomarkers in adults with abdominal obesity: a randomized controlled trial<br>PMID 37749703 | Grace Kim: No saffron | Alexander Tai Nguyen: No saffron | – |
| 479. Li 2023. The directed acyclic graph helped identify confounders in the association between coronary heart disease and pesticide exposure among greenhouse vegetable farmers<br>PMID 37746981 | Grace Kim: No saffron | Alexander Tai Nguyen: No saffron | – |
| 480. Chhimwal 2023. Metagenomic signatures reveal the key role of phloretin in amelioration of gut dysbiosis attributed to metabolic dysfunction-associated fatty liver disease by time-dependent modulation of gut microbiome<br>PMID 37744933 | Grace Kim: No saffron | Alexander Tai Nguyen: No saffron | – |

Table S2 continued

| Paper | Reviewer 1 | Reviewer 2 | Additional recorded reviews |
|---|---|---|---|
| 481. Esmaealzadeh 2023. Pharmacological effects of Safranal: An updated review<br>PMID 37736506 | Grace Kim: No saffron | Alexander Tai Nguyen: No saffron | – |
| 482. Chen 2023. Mechanism, prevention and treatment of cognitive impairment caused by high altitude exposure<br>PMID 37731540 | Grace Kim: No saffron | Alexander Tai Nguyen: No saffron | – |
| 483. Torres-Sánchez 2023. Effect of pesticides on phosphorylation of tau protein, and its influence on Alzheimer's disease<br>PMID 37727721 | Grace Kim: No saffron | Alexander Tai Nguyen: No saffron | – |
| 484. Mohsin 2023. Prospective Nutraceutical Effects of Cinnamon Derivatives Against Insulin Resistance in Type II Diabetes Mellitus-Evidence From the Literature<br>PMID 37701673 | Grace Kim: No saffron | Alexander Tai Nguyen: No saffron | – |
| 485. Tahmouzi 2023. Application of guar (Cyamopsis tetragonoloba L.) gum in food technologies: A review of properties and mechanisms of action<br>PMID 37701200 | Grace Kim: No saffron | Alexander Tai Nguyen: No saffron | – |
| 486. Xu 2023. Trends in metabolic dysfunction in polycystic ovary syndrome: a bibliometric analysis<br>PMID 37701893 | Grace Kim: No saffron | Alexander Tai Nguyen: No saffron | – |
| 487. Luo 2023. Integrating serum metabolomics and network analysis to explore the antidepressant activity of crocin in rats with chronic unexpected mild stress-induced depression<br>PMID 37688460 | Grace Kim: No saffron | Alexander Tai Nguyen: Not clinical | – |
| 488. Bukvicki 2023. Essential Oils and Their Combination with Lactic Acid Bacteria and Bacteriocins to Improve the Safety and Shelf Life of Foods: A Review<br>PMID 37685221 | Grace Kim: No saffron | Alexander Tai Nguyen: No saffron | – |
| 489. Gil 2023. Investigation into Polyphenol Profile and Biological Activities of Enriched Persimmon/Apple Smoothies during Storage<br>PMID 37685183 | Bryan Anh Nguyen: No saffron | Pranav Potluri: No saffron | Alexander Tai Nguyen: No saffron |
| 490. Kim 2023. Interactions between Polygenetic Variants and Lifestyle Factors in Hypothyroidism: A Hospital-Based Cohort Study<br>PMID 37686882 | Bryan Anh Nguyen: No saffron | Pranav Potluri: No saffron | Alexander Tai Nguyen: No saffron |
| 491. Al-Ishaq 2023. Sweeteners and the Gut Microbiome: Effects on Gastrointestinal Cancers<br>PMID 37686707 | Bryan Anh Nguyen: No saffron | Pranav Potluri: No saffron | Alexander Tai Nguyen: No saffron |

Table S2 continued

| Paper | Reviewer 1 | Reviewer 2 | Additional recorded reviews |
|---|---|---|---|
| 492. Jomova 2023. Reactive oxygen species, toxicity, oxidative stress, and antioxidants: chronic diseases and aging PMID 37597078 | Bryan Anh Nguyen: No saffron | Pranav Potluri: No saffron | Alexander Tai Nguyen: No saffron |
| 493. Alsharif 2023. Melatonin downregulates the increased hepatic alpha-fetoprotein expression and restores pancreatic beta cells in a streptozotocin-induced diabetic rat model: a clinical, biochemical, immunohistochemical, and descriptive histopathological study PMID 37655263 | Bryan Anh Nguyen: No saffron | Pranav Potluri: No saffron | Alexander Tai Nguyen: No saffron |
| 494. Ruggieri 2023. Comprehensive Extraction and Chemical Characterization of Bioactive Compounds in Tepals of Crocus sativus L PMID 37630227 | Bryan Anh Nguyen: Not clinical | Pranav Potluri: No saffron | Alexander Tai Nguyen: No saffron |
| 495. Rojas-Aguilar 2023. The Additive Antinociceptive Effect of Resveratrol and Ketorolac in the Formalin Test in Mice PMID 37630993 | Bryan Anh Nguyen: No saffron | Pranav Potluri: No saffron | Alexander Tai Nguyen: No saffron |
| 496. Piotrowska 2023. Physical Activity as a Modern Intervention in the Fight against Obesity-Related Inflammation in Type 2 Diabetes Mellitus and Gestational Diabetes PMID 37627482 | Bryan Anh Nguyen: Not clinical | Pranav Potluri: No saffron | Alexander Tai Nguyen: No saffron |
| 497. Raut 2023. Hypometabolism, Alzheimer's Disease, and Possible Therapeutic Targets: An Overview PMID 37626828 | Bryan Anh Nguyen: No saffron | Pranav Potluri: No saffron | Alexander Tai Nguyen: No saffron |
| 498. González-Domínguez Á 2023. Altered insulin secretion dynamics relate to oxidative stress and inflammasome activation in children with obesity and insulin resistance PMID 37599368 | Bryan Anh Nguyen: No saffron | Pranav Potluri: No saffron | Alexander Tai Nguyen: No saffron |
| 499. Zhao 2023. Multi-omics reveals the mechanisms underlying Lactiplantibacillus plantarum P8-mediated attenuation of oxidative stress in broilers challenged with dexamethasone PMID 37600839 | Bryan Anh Nguyen: No saffron | Pranav Potluri: No saffron | Alexander Tai Nguyen: No saffron |
| 500. Wu 2023. PECAM-1 drives $\beta$-catenin-mediated EndMT via internalization in colon cancer with diabetes mellitus PMID 37580771 | Bryan Anh Nguyen: No saffron | Alexander Tai Nguyen: No saffron | – |
| 501. Li 2023. Effects of Shenlian formula on microbiota and inflammatory cytokines in adults with type 2 diabetes: a double-blind randomized clinical trial PMID 37454261 | Bryan Anh Nguyen: No saffron | Alexander Tai Nguyen: No saffron | – |

Table S2 continued

| Paper | Reviewer 1 | Reviewer 2 | Additional recorded reviews |
|---|---|---|---|
| 502. Cesaro 2023. Visceral adipose tissue and residual cardiovascular risk: a pathological link and new therapeutic options<br>PMID 37576108 | Bryan Anh Nguyen: No saffron | Alexander Tai Nguyen: No saffron | – |
| 503. Darenskaya 2023. Diabetic Nephropathy: Significance of Determining Oxidative Stress and Opportunities for Antioxidant Therapies<br>PMID 37569752 | Bryan Anh Nguyen: No saffron | Alexander Tai Nguyen: No saffron | – |
| 504. Tiwari 2023. Network pharmacology-based strategic prediction and target identification of apocarotenoids and carotenoids from standardized Kashmir saffron (Crocus sativus L.) extract against polycystic ovary syndrome<br>PMID 37565925 | Bryan Anh Nguyen: No saffron | Alexander Tai Nguyen: No saffron | – |
| 505. Ishiwata 2023. Recent Progress in 1,2-cis glycosylation for Glucan Synthesis<br>PMID 37570614 | Bryan Anh Nguyen: No saffron | Alexander Tai Nguyen: No saffron | – |
| 506. Persson 2023. Impact of an Okinawa/Nordic based diet on endocrinological and periodontal conditions in individuals with type 2 diabetes. A randomized case-control study<br>PMID 37553617 | Bryan Anh Nguyen: No saffron | Alexander Tai Nguyen: Not isolated | – |
| 507. Mathew 2023. Phosphorylation of insulin receptor substrates (IRS-1 and IRS-2) is attenuated following cecal ligation and puncture in mice<br>PMID 37550630 | Bryan Anh Nguyen: No saffron | Alexander Tai Nguyen: No saffron | – |
| 508. Ejaz 2023. A comprehensive review of summer savory (Satureja hortensis L.): promising ingredient for production of functional foods<br>PMID 37554989 | Bryan Anh Nguyen: No saffron | Alexander Tai Nguyen: No saffron | – |
| 509. Rahimpour 2023. The Effect of Training Type on The Signaling P athway of Ceramide-Dependent Insulin Resistance in The Flexor Hallucis Longus Muscle of Streptozotocin-Induced Diabetic Rats<br>PMID 37543859 | Bryan Anh Nguyen: No saffron | Alexander Tai Nguyen: No saffron | – |
| 510. Li 2023. Acupuncture improves immunity and fatigue after chemotherapy in breast cancer patients by inhibiting the Leptin/AMPK signaling pathway<br>PMID 37542585 | Bryan Anh Nguyen: No saffron | Alexander Tai Nguyen: No saffron | – |
| 511. Kokts-Porietis 2023. Prospective Cohort of Pre- and Post-Diagnosis Diet with Survival Outcomes: an Alberta Endometrial Cancer Cohort Study<br>PMID 36477189 | Bryan Anh Nguyen: No saffron | Alexander Tai Nguyen: No saffron | – |

Table S2 continued

| Paper | Reviewer 1 | Reviewer 2 | Additional recorded reviews |
|---|---|---|---|
| 512. Ashraf Ganjooei 2022. Association between Thyroid Hormones and Non-alcoholic Fatty Liver Disease and Non-alcoholic Steatohepatitis in Obese Individuals Undergoing Bariatric Surgery PMID 37547501 | Bryan Anh Nguyen: No saffron | Alexander Tai Nguyen: No saffron | – |
| 513. Anaeigoudari 2023. A review of therapeutic impacts of saffron (Crocus sativus L.) and its constituents PMID 37537722 | Bryan Anh Nguyen: No saffron | Alexander Tai Nguyen: No saffron | – |
| 514. Garza-Campos 2023. Implications of receptor for advanced glycation end products for progression from obesity to diabetes and from diabetes to cancer PMID 37547586 | Bryan Anh Nguyen: No saffron | Alexander Tai Nguyen: No saffron | – |
| 515. Mohammadi 2023. Effect of crocin and losartan on biochemical parameters and genes expression of FRMD3 and BMP7 in diabetic rats PMID 36945919 | Bryan Anh Nguyen: Not clinical | Alexander Tai Nguyen: Not clinical | – |
| 516. Umar 2023. Phenolics Extracted from Jasminum sambac Mitigates Diabetic Cardiomyopathy by Modulating Oxidative Stress, Apoptotic Mediators and the Nfr-2/HO-1 Pathway in Alloxan-Induced Diabetic Rats PMID 37513325 | Bryan Anh Nguyen: No saffron | Alexander Tai Nguyen: No saffron | – |
| 517. Ansari 2023. Protective Effects of Medicinal Plant-Based Foods against Diabetes: A Review on Pharmacology, Phytochemistry, and Molecular Mechanisms PMID 37513684 | Bryan Anh Nguyen: No saffron | Alexander Tai Nguyen: No saffron | – |
| 518. Fanelli Kuczmarski 2023. Diet Quality Trajectories over Adulthood in a Biracial Urban Sample from the Healthy Aging in Neighborhoods of Diversity across the Life Span Study PMID 37513517 | Bryan Anh Nguyen: No saffron | Alexander Tai Nguyen: No saffron | – |
| 519. Uzelac 2023. Invasive Alien Species as a Potential Source of Phytopharmaceuticals: Phenolic Composition and Antimicrobial and Cytotoxic Activity of Robinia pseudoacacia L. Leaf and Flower Extracts PMID 37514330 | Bryan Anh Nguyen: No saffron | Alexander Tai Nguyen: No saffron | – |
| 520. Maneesai 2023. Cardiovascular complications are resolved by tuna protein hydrolysate supplementation in rats fed with a high-fat diet PMID 37507421 | Bryan Anh Nguyen: No saffron | Alexander Tai Nguyen: No saffron | – |
| 521. Lis 2023. Plant Food Dyes with Antioxidant Properties and Allergies-Friend or Enemy? PMID 37507897 | Bryan Anh Nguyen: No saffron | Alexander Tai Nguyen: No saffron | – |

Table S2 continued

| Paper | Reviewer 1 | Reviewer 2 | Additional recorded reviews |
|---|---|---|---|
| 522. Białkowska 2023. Plasma Carotenoids and Polyphenols and Their Association with MetS: The Need for Nutritional Interventions<br>PMID 37507876 | Bryan Anh Nguyen: No saffron | Alexander Tai Nguyen: No saffron | – |
| 523. Lejman-Larysz 2023. Influence of hsCRP Parameter on the Occurrence of Metabolic Syndrome in Patients with Polycystic Ovary Syndrome<br>PMID 37509592 | Bryan Anh Nguyen: No saffron | Alexander Tai Nguyen: No saffron | – |
| 524. Torraville 2023. Life Experience Matters: Enrichment and Stress Can Influence the Likelihood of Developing Alzheimer's Disease via Gut Microbiome<br>PMID 37509523 | Bryan Anh Nguyen: No saffron | Alexander Tai Nguyen: No saffron | – |
| 525. Liu 2023. Bioactives and Inflammation<br>PMID 37504284 | Bryan Anh Nguyen: Not clinical | Alexander Tai Nguyen: No saffron | – |
| 526. Chu 2023. Compound K Production: Achievements and Perspectives<br>PMID 37511939 | Bryan Anh Nguyen: No saffron | Alexander Tai Nguyen: No saffron | – |
| 527. Mohamed 2023. Gallic acid and metformin co-administration reduce oxidative stress, apoptosis and inflammation via Fas/caspase-3 and NF-$\kappa$B signaling pathways in thioacetamide-induced acute hepatic encephalopathy in rats<br>PMID 37491245 | Bryan Anh Nguyen: No saffron | Alexander Tai Nguyen: No saffron | – |
| 528. He 2023. Polysaccharides from natural resource: ameliorate type 2 diabetes mellitus via regulation of oxidative stress network<br>PMID 37497112 | Bryan Anh Nguyen: No saffron | Alexander Tai Nguyen: No saffron | – |
| 529. Zhu 2023. Regulation of Long Noncoding RNA NEAT1/miR-320a/HIF-1$\alpha$ Competitive Endogenous RNA Regulatory Network in Diabetic Retinopathy<br>PMID 37432846 | Bryan Anh Nguyen: No saffron | Alexander Tai Nguyen: No saffron | – |
| 530. Yu 2023. The Effect of Cinnamon on Glycolipid Metabolism: A Dose-Response Meta-Analysis of Randomized Controlled Trials<br>PMID 37447309 | Bryan Anh Nguyen: No saffron | Alexander Tai Nguyen: No saffron | – |
| 531. AlAfaleq 2023. Proinflammatory cytokine profiles in prediabetic Saudi patients<br>PMID 37457235 | Bryan Anh Nguyen: No saffron | Alexander Tai Nguyen: No saffron | – |
| 532. Ozyigit 2023. Production of secondary metabolites using tissue culture-based biotechnological applications<br>PMID 37457343 | Bryan Anh Nguyen: No saffron | Alexander Tai Nguyen: No saffron | – |

Table S2 continued

| Paper | Reviewer 1 | Reviewer 2 | Additional recorded reviews |
|---|---|---|---|
| 533. Zahra 2023. Efficacy of Green Synthesized Nanoparticles in Photodynamic Therapy: A Therapeutic Approach PMID 37446109 | Bryan Anh Nguyen: No saffron | Alexander Tai Nguyen: No saffron | – |
| 534. Fusi 2023. Photon- and Singlet-Oxygen-Induced Cis-Trans Isomerization of the Water-Soluble Carotenoid Crocin PMID 37445961 | Bryan Anh Nguyen: No saffron | Alexander Tai Nguyen: No saffron | – |
| 535. Tsilingiris 2023. Laboratory Findings and Biomarkers in Long COVID: What Do We Know So Far? Insights into Epidemiology, Pathogenesis, Therapeutic Perspectives and Challenges PMID 37445634 | Bryan Anh Nguyen: No saffron | Alexander Tai Nguyen: No saffron | – |
| 536. Shahinfar 2023. The combined association of dietary inflammatory index and resting metabolic rate on cardiorespiratory fitness in adults PMID 37443056 | Bryan Anh Nguyen: No saffron | Alexander Tai Nguyen: No saffron | – |
| 537. Millar 2023. Association of Proinflammatory Diet With Frailty Onset Among Adults With and Without Depressive Symptoms: Results From the Framingham Offspring Study PMID 35830506 | Bryan Anh Nguyen: No saffron | Alexander Tai Nguyen: No saffron | – |
| 538. Valizadeh 2023. Association between the rs2241883 polymorphism of the fatty acid-binding protein-1 (FABP1) gene and obesity in a population of MASHAD study cohort PMID 37026586 | Bryan Anh Nguyen: No saffron | Alexander Tai Nguyen: No saffron | – |
| 539. Khoshandam 2023. Propolis and its constituents against cardiovascular risk factors including obesity, hypertension, atherosclerosis, diabetes, and dyslipidemia: A comprehensive review PMID 37427329 | Bryan Anh Nguyen: No saffron | Alexander Tai Nguyen: No saffron | – |
| 540. Urizar 2023. Beta cell dysfunction induced by bone morphogenetic protein (BMP)-2 is associated with histone modifications and decreased NeuroD1 chromatin binding PMID 37407581 | Bryan Anh Nguyen: No saffron | Alexander Tai Nguyen: No saffron | – |
| 541. In Het Panhuis 2023. Time-restricted feeding attenuates hypercholesterolaemia and atherosclerosis development during circadian disturbance in APOE∗3-Leiden.CETP mice PMID 37356205 | Bryan Anh Nguyen: No saffron | Alexander Tai Nguyen: No saffron | – |
| 542. Shaik Mohamed Sayed 2023. Natural products as novel anti-obesity agents: insights into mechanisms of action and potential for therapeutic management PMID 37408757 | Bryan Anh Nguyen: No saffron | Alexander Tai Nguyen: No saffron | – |

Table S2 continued

| Paper | Reviewer 1 | Reviewer 2 | Additional recorded reviews |
|---|---|---|---|
| 543. Oskouei 2023. The effects of Dendrobium species on the metabolic syndrome: A review<br>PMID 37396948 | Bryan Anh Nguyen: No saffron | Alexander Tai Nguyen: No saffron | – |
| 544. Xu 2023. Yinchenhao Tang alleviates high fat diet induced NAFLD by increasing NR1H4 and APOA1 expression<br>PMID 37396157 | Bryan Anh Nguyen: No saffron | Alexander Tai Nguyen: Not clinical | – |
| 545. Basta 2023. Resveratrol ameliorates the behavioural and molecular changes in rats exposed to uninephrectomy: role of hippocampal SIRT1, BDNF and AChE<br>PMID 36469209 | Bryan Anh Nguyen: No saffron | Alexander Tai Nguyen: No saffron | – |
| 546. Charneca 2023. Beyond Seasoning-The Role of Herbs and Spices in Rheumatic Diseases<br>PMID 37375716 | Bryan Anh Nguyen: No saffron | Alexander Tai Nguyen: No saffron | – |
| 547. Farhan 2023. Insights on the Role of Polyphenols in Combating Cancer Drug Resistance<br>PMID 37371804 | Bryan Anh Nguyen: No saffron | Alexander Tai Nguyen: No saffron | – |
| 548. Pourreza 2022. Therapeutic Effects and Mechanisms of Action of Garlic (Allium sativum) on Nonalcoholic Fatty Liver Disease: A Comprehensive Systematic Literature Review<br>PMID 37377647 | Bryan Anh Nguyen: No saffron | Alexander Tai Nguyen: No saffron | – |
| 549. Cheng-Yuan 2023. Research progress on the prevention and treatment of hyperuricemia by medicinal and edible plants and its bioactive components<br>PMID 37377486 | Bryan Anh Nguyen: No saffron | Alexander Tai Nguyen: No saffron | – |
| 550. Wei 2023. Ferroptosis: a new strategy for Chinese herbal medicine treatment of diabetic nephropathy<br>PMID 37361521 | Bryan Anh Nguyen: Not clinical | Alexander Tai Nguyen: No saffron | – |
| 551. de Mello 2023. Dietary inflammatory index and its relation to the pathophysiological aspects of obesity: a narrative review<br>PMID 37364142 | Bryan Anh Nguyen: No saffron | Alexander Tai Nguyen: No saffron | – |
| 552. Mohammadi 2023. The effect of crocin and losartan on TGF-$\beta$ gene expression and histopathology of kidney tissue in a rat model of diabetic nephropathy<br>PMID 37333473 | Bryan Anh Nguyen: Not clinical | Alexander Tai Nguyen: No saffron | – |
| 553. Mao 2023. Protective effects of dioscin against Parkinson's disease via regulating bile acid metabolism through remodeling gut microbiome/GLP-1 signaling<br>PMID 38024855 | Bryan Anh Nguyen: No saffron | Alexander Tai Nguyen: No saffron | – |

Table S2 continued

| Paper | Reviewer 1 | Reviewer 2 | Additional recorded reviews |
|---|---|---|---|
| 554. Jandari 2023. The effect of low dietary inflammatory index score formula on inflammatory, metabolic, and clinical outcomes in critically ill traumatic brain injury patients: A single-blind randomized controlled pilot study<br>PMID 37324871 | Bryan Anh Nguyen: No saffron | Alexander Tai Nguyen: No saffron | – |
| 555. Rondanelli 2023. Diet and ideal food pyramid to prevent or support the treatment of diabetic retinopathy, age-related macular degeneration, and cataracts<br>PMID 37324128 | Bryan Anh Nguyen: No saffron | Alexander Tai Nguyen: No saffron | – |
| 556. Ghazizadeh 2023. Use of data mining approaches to explore the association between type 2 diabetes mellitus with SARS-CoV-2<br>PMID 37308948 | Bryan Anh Nguyen: No saffron | Alexander Tai Nguyen: No saffron | – |
| 557. González 2023. Hyperglycemia and Oxidative Stress: An Integral, Updated and Critical Overview of Their Metabolic Interconnections<br>PMID 37298303 | Bryan Anh Nguyen: No saffron | Alexander Tai Nguyen: No saffron | – |
| 558. Reyes-Castellanos 2023. Combining the antianginal drug perhexiline with chemotherapy induces complete pancreatic cancer regression in vivo<br>PMID 37305702 | Bryan Anh Nguyen: No saffron | Alexander Tai Nguyen: No saffron | – |
| 559. Yaribeygi 2023. Sodium Glucose Cotransporter-2 Inhibitor Empagliflozin Increases Antioxidative Capacity and Improves Renal Function in Diabetic Rats<br>PMID 37298010 | Bryan Anh Nguyen: No saffron | Alexander Tai Nguyen: No saffron | – |
| 560. Badaeva 2023. Perspectives on Neuronutrition in Prevention and Treatment of Neurological Disorders<br>PMID 37299468 | Bryan Anh Nguyen: No saffron | Alexander Tai Nguyen: Not clinical | – |
| 561. Khakisahneh 2023. Yijung-tang improves thermogenesis and reduces inflammation associated with gut microbiota in hypothyroid rats<br>PMID 37270649 | Bryan Anh Nguyen: No saffron | Alexander Tai Nguyen: No saffron | – |
| 562. Jafari 2023. Pharmacological Effects of Genistein on Cardiovascular Diseases<br>PMID 37275572 | Bryan Anh Nguyen: No saffron | Alexander Tai Nguyen: No saffron | – |
| 563. Wijdeveld 2023. Identifying Gut Microbiota associated with Gastrointestinal Symptoms upon Roux-en-Y Gastric Bypass<br>PMID 37093508 | Bryan Anh Nguyen: No saffron | Alexander Tai Nguyen: No saffron | – |

Table S2 continued

| Paper | Reviewer 1 | Reviewer 2 | Additional recorded reviews |
|---|---|---|---|
| 564. Hassan 2023. Genistein ameliorated experimentally induced gastric ulcer in rats via inhibiting gastric tissues fibrosis by modulating Wnt/$\beta$-catenin/TGF-$\beta$/PKB pathway PMID 37260037 | Bryan Anh Nguyen: No saffron | Alexander Tai Nguyen: No saffron | – |
| 565. Eroglu 2023. Carotenoids and Their Health Benefits as Derived via Their Interactions with Gut Microbiota PMID 36775788 | Bryan Anh Nguyen: No saffron | Alexander Tai Nguyen: No saffron | – |
| 566. Hao 2023. Pharmacological Mechanisms and Clinical Applications of Curcumin: Update PMID 37191432 | Bryan Anh Nguyen: No saffron | Alexander Tai Nguyen: No saffron | – |
| 567. Lin 2023. Extracellular status of thrombospondin-2 in type 2 diabetes mellitus and utility as a biomarker in the determination of early diabetic kidney disease PMID 37259071 | Bryan Anh Nguyen: No saffron | Alexander Tai Nguyen: No saffron | – |
| 568. Saidi 2023. Phytochemical Profile, Antioxidant, Antimicrobial, and Antidiabetic Activities of Ajuga iva (L.) PMID 37240812 | Bryan Anh Nguyen: No saffron | Alexander Tai Nguyen: No saffron | – |
| 569. Shao 2023. Anti-Diabetic Activity of a Novel Exopolysaccharide Produced by the Mangrove Endophytic Fungus Penicillium janthinellum N29 PMID 37233464 | Bryan Anh Nguyen: No saffron | Alexander Tai Nguyen: No saffron | – |
| 570. Biernacka 2023. The Potential of Ginkgo biloba as a Source of Biologically Active Compounds-A Review of the Recent Literature and Patents PMID 37241734 | Bryan Anh Nguyen: No saffron | Alexander Tai Nguyen: No saffron | – |
| 571. Gambino 2023. "Golden" Tomato Consumption Ameliorates Metabolic Syndrome: A Focus on the Redox Balance in the High-Fat-Diet-Fed Rat PMID 37237987 | Bryan Anh Nguyen: No saffron | Alexander Tai Nguyen: No saffron | – |
| 572. Maiese 2023. Cellular Metabolism: A Fundamental Component of Degeneration in the Nervous System PMID 37238686 | Bryan Anh Nguyen: No saffron | Alexander Tai Nguyen: No saffron | – |
| 573. Jiang 2023. The Nutrients and Volatile Compounds in Stropharia rugoso-annulata by Three Drying Treatments PMID 37238895 | Bryan Anh Nguyen: No saffron | Alexander Tai Nguyen: No saffron | – |
| 574. De la Fuente-Muñoz 2023. Supplementation with a New Standardized Extract of Green and Black Tea Exerts Antiadipogenic Effects and Prevents Insulin Resistance in Mice with Metabolic Syndrome PMID 37239868 | Bryan Anh Nguyen: No saffron | Alexander Tai Nguyen: No saffron | – |

Table S2 continued

| Paper | Reviewer 1 | Reviewer 2 | Additional recorded reviews |
|---|---|---|---|
| 575. Inia 2023. Semaglutide Has Beneficial Effects on Non-Alcoholic Steatohepatitis in Ldlr-/-.Leiden Mice<br>PMID 37239841 | Bryan Anh Nguyen: No saffron | Alexander Tai Nguyen: No saffron | – |
| 576. Veisi 2023. Effect of Zingiber officinale on Lipid Profile and Some Inflammatory Markers in Diabetic Hemodialysis Patients: A Randomized Double-Blind Placebo-Controlled Clinical Trial<br>PMID 37228303 | Bryan Anh Nguyen: No saffron | Alexander Tai Nguyen: No saffron | – |
| 577. Hall 2023. Effect of Dietary Intervention, with or without Cointerventions, on Inflammatory Markers in Patients with Nonalcoholic Fatty Liver Disease: A Systematic Review and Meta-Analysis<br>PMID 36796436 | Bryan Anh Nguyen: No saffron | Alexander Tai Nguyen: Not clinical | – |
| 578. Li 2020. Association of sugar-sweetened beverage intake with risk of metabolic syndrome among children and adolescents in urban China<br>PMID 31915093 | Bryan Anh Nguyen: No saffron | Alexander Tai Nguyen: No saffron | – |
| 579. Tsilimigras 2023. Rosuvastatin Attenuates Progression of Atherosclerosis and Reduces Serum IL6 and CCL2 Levels in Apolipoprotein-E-deficient Mice<br>PMID 37103114 | Bryan Anh Nguyen: No saffron | Alexander Tai Nguyen: No saffron | – |
| 580. Mirenayat 2023. Effects of sumac supplementation on metabolic markers in adults with metabolic syndrome: a triple-blinded randomized placebo-controlled cross-over clinical trial<br>PMID 37189189 | Bryan Anh Nguyen: No saffron | Alexander Tai Nguyen: No saffron | – |
| 581. Andrés-Blasco 2023. Oxidative Stress, Inflammatory, Angiogenic, and Apoptotic molecules in Proliferative Diabetic Retinopathy and Diabetic Macular Edema Patients<br>PMID 37175931 | Bryan Anh Nguyen: No saffron | Alexander Tai Nguyen: No saffron | – |
| 582. Inia 2023. Atorvastatin Attenuates Diet-Induced Non-Alcoholic Steatohepatitis in APOE*3-Leiden Mice by Reducing Hepatic Inflammation<br>PMID 37175538 | Bryan Anh Nguyen: No saffron | Alexander Tai Nguyen: No saffron | – |
| 583. Tommerdahl 2022. Youth versus adult-onset type 2 diabetic kidney disease: Insights into currently known structural differences and the potential underlying mechanisms<br>PMID 36326718 | Bryan Anh Nguyen: No saffron | Alexander Tai Nguyen: No saffron | – |
| 584. Abedi 2023. HDL-cholesterol concentration and its association with coronary artery calcification: a systematic review and meta-analysis<br>PMID 37158895 | Bryan Anh Nguyen: No saffron | Alexander Tai Nguyen: No saffron | – |

Table S2 continued

| Paper | Reviewer 1 | Reviewer 2 | Additional recorded reviews |
|---|---|---|---|
| 585. Zancanella 2023. Proof-of-concept study for liver-directed miQURE technology in a dyslipidemic mouse model<br>PMID 37168797 | Bryan Anh Nguyen: No saffron | Alexander Tai Nguyen: No saffron | – |
| 586. Xu 2022. Diabetes Technology Meeting 2021<br>PMID 35499170 | Prisha Batra: Not clinical | Alexander Tai Nguyen: No saffron | – |
| 587. Heindel 2022. Obesity II: Establishing causal links between chemical exposures and obesity<br>PMID 35395240 | Prisha Batra: Not clinical | Alexander Tai Nguyen: No saffron | – |
| 588. Ai 2023. Hepatotoxic Components Effect of Chebulae Fructus and Associated Molecular Mechanism by Integrated Transcriptome and Molecular Docking<br>PMID 37110661 | Prisha Batra: Not clinical | Alexander Tai Nguyen: No saffron | – |
| 589. Drioiche 2023. Identification of Compounds of Crocus sativus by GC-MS and HPLC/UV-ESI-MS and Evaluation of Their Antioxidant, Antimicrobial, Anticoagulant, and Antidiabetic Properties<br>PMID 37111302 | Prisha Batra: Not clinical | Alexander Tai Nguyen: Not isolated | – |
| 590. Seo 2023. New Sesquiterpene Glycosides from the Flowers of Aster koraiensis and Their Inhibition Activities on EGF- and TPA-Induced Cell Transformation<br>PMID 37111949 | Prisha Batra: Not clinical | Alexander Tai Nguyen: No saffron | – |
| 591. Qiao 2023. Herbal/Natural Compounds Resist Hallmarks of Brain Aging: From Molecular Mechanisms to Therapeutic Strategies<br>PMID 37107295 | Prisha Batra: Not clinical | Alexander Tai Nguyen: No saffron | – |
| 592. Gil 2023. Antioxidant Activity and Inhibition of Digestive Enzymes of New Strawberry Tree Fruit/Apple Smoothies<br>PMID 37107180 | Prisha Batra: Not clinical | Alexander Tai Nguyen: Not clinical | – |
| 593. Kondo 2023. Rheumatoid Arthritis and Reactive Oxygen Species: A Review<br>PMID 37185721 | Prisha Batra: Not clinical | Alexander Tai Nguyen: No saffron | – |
| 594. Zhou 2023. Effective Therapeutic Verification of Crocin I, Geniposide, and Gardenia (Gardenia jasminoides Ellis) on Type 2 Diabetes Mellitus In Vivo and In Vitro<br>PMID 37107463 | Prisha Batra: Not clinical | Alexander Tai Nguyen: Not clinical | – |
| 595. Alshammary 2023. Saudi Community-Based Screening Study on Genetic Variants in $\beta$-Cell Dysfunction and Its Role in Women with Gestational Diabetes Mellitus<br>PMID 37107681 | Prisha Batra: Not clinical | Alexander Tai Nguyen: No saffron | – |

Table S2 continued

| Paper | Reviewer 1 | Reviewer 2 | Additional recorded reviews |
|---|---|---|---|
| 596. Ojo 2023. Apoptosis, inflammation, and oxidative stress in infertility: A mini review<br>PMID 37125147 | Prisha Batra: Not clinical | Alexander Tai Nguyen: No saffron | – |
| 597. Giri 2023. Treatment on Nature's lap: Use of herbal products in the management of hyperglycemia<br>PMID 37122430 | Bryan Anh Nguyen: No saffron | Alexander Tai Nguyen: No saffron | – |
| 598. Wijeweera 2023. Therapeutic Implications of Some Natural Products for Neuroimmune Diseases: A Narrative of Clinical Studies Review<br>PMID 37089709 | Bryan Anh Nguyen: No saffron | Alexander Tai Nguyen: No saffron | – |
| 599. EFSA Panel on Food Contact Materials, Enzymes and Processing Aids (CEP) 2023. Re-evaluation of the risks to public health related to the presence of bisphenol A (BPA) in foodstuffs<br>PMID 37089179 | Bryan Anh Nguyen: No saffron | Alexander Tai Nguyen: No saffron | – |
| 600. Ding 2023. Meta-analysis of the association between dietary inflammatory index and cognitive health<br>PMID 37081917 | Bryan Anh Nguyen: No saffron | Alexander Tai Nguyen: Not clinical | – |
| 601. Aghaei 2023. Neuroprotective Effect of Wharton's Jelly-Derived Mesenchymal Stem Cell-Conditioned Medium (WJMSC-CM) on Diabetes-Associated Cognitive Impairment by Improving Oxidative Stress, Neuroinflammation, and Apoptosis<br>PMID 37081849 | Bryan Anh Nguyen: No saffron | Alexander Tai Nguyen: No saffron | – |
| 602. Lin 2023. Anthocyanins: Modified New Technologies and Challenges<br>PMID 37048188 | Bryan Anh Nguyen: No saffron | Alexander Tai Nguyen: No saffron | – |
| 603. Yang 2023. The relationship between HMGB1 and autophagy in the pathogenesis of diabetes and its complications<br>PMID 37065747 | Bryan Anh Nguyen: No saffron | Alexander Tai Nguyen: No saffron | – |
| 604. Hikisz 2023. Diet as a Source of Acrolein: Molecular Basis of Aldehyde Biological Activity in Diabetes and Digestive System Diseases<br>PMID 37047550 | Bryan Anh Nguyen: No saffron | Alexander Tai Nguyen: No saffron | – |
| 605. Mokhtari 2023. Comparative Effect of the Active Substance of Thyme with N-Acetyl Cysteine on Hematological Parameters and Histopathological Changes of Bone Marrow and Liver in Rat Models of Acetaminophen Toxicity<br>PMID 37056637 | Bryan Anh Nguyen: No saffron | Alexander Tai Nguyen: No saffron | – |

Table S2 continued

| Paper | Reviewer 1 | Reviewer 2 | Additional recorded reviews |
|---|---|---|---|
| 606. Li 2023. Nigella sativa and health outcomes: An overview of systematic reviews and meta-analyses PMID 37057067 | Bryan Anh Nguyen: No saffron | Alexander Tai Nguyen: Not clinical | – |
| 607. Kamari 2023. Combination of the effect of ginger and anti-inflammatory diet on children with obesity with nonalcoholic fatty liver disease: A randomized clinical trial PMID 37051346 | Bryan Anh Nguyen: No saffron | Alexander Tai Nguyen: No saffron | – |
| 608. Jamialahmadi 2023. Association of C-reactive protein with histological, elastographic, and sonographic indices of non-alcoholic fatty liver disease in individuals with severe obesity PMID 37029427 | Bryan Anh Nguyen: No saffron | Alexander Tai Nguyen: No saffron | – |
| 609. Wang 2023. The lipid-lowering effects of fenugreek gum, hawthorn pectin, and burdock inulin PMID 37032784 | Bryan Anh Nguyen: No saffron | Alexander Tai Nguyen: No saffron | – |
| 610. Mondal 2023. An overview of nutritional profiling in foods: Bioanalytical techniques and useful protocols PMID 37025612 | Bryan Anh Nguyen: No saffron | Alexander Tai Nguyen: No saffron | – |
| 611. Park 2023. Umbelliferone Ameliorates Hepatic Steatosis and Lipid-Induced ER Stress in High-Fat Diet-Induced Obese Mice PMID 36996895 | Bryan Anh Nguyen: No saffron | Alexander Tai Nguyen: No saffron | – |
| 612. Rahbarghazi 2023. Co-administration of exercise training and melatonin on the function of diabetic heart tissue: a systematic review and meta-analysis of rodent models PMID 37005639 | Bryan Anh Nguyen: No saffron | Alexander Tai Nguyen: No saffron | – |
| 613. Yang 2023. Sirtuin-3: A potential target for treating several types of brain injury PMID 37009480 | Bryan Anh Nguyen: No saffron | Alexander Tai Nguyen: No saffron | – |
| 614. Patel 2023. Nanocarriers for the Delivery of Neuroprotective Agents in the Treatment of Ocular Neurodegenerative Diseases PMID 36986699 | Bryan Anh Nguyen: No saffron | Alexander Tai Nguyen: No saffron | – |
| 615. Testa 2023. 24-Hydroxycholesterol Induces Tau Proteasome-Dependent Degradation via the SIRT1/PGC1$\alpha$/Nrf2 Pathway: A Potential Mechanism to Counteract Alzheimer's Disease PMID 36978879 | Bryan Anh Nguyen: No saffron | Alexander Tai Nguyen: No saffron | – |
| 616. Longoni 2023. Polyunsaturated Lipids in the Light-Exposed and Prooxidant Retinal Environment PMID 36978865 | Bryan Anh Nguyen: No saffron | Alexander Tai Nguyen: No saffron | – |

Table S2 continued

| Paper | Reviewer 1 | Reviewer 2 | Additional recorded reviews |
|---|---|---|---|
| 618. Alatorre-Cruz 2023. Traditional Mexican Food: Phenolic Content and Public Health Relationship<br>PMID 36981159 | Prisha Batra: Not clinical | Alexander Tai Nguyen: No saffron | – |
| 619. Jeong 2023. Inverse association of daily fermented soybean paste ("Jang") intake with metabolic syndrome risk, especially body fat and hypertension, in men of a large hospital-based cohort<br>PMID 36992908 | Prisha Batra: Not clinical | Alexander Tai Nguyen: No saffron | – |
| 620. Nowicka 2023. Quantitative and qualitative determination of carotenoids and polyphenolics compounds in selected cultivars of Prunus persica L. and their ability to in vitro inhibit lipoxygenase, cholinoesterase, $\alpha$-amylase, $\alpha$-glucosidase and pancreatic lipase<br>PMID 36974173 | Prisha Batra: Not clinical | Alexander Tai Nguyen: No saffron | – |
| 621. Towanou 2023. Phytochemical Screening, Antioxidant Activity, and Acute Toxicity Evaluation of Senna italica Extract Used in Traditional Medicine<br>PMID 36968147 | Prisha Batra: Not clinical | Alexander Tai Nguyen: No saffron | – |
| 622. Soussi 2023. A Multidimensional Review of<br>PMID #N/A | Prisha Batra: Not clinical | Alexander Tai Nguyen: No saffron | – |
| 623. Gouirand 2022. Ketogenic HMG-CoA lyase and its product $\beta$-hydroxybutyrate promote pancreatic cancer progression<br>PMID 35307861 | Prisha Batra: Not clinical | Alexander Tai Nguyen: No saffron | – |
| 624. Zadhoush 2023. The Therapeutic Impact of Garlic (Allium Sativum) on Oxidative Stress Markers among Polycystic Ovary Syndrome Patients: A Randomized, Double-Blinded Clinical Trial<br>PMID 36942042 | Prisha Batra: No saffron | Alexander Tai Nguyen: No saffron | – |
| 625. Fourati 2023. Fecal microbiota transplantation in a rodent model of short bowel syndrome: A therapeutic approach?<br>PMID 36936775 | Prisha Batra: Not clinical | Alexander Tai Nguyen: No saffron | – |
| 626. Rezaeinezhad 2022. Short-term circuit resistance training improves insulin resistance probably via increasing circulating Adropin<br>PMID 35673513 | Prisha Batra: No saffron | Alexander Tai Nguyen: No saffron | – |
| 627. Gart 2023. Translational characterization of the temporal dynamics of metabolic dysfunctions in liver, adipose tissue and the gut during diet-induced NASH development in Ldlr-/-.Leiden mice<br>PMID 36915476 | Prisha Batra: No saffron | Alexander Tai Nguyen: No saffron | – |

Table S2 continued

| Paper | Reviewer 1 | Reviewer 2 | Additional recorded reviews |
|---|---|---|---|
| 628. Fereidouni 2022. Diet quality and dietary acid load in relation to cardiovascular disease mortality: Results from Fasa PERSIAN cohort study<br>PMID 36911829 | Prisha Batra: Not clinical | Alexander Tai Nguyen: No saffron | – |
| 629. Lotfi 2022. Plant-based diets could ameliorate the risk factors of cardiovascular diseases in adults with chronic diseases<br>PMID 36911818 | Prisha Batra: Not clinical | Alexander Tai Nguyen: No saffron | – |
| 630. Brockmueller 2023. Resveratrol Modulates Chemosensitisation to 5-FU via $\beta$1-Integrin/HIF-1$\alpha$ Axis in CRC Tumor Microenvironment<br>PMID 36902421 | Prisha Batra: Not clinical | Alexander Tai Nguyen: No saffron | – |
| 631. Panknin 2023. Curcumin Supplementation and Human Disease: A Scoping Review of Clinical Trials<br>PMID 36901908 | Prisha Batra: Not clinical | Alexander Tai Nguyen: No saffron | – |
| 632. Jin 2023. Costunolide alleviates hyperglycaemia-induced diabetic cardiomyopathy via inhibiting inflammatory responses and oxidative stress<br>PMID 36810875 | Prisha Batra: Not clinical | Alexander Tai Nguyen: No saffron | – |
| 633. Kosmalski 2023. What's New in the Treatment of Non-Alcoholic Fatty Liver Disease (NAFLD)<br>PMID 36902639 | Prisha Batra: Not clinical | Alexander Tai Nguyen: Not clinical | – |
| 634. Musazadeh 2023. Effects of synbiotics supplementation on anthropometric and lipid profile parameters: Finding from an umbrella meta-analysis<br>PMID 36908920 | Prisha Batra: Not clinical | Alexander Tai Nguyen: No saffron | – |
| 635. Ferreira 2021. The association between an energy-adjusted dietary inflammatory index and inflammation in rural and urban Black South Africans<br>PMID 34955112 | Prisha Batra: Not clinical | Alexander Tai Nguyen: No saffron | – |
| 636. Soetikno 2023. Alpha-Mangosteen lessens high-fat/high-glucose diet and low-dose streptozotocin induced-hepatic manifestations in the insulin resistance rat model<br>PMID 36655319 | Prisha Batra: Not clinical | Alexander Tai Nguyen: No saffron | – |
| 637. Delgadillo-Puga 2023. Cardamom (Elettaria cardamomum (L.) Maton) Seeds Intake Increases Energy Expenditure and Reduces Fat Mass in Mice by Modulating Neural Circuits That Regulate Adipose Tissue Lipolysis and Mitochondrial Oxidative Metabolism in Liver and Skeletal Muscle<br>PMID 36835337 | Prisha Batra: Not clinical | Alexander Tai Nguyen: No saffron | – |

Table S2 continued

| Paper | Reviewer 1 | Reviewer 2 | Additional recorded reviews |
|---|---|---|---|
| 638. Lawson 2023. Improvement of Therapeutic Value of Quercetin with Chitosan Nanoparticle Delivery Systems and Potential Applications<br>PMID 36834702 | Prisha Batra: Not clinical | Alexander Tai Nguyen: No saffron | – |
| 639. Sehaki 2023. Metabolomics-Based Profiling via a Chemometric Approach to Investigate the Antidiabetic Property of Different Parts and Origins of Pistacia lentiscus L<br>PMID 36837894 | Prisha Batra: Not clinical | Alexander Tai Nguyen: No saffron | – |
| 640. Daily 2023. Association of Plant-Based and High-Protein Diets with a Lower Obesity Risk Defined by Fat Mass in Middle-Aged and Elderly Persons with a High Genetic Risk of Obesity<br>PMID 36839421 | Prisha Batra: Not clinical | Alexander Tai Nguyen: No saffron | – |
| 641. Park 2023. Protective Effects of Coumestrol on Metabolic Dysfunction and Its Estrogen Receptor-Mediated Action in Ovariectomized Mice<br>PMID 36839308 | Prisha Batra: Not clinical | Alexander Tai Nguyen: No saffron | – |
| 642. Milić 2023. Combined Approach: FFQ, DII, Anthropometric, Biochemical and DNA Damage Parameters in Obese with BMI $\geq$ 35 kg m(-2)<br>PMID 36839257 | Prisha Batra: Not clinical | Alexander Tai Nguyen: No saffron | – |
| 643. Gasmi 2023. Pharmacological Treatments and Natural Biocompounds in Weight Management<br>PMID 37139804 | Prisha Batra: Not clinical | Alexander Tai Nguyen: Not clinical | – |
| 644. Szypowska 2023. Comparison of Intake of Food Groups Based on Dietary Inflammatory Index (DII) and Cardiovascular Risk Factors in the Middle-Age Population of Lower Silesia: Results of the PURE Poland Study<br>PMID 36829844 | Prisha Batra: Not clinical | Alexander Tai Nguyen: No saffron | – |
| 645. He 2023. The Regulatory Effect of Phytochemicals on Chronic Diseases by Targeting Nrf2-ARE Signaling Pathway<br>PMID 36829795 | Prisha Batra: Not clinical | Alexander Tai Nguyen: No saffron | – |
| 646. Ashtary-Larky 2023. Are Women with Normal-Weight Obesity at Higher Risk for Cardiometabolic Disorders?<br>PMID 36830878 | Prisha Batra: Not clinical | Alexander Tai Nguyen: No saffron | – |
| 647. Satyam 2023. Metformin and Dapagliflozin Attenuate Doxorubicin-Induced Acute Cardiotoxicity in Wistar Rats: An Electrocardiographic, Biochemical, and Histopathological Approach<br>PMID 36790727 | Prisha Batra: Not clinical | Alexander Tai Nguyen: No saffron | – |

Table S2 continued

| Paper | Reviewer 1 | Reviewer 2 | Additional recorded reviews |
| --- | --- | --- | --- |
| 648. Wang 2023. Treatment of Diabetes Nephropathy in Mice by Germinating Seeds of Euryale ferox through Improving Oxidative Stress<br>PMID 36832842 | Prisha Batra: Not clinical | Alexander Tai Nguyen: No saffron | – |
| 649. Kumar 2022. Mucoadhesive carriers for oral drug delivery<br>PMID 36116580 | Prisha Batra: Not clinical | Alexander Tai Nguyen: No saffron | – |
| 650. Buller 2023. Median eminence myelin continuously turns over in adult mice<br>PMID 36739968 | Prisha Batra: Not clinical | Alexander Tai Nguyen: No saffron | – |
| 651. Khoshnood 2023. Amoxicillin-docosahexaenoic acid encapsulated chitosan-alginate nanoparticles as a delivery system with enhanced biocidal activities against Helicobacter pylori and improved ulcer healing<br>PMID 36846798 | Prisha Batra: Not clinical | Alexander Tai Nguyen: No saffron | – |
| 652. Teo 2023. Associations between maternal dietary scores during early pregnancy with placental outcomes<br>PMID 36845057 | Prisha Batra: Not clinical | Alexander Tai Nguyen: No saffron | – |
| 653. Govindugari 2023. Thwarting Alzheimer's Disease through Healthy Lifestyle Habits: Hope for the Future<br>PMID 36810468 | Prisha Batra: Not clinical | Alexander Tai Nguyen: No saffron | – |
| 654. Takayama 2023. Efficacy and safety of anamorelin in patients with cancer cachexia: Post-hoc subgroup analyses of a placebo-controlled study<br>PMID 36394148 | Michele Schaffer: Not isolated | Prisha Batra: Not clinical | – |
| 655. Zhang 2023. Investigation of the Potential Mechanism of Alpinia officinarum Hance in Improving Type 2 Diabetes Mellitus Based on Network Pharmacology and Molecular Docking<br>PMID 36818229 | Michele Schaffer: Not isolated | Prisha Batra: Not clinical | – |
| 656. Abedi 2023. Effect of saffron supplementation on oxidative stress markers (MDA, TAC, TOS, GPx, SOD, and pro-oxidant/antioxidant balance): An updated systematic review and meta-analysis of randomized placebo-controlled trials<br>PMID 36817799 | Michele Schaffer: Not clinical | Prisha Batra: Not clinical | – |
| 657. Moreira 2023. Cinnamon: an aromatic condiment applicable to chronic kidney disease<br>PMID 36747357 | Michele Schaffer: Not isolated | Prisha Batra: Not clinical | – |

Table S2 continued

| Paper | Reviewer 1 | Reviewer 2 | Additional recorded reviews |
|---|---|---|---|
| 658. Alshabi 2022. Antidiabetic and antioxidant potential of Gardenia latifolia in type-2 diabetic rats fed with high-fat diet plus low-dose streptozotocin PMID 35964948 | Michele Schaffer: Not isolated | Prisha Batra: Not clinical | – |
| 659. Chattopadhyay 2023. Development process of a clinical guideline to manage type 2 diabetes in adults by Ayurvedic practitioners PMID 36793876 | Michele Schaffer: Not clinical | Prisha Batra: Not clinical | – |
| 660. Deng 2023. $\alpha$-Linolenic Acid Inhibits RANKL-Induced Osteoclastogenesis In Vitro and Prevents Inflammation In Vivo PMID 36766210 | Michele Schaffer: Not isolated | Prisha Batra: Not clinical | – |
| 661. Ma 2022. Nicotinamide mononucleotide improves spermatogenic function in streptozotocin-induced diabetic mice via modulating the glycolysis pathway PMID 35929593 | Michele Schaffer: Not isolated | Prisha Batra: Not clinical | – |
| 662. Prakasam 2023. LSD1/PRMT6-targeting gene therapy to attenuate androgen receptor toxic gain-of-function ameliorates spinobulbar muscular atrophy phenotypes in flies and mice PMID 36746939 | Michele Schaffer: No saffron | Prisha Batra: Not clinical | – |
| 663. Koeder 2023. Exploratory analysis of the effect of a controlled lifestyle intervention on inflammatory markers - the Healthy Lifestyle Community Programme (cohort 2) PMID 36747285 | Michele Schaffer: Not isolated | Prisha Batra: Not clinical | – |
| 664. Betanzos-Robledo 2020. Greater cumulative exposure to a pro-inflammatory diet is associated with higher metabolic syndrome score and blood pressure in young Mexican adults PMID 32942060 | Michele Schaffer: Not isolated | Prisha Batra: Not clinical | – |
| 665. El-Saadony 2023. Impacts of turmeric and its principal bioactive curcumin on human health: Pharmaceutical, medicinal, and food applications: A comprehensive review PMID 36712505 | Michele Schaffer: No saffron | Prisha Batra: Not clinical | – |
| 666. Luo 2023. Advancements in lead therapeutic phytochemicals polycystic ovary syndrome: A review PMID 36699064 | Michele Schaffer: Not clinical | Prisha Batra: Not clinical | – |
| 667. Ying 2023. Phenomic Studies on Diseases: Potential and Challenges PMID 36714223 | Michele Schaffer: Not isolated | Prisha Batra: Not clinical | – |
| 668. Heres 2023. Bioactive and Sensory Di- and Tripeptides Generated during Dry-Curing of Pork Meat PMID 36675084 | Michele Schaffer: Not isolated | Prisha Batra: Not clinical | – |

Table S2 continued

| Paper | Reviewer 1 | Reviewer 2 | Additional recorded reviews |
|---|---|---|---|
| 669. Eltimamy 2022. Effects of alcoholic extract of Terminalia Chebula dried fruit on blood biochemical profile in diabetic rats<br>PMID 35673508 | Michele Schaffer: Not isolated | Prisha Batra: Not clinical | – |
| 670. Devranis 2023. Mediterranean Diet, Ketogenic Diet or MIND Diet for Aging Populations with Cognitive Decline: A Systematic Review<br>PMID 36676122 | Michele Schaffer: Not isolated | Prisha Batra: Not clinical | – |
| 671. Ashruf 2022. Natural Compounds: Potential Therapeutics for the Inhibition of Cartilage Matrix Degradation in Osteoarthritis<br>PMID 36676051 | Michele Schaffer: Not clinical | Prisha Batra: Not clinical | – |
| 672. Rahmani 2023. Quercetin, a Plant Flavonol Attenuates Diabetic Complications, Renal Tissue Damage, Renal Oxidative Stress and Inflammation in Streptozotocin-Induced Diabetic Rats<br>PMID 36677055 | Michele Schaffer: Not isolated | Prisha Batra: Not clinical | – |
| 673. Alqudah 2023. Isorhamnetin Reduces Glucose Level, Inflammation, and Oxidative Stress in High-Fat Diet/Streptozotocin Diabetic Mice Model<br>PMID 36677559 | Michele Schaffer: Not clinical | Prisha Batra: Not clinical | – |
| 674. Monteiro 2023. Identifying and Analyzing Topic Clusters in a Nutri-, Food-, and Diet-Proteomic Corpus Using Machine Reading<br>PMID 36678141 | Michele Schaffer: Not clinical | Prisha Batra: Not clinical | – |
| 675. Matraszek-Gawron 2022. Current Knowledge of the Antidepressant Activity of Chemical Compounds from Crocus sativus L<br>PMID 36678554 | Michele Schaffer: Not clinical | Prisha Batra: Not clinical | – |
| 676. Gu 2022. Advances in Anti-Diabetic Cognitive Dysfunction Effect of Erigeron Breviscapus (Vaniot) Hand-Mazz<br>PMID 36678547 | Michele Schaffer: Not isolated | Prisha Batra: Not clinical | – |
| 677. González-Peña 2023. Chemistry, Occurrence, Properties, Applications, and Encapsulation of Carotenoids-A Review<br>PMID 36679026 | Michele Schaffer: Not clinical | Prisha Batra: Not clinical | – |
| 678. Petrarca 2023. Redox Remodeling by Nutraceuticals for Prevention and Treatment of Acute and Chronic Inflammation<br>PMID 36670995 | Michele Schaffer: Not isolated | Prisha Batra: Not clinical | – |

Table S2 continued

| Paper | Reviewer 1 | Reviewer 2 | Additional recorded reviews |
|---|---|---|---|
| 679. Feldman 2022. Cranberry Proanthocyanidins as a Therapeutic Strategy to Curb Metabolic Syndrome and Fatty Liver-Associated Disorders<br>PMID 36670951 | Michele Schaffer: No saffron | Prisha Batra: Not clinical | – |
| 680. Kumar 2023. Optically Active Nanomaterials and Its Biosensing Applications-A Review<br>PMID 36671920 | Michele Schaffer: Not clinical | Prisha Batra: Not clinical | – |
| 681. Fang 2023. Konjac Glucomannan: An Emerging Specialty Medical Food to Aid in the Treatment of Type 2 Diabetes Mellitus<br>PMID 36673456 | Michele Schaffer: Not isolated | Prisha Batra: Not clinical | – |
| 682. Szypowska 2023. Dietary Inflammatory Index (DII)(®) and Metabolic Syndrome in the Selected Population of Polish Adults: Results of the PURE Poland Sub-Study<br>PMID 36673811 | Michele Schaffer: No saffron | Prisha Batra: Not clinical | – |
| 683. Lakshmi 2023. Herbs as a Source for the Treatment of Polycystic Ovarian Syndrome: A Systematic Review<br>PMID 36648830 | Michele Schaffer: No saffron | Prisha Batra: Not clinical | – |
| 684. Elkattawy 2022. Rho-kinase inhibition ameliorates non-alcoholic fatty liver disease in type 2 diabetic rats<br>PMID 36047723 | Michele Schaffer: No saffron | Prisha Batra: Not clinical | – |
| 685. Saberi-Karimian 2023. Data mining approaches for type 2 diabetes mellitus prediction using anthropometric measurements<br>PMID 36510349 | Michele Schaffer: Not isolated | Prisha Batra: Not clinical | – |
| 686. Zuercher 2023. Dietary inflammatory index and cardiovascular disease risk in Hispanic women from the Women's Health Initiative<br>PMID 36631866 | Michele Schaffer: No outcome | Prisha Batra: Not clinical | – |
| 687. Zhao 2023. Insulin resistance in polycystic ovary syndrome across various tissues: an updated review of pathogenesis, evaluation, and treatment<br>PMID 36631836 | Michele Schaffer: Not isolated | Prisha Batra: Not clinical | – |
| 688. Sun 2023. Effects of antioxidants on diabetic kidney diseases: mechanistic interpretations and clinical assessment<br>PMID 36624538 | Michele Schaffer: Not clinical | Prisha Batra: Not clinical | – |
| 689. Tibarewal 2022. Long-term treatment of cancer-prone germline PTEN mutant mice with low-dose rapamycin extends lifespan and delays tumour development<br>PMID 36073856 | Michele Schaffer: No saffron | Prisha Batra: Not clinical | – |

Table S2 continued

| Paper | Reviewer 1 | Reviewer 2 | Additional recorded reviews |
|---|---|---|---|
| 690. Omale 2023. A systematic analysis of anti-diabetic medicinal plants from cells to clinical trials<br>PMID 36627919 | Michele Schaffer: Not clinical | Prisha Batra: Not clinical | – |
| 691. Yan 2022. Exploring the role of Tibetan medicinal formula Qishiwei Zhenzhu Pills (Ranasampel) against diabetes mellitus-linked cognitive impairment of db/db mice through serum pharmacochemistry and microarray data analysis<br>PMID 36620773 | Michele Schaffer: Not isolated | Prisha Batra: Not clinical | – |
| 692. Bergomi 2022. Insulin expression in $\beta$ cells is reduced within islets before islet loss in diabetic cats<br>PMID 35986507 | Michele Schaffer: No saffron | Prisha Batra: Not clinical | – |
| 693. Fadahunsi 2021. Phytochemistry, nutritional composition, and pharmacological activities of Thaumatococcus daniellii (Benth): a review<br>PMID 36605714 | Michele Schaffer: Not isolated | Prisha Batra: Not clinical | – |
| 694. Neto 2023. The complex relationship between obesity and neurodegenerative diseases: an updated review<br>PMID 38026693 | Michele Schaffer: Not isolated | Prisha Batra: Not clinical | – |
| 695. Maiese 2023. The impact of aging and oxidative stress in metabolic and nervous system disorders: programmed cell death and molecular signal transduction crosstalk<br>PMID 38022638 | Michele Schaffer: Not isolated | Prisha Batra: Not clinical | – |
| 696. Naz 2022. Hyperglycemia-associated Alzheimer's-like symptoms and other behavioral effects attenuated by Plumeria obtusa L. Extract in alloxan-induced diabetic rats<br>PMID 36588726 | Michele Schaffer: Not isolated | Prisha Batra: Not clinical | – |
| 697. Wu 2022. The sirtuin family in health and disease<br>PMID 36581622 | Michele Schaffer: Not clinical | Prisha Batra: Not clinical | – |
| 698. Ghanbari 2022. The effect of high intensity interval training with beetroot (Beta vulgaris) juice supplementation on serotonin and dopamine receptors expression, anxiety and depression in middle-aged diabetic rats<br>PMID 36583177 | Michele Schaffer: Not clinical | Prisha Batra: Not clinical | – |
| 699. Nafees 2022. Indian traditional medicinal plants in ophthalmic diseases<br>PMID 36583172 | Michele Schaffer: Not clinical | Prisha Batra: Not clinical | – |
| 700. Kim 2022. Marine Polyphenol Phlorotannins as a Natural Sleep Aid for Treatment of Insomnia: A Review of Sedative-Hypnotic Effects and Mechanism of Action<br>PMID 36547921 | Michele Schaffer: Not clinical | Prisha Batra: Not clinical | – |

Table S2 continued

| Paper | Reviewer 1 | Reviewer 2 | Additional recorded reviews |
|---|---|---|---|
| 701. Farouil 2022. The Metabolomic Profile of the Essential Oil from Zanthoxylum caribaeum (syn. chiloperone) Growing in Guadeloupe FWI using GC × GC-TOFMS<br>PMID 36557331 | Michele Schaffer: Not clinical | Prisha Batra: Not clinical | – |
| 702. Dini 2022. Recent Advances in Natural Polyphenol Research<br>PMID 36557912 | Michele Schaffer: Not isolated | Prisha Batra: Not clinical | – |
| 703. Cerdá-Bernad 2022. Saffron against Neuro-Cognitive Disorders: An Overview of Its Main Bioactive Compounds, Their Metabolic Fate and Potential Mechanisms of Neurological Protection<br>PMID 36558528 | Michele Schaffer: Not clinical | Prisha Batra: Not clinical | – |
| 704. Nacka-Aleksić 2022. The Role of Dietary Polyphenols in Pregnancy and Pregnancy-Related Disorders<br>PMID 36558404 | Michele Schaffer: Not isolated | Prisha Batra: Not clinical | – |
| 705. Ali 2022. Boosting Tadalafil Bioavailability via Sono-Assisted Nano-Emulsion-Based Oral Jellies: Box-Behnken Optimization and Assessment<br>PMID 36559086 | Michele Schaffer: Not clinical | Prisha Batra: Not clinical | – |
| 706. Imtiaz 2022. Prediction of $\alpha$-Glucosidase Inhibitory Activity of LC-ESI-TQ-MS/MS-Identified Compounds from Tradescantia pallida Leaves<br>PMID 36559071 | Michele Schaffer: Not isolated | Prisha Batra: Not clinical | – |
| 707. Foshati 2022. The efficacy and safety of ginger supplementation in patients with multiple sclerosis: A rationale and study protocol for a double-blind randomized controlled trial<br>PMID 36570345 | Michele Schaffer: Not isolated | Prisha Batra: No saffron | – |
| 708. Hosseini 2022. LC-MS-based metabolite profiling of aqueous extract of Pergularia tomentosa L. and its anti-hyperglycemic effect<br>PMID 36544524 | Michele Schaffer: Not isolated | Prisha Batra: Not clinical | – |
| 709. Vangoori 2022. A Review on Drug Induced Obesity and Rodent Experimental Models of Obesity in Animals<br>PMID 36540593 | Michele Schaffer: Not isolated | Prisha Batra: Not clinical | – |
| 710. Karampela 2023. Perspectives in vaccines, immune response, therapeutic interventions and COVID-19<br>PMID 36570684 | Michele Schaffer: Not isolated | Prisha Batra: Not clinical | – |

Table S2 continued

| Paper | Reviewer 1 | Reviewer 2 | Additional recorded reviews |
|---|---|---|---|
| 711. Mohammadi 2023. An overview on role of nutrition on COVID-19 immunity: Accumulative review from available studies<br>PMID 36540357 | Michele Schaffer: Not isolated | Prisha Batra: Not clinical | – |
| 712. Cheshmeh 2022. Effect of green cardamom on the expression of genes implicated in obesity and diabetes among obese women with polycystic ovary syndrome: a double blind randomized controlled trial<br>PMID 36522620 | Michele Schaffer: Not isolated | Prisha Batra: No saffron | – |
| 713. Chua 2022. Utility of Human Relevant Preclinical Animal Models in Navigating NAFLD to MAFLD Paradigm<br>PMID 36499091 | Michele Schaffer: Not isolated | Prisha Batra: Not clinical | – |
| 714. Theodosis-Nobelos 2022. The Multiple Sclerosis Modulatory Potential of Natural Multi-Targeting Antioxidants<br>PMID 36500494 | Michele Schaffer: No saffron | Prisha Batra: Not clinical | – |
| 715. Zhou 2022. Efficacy of cinnamon supplementation on glycolipid metabolism in T2DM diabetes: A meta-analysis and systematic review<br>PMID 36505061 | Michele Schaffer: Not isolated | Prisha Batra: Not clinical | – |
| 716. Aragam 2022. Discovery and systematic characterization of risk variants and genes for coronary artery disease in over a million participants<br>PMID 36474045 | Michele Schaffer: No saffron | Prisha Batra: Not clinical | – |
| 717. Karamali 2022. Potential therapeutic strategies for photoreceptor degeneration: the path to restore vision<br>PMID 36476500 | Michele Schaffer: Not isolated | Prisha Batra: Not clinical | – |
| 718. Lozano 2022. Associations of the Dietary Inflammatory Index with total adiposity and ectopic fat through the gut microbiota, LPS, and C-reactive protein in the Multiethnic Cohort-Adiposity Phenotype Study<br>PMID 34871345 | Michele Schaffer: No saffron | Prisha Batra: Not clinical | – |
| 719. Dana 2022. Effects of empagliflozin on the expression of kisspeptin gene and reproductive system function in streptozotocin-induced diabetic male rats<br>PMID 36479221 | Michele Schaffer: Not isolated | Prisha Batra: Not clinical | – |
| 720. Li 2022. Role of potential bioactive metabolites from traditional Chinese medicine for type 2 diabetes mellitus: An overview<br>PMID 36479195 | Michele Schaffer: Not isolated | Prisha Batra: Not clinical | – |

Table S2 continued

| Paper | Reviewer 1 | Reviewer 2 | Additional recorded reviews |
|---|---|---|---|
| 721. Ebrahimian 2022. Effects of Portulaca oleracea L. (purslane) on the metabolic syndrome: A review<br>PMID 36474567 | Michele Schaffer: Not isolated | Prisha Batra: Not clinical | – |
| 722. Afzaal 2022. Virtual screening and drug repositioning of FDA-approved drugs from the ZINC database to identify the potential hTERT inhibitors<br>PMID 36467041 | Michele Schaffer: Not isolated | Prisha Batra: Not clinical | – |
| 723. Izhar Ariff Mohd Kashim 2023. Scientific and Islamic perspectives in relation to the Halal status of cultured meat<br>PMID 36466219 | Michele Schaffer: No saffron | Prisha Batra: Not clinical | – |
| 724. Wu 2022. Amyloid Cascade Hypothesis for the Treatment of Alzheimer's Disease: Progress and Challenges<br>PMID 36465173 | Michele Schaffer: Not isolated | Prisha Batra: Not clinical | – |
| 725. Rausch 2022. The Farnesoid X Receptor as a Master Regulator of Hepatotoxicity<br>PMID 36430444 | Michele Schaffer: Not isolated | Bryan Anh Nguyen: No saffron | – |
| 726. Amin 2022. Chemical Profiling and Molecular Docking Study of Agathophora alopecuroides<br>PMID 36430987 | Michele Schaffer: Not clinical | Bryan Anh Nguyen: No saffron | – |
| 727. Giani 2022. Carbon Source Influences Antioxidant, Antiglycemic, and Antilipidemic Activities of Haloferax mediterranei Carotenoid Extracts<br>PMID 36354982 | Michele Schaffer: No saffron | Bryan Anh Nguyen: No saffron | – |
| 728. Brandão-Lima 2022. Circulating microRNA Related to Cardiometabolic Risk Factors for Metabolic Syndrome: A Systematic Review<br>PMID 36355127 | Michele Schaffer: Not isolated | Bryan Anh Nguyen: No saffron | – |
| 729. Sapian 2022. The Role of Anthocyanin in Modulating Diabetic Cardiovascular Disease and Its Potential to Be Developed as a Nutraceutical<br>PMID 36355516 | Michele Schaffer: Not clinical | Bryan Anh Nguyen: No saffron | – |
| 730. Gonçalves 2022. Employ of Anthocyanins in Nanocarriers for Nano Delivery: In Vitro and In Vivo Experimental Approaches for Chronic Diseases<br>PMID 36365091 | Michele Schaffer: Not clinical | Bryan Anh Nguyen: No saffron | – |
| 731. Gani 2022. Upscaling of Apple By-Product by Utilising Apple Seed Protein as a Novel Wall Material for Encapsulation of Chlorogenic Acid as Model Bioactive Compound<br>PMID 36429294 | Grace Kim: No saffron | Michele Schaffer: Not isolated | – |

Table S2 continued

| Paper | Reviewer 1 | Reviewer 2 | Additional recorded reviews |
|---|---|---|---|
| 732. Kim 2022. Inverse association of a traditional Korean diet composed of a multigrain rice-containing meal with fruits and nuts with metabolic syndrome risk: The KoGES PMID 36438748 | Grace Kim: No saffron | Michele Schaffer: No saffron | – |
| 733. Zhao 2022. Natural compounds targeting glycolysis as promising therapeutics for gastric cancer: A review PMID 36438836 | Grace Kim: Not clinical | Michele Schaffer: Not clinical | – |
| 734. Lepić 2022. A review of the diet, nutrients, and supplementation potential for the outcome augmentation in surgical treatment of peripheral nerve injuries PMID 36439529 | Michele Schaffer: Not isolated | Bryan Anh Nguyen: No saffron | – |
| 735. Gao 2022. Dietary Oregano Essential Oil Supplementation Influences Production Performance and Gut Microbiota in Late-Phase Laying Hens Fed Wheat-Based Diets PMID 36359131 | Michele Schaffer: Not isolated | Bryan Anh Nguyen: No saffron | – |
| 736. Gomaa 2022. Polyphenols from Conyza dioscoridis (L.) ameliorate Alzheimer's disease- like alterations through multi-targeting activities in two animal models PMID 36348329 | Michele Schaffer: Retracted | Bryan Anh Nguyen: No saffron | – |
| 737. Ma 2022. Roles of physical exercise-induced MiR-126 in cardiovascular health of type 2 diabetes PMID 36376958 | Michele Schaffer: Not isolated | Bryan Anh Nguyen: No saffron | – |
| 738. Dolgacheva 2022. Molecular and Cellular Interactions in Pathogenesis of Sporadic Parkinson Disease PMID 36361826 | Michele Schaffer: Not isolated | Bryan Anh Nguyen: No saffron | – |
| 739. Kaczmarczyk 2022. The Importance of Nutritional Aspects in the Assessment of Inflammation and Intestinal Barrier in Patients with Inflammatory Bowel Disease PMID 36364889 | Michele Schaffer: No saffron | Bryan Anh Nguyen: No saffron | – |
| 740. Ahmed 2022. Luteolin loaded on zinc oxide nanoparticles ameliorates non-alcoholic fatty liver disease associated with insulin resistance in diabetic rats via regulation of PI3K/AKT/FoxO1 pathway PMID 36319192 | Michele Schaffer: Not clinical | Bryan Anh Nguyen: No saffron | – |
| 741. Dias de Castro 2022. Dietary parameters in patients with drug allergy: Assessing dietary inflammatory index PMID 36327304 | Michele Schaffer: No saffron | Bryan Anh Nguyen: No saffron | – |
| 742. Al-Shaeli 2022. Effect of honey bee venom on the histological changes of testes and hormonal disturbance in diabetic mice PMID 36341058 | Michele Schaffer: Not clinical | Bryan Anh Nguyen: No saffron | – |

Table S2 continued

| Paper | Reviewer 1 | Reviewer 2 | Additional recorded reviews |
|---|---|---|---|
| 743. Yan 2022. Current advances on the phytochemical composition, pharmacologic effects, toxicology, and product development of Phyllanthi Fructus PMID 36339628 | Michele Schaffer: Not isolated | Bryan Anh Nguyen: No saffron | – |
| 744. Zheng 2022. Systematic review of Lonicerae Japonicae Flos: A significant food and traditional Chinese medicine PMID 36339557 | Michele Schaffer: Not clinical | Bryan Anh Nguyen: No saffron | – |
| 745. Lei 2022. Bergenin inhibits palmitic acid-induced pancreatic $\beta$-cell inflammatory death via regulating NLRP3 inflammasome activation PMID 36330410 | Bryan Anh Nguyen: No saffron | Prisha Batra: Not clinical | – |
| 746. Karigidi 2021. Effects of Curculigo pilosa supplementation on antioxidant and antidiabetic activities of yam flour PMID 34538895 | Bryan Anh Nguyen: No saffron | Prisha Batra: Not clinical | – |
| 747. Renai 2022. Combining Feature-Based Molecular Networking and Contextual Mass Spectral Libraries to Decipher Nutrimetabolomics Profiles PMID 36295906 | Bryan Anh Nguyen: No saffron | Prisha Batra: Not clinical | – |
| 748. Milluzzo 2022. Do Nutrients and Nutraceuticals Play a Role in Diabetic Retinopathy? A Systematic Review PMID 36297113 | Bryan Anh Nguyen: No saffron | Prisha Batra: Not clinical | – |
| 749. Ahmad 2022. Ameliorative effects of half-dose saffron and chamomile combination on Psycho-endocrinological changes in a diabetic murine model PMID 36302045 | Bryan Anh Nguyen: Not clinical | Prisha Batra: Not clinical | – |
| 750. Bakrim 2022. Health Benefits and Pharmacological Properties of Stigmasterol PMID 36290632 | Bryan Anh Nguyen: No saffron | Prisha Batra: Not clinical | – |
| 751. Zhang 2022. Observation on clinical effect of Huoxue-Jiangtang decoction formula granules in treating prediabetes: a randomized prospective placebo-controlled double-blind trial protocol PMID 36261813 | Bryan Anh Nguyen: No saffron | Prisha Batra: No saffron | – |
| 752. Ramos-Lopez 2022. The Role of Nutrition on Meta-inflammation: Insights and Potential Targets in Communicable and Chronic Disease Management PMID 36258149 | Bryan Anh Nguyen: No saffron | Prisha Batra: Not clinical | – |
| 753. Harirchian 2022. Diet and disease-related outcomes in multiple sclerosis: A systematic review of clinical trials PMID 38011464 | Bryan Anh Nguyen: No saffron | Prisha Batra: Not clinical | – |

Table S2 continued

| Paper | Reviewer 1 | Reviewer 2 | Additional recorded reviews |
|---|---|---|---|
| 754. Jamil 2022. Carduus edelbergii Rech. f. Mediated Fabrication of Gold Nanoparticles; Characterization and Evaluation of Antimicrobial, Antioxidant and Antidiabetic Potency of the Synthesized AuNPs<br>PMID 36235206 | Bryan Anh Nguyen: No saffron | Prisha Batra: Not clinical | – |
| 755. Gasmi 2022. Polyphenols in Metabolic Diseases<br>PMID 36234817 | Bryan Anh Nguyen: No saffron | Prisha Batra: Not clinical | – |
| 756. Zhou 2022. Molecular Mechanistic Pathways Targeted by Natural Compounds in the Prevention and Treatment of Diabetic Kidney Disease<br>PMID 36234757 | Bryan Anh Nguyen: No saffron | Prisha Batra: Not clinical | – |
| 757. Yin 2022. Recent Progress on Techniques in the Detection of Aflatoxin B(1) in Edible Oil: A Mini Review<br>PMID 36234684 | Bryan Anh Nguyen: No saffron | Prisha Batra: Not clinical | – |
| 758. Tieu 2022. Immunoceuticals: Harnessing Their Immunomodulatory Potential to Promote Health and Wellness<br>PMID 36235727 | Bryan Anh Nguyen: No saffron | Prisha Batra: Not clinical | – |
| 759. Zhang 2022. Regulation of nuclear factor erythroid-2-related factor 2 as a potential therapeutic target in intracerebral hemorrhage<br>PMID 36245922 | Bryan Anh Nguyen: No saffron | Prisha Batra: Not clinical | – |
| 760. Marhuenda-Muñoz 2022. Circulating carotenoids are associated with favorable lipid and fatty acid profiles in an older population at high cardiovascular risk<br>PMID 36245542 | Bryan Anh Nguyen: No saffron | Prisha Batra: Not clinical | – |
| 761. Mandal 2023. Critical Review on Nutritional, Bioactive, and Medicinal Potential of Spices and Herbs and Their Application in Food Fortification and Nanotechnology<br>PMID 36219334 | Bryan Anh Nguyen: No saffron | Prisha Batra: Not clinical | – |
| 762. Jaafarinia 2022. Evaluation of therapeutic effects of crocin in attenuating the progression of diabetic nephropathy: a preliminary randomized triple-blind placebo-controlled trial<br>PMID 36209091 | Bryan Anh Nguyen: Candidate | Prisha Batra: Candidate | – |
| 763. Deshayes 2022. Role of non-invasive methods in detecting liver impairment in familial Mediterranean fever adult patients with persistent hepatic cytolysis<br>PMID 36198677 | Bryan Anh Nguyen: No saffron | Prisha Batra: Not clinical | – |
| 764. Deng 2022. Roles of traditional chinese medicine regulating neuroendocrinology on AD treatment<br>PMID 36213283 | Bryan Anh Nguyen: No saffron | Prisha Batra: Not clinical | – |

Table S2 continued

| Paper | Reviewer 1 | Reviewer 2 | Additional recorded reviews |
|---|---|---|---|
| 765. Fang 2022. The AMPK pathway in fatty liver disease PMID 36203933 | Bryan Anh Nguyen: No saffron | Prisha Batra: Not clinical | – |
| 766. Hu 2022. Medicinal and edible plants in the treatment of dyslipidemia: advances and prospects PMID 36175900 | Bryan Anh Nguyen: No saffron | Prisha Batra: Not clinical | – |
| 767. Ji 2022. Therapeutic potential of traditional Chinese medicine for the treatment of NAFLD: A promising drug Potentilla discolor Bunge PMID 36176915 | Bryan Anh Nguyen: No saffron | Prisha Batra: Not clinical | – |
| 768. Yang 2022. Potential lipolytic regulators derived from natural products as effective approaches to treat obesity PMID 36176469 | Bryan Anh Nguyen: No saffron | Prisha Batra: Not clinical | – |
| 769. Millar 2022. A proinflammatory diet is associated with increased odds of frailty after 12-year follow-up in a cohort of adults PMID 34558613 | Bryan Anh Nguyen: No saffron | Prisha Batra: Not clinical | – |
| 770. Rakhshandeh 2022. Protective Effect of Portulaca oleracea on Streptozotocin-Induced Type I Diabetes-Associated Reproductive System Dysfunction and Inflammation PMID 36144807 | Bryan Anh Nguyen: No saffron | Prisha Batra: Not clinical | – |
| 771. Metibemu 2022. Carotenoids in Drug Discovery and Medicine: Pathways and Molecular Targets Implicated in Human Diseases PMID 36144741 | Bryan Anh Nguyen: No saffron | Prisha Batra: Not clinical | – |
| 772. Farzana 2022. Phenolic Constituents from Wendlandia tinctoria var. grandis (Roxb.) DC. Stem Deciphering Pharmacological Potentials against Oxidation, Hyperglycemia, and Diarrhea: Phyto-Pharmacological and Computational Approaches PMID 36144691 | Bryan Anh Nguyen: No saffron | Prisha Batra: Not clinical | – |
| 773. Trandafir 2022. Tackling Dyslipidemia in Obesity from a Nanotechnology Perspective PMID 36145147 | Bryan Anh Nguyen: No saffron | Prisha Batra: Not clinical | – |
| 774. Salinas-Roca 2022. Polyphenol Intake in Pregnant Women on Gestational Diabetes Risk and Neurodevelopmental Disorders in Offspring: A Systematic Review PMID 36145129 | Bryan Anh Nguyen: No saffron | Prisha Batra: Not clinical | – |

Table S2 continued

| Paper | Reviewer 1 | Reviewer 2 | Additional recorded reviews |
|---|---|---|---|
| 775. Puttasiddaiah 2022. Advances in Nanofabrication Technology for Nutraceuticals: New Insights and Future Trends PMID 36135026 | Bryan Anh Nguyen: No saffron | Prisha Batra: Not clinical | – |
| 776. Yan 2022. The Nicotinamide/Streptozotocin Rodent Model of Type 2 Diabetes: Renal Pathophysiology and Redox Imbalance Features PMID 36139064 | Bryan Anh Nguyen: No saffron | Prisha Batra: Not clinical | – |
| 777. Liu 2022. Synergistic Hypolipidemic Effects and Mechanisms of Phytochemicals: A Review PMID 36140902 | Bryan Anh Nguyen: No saffron | Prisha Batra: Not clinical | – |
| 778. Omidkhoda 2022. Saffron and its active ingredients against human disorders: A literature review on existing clinical evidence PMID 36159329 | Bryan Anh Nguyen: No saffron | Prisha Batra: Not clinical | – |
| 779. Hassanzadeh-Taheri 2021. Investigating the effect of ethanolic extract of Commiphora myrrha (Nees) Engl. gum-resin against hepatorenal injury in diabetic rats PMID 34900809 | Bryan Anh Nguyen: No saffron | Prisha Batra: Not clinical | – |
| 780. Zhang 2022. Integrating pharmacological evaluation and computational identification for deciphering the action mechanism of Yunpi-Huoxue-Sanjie formula alleviates diabetic cardiomyopathy PMID 36147338 | Bryan Anh Nguyen: No saffron | Prisha Batra: Not clinical | – |
| 781. Olga 2022. Lipid profiling analyses from mouse models and human infants PMID 36115026 | Bryan Anh Nguyen: No saffron | Prisha Batra: Not clinical | – |
| 782. Singh 2022. A review on health benefits of phenolics derived from dietary spices PMID 36132490 | Bryan Anh Nguyen: No saffron | Prisha Batra: Not clinical | – |
| 783. Dulan 2022. Famous traditional Mongolian medicine Xieriga-4 (Turmeric-4) decoction: A review PMID 36118011 | Bryan Anh Nguyen: No saffron | Prisha Batra: Not clinical | – |
| 784. Xie 2022. Gastroprotective mechanism of modified lvdou gancao decoction on ethanol-induced gastric lesions in mice: Involvement of Nrf-2/HO-1/NF-$\kappa$B signaling pathway PMID 36120337 | Bryan Anh Nguyen: No saffron | Prisha Batra: Not clinical | – |
| 785. Wang 2022. LncRNA H19 inhibits ER stress induced apoptosis and improves diabetic cardiomyopathy by regulating PI3K/AKT/mTOR axis PMID 36044268 | Bryan Anh Nguyen: No saffron | Prisha Batra: Not clinical | – |

Table S2 continued

| Paper | Reviewer 1 | Reviewer 2 | Additional recorded reviews |
|---|---|---|---|
| 786. Karimi 2022. The Effect of Saffron (Crocus sativus L.) Supplementation on Renal Function: A Systematic Review and Meta-Analysis of Randomized Controlled Clinical Trials<br>PMID 36105788 | Bryan Anh Nguyen: Not clinical | Prisha Batra: Not clinical | – |
| 787. Roham 2022. Human islet amyloid polypeptide: A therapeutic target for the management of type 2 diabetes mellitus<br>PMID 36105173 | Bryan Anh Nguyen: No saffron | Prisha Batra: Not clinical | – |
| 788. Nakhate 2022. Therapeutic Potential and Pharmaceutical Development of a Multitargeted Flavonoid Phloretin<br>PMID 36079895 | Bryan Anh Nguyen: No saffron | Prisha Batra: Not clinical | – |
| 789. Agarwal 2022. Comparing the Effects of Concord Grape (Vitis labrusca L.) Puree, Juice, and Pomace on Intestinal Morphology, Functionality, and Bacterial Populations In Vivo (Gallus gallus)<br>PMID 36079797 | Bryan Anh Nguyen: No saffron | Prisha Batra: Not clinical | – |
| 790. Arzi 2022. Countering Triple Negative Breast Cancer via Impeding Wnt/$\beta$-Catenin Signaling, a Phytotherapeutic Approach<br>PMID 36079579 | Bryan Anh Nguyen: No saffron | Prisha Batra: Not clinical | – |
| 791. Chandel 2022. Current Advancements in Pectin: Extraction, Properties and Multifunctional Applications<br>PMID 36076865 | Bryan Anh Nguyen: No saffron | Prisha Batra: Not clinical | – |
| 792. Machado 2022. Browning of the white adipose tissue regulation: new insights into nutritional and metabolic relevance in health and diseases<br>PMID 36068578 | Bryan Anh Nguyen: No saffron | Prisha Batra: Not clinical | – |
| 793. Jin 2022. The Efficacy and Safety of Dingkun Pill in Women with Polycystic Ovary Syndrome: A Systematic Review and Meta-Analysis of Randomized Controlled Trials<br>PMID 36062168 | Bryan Anh Nguyen: No saffron | Prisha Batra: Not clinical | – |
| 794. Lu 2022. The landscape of potential health benefits of carotenoids as natural supportive therapeutics in protecting against Coronavirus infection<br>PMID 36058151 | Bryan Anh Nguyen: No saffron | Prisha Batra: Not clinical | – |
| 795. Vecchia 2022. Sonchus oleraceus L. Promotes Gastroprotection in Rodents via Antioxidant, Anti-Inflammatory, and Antisecretory Activities<br>PMID 36051492 | Bryan Anh Nguyen: No saffron | Prisha Batra: Not clinical | – |

Table S2 continued

| Paper | Reviewer 1 | Reviewer 2 | Additional recorded reviews |
|---|---|---|---|
| 796. Liang 2022. Relationship between quality of life and adolescent glycolipid metabolism disorder: A cohort study PMID 36051423 | Bryan Anh Nguyen: No saffron | Prisha Batra: Not clinical | – |
| 797. #N/A 2022. 21st ISoP Annual Meeting "A New Era of Pharmacovigilance: Challenges and Opportunities" 20–23 September 2022 Verona, Italy PMC9421640 | Bryan Anh Nguyen: No saffron | Prisha Batra: Not clinical | – |
| 798. Mathakala 2022. Halophila beccarii extract ameliorate glucose uptake in 3T3-L1 adipocyte cells and improves glucose homeostasis in streptozotocin-induced diabetic rats PMID 36042748 | Bryan Anh Nguyen: No saffron | Prisha Batra: Not clinical | – |
| 799. Jeziorek 2022. The Effect of a Low-Carbohydrate, High-Fat Diet versus Moderate-Carbohydrate and Fat Diet on Body Composition in Patients with Lipedema PMID 36035515 | Bryan Anh Nguyen: No saffron | Prisha Batra: Not clinical | – |
| 800. Batiha 2022. Rhus coriaria L. (Sumac), a Versatile and Resourceful Food Spice with Cornucopia of Polyphenols PMID 36014419 | Bryan Anh Nguyen: No saffron | Prisha Batra: Not clinical | – |
| 801. Mustapha 2022. Inhibition of Endoplasmic Reticulum Stress Improves Acetylcholine-Mediated Relaxation in the Aorta of Type-2 Diabetic Rats PMID 36014347 | Bryan Anh Nguyen: No saffron | Prisha Batra: Not clinical | – |
| 802. Muscoli 2022. The New Role of SGLT2 Inhibitors in the Management of Heart Failure: Current Evidence and Future Perspective PMID 36015359 | Bryan Anh Nguyen: No saffron | Prisha Batra: Not clinical | – |
| 803. Hossain 2022. Propolis: An update on its chemistry and pharmacological applications PMID 36028892 | Bryan Anh Nguyen: No saffron | Prisha Batra: Not clinical | – |
| 804. Fu 2022. Antioxidant and Anti-Inflammatory Properties of Hydroxyl Safflower Yellow a in Diabetic Nephropathy: A Meta-Analysis of Randomized Controlled Trials PMID 36034830 | Bryan Anh Nguyen: No saffron | Prisha Batra: Not clinical | – |
| 805. Xie 2022. Crocin-I Protects Against High-Fat Diet-Induced Obesity via Modulation of Gut Microbiota and Intestinal Inflammation in Mice PMID 36034852 | Bryan Anh Nguyen: Not clinical | Prisha Batra: Not clinical | – |
| 806. Khoshandam 2022. Interaction of saffron and its constituents with Nrf2 signaling pathway: A review PMID 36033950 | Bryan Anh Nguyen: No saffron | Prisha Batra: Not clinical | – |

Table S2 continued

| Paper | Reviewer 1 | Reviewer 2 | Additional recorded reviews |
|---|---|---|---|
| 807. Aramjoo 2022. Antihyperglycemic and hepatoprotective effects of Salvia tebesana Bunge in diabetic rats<br>PMID 36034081 | Bryan Anh Nguyen: No saffron | Prisha Batra: Not clinical | – |
| 808. Heurtaux 2022. Normal and Pathological NRF2 Signalling in the Central Nervous System<br>PMID 35892629 | Bryan Anh Nguyen: No saffron | Prisha Batra: Not clinical | – |
| 809. Shahbaz 2022. Crocins for Ischemic Stroke: A Review of Current Evidence<br>PMID 35991882 | Bryan Anh Nguyen: No saffron | Prisha Batra: Not clinical | – |
| 810. Jin 2022. Corresponding risk factors between cognitive impairment and type 1 diabetes mellitus: A narrative review<br>PMID 35991978 | Bryan Anh Nguyen: No saffron | Prisha Batra: Not clinical | – |
| 811. Mahmod 2022. Chemoprevention effect of the Mediterranean diet on colorectal cancer: Current studies and future prospects<br>PMID 35990343 | Bryan Anh Nguyen: No saffron | Prisha Batra: Not clinical | – |
| 812. Kianmehr 2022. The effects of Crocus sativus L. (Saffron) and its ingredients on dietary intakes in cardiovascular disease in Iranian population: A systematic review and meta-analysis<br>PMID 35990354 | Bryan Anh Nguyen: No saffron | Prisha Batra: Not clinical | – |
| 813. Lanspa 2022. A Systematic Review of Nutritional Interventions on Key Cytokine Pathways in Rheumatoid Arthritis and Its Implications for Comorbid Depression: Is a More Comprehensive Approach Required?<br>PMID 35990558 | Bryan Anh Nguyen: No saffron | Prisha Batra: Not clinical | – |
| 814. Luo 2022. Reducing VEGFB expression regulates the balance of glucose and lipid metabolism in mice via VEGFR1<br>PMID 35894135 | Bryan Anh Nguyen: No saffron | Prisha Batra: Not clinical | – |
| 815. Shi 2022. Beneficial Effects of lncRNA-UC.360+ shRNA on Diabetic Cardiac Sympathetic Damage via NLRP3 Inflammasome-Induced Pyroptosis in Stellate Ganglion<br>PMID 35967043 | Bryan Anh Nguyen: No saffron | Prisha Batra: Not clinical | – |
| 816. Sahebnasagh 2022. Neurohormetic phytochemicals in the pathogenesis of neurodegenerative diseases<br>PMID 35953850 | Bryan Anh Nguyen: No saffron | Prisha Batra: Not clinical | – |
| 817. Hur 2022. Association of Polygenic Variants with Type 2 Diabetes Risk and Their Interaction with Lifestyles in Asians<br>PMID 35956399 | Bryan Anh Nguyen: No saffron | Prisha Batra: Not clinical | – |

Table S2 continued

| Paper | Reviewer 1 | Reviewer 2 | Additional recorded reviews |
|---|---|---|---|
| 818. Zasowska-Nowak 2022. Carotenoids in Palliative Care-Is There Any Benefit from Carotenoid Supplementation in the Adjuvant Treatment of Cancer-Related Symptoms? PMID 35956359 | Bryan Anh Nguyen: No saffron | Prisha Batra: Not clinical | – |
| 819. Liu 2022. Sparsely Granulated Corticotroph Pituitary Macroadenoma Presenting With Pituitary Apoplexy Resulting in Remission of Hypercortisolism PMID 35959088 | Bryan Anh Nguyen: No saffron | Prisha Batra: Not clinical | – |
| 820. Fakhri 2022. Effect of Medicinal Plants and Natural Products on Liver Enzymes in Non-alcoholic Fatty Liver Patients in Iran: A Systematic Review and Meta-Analysis PMID 35958359 | Bryan Anh Nguyen: No saffron | Prisha Batra: Not clinical | – |
| 821. Liu 2022. Gardenia jasminoides J. Ellis extract GJ-4 attenuates hyperlipidemic vascular dementia in rats via regulating PPAR-$\gamma$-mediated microglial polarization PMID 35950104 | Bryan Anh Nguyen: Not clinical | Prisha Batra: Not clinical | – |
| 822. Salaramoli 2022. The effects of ginger and its constituents in the prevention of metabolic syndrome: A review PMID 35949312 | Bryan Anh Nguyen: No saffron | Prisha Batra: Not clinical | – |
| 823. Kuo 2019. Identification of baicalin from Bofutsushosan and Daisaikoto as a potent inducer of glucose uptake and modulator of insulin signaling-associated pathways PMID 30648577 | Bryan Anh Nguyen: No saffron | Prisha Batra: Not clinical | – |
| 824. Mard 2022. Therapeutic Effect of Crocin on the NASH Model by Targeting the Fas Receptor Signaling Pathway PMID 35786619 | Bryan Anh Nguyen: No saffron | Prisha Batra: Not clinical | – |
| 825. Sani 2022. A review of the anti-diabetic potential of saffron PMID 35911474 | Bryan Anh Nguyen: Not clinical | Prisha Batra: Not clinical | – |
| 826. van den Hoek 2022. Unraveling the Transcriptional Dynamics of NASH Pathogenesis Affecting Atherosclerosis PMID 35897797 | Bryan Anh Nguyen: No saffron | Prisha Batra: Not clinical | – |
| 827. Samakar 2022. A review of the effects of Urtica dioica (nettle) in metabolic syndrome PMID 35911652 | Bryan Anh Nguyen: No saffron | Prisha Batra: Not clinical | – |
| 828. Brunetti 2022. Food Industry Byproducts as Starting Material for Innovative, Green Feed Formulation: A Sustainable Alternative for Poultry Feeding PMID 35897911 | Bryan Anh Nguyen: No saffron | Prisha Batra: Not clinical | – |

Table S2 continued

| Paper | Reviewer 1 | Reviewer 2 | Additional recorded reviews |
|---|---|---|---|
| 829. Zhang 2022. Angiopoietin-like protein 4 treated bone marrow-derived mesenchymal stem cells alleviate myocardial injury of patients with myocardial infarction PMID 35157362 | Bryan Anh Nguyen: No saffron | Prisha Batra: Not clinical | – |
| 830. Remigante 2022. Oxidation Stress as a Mechanism of Aging in Human Erythrocytes: Protective Effect of Quercetin PMID 35887126 | Bryan Anh Nguyen: No saffron | Prisha Batra: Not clinical | – |
| 831. Kozlovskaya 2022. Comparative Study of the Pharmacological Properties of Luteolin and Its 7,3'-Disulfate PMID 35877719 | Bryan Anh Nguyen: No saffron | Prisha Batra: Not clinical | – |
| 832. Sivani 2022. Reconnoitering the Therapeutic Role of Curcumin in Disease Prevention and Treatment: Lessons Learnt and Future Directions PMID 35888763 | Bryan Anh Nguyen: No saffron | Prisha Batra: Not clinical | – |
| 833. Guo 2022. Lactobacillus paracasei CCFM1223 Protects against Lipopolysaccharide-Induced Acute Liver Injury in Mice by Regulating the "Gut-Liver" Axis PMID 35889040 | Bryan Anh Nguyen: No saffron | Prisha Batra: Not clinical | – |
| 834. Sultana 2022. Ethnobotanical Uses, Phytochemistry, Toxicology, and Pharmacological Properties of Euphorbia neriifolia Linn. against Infectious Diseases: A Comprehensive Review PMID 35889247 | Bryan Anh Nguyen: No saffron | Prisha Batra: Not clinical | – |
| 835. Albrahim 2022. Effect of Blueberry Extract on Liver in Aged Rats PMID 35898615 | Bryan Anh Nguyen: No saffron | Prisha Batra: Not clinical | – |
| 836. Karimzadeh 2023. Effects of concentrated beetroot juice consumption on glycemic control, blood pressure, and lipid profile in type 2 diabetes patients: randomized clinical trial study PMID 35869311 | Bryan Anh Nguyen: No saffron | Prisha Batra: Not clinical | – |
| 837. Komolafe 2021. Nutritional supplementation for nonalcohol-related fatty liver disease: a network meta-analysis PMID 34280304 | Bryan Anh Nguyen: No saffron | Prisha Batra: Not clinical | – |
| 838. Faria 2020. Acute and subacute oral toxicity assessment of dry encapsulated and non-encapsulated green coffee fruit extracts PMID 35696115 | Bryan Anh Nguyen: No saffron | Prisha Batra: Not clinical | – |
| 839. Sawong 2022. Calotropis gigantea stem bark extracts inhibit liver cancer induced by diethylnitrosamine PMID 35840761 | Bryan Anh Nguyen: No saffron | Prisha Batra: Not clinical | – |

Table S2 continued

| Paper | Reviewer 1 | Reviewer 2 | Additional recorded reviews |
|---|---|---|---|
| 840. Gharanjik 2022. The Effect of Hydroalcoholic Calendula Officinalis Extract on Androgen-Induced Polycystic Ovary Syndrome Model in Female Rat<br>PMID 35845946 | Bryan Anh Nguyen: No saffron | Prisha Batra: Not clinical | – |
| 841. Wang 2022. Study on Antidepressant Effect and Mechanism of Crocin Mediated by the mTOR Signaling Pathway<br>PMID 35804209 | Bryan Anh Nguyen: Not clinical | Prisha Batra: Not clinical | – |
| 842. Li 2022. G Allele of the rs1801282 Polymorphism in PPAR$\gamma$ Gene Confers an Increased Risk of Obesity and Hypercholesterolemia, While T Allele of the rs3856806 Polymorphism Displays a Protective Role Against Dyslipidemia: A Systematic Review and Meta-Analysis<br>PMID 35846293 | Bryan Anh Nguyen: No saffron | Prisha Batra: Not clinical | – |
| 843. Kotowska 2022. IL-6 and HSPA1A Gene Polymorphisms May Influence the Levels of the Inflammatory and Oxidative Stress Parameters and Their Response to a Chronic Swimming Training<br>PMID 35805787 | Grace Kim: No saffron | Bryan Anh Nguyen: No saffron | Prisha Batra: Not clinical |
| 844. Park 2022. Naringenin and Phytoestrogen 8-Prenylnaringenin Protect against Islet Dysfunction and Inhibit Apoptotic Signaling in Insulin-Deficient Diabetic Mice<br>PMID 35807469 | Grace Kim: No saffron | Pranav Potluri: No saffron | Prisha Batra: Not clinical |
| 845. Lopresti 2022. An examination into the mental and physical effects of a saffron extract (affron®) in recreationally-active adults: A randomized, double-blind, placebo-controlled study<br>PMID 35813851 | Grace Kim: No outcome | Pranav Potluri: Intervention + outcome but no endpoint outcome | Prisha Batra: Not clinical |
| 846. Zhao 2022. A Mechanistic Exploratory Study on the Therapeutic Efficacy of Astragaloside IV Against Diabetic Retinopathy Revealed by Network Pharmacology<br>PMID 35814228 | Grace Kim: No saffron | Pranav Potluri: No saffron | – |
| 847. Mogi 2022. Update on Hypertension Research in 2021<br>PMID 35790879 | Grace Kim: No saffron | Pranav Potluri: No saffron | – |
| 848. Miri 2021. Association of ACE I/D and PAI-1 4G/5G polymorphisms with susceptibility to type 2 diabetes mellitus<br>PMID 34900771 | Grace Kim: No saffron | Pranav Potluri: No saffron | – |
| 849. Tonade 2021. Photoreceptor cells and RPE contribute to the development of diabetic retinopathy<br>PMID 33188897 | Grace Kim: No saffron | Pranav Potluri: No saffron | – |

Table S2 continued

| Paper | Reviewer 1 | Reviewer 2 | Additional recorded reviews |
|---|---|---|---|
| 850. Babaei 2022. Exercise training modulates adipokine dysregulations in metabolic syndrome<br>PMID 35782776 | Grace Kim: No saffron | Pranav Potluri: No saffron | – |
| 851. Qiu 2022. Metabolomics and biochemical insights on the regulation of aging-related diabetes by a low-molecular-weight polysaccharide from green microalga Chlorella pyrenoidosa<br>PMID 35774637 | Grace Kim: Not clinical | Pranav Potluri: No saffron | – |
| 852. Roth-Walter 2022. Iron-Deficiency in Atopic Diseases: Innate Immune Priming by Allergens and Siderophores<br>PMID 35769558 | Grace Kim: Not clinical | Pranav Potluri: No saffron | – |
| 853. Xu 2022. Demethylzeylasteral reduces the level of proteinuria in diabetic nephropathy: Screening of network pharmacology and verification by animal experiment<br>PMID 35716119 | Grace Kim: Not clinical | Pranav Potluri: No saffron | – |
| 854. Blevins 2022. The NLRP3 Inflammasome Pathway: A Review of Mechanisms and Inhibitors for the Treatment of Inflammatory Diseases<br>PMID 35754962 | Grace Kim: No saffron | Pranav Potluri: No saffron | – |
| 855. El-Hawary 2021. In vivo antidiabetic potential of standardized Gymnocarpos decandrus Forssk. Extract<br>PMID 34900766 | Grace Kim: No saffron | Pranav Potluri: No saffron | – |
| 856. Zhao 2022. Neuroendocrine-Immune Regulatory Network of Eucommia ulmoides Oliver<br>PMID 35744822 | Grace Kim: No saffron | Pranav Potluri: No saffron | – |
| 857. Hatziagapiou 2022. An In Vitro Study of Saffron Carotenoids: The Effect of Crocin Extracts and Dimethylcrocetin on Cancer Cell Lines<br>PMID 35739971 | Grace Kim: No saffron | Pranav Potluri: Not clinical | – |
| 858. Chouh 2022. Phlorotannins of the Brown Algae Sargassum vulgare from the Mediterranean Sea Coast<br>PMID 35739951 | Grace Kim: No saffron | Pranav Potluri: No saffron | – |
| 859. Chattopadhyay 2022. Effectiveness and Safety of Ayurvedic Medicines in Type 2 Diabetes Mellitus Management: A Systematic Review and Meta-Analysis<br>PMID 35754481 | Grace Kim: Not isolated | Pranav Potluri: No saffron | – |
| 860. de Britto Rosa 2022. Fatty acids composition and in vivo biochemical effects of Aleurites moluccana seed (Candlenut) in obese wistar rats<br>PMID 35676689 | Grace Kim: No saffron | Pranav Potluri: No saffron | – |

Table S2 continued

| Paper | Reviewer 1 | Reviewer 2 | Additional recorded reviews |
|---|---|---|---|
| 861. Prokisch 2022. Functional Yogurt Fortified with Honey Produced by Feeding Bees Natural Plant Extracts for Controlling Human Blood Sugar Level<br>PMID 35684164 | Grace Kim: No saffron | Pranav Potluri: No saffron | – |
| 862. Kellow 2022. The Effect of Dietary Patterns on Clinical Pregnancy and Live Birth Outcomes in Men and Women Receiving Assisted Reproductive Technologies: A Systematic Review and Meta-Analysis<br>PMID 35293975 | Grace Kim: No saffron | Pranav Potluri: No saffron | – |
| 863. Ardakanian 2022. Effect of alpha-mangostin on olanzapine-induced metabolic disorders in rats<br>PMID 35655598 | Grace Kim: No saffron | Pranav Potluri: No saffron | – |
| 864. Bhat 2022. Natural Therapeutics in Aid of Treating Alzheimer's Disease: A Green Gateway Toward Ending Quest for Treating Neurological Disorders<br>PMID 35651632 | Grace Kim: No saffron | Pranav Potluri: No saffron | – |
| 865. Sisay 2022. Glinus lotoides linn. Seed extract as antidiabetic agent: In vitro and in vivo anti-glucolipotoxicity efficacy in Type-II diabetes mellitus<br>PMID 35637658 | Grace Kim: No saffron | Pranav Potluri: No saffron | – |
| 866. Kim 2022. Anti-Obesity Effect of Dyglomera(®) Is Associated with Activation of the AMPK Signaling Pathway in 3T3-L1 Adipocytes and Mice with High-Fat Diet-Induced Obesity<br>PMID 35630765 | Grace Kim: No saffron | Pranav Potluri: No saffron | – |
| 867. Tao 2022. Ethanolic Extract from Pteris wallichiana Alleviates DSS-Induced Intestinal Inflammation and Intestinal Barrier Dysfunction by Inhibiting the TLR4/NF-$\kappa$B Pathway and Regulating Tight Junction Proteins<br>PMID 35630568 | Grace Kim: No saffron | Pranav Potluri: No saffron | – |
| 868. Unnisa 2022. Development of Dapagliflozin Solid Lipid Nanoparticles as a Novel Carrier for Oral Delivery: Statistical Design, Optimization, In-Vitro and In-Vivo Characterization, and Evaluation<br>PMID 35631394 | Grace Kim: No saffron | Pranav Potluri: No saffron | – |
| 869. Kadoglou 2022. A Comprehensive Review of the Cardiovascular Protective Properties of Silibinin/Silymarin: A New Kid on the Block<br>PMID 35631363 | Grace Kim: No saffron | Pranav Potluri: No saffron | – |

Table S2 continued

| Paper | Reviewer 1 | Reviewer 2 | Additional recorded reviews |
|---|---|---|---|
| 870. Liu 2022. A Causal Relationship between Vitamin C Intake with Hyperglycemia and Metabolic Syndrome Risk: A Two-Sample Mendelian Randomization Study PMID 35624721 | Grace Kim: No saffron | Pranav Potluri: No saffron | – |
| 871. Safriani 2022. Using metabolomics to discover the immunomodulator activity of food plants PMID 35647332 | Grace Kim: No saffron | Pranav Potluri: No saffron | – |
| 872. Mavropalias 2022. Exercise medicine for cancer cachexia: targeted exercise to counteract mechanisms and treatment side effects PMID 35088134 | Grace Kim: No saffron | Pranav Potluri: No saffron | – |
| 873. Mohammad 2022. Antioxidant Effects of Curcumin Gel in Experimental Induced Diabetes and Periodontitis in Rats PMID 35592526 | Grace Kim: No saffron | Bryan Anh Nguyen: Not clinical | – |
| 874. Al-Khayri 2022. Flavonoids as Potential Anti-Inflammatory Molecules: A Review PMID 35566252 | Grace Kim: No saffron | Bryan Anh Nguyen: No saffron | – |
| 875. Nasri 2018. Preventive effect of goby fish protein hydrolysates on hyperlipidemia and cardiovascular disease in Wistar rats fed a high-fat/fructose diet PMID 35541829 | Grace Kim: No saffron | Bryan Anh Nguyen: No saffron | – |
| 876. Zarafshan 2022. Type 2 Diabetes Mellitus Provokes Rat Immune Cells Recruitment into the Pulmonary Niche by Up-regulation of Endothelial Adhesion Molecules PMID 35517882 | Grace Kim: No saffron | Bryan Anh Nguyen: No saffron | – |
| 877. Li 2022. Glucagon-Like Peptide 1 Receptor Agonist Improves Renal Tubular Damage in Mice with Diabetic Kidney Disease PMID 35519661 | Grace Kim: No saffron | Bryan Anh Nguyen: No saffron | – |
| 878. Hausman-Cohen 2022. Genomics as a Clinical Decision Support Tool for Identifying and Addressing Modifiable Causes of Cognitive Decline and Improving Outcomes: Proof of Concept Support for This Personalized Medicine Strategy PMID 35517054 | Grace Kim: No saffron | Bryan Anh Nguyen: No saffron | – |
| 879. Fernández-López 2020. New insights into red plant pigments: more than just natural colorants PMID 35516216 | Grace Kim: No saffron | Bryan Anh Nguyen: No saffron | – |
| 880. Lin 2019. Gut microbiota: a new angle for traditional herbal medicine research PMID 35519900 | Grace Kim: No saffron | Bryan Anh Nguyen: No saffron | – |

Table S2 continued

| Paper | Reviewer 1 | Reviewer 2 | Additional recorded reviews |
|---|---|---|---|
| 881. Leitner 2022. Insulin and cancer: a tangled web PMID 35244142 | Grace Kim: No saffron | Bryan Anh Nguyen: No saffron | – |
| 882. Zhou 2022. The Effect of Chinese Medicinal Formulas on Biomarkers of Oxidative Stress in STZ-Induced Diabetic Kidney Disease Rats: A Meta-Analysis and Systematic Review PMID 35492300 | Grace Kim: No saffron | Bryan Anh Nguyen: No saffron | – |
| 883. Asadi 2022. Association between Dietary Inflammatory Index Scores and Diabetes Sensorimotor Polyneuropathy in Patients with Type 2 Diabetes Mellitus: A Case-Control Study PMID 35497930 | Grace Kim: No saffron | Bryan Anh Nguyen: No saffron | – |
| 884. Lindsay 2021. Maternal mindful eating as a target for improving metabolic outcomes in pregnant women with obesity PMID 34994169 | Grace Kim: No saffron | Bryan Anh Nguyen: No saffron | – |
| 885. Author not recorded 2021. Prize Presentations: ACHARI PRIZE PMC9045589 | Grace Kim: No saffron | Bryan Anh Nguyen: No saffron | – |
| 886. Samaha 2022. Indapamide Increases IRS1 Expression and Modifies Adiponectin/NLRP3/PPAR$\gamma$ Crosstalk in Type 2 Diabetic Rats PMID 35453376 | Grace Kim: No saffron | Bryan Anh Nguyen: No saffron | – |
| 887. Sonali 2022. Mechanistic Insights into the Link between Gut Dysbiosis and Major Depression: An Extensive Review PMID 35456041 | Grace Kim: No saffron | Bryan Anh Nguyen: No saffron | – |
| 888. Clemente-Suárez 2022. Nutritional and Exercise Interventions in Cancer-Related Cachexia: An Extensive Narrative Review PMID 35457471 | Grace Kim: No saffron | Bryan Anh Nguyen: No saffron | – |
| 889. Wolosowicz 2022. Recent Advances in the Treatment of Insulin Resistance Targeting Molecular and Metabolic Pathways: Fighting a Losing Battle? PMID 35454311 | Grace Kim: No saffron | Bryan Anh Nguyen: No saffron | – |
| 890. Deepika 2022. Health Benefits of Quercetin in Age-Related Diseases PMID 35458696 | Grace Kim: No saffron | Bryan Anh Nguyen: No saffron | – |
| 891. Bari 2022. Effects of Artemisia macrocephala Jacquem on Memory Deficits and Brain Oxidative Stress in Streptozotocin-Induced Diabetic Mice PMID 35458597 | Grace Kim: No saffron | Bryan Anh Nguyen: No saffron | – |

Table S2 continued

| Paper | Reviewer 1 | Reviewer 2 | Additional recorded reviews |
|---|---|---|---|
| 892. Inam 2022. Development and Characterization of Eudragit(®) EPO-Based Solid Dispersion of Rosuvastatin Calcium to Foresee the Impact on Solubility, Dissolution and Antihyperlipidemic Activity<br>PMID 35455489 | Grace Kim: No saffron | Bryan Anh Nguyen: No saffron | – |
| 893. Chen 2022. Glucocorticoid-induced loss of beneficial gut bacterial extracellular vesicles is associated with the pathogenesis of osteonecrosis<br>PMID 35417243 | Grace Kim: No saffron | Bryan Anh Nguyen: No saffron | – |
| 894. Tomas 2022. The direct and indirect effects of bioactive compounds against coronavirus<br>PMID 35462942 | Grace Kim: No saffron | Bryan Anh Nguyen: No saffron | – |
| 895. Gidwani 2022. Indian spices: past, present and future challenges as the engine for bio-enhancement of drugs: impact of COVID-19<br>PMID 35043421 | Grace Kim: No saffron | Bryan Anh Nguyen: No saffron | – |
| 896. Ng 2022. Operational definition of complementary, alternative, and integrative medicine derived from a systematic search<br>PMID 35413882 | Grace Kim: No saffron | Bryan Anh Nguyen: No saffron | – |
| 897. Jiménez-Sánchez 2022. Therapeutic Properties and Use of Extra Virgin Olive Oil in Clinical Nutrition: A Narrative Review and Literature Update<br>PMID 35406067 | Grace Kim: No saffron | Bryan Anh Nguyen: No saffron | – |
| 898. Kozłowska 2022. Targeting Cardiovascular Diseases by Flavonols: An Update<br>PMID 35406050 | Grace Kim: No saffron | Bryan Anh Nguyen: No saffron | – |
| 899. Pogány 2022. Redox and Hormonal Changes in the Transcriptome of Grape (Vitis vinifera) Berries during Natural Noble Rot Development<br>PMID 35406844 | Grace Kim: No saffron | Bryan Anh Nguyen: No saffron | – |
| 900. Ren 2022. Influence of gardenia yellow on in vitro slow starch digestion and its action mechanism<br>PMID 35424638 | Grace Kim: No saffron | Bryan Anh Nguyen: No saffron | – |
| 901. Allegrini 2022. Curcumin in Retinal Diseases: A Comprehensive Review from Bench to Bedside<br>PMID 35408920 | Grace Kim: No saffron | Bryan Anh Nguyen: No saffron | – |
| 902. Leh 2022. Lycopene: A Potent Antioxidant for the Amelioration of Type II Diabetes Mellitus<br>PMID 35408734 | Grace Kim: Not isolated | Bryan Anh Nguyen: No saffron | – |

Table S2 continued

| Paper | Reviewer 1 | Reviewer 2 | Additional recorded reviews |
|---|---|---|---|
| 903. Fuloria 2022. A Comprehensive Review on the Therapeutic Potential of Curcuma longa Linn. in Relation to its Major Active Constituent Curcumin<br>PMID 35401176 | Grace Kim: No saffron | Bryan Anh Nguyen: No saffron | – |
| 904. Laka 2022. Cholesterol-Lowering Phytochemicals: Targeting the Mevalonate Pathway for Anticancer Interventions<br>PMID 35391801 | Grace Kim: No saffron | Bryan Anh Nguyen: No saffron | – |
| 905. Olivares-González 2022. NUTRARET: Effect of 2-Year Nutraceutical Supplementation on Redox Status and Visual Function of Patients With Retinitis Pigmentosa: A Randomized, Double-Blind, Placebo-Controlled Trial<br>PMID 35387197 | Grace Kim: No saffron | Bryan Anh Nguyen: No saffron | – |
| 906. Arcusa 2022. Potential Role of Ginger (Zingiber officinale Roscoe) in the Prevention of Neurodegenerative Diseases<br>PMID 35369082 | Grace Kim: No saffron | Bryan Anh Nguyen: No saffron | – |
| 907. Yaribeygi 2022. Molecular mechanisms linking stress and insulin resistance<br>PMID 35368460 | Grace Kim: No saffron | Bryan Anh Nguyen: No saffron | – |
| 908. Attlee 2022. Higher Dietary Inflammatory Index Scores Are Associated With Stress and Anxiety in Dormitory-Residing Female University Students in the United Arab Emirates<br>PMID 35360682 | Grace Kim: No saffron | Bryan Anh Nguyen: No saffron | – |
| 909. Li 2022. Recent advance in treatment of atherosclerosis: Key targets and plaque-positioned delivery strategies<br>PMID 35356091 | Grace Kim: No saffron | Bryan Anh Nguyen: No saffron | – |
| 910. Nethengwe 2022. Ameliorative effects of Anchomanes difformis aqueous extract against oxidative stress in the testes and epididymis of streptozotocin-induced diabetic male Wistar rats<br>PMID 35355957 | Grace Kim: No saffron | Bryan Anh Nguyen: No saffron | – |
| 911. Younis 2022. Metabolomics-based profiling of 4 avocado varieties using HPLC-MS/MS and GC/MS and evaluation of their antidiabetic activity<br>PMID 35322072 | Grace Kim: No saffron | Bryan Anh Nguyen: No saffron | – |
| 912. Pollini 2022. Ultrasound-Assisted Extraction and Characterization of Polyphenols from Apple Pomace, Functional Ingredients for Beef Burger Fortification<br>PMID 35335297 | Grace Kim: No saffron | Bryan Anh Nguyen: No saffron | – |

Table S2 continued

| Paper | Reviewer 1 | Reviewer 2 | Additional recorded reviews |
|---|---|---|---|
| 913. Kamal 2022. Genesis and Mechanism of Some Cancer Types and an Overview on the Role of Diet and Nutrition in Cancer Prevention<br>PMID 35335158 | Grace Kim: No saffron | Bryan Anh Nguyen: No saffron | – |
| 914. Tripathy 2021. Trends in diabetes care with special emphasis to medicinal plants: Advancement and treatment<br>PMID 35342487 | Grace Kim: Not isolated | Bryan Anh Nguyen: No saffron | – |
| 915. Mokhtary 2022. Deletion allele of Apo B gene is associated with higher inflammation, oxidative stress and dyslipidemia in obese type 2 diabetic patients: an analytical cross-sectional study<br>PMID 35317787 | Grace Kim: No saffron | Bryan Anh Nguyen: No saffron | – |
| 916. Yahyazadeh 2021. The effect of Elettaria cardamomum (cardamom) on the metabolic syndrome: Narrative review<br>PMID 35317114 | Grace Kim: No saffron | Bryan Anh Nguyen: No saffron | – |
| 917. Silva 2022. Bioactive Compounds from Cardoon as Health Promoters in Metabolic Disorders<br>PMID 35159487 | Grace Kim: No saffron | Bryan Anh Nguyen: No saffron | – |
| 918. Noor 2022. Ferulic acid loaded pickering emulsions stabilized by resistant starch nanoparticles using ultrasonication: Characterization, in vitro release and nutraceutical potential<br>PMID 35279632 | Grace Kim: No saffron | Bryan Anh Nguyen: No saffron | – |
| 919. Paul 2022. The Role of Medicinal and Aromatic Plants against Obesity and Arthritis: A Review<br>PMID 35267958 | Grace Kim: No saffron | Bryan Anh Nguyen: No saffron | – |
| 920. Ghorbanalizadeh 2021. Plant diversity of Hyrcanian relict forests: An annotated checklist, chorology and threat categories of endemic and near endemic vascular plant species<br>PMID 35281126 | Grace Kim: No saffron | Bryan Anh Nguyen: No saffron | – |
| 921. Rad 2022. Antioxidant effects of astaxanthin and metformin combined therapy in type 2 diabetes mellitus patients: a randomized double-blind controlled clinical trial<br>PMID 35280834 | Grace Kim: No saffron | Bryan Anh Nguyen: No saffron | – |
| 922. Huang 2022. Endothelial Function and Postprandial Glucose Control in Response to Test-Meals Containing Herbs and Spices in Adults With Overweight/Obesity<br>PMID 35273988 | Grace Kim: No saffron | Bryan Anh Nguyen: No saffron | – |
| 923. Shu 2022. Associations of Dietary Inflammatory Index With Prediabetes and Insulin Resistance<br>PMID 35250879 | Grace Kim: No saffron | Bryan Anh Nguyen: No saffron | – |

Table S2 continued

| Paper | Reviewer 1 | Reviewer 2 | Additional recorded reviews |
|---|---|---|---|
| 924. Sabir 2022. Reduction of Hepatic Steatosis, Oxidative Stress, Inflammation, Ballooning and Insulin Resistance After Therapy with Safranal in NAFLD Animal Model: A New Approach PMID 35241921 | Grace Kim: Not clinical | Michele Schaffer: Not clinical | – |
| 925. Qin 2022. Instant Dark Tea Alleviates Hyperlipidaemia in High-Fat Diet-Fed Rat: From Molecular Evidence to Redox Balance and Beyond PMID 35223953 | Grace Kim: No saffron | Michele Schaffer: Not clinical | – |
| 926. Zhao 2022. The Potential to Fight Obesity with Adipogenesis Modulating Compounds PMID 35216415 | Grace Kim: No saffron | Michele Schaffer: Not clinical | – |
| 927. Morrison 2022. Heat-Inactivated Akkermansia muciniphila Improves Gut Permeability but Does Not Prevent Development of Non-Alcoholic Steatohepatitis in Diet-Induced Obese Ldlr-/-.Leiden Mice PMID 35216439 | Grace Kim: No saffron | Michele Schaffer: No saffron | – |
| 928. Azemi 2022. Phytochemical Contents and Pharmacological Potential of Parkia speciosa Hassk. for Diabetic Vasculopathy: A Review PMID 35204313 | Grace Kim: No saffron | Michele Schaffer: Not clinical | – |
| 929. Lamanna-Rama 2022. An Update on the Exploratory Use of Curcumin in Neuropsychiatric Disorders PMID 35204235 | Grace Kim: No saffron | Michele Schaffer: Not clinical | – |
| 930. Donati Zeppa 2022. Nutraceuticals and Physical Activity as Antidepressants: The Central Role of the Gut Microbiota PMID 35204119 | Grace Kim: No saffron | Michele Schaffer: Not clinical | – |
| 931. Pritam 2022. Antioxidants in Alzheimer's Disease: Current Therapeutic Significance and Future Prospects PMID 35205079 | Grace Kim: No saffron | Michele Schaffer: Not clinical | – |
| 932. Szot 2022. Can Nutrients and Dietary Supplements Potentially Improve Cognitive Performance Also in Esports? PMID 35206801 | Grace Kim: No saffron | Michele Schaffer: Not clinical | – |
| 933. Butnariu 2022. The Pharmacological Activities of Crocus sativus L.: A Review Based on the Mechanisms and Therapeutic Opportunities of its Phytoconstituents PMID 35198096 | Grace Kim: Not clinical | Michele Schaffer: Not clinical | – |
| 934. Wang 2022. Oral Administration of Bacterial $\beta$ Cell Expansion Factor A (BefA) Alleviates Diabetes in Mice with Type 1 and Type 2 Diabetes PMID 35186190 | Grace Kim: No saffron | Michele Schaffer: Not clinical | – |

Table S2 continued

| Paper | Reviewer 1 | Reviewer 2 | Additional recorded reviews |
|---|---|---|---|
| 935. Mishu 2021. Interventions for preventing type 2 diabetes in adults with mental disorders in low- and middle-income countries<br>PMID 33591592 | Grace Kim: Not isolated | Michele Schaffer: Not clinical | – |
| 936. El Midaoui 2022. Saffron (Crocus sativus L.): A Source of Nutrients for Health and for the Treatment of Neuropsychiatric and Age-Related Diseases<br>PMID 35276955 | Grace Kim: Not clinical | Michele Schaffer: Not clinical | – |
| 937. Valls 2022. Effects of an Optimized Aged Garlic Extract on Cardiovascular Disease Risk Factors in Moderate Hypercholesterolemic Subjects: A Randomized, Crossover, Double-Blind, Sustainedand Controlled Study<br>PMID 35276764 | Grace Kim: No saffron | Michele Schaffer: Not isolated | – |
| 938. Zhang 2022. Crocin ameliorates atherosclerosis by promoting the reverse cholesterol transport and inhibiting the foam cell formation via regulating PPAR$\gamma$/LXR-$\alpha$<br>PMID 34978526 | Grace Kim: Not clinical | Michele Schaffer: Not clinical | – |
| 939. Wan 2022. Quercetin attenuates the proliferation, inflammation, and oxidative stress of high glucose-induced human mesangial cells by regulating the miR-485-5p/YAP1 pathway<br>PMID 35129398 | Grace Kim: No saffron | Michele Schaffer: Not isolated | – |
| 940. Gervasi 2022. Biotechnological Applications and Health-Promoting Properties of Flavonols: An Updated View<br>PMID 35163632 | Grace Kim: No saffron | Michele Schaffer: Not isolated | – |
| 941. Wei 2022. Preliminary evidence for the presence of multiple forms of cell death in diabetes cardiomyopathy<br>PMID 35127369 | Grace Kim: No saffron | Michele Schaffer: Not isolated | – |
| 942. Anusiya 2021. A review of the therapeutic and biological effects of edible and wild mushrooms<br>PMID 34738876 | Grace Kim: No saffron | Michele Schaffer: No saffron | – |
| 943. Jin 2021. Ibrutinib ameliorates cerebral ischemia/reperfusion injury through autophagy activation and PI3K/Akt/mTOR signaling pathway in diabetic mice<br>PMID 34605340 | Grace Kim: No saffron | Michele Schaffer: Not isolated | – |
| 944. Xu 2022. Hypoglycemic Effect of Electroacupuncture at ST25 Through Neural Regulation of the Pancreatic Intrinsic Nervous System<br>PMID 34757591 | Grace Kim: No saffron | Michele Schaffer: Not isolated | – |

Table S2 continued

| Paper | Reviewer 1 | Reviewer 2 | Additional recorded reviews |
|---|---|---|---|
| 945. Liu 2022. Pharmacological treatment with FGF21 strongly improves plasma cholesterol metabolism to reduce atherosclerosis<br>PMID 33693480 | Grace Kim: No saffron | Michele Schaffer: No saffron | – |
| 946. Liu 2022. Emerging Theranostic Nanomaterials in Diabetes and Its Complications<br>PMID 34825525 | Grace Kim: No saffron | Michele Schaffer: Not isolated | – |
| 947. Guo 2022. Crocetin: A Systematic Review<br>PMID 35095483 | Grace Kim: No saffron | Michele Schaffer: Not clinical | – |
| 948. Esmaeelpanah 2021. Green tea and metabolic syndrome: A 10-year research update review<br>PMID 35083002 | Grace Kim: No saffron | Bryan Anh Nguyen: No saffron | – |
| 949. Galanakis 2022. Sustainable Applications for the Valorization of Cereal Processing By-Products<br>PMID 35053973 | Grace Kim: No saffron | Pranav Potluri: No saffron | – |
| 950. Jazayeri 2021. Up-regulation of Arl4a gene expression by broccoli aqueous extract is associated with improved spermatogenesis in mouse testes<br>PMID 34936255 | Grace Kim: No saffron | Pranav Potluri: No saffron | – |
| 951. Gao 2022. Maslinic acid activates renal AMPK/SIRT1 signaling pathway and protects against diabetic nephropathy in mice<br>PMID 35042497 | Grace Kim: No saffron | Pranav Potluri: No saffron | – |
| 952. Heidary Moghaddam 2022. Natural AMPK Activators in Cardiovascular Disease Prevention<br>PMID 35046800 | Grace Kim: No saffron | Pranav Potluri: No saffron | – |
| 953. Yao 2021. The Roles of Carotenoid Consumption and Bioavailability in Cardiovascular Health<br>PMID 34943081 | Grace Kim: Not clinical | Pranav Potluri: No saffron | – |
| 954. Chandrasekaran 2022. Role of Curcumin in Retinal Diseases-A review<br>PMID 35015114 | Grace Kim: No saffron | Pranav Potluri: No saffron | – |
| 955. Zolkeflee 2022. In Vitro Anti-Diabetic Activities and UHPLC-ESI-MS/MS Profile of Muntingia calabura Leaves Extract<br>PMID 35011523 | Grace Kim: No saffron | Pranav Potluri: No saffron | – |
| 956. D'Alessandro 2022. Interactions between Food and Drugs, and Nutritional Status in Renal Patients: A Narrative Review<br>PMID 35011087 | Grace Kim: No saffron | Pranav Potluri: No saffron | – |

Table S2 continued

| Paper | Reviewer 1 | Reviewer 2 | Additional recorded reviews |
|---|---|---|---|
| 957. Alkhatib 2021. The Role of Bioactive Compounds from Dietary Spices in the Management of Metabolic Syndrome: An Overview<br>PMID 35011050 | Grace Kim: Not clinical | Pranav Potluri: No saffron | – |
| 958. Alemán-González-Duhart 2021. In Vivo and Ex Vivo Evaluation of 1,3-Thiazolidine-2,4-Dione Derivatives as Euglycemic Agents<br>PMID 35003235 | Grace Kim: No saffron | Pranav Potluri: No saffron | – |
| 959. Xu 2021. Mitochondrial Targeting Therapeutics: Promising Role of Natural Products in Non-alcoholic Fatty Liver Disease<br>PMID 35002729 | Grace Kim: No saffron | Pranav Potluri: No saffron | – |
| 960. Jiang 2021. Spotlight on NLRP3 Inflammasome: Role in Pathogenesis and Therapies of Atherosclerosis<br>PMID 34992411 | Grace Kim: No saffron | Pranav Potluri: No saffron | – |
| 961. Author not recorded 2021. 2021 ACVIM Forum Research Abstract Program<br>PMID 34351021 | Grace Kim: No saffron | Pranav Potluri: No saffron | – |
| 962. von Lintig 2021. Molecular components affecting ocular carotenoid and retinoid homeostasis<br>PMID 32339666 | Grace Kim: No saffron | Pranav Potluri: No saffron | – |
| 963. Berta 2021. A Review on Plant-Mediated Synthesis of Bimetallic Nanoparticles, Characterisation and Their Biological Applications<br>PMID 34947271 | Grace Kim: Not clinical | Pranav Potluri: No saffron | – |
| 964. Jakše 2021. Placing a Well-Designed Vegan Diet for Slovenes<br>PMID 34960098 | Grace Kim: No saffron | Pranav Potluri: No saffron | – |
| 965. Salis 2021. Old Is Gold': How Traditional Indian Dietary Practices Can Support Pediatric Diabetes Management<br>PMID 34959978 | Grace Kim: No saffron | Pranav Potluri: No saffron | – |
| 966. Gart 2021. Butyrate Protects against Diet-Induced NASH and Liver Fibrosis and Suppresses Specific Non-Canonical TGF-$\beta$ Signaling Pathways in Human Hepatic Stellate Cells<br>PMID 34944770 | Grace Kim: No saffron | Pranav Potluri: No saffron | – |
| 967. Majka 2021. Impact of the Main Cardiovascular Risk Factors on Plasma Extracellular Vesicles and Their Influence on the Heart's Vulnerability to Ischemia-Reperfusion Injury<br>PMID 34943838 | Grace Kim: No saffron | Pranav Potluri: No saffron | – |
| 968. Wilburn 2004. The natural treatment of hypertension<br>PMID 15133406 | Grace Kim: No saffron | Pranav Potluri: No saffron | – |

Table S2 continued

| Paper | Reviewer 1 | Reviewer 2 | Additional recorded reviews |
|---|---|---|---|
| 969. Zhou 2021. Astragalus-Saffron-Rhubarb Mixture Delays the Progress of Diabetic Nephropathy in db/db Mice PMID 34916815 | Grace Kim: Not clinical | Pranav Potluri: Not isolated | – |
| 970. Tang 2021. The effect and mechanism of Jiao-tai-wan in the treatment of diabetes mellitus with depression based on network pharmacology and experimental analysis PMID 34875999 | Grace Kim: No saffron | Pranav Potluri: No saffron | – |
| 971. Arnone 2021. Long-Term Overconsumption of Fat and Sugar Causes a Partially Reversible Pre-inflammatory Bowel Disease State PMID 34869528 | Grace Kim: No saffron | Pranav Potluri: No saffron | – |
| 972. Anaeigoudari 2021. Effects of Nigella sativa, Camellia sinensis, and Allium sativum as Food Additives on Metabolic Disorders, a Literature Review PMID 34867384 | Grace Kim: No saffron | Pranav Potluri: No saffron | – |
| 973. Andreadou 2020. Hyperlipidaemia and cardioprotection: Animal models for translational studies PMID 31769007 | Grace Kim: No saffron | Pranav Potluri: No saffron | – |
| 974. van Bilsen 2021. Mechanism-Based Biomarker Prediction for Low-Grade Inflammation in Liver and Adipose Tissue PMID 34858196 | Grace Kim: No saffron | Pranav Potluri: No saffron | – |
| 975. Bai 2021. Novel Antioxidants and $\alpha$-Glycosidase and Protein Tyrosine Phosphatase 1B Inhibitors from an Endophytic Fungus Penicillium brefeldianum F4a PMID 34829202 | Grace Kim: No saffron | Pranav Potluri: No saffron | – |
| 976. Cicalău 2021. Anti-Inflammatory and Antioxidant Properties of Carvacrol and Magnolol, in Periodontal Disease and Diabetes Mellitus PMID 34833990 | Grace Kim: No saffron | Pranav Potluri: No saffron | – |
| 977. Chung 2021. AMPK Activity: A Primary Target for Diabetes Prevention with Therapeutic Phytochemicals PMID 34836306 | Grace Kim: No saffron | Pranav Potluri: No saffron | – |
| 978. Park 2021. Interactions between Polygenic Risk Scores, Dietary Pattern, and Menarche Age with the Obesity Risk in a Large Hospital-Based Cohort PMID 34836030 | Grace Kim: No saffron | Pranav Potluri: No saffron | – |
| 979. Contreras-Zentella 2021. Possible Gender Influence in the Mechanisms Underlying the Oxidative Stress, Inflammatory Response, and the Metabolic Alterations in Patients with Obesity and/or Type 2 Diabetes PMID 34829598 | Grace Kim: No saffron | Pranav Potluri: No saffron | – |

Table S2 continued

| Paper | Reviewer 1 | Reviewer 2 | Additional recorded reviews |
|---|---|---|---|
| 980. Abedini 2021. Evaluation of the in vivo and in vitro safety profile of Cuscuta epithymum ethanolic extract PMID 34804901 | Grace Kim: No saffron | Pranav Potluri: No saffron | – |
| 981. Badavi 2020. Crocin attenuates oxidative stress and inflammation in myocardial infarction induced by isoprenaline via PPAR$\gamma$ activation in diabetic rats PMID 33553037 | Grace Kim: Not clinical | Pranav Potluri: No saffron | – |
| 982. Yamada 2021. Involvement of Ghrelin Dynamics in Stress-Induced Eating Disorder: Effects of Sex and Aging PMID 34769125 | Grace Kim: No saffron | Pranav Potluri: No saffron | – |
| 983. Zhang 2021. Integrative Analysis of Gut Microbiota and Fecal Metabolites in Rats after Prednisone Treatment PMID 34756061 | Grace Kim: No saffron | Pranav Potluri: No saffron | – |
| 984. Wei 2021. Traditional Uses, Chemistry, Pharmacology, Toxicology and Quality Control of Alhagi sparsifolia Shap.: A Review PMID 34721046 | Grace Kim: No saffron | Pranav Potluri: No saffron | – |
| 985. Tousian 2021. In search of elixir: Pharmacological agents against stem cell senescence PMID 34712416 | Grace Kim: No saffron | Pranav Potluri: No saffron | – |
| 986. Mulette 2021. Pulmonary cavitations with increased 18F-FDG uptake revealing a thoracic endometriosis: A case report PMID 34678890 | Grace Kim: No saffron | Pranav Potluri: No saffron | – |
| 987. Abbasi 2021. Favorable effects of Anethum graveolens on liver oxidative stress and cholesterol 7 alpha-hydroxylase levels in non-alcoholic fatty liver disease (NAFLD) rat models PMID 34704009 | Grace Kim: No saffron | Pranav Potluri: No saffron | – |
| 988. Yang 2021. Protective Effects of Ulva lactuca Polysaccharide Extract on Oxidative Stress and Kidney Injury Induced by D-Galactose in Mice PMID 34677438 | Grace Kim: No saffron | Pranav Potluri: No saffron | – |
| 989. Sahukari 2021. Phytochemical Profile, Free Radical Scavenging and Anti-Inflammatory Properties of Acalypha Indica Root Extract: Evidence from In Vitro and In Vivo Studies PMID 34684831 | Grace Kim: No saffron | Pranav Potluri: No saffron | – |
| 990. Nakhaee 2021. The effects of quercetin on seizure, inflammation parameters and oxidative stress in acute on chronic tramadol intoxication PMID 34666816 | Grace Kim: No saffron | Pranav Potluri: No saffron | – |

Table S2 continued

| Paper | Reviewer 1 | Reviewer 2 | Additional recorded reviews |
|---|---|---|---|
| 991. Rostoker 2021. Analysis of liver iron concentration in an elderly female undergoing hemodialysis with calcific uremic arteriolopathy does not support the role of iron overload in calciphylaxis: lesson for the clinical nephrologist<br>PMID 33484427 | Grace Kim: No saffron | Pranav Potluri: No saffron | – |
| 992. Yaribeygi 2021. Pathophysiology of Physical Inactivity-Dependent Insulin Resistance: A Theoretical Mechanistic Review Emphasizing Clinical Evidence<br>PMID 34660812 | Grace Kim: No saffron | Pranav Potluri: No saffron | – |
| 993. Morvaridzadeh 2021. Effect of saffron supplementation on oxidative stress parameters: A systematic review and meta-analysis of randomized placebo-controlled trials<br>PMID 34646548 | Grace Kim: Not clinical | Pranav Potluri: Not clinical | – |
| 994. Asdaq 2021. The Potential Benefits of Using Garlic Oil and Its Active Constituent, Dially Disulphide, in Combination With Carvedilol in Ameliorating Isoprenaline-Induced Cardiac Damage in Rats<br>PMID 34646139 | Grace Kim: No saffron | Pranav Potluri: No saffron | – |
| 995. Hasani 2021. Beneficial effects of Se/Zn co-supplementation on body weight and adipose tissue inflammation in high-fat diet-induced obese rats<br>PMID 34631042 | Grace Kim: No saffron | Pranav Potluri: No saffron | – |
| 996. Mirazi 2020. Attenuating properties of Rubus fruticosus L. on oxidative damage and inflammatory response following streptozotocin-induced diabetes in the male Wistar rats<br>PMID 33520837 | Grace Kim: No saffron | Pranav Potluri: No saffron | – |
| 997. Mohammadi 2021. Effect of Total Suspended Particulate Matter in the Air on Inflammation Factors and Apoptotic Markers in Diabetic Rats: The Protective Effect of Insulin and Crocin<br>PMID 34604423 | Grace Kim: Not clinical | Pranav Potluri: No saffron | – |
| 998. Robertson 2020. Pine bark (Pinus spp.) extract for treating chronic disorders<br>PMID 32990945 | Grace Kim: No saffron | Pranav Potluri: No saffron | – |
| 999. Namgyal 2021. Aberrant Lighting Causes Anxiety-like Behavior in Mice but Curcumin Ameliorates the Symptoms<br>PMID 34573555 | Grace Kim: No saffron | Pranav Potluri: No saffron | – |
| 1000. Tamagno 2021. Oxidative Stress and Beta Amyloid in Alzheimer's Disease. Which Comes First: The Chicken or the Egg?<br>PMID 34573112 | Grace Kim: No saffron | Pranav Potluri: No saffron | – |

Table S2 continued

| Paper | Reviewer 1 | Reviewer 2 | Additional recorded reviews |
|---|---|---|---|
| 1001. Sauvé 2021. Glycomacropeptide for Management of Insulin Resistance and Liver Metabolic Perturbations<br>PMID 34572325 | Grace Kim: No saffron | Pranav Potluri: No saffron | – |
| 1002. Kim 2021. An Updated Comprehensive Review on Vitamin A and Carotenoids in Breast Cancer: Mechanisms, Genetics, Assessment, Current Evidence, and Future Clinical Implications<br>PMID 34579037 | Grace Kim: No saffron | Pranav Potluri: No saffron | – |
| 1003. Lecorguillé 2021. Maternal Dietary Quality and Dietary Inflammation Associations with Offspring Growth, Placental Development, and DNA Methylation<br>PMID 34579008 | Grace Kim: No saffron | Pranav Potluri: No saffron | – |
| 1004. Klonizakis 2021. Assessing the Physiological Effects of Traditional Regional Diets Targeting the Prevention of Cardiovascular Disease: A Systematic Review of Randomized Controlled Trials Implementing Mediterranean, New Nordic, Japanese, Atlantic, Persian and Mexican Dietary Interventions<br>PMID 34578911 | Grace Kim: No saffron | Pranav Potluri: No saffron | – |
| 1005. Dammak 2021. From Oxidative Stress to Inflammation in the Posterior Ocular Diseases: Diagnosis and Treatment<br>PMID 34575451 | Grace Kim: No saffron | Pranav Potluri: No saffron | – |
| 1006. Beddiar 2021. Chemical Composition, Anti-Quorum Sensing, Enzyme Inhibitory, and Antioxidant Properties of Phenolic Extracts of Clinopodium nepeta L. Kuntze<br>PMID 34579487 | Grace Kim: No saffron | Pranav Potluri: No saffron | – |
| 1007. Bahramsoltani 2021. Bioactive Foods and Medicinal Plants for Cardiovascular Complications of Type II Diabetes: Current Clinical Evidence and Future Perspectives<br>PMID 34567218 | Grace Kim: Not clinical | Pranav Potluri: No saffron | – |
| 1008. Song 2021. Association of Polygenetic Risk Scores Related to Immunity and Inflammation with Hyperthyroidism Risk and Interactions between the Polygenetic Scores and Dietary Factors in a Large Cohort<br>PMID 34567510 | Grace Kim: No saffron | Pranav Potluri: No saffron | – |
| 1009. Dong 2021. Exploration of the profile-effect relationship of Siraitia grosvenorii aqueous extracts related to their laxative effect on the basis of gray correlation analysis<br>PMID 34544411 | Grace Kim: No saffron | Pranav Potluri: No saffron | – |
| 1010. Taheri 2021. Cyperus spp.: A Review on Phytochemical Composition, Biological Activity, and Health-Promoting Effects<br>PMID 34539969 | Grace Kim: No saffron | Pranav Potluri: No saffron | – |

Table S2 continued

| Paper | Reviewer 1 | Reviewer 2 | Additional recorded reviews |
|---|---|---|---|
| 1011. Aminifard 2021. The effects of ginseng on the metabolic syndrome: An updated review<br>PMID 34532035 | Grace Kim: No saffron | Pranav Potluri: No saffron | – |
| 1012. Qiu 2021. Update on the Relationship Between Depression and Neuroendocrine Metabolism<br>PMID 34531719 | Grace Kim: No saffron | Pranav Potluri: No saffron | – |
| 1013. Zhang 2021. Protective Effect of Yi Shen Pai Du Formula against Diabetic Kidney Injury via Inhibition of Oxidative Stress, Inflammation, and Epithelial-to-Mesenchymal Transition in db/db Mice<br>PMID 34504642 | Grace Kim: No saffron | Pranav Potluri: No saffron | – |
| 1014. Samarghandian 2020. Toxicity Evaluation of the Subacute Diazinon in Aged Male Rats: Hematological Aspects<br>PMID 32133967 | Grace Kim: No saffron | Pranav Potluri: No saffron | – |
| 1015. Zhang 2021. Efficacy and Safety of Curcumin Supplement on Improvement of Insulin Resistance in People with Type 2 Diabetes Mellitus: A Systematic Review and Meta-Analysis of Randomized Controlled Trials<br>PMID 34484389 | Grace Kim: No saffron | Pranav Potluri: No saffron | – |
| 1016. Yaribeygi 2021. Renoprotective Effects of Incretin-Based Therapy in Diabetes Mellitus<br>PMID 34471642 | Grace Kim: No saffron | Pranav Potluri: No saffron | – |
| 1017. Adalat 2019. Antidepressant Effects of a Persian Medicine Remedy on Multiple Sclerosis Patients: A Double-Blinded Randomized Clinical Trial<br>PMID 34466472 | Grace Kim: Not clinical | Pranav Potluri: No outcome | – |
| 1018. Park 2021. Association between Polygenetic Risk Scores of Low Immunity and Interactions between These Scores and Moderate Fat Intake in a Large Cohort<br>PMID 34445011 | Grace Kim: No saffron | Pranav Potluri: No saffron | – |
| 1019. Gart 2021. Krill Oil Treatment Increases Distinct PUFAs and Oxylipins in Adipose Tissue and Liver and Attenuates Obesity-Associated Inflammation via Direct and Indirect Mechanisms<br>PMID 34444996 | Grace Kim: No saffron | Pranav Potluri: No saffron | – |
| 1020. Setayesh 2021. The Effect of Saffron Supplementation on Blood Pressure in Adults: A Systematic Review and Dose-Response Meta-Analysis of Randomized Controlled Trials<br>PMID 34444896 | Grace Kim: Not clinical | Pranav Potluri: Not clinical | – |

Table S2 continued

| Paper | Reviewer 1 | Reviewer 2 | Additional recorded reviews |
|---|---|---|---|
| 1021. Wang 2021. Dietary Inflammatory Index and Cardiometabolic Risk in Ecuadorian Women<br>PMID 34444800 | Grace Kim: No saffron | Pranav Potluri: No saffron | – |
| 1022. Andjić 2021. Formulation and Evaluation of Helichrysum italicum Essential Oil-Based Topical Formulations for Wound Healing in Diabetic Rats<br>PMID 34451910 | Grace Kim: No saffron | Pranav Potluri: No saffron | – |
| 1023. Kim 2021. Anti-Obesity Effects of Morus alba L. and Aronia melanocarpa in a High-Fat Diet-Induced Obese C57BL/6J Mouse Model<br>PMID 34441691 | Grace Kim: No saffron | Pranav Potluri: No saffron | – |
| 1024. Kapar 2020. The effects of curcumin and Lactobacillus acidophilus on certain hormones and insulin resistance in rats with metabolic syndrome<br>PMID 33553015 | Grace Kim: No saffron | Pranav Potluri: No saffron | – |
| 1025. Karigidi 2020. Effect of Curculigo pilosa supplemented diet on blood sugar, lipid metabolism, hepatic oxidative stress and carbohydrate metabolism enzymes in streptozotocin-induced diabetic rats<br>PMID 33520833 | Grace Kim: No saffron | Pranav Potluri: No saffron | – |
| 1026. Li 2021. Effect of Lactobacillus casei on lipid metabolism and intestinal microflora in patients with alcoholic liver injury<br>PMID 33514869 | Grace Kim: No saffron | Pranav Potluri: No saffron | – |
| 1027. Dhakal 2021. Antioxidant, Cytotoxic, and Antibacterial Activities of the Selected Tibetan Formulations Used in Gandaki Province, Nepal<br>PMID 34422072 | Grace Kim: No saffron | Pranav Potluri: No saffron | – |
| 1028. Abdelghani 2021. Therapeutic applications and biological activities of bacterial bioactive extracts<br>PMID 34370077 | Grace Kim: No saffron | Pranav Potluri: No saffron | – |
| 1029. Sabzevari Rad 2020. Micro RNA-126 promoting angiogenesis in diabetic heart by VEGF/Spred-1/Raf-1 pathway: effects of high-intensity interval training<br>PMID 33520826 | Grace Kim: No saffron | Pranav Potluri: No saffron | – |
| 1030. Di Nardo 2021. Ten Years of Lateral Flow Immunoassay Technique Applications: Trends, Challenges and Future Perspectives<br>PMID 34372422 | Grace Kim: No saffron | Pranav Potluri: No saffron | – |
| 1031. Felisbino 2021. Nutrigenomics in Regulating the Expression of Genes Related to Type 2 Diabetes Mellitus<br>PMID 34366888 | Grace Kim: No saffron | Pranav Potluri: No saffron | – |

Table S2 continued

| Paper | Reviewer 1 | Reviewer 2 | Additional recorded reviews |
|---|---|---|---|
| 1032. Liu 2021. Serum cardiovascular-related metabolites disturbance exposed to different heavy metal exposure scenarios<br>PMID 33740723 | Grace Kim: No saffron | Pranav Potluri: No saffron | – |
| 1033. Martinez-Arroyo 2020. The Rab-Rabphilin system in injured human podocytes stressed by glucose overload and angiotensin II<br>PMID 32567349 | Grace Kim: No saffron | Pranav Potluri: No saffron | – |
| 1034. Wickramasinghe 2021. Herbal Medicines Targeting the Improved $\beta$-Cell Functions and $\beta$-Cell Regeneration for the Management of Diabetes Mellitus<br>PMID 34335803 | Grace Kim: No saffron | Pranav Potluri: No saffron | – |
| 1035. Zhu 2021. The effects of compound centella formula on OxInflammation and silent information regulator 1 in a high-fat diet/streptozotocin-induced diabetic kidney disease rat model<br>PMID 34335904 | Grace Kim: No saffron | Pranav Potluri: No saffron | – |
| 1036. Paratthakonkun 2021. The Effects of Crocodile Blood Supplementation on Delayed-Onset Muscle Soreness<br>PMID 34371824 | Grace Kim: No saffron | Pranav Potluri: No saffron | – |
| 1037. Terasaki 2021. Glucose-Dependent Insulinotropic Polypeptide Suppresses Foam Cell Formation of Macrophages through Inhibition of the Cyclin-Dependent Kinase 5-CD36 Pathway<br>PMID 34356896 | Grace Kim: No saffron | Pranav Potluri: No saffron | – |
| 1038. Jin 2021. MicroRNA-126 Level Increases During Exercise Rehabilitation of Heart Failure with a Preserved Ejection Fraction<br>PMID 34285564 | Grace Kim: No saffron | Pranav Potluri: No saffron | – |
| 1039. Gorka 2021. A dimeric chlorophyll electron acceptor differentiates type I from type II photosynthetic reaction centers<br>PMID 34278250 | Grace Kim: No saffron | Pranav Potluri: No saffron | – |
| 1040. Gilmartin 2021. In vitro and in vivo induction of fetal hemoglobin with a reversible and selective DNMT1 inhibitor<br>PMID 32586904 | Grace Kim: No saffron | Pranav Potluri: No saffron | – |
| 1041. Jakubiak 2021. Oxidative Stress in Association with Metabolic Health and Obesity in Young Adults<br>PMID 34257828 | Grace Kim: No saffron | Pranav Potluri: No saffron | – |

Table S2 continued

| Paper | Reviewer 1 | Reviewer 2 | Additional recorded reviews |
|---|---|---|---|
| 1042. Vecchiatto 2021. Oxidative phenotype induced by aerobic physical training prevents the obesity-linked insulin resistance without changes in gastrocnemius muscle ACE2-Angiotensin(1-7)-Mas axis<br>PMID 34229719 | Grace Kim: No saffron | Pranav Potluri: No saffron | – |
| 1043. Morin 2021. Pan-European meteorological and snow indicators of climate change impact on ski tourism<br>PMID 34239989 | Grace Kim: No saffron | Pranav Potluri: No saffron | – |
| 1044. Kaur 2021. Mechanisms and Therapeutic Prospects of Diabetic Cardiomyopathy Through the Inflammatory Response<br>PMID 34234695 | Grace Kim: No saffron | Pranav Potluri: No saffron | – |
| 1045. Al-Awaida 2021. Evaluation of the Genetic Association and Expressions of Notch-2 /Jagged-1 in Patients with Type 2 Diabetes Mellitus<br>PMID 34219868 | Grace Kim: No saffron | Pranav Potluri: No saffron | – |
| 1046. Khazraei 2021. Mechanical effects of ranolazine on normal and diabetic-isolated rat heart<br>PMID 34221060 | Grace Kim: No saffron | Pranav Potluri: No saffron | – |
| 1047. Mohammed Basheeruddin Asdaq 2021. Anti-obesity potential of almond (Prunus dulcis) in experimental animals under cafeteria and atherogenic diets<br>PMID 34220264 | Grace Kim: No saffron | Pranav Potluri: No saffron | – |
| 1048. Duggan 2021. Changes in Dietary Inflammatory Index Patterns with Weight Loss in Women: A Randomized Controlled Trial<br>PMID 32859616 | Grace Kim: No saffron | Alexander Tai Nguyen: No saffron | – |
| 1049. Samadi 2021. The effect of tropisetron on oxidative stress, SIRT1, FOXO3a, and claudin-1 in the renal tissue of STZ-induced diabetic rats<br>PMID 33047279 | Grace Kim: No saffron | Alexander Tai Nguyen: No saffron | – |
| 1050. Zaazaa 2021. Determination of Mineral Composition and Phenolic Content and Investigation of Antioxidant, Antidiabetic, and Antibacterial Activities of Crocus sativus L. Aqueous Stigmas Extracts<br>PMID 34195613 | Grace Kim: Not clinical | Alexander Tai Nguyen: Not clinical | – |
| 1051. Eslampour 2021. "Association between dietary inflammatory index (DII) and risk of irritable bowel syndrome: a case-control study"<br>PMID 34183016 | Grace Kim: No saffron | Alexander Tai Nguyen: Not isolated | – |

Table S2 continued

| Paper | Reviewer 1 | Reviewer 2 | Additional recorded reviews |
|---|---|---|---|
| 1052. Kuczmarski 2021. Healthy Behaviors Associated with Changes in Mental and Physical Strength in Urban African American and White Adults<br>PMID 34071874 | Grace Kim: No saffron | Alexander Tai Nguyen: No saffron | – |
| 1053. Ayoub 2021. Anti-Allergic, Anti-Inflammatory and Anti-Hyperglycemic Activity of Chasmanthe aethiopica Leaf Extract and Its Profiling Using LC/MS and GLC/MS<br>PMID 34073129 | Grace Kim: No saffron | Alexander Tai Nguyen: No saffron | – |
| 1054. Kamil 2021. Toxicological Evaluation of Novel Cyclohexenone Derivative in an Animal Model through Histopathological and Biochemical Techniques<br>PMID 34070633 | Grace Kim: No saffron | Alexander Tai Nguyen: No saffron | – |
| 1055. Xu 2021. Exploring the Protective Effects and Mechanism of Crocetin From Saffron Against NAFLD by Network Pharmacology and Experimental Validation<br>PMID 34179049 | Grace Kim: Not clinical | Alexander Tai Nguyen: No saffron | – |
| 1056. Maita 2021. An undiagnosed patient with skin rash, polyarthritis, and edema responding to low-dose colchicine: A case report<br>PMID 34178350 | Grace Kim: No saffron | Alexander Tai Nguyen: No saffron | – |
| 1057. Hawash 2021. Molecular docking, chemo-informatic properties, alpha-amylase, and lipase inhibition studies of benzodioxol derivatives<br>PMID 34162414 | Grace Kim: No saffron | Alexander Tai Nguyen: Not clinical | – |
| 1058. Zia 2021. Molecular and cellular pathways contributing to brain aging<br>PMID 34118939 | Grace Kim: No saffron | Alexander Tai Nguyen: No saffron | – |
| 1059. Ganjooei 2021. The Role of Lipid Profile as an Independent Predictor of Non-alcoholic Steatosis and Steatohepatitis in Morbidly Obese Patients<br>PMID 34136549 | Grace Kim: No saffron | Alexander Tai Nguyen: No saffron | – |
| 1060. Adamska 2021. Bioactive Substances in Safflower Flowers and Their Applicability in Medicine and Health-Promoting Foods<br>PMID 34136564 | Grace Kim: Not clinical | Alexander Tai Nguyen: No saffron | – |
| 1061. Himori 2021. The Effect of Dietary Antioxidant Supplementation in Patients with Glaucoma<br>PMID 34113073 | Grace Kim: No saffron | Alexander Tai Nguyen: Not isolated | – |

Table S2 continued

| Paper | Reviewer 1 | Reviewer 2 | Additional recorded reviews |
|---|---|---|---|
| 1062. Zhang 2021. Turmeric and its bioactive constituents trigger cell signaling mechanisms that protect against diabetes and cardiovascular diseases<br>PMID 34106380 | Grace Kim: No saffron | Alexander Tai Nguyen: No saffron | – |
| 1063. Zobeiri 2021. Evaluation of the Effectiveness of Cinnamon Oil Soft Capsule in Patients with Functional Dyspepsia: A Randomized Double-Blind Placebo-Controlled Clinical Trial<br>PMID 34093719 | Grace Kim: No saffron | Alexander Tai Nguyen: No saffron | – |
| 1064. Soltani 2021. Association between dietary inflammatory potential and risk of developing gestational diabetes: a prospective cohort study<br>PMID 34078385 | Grace Kim: No saffron | Alexander Tai Nguyen: No saffron | – |
| 1065. Cheshmeh 2022. Green cardamom plus low-calorie diet can decrease the expression of inflammatory genes among obese women with polycystic ovary syndrome: a double-blind randomized clinical trial<br>PMID 34057705 | Grace Kim: No saffron | Alexander Tai Nguyen: No saffron | – |
| 1066. Islam 2021. Recent Molecular Mechanisms and Beneficial Effects of Phytochemicals and Plant-Based Whole Foods in Reducing LDL-C and Preventing Cardiovascular Disease<br>PMID 34063371 | Grace Kim: No saffron | Alexander Tai Nguyen: No saffron | – |
| 1067. Balejko 2021. Use of a Highly Antioxidant Diet in the Regulation of Adipose Tissue Secretion in Patients after the BIB Procedure<br>PMID 34067712 | Grace Kim: No saffron | Alexander Tai Nguyen: No saffron | – |
| 1068. Miziak 2021. Some Candidate Drugs for Pharmacotherapy of Alzheimer's Disease<br>PMID 34068096 | Grace Kim: No saffron | Alexander Tai Nguyen: No saffron | – |
| 1069. Sharifi-Rad 2021. Cinnamomum Species: Bridging Phytochemistry Knowledge, Pharmacological Properties and Toxicological Safety for Health Benefits<br>PMID 34045956 | Grace Kim: No saffron | Alexander Tai Nguyen: No saffron | – |
| 1070. Ungurianu 2021. Vitamin E beyond Its Antioxidant Label<br>PMID 33919211 | Grace Kim: No saffron | Alexander Tai Nguyen: No saffron | – |
| 1071. Zhao 2021. NmFGF1-Regulated Glucolipid Metabolism and Angiogenesis Improves Functional Recovery in a Mouse Model of Diabetic Stroke and Acts via the AMPK Signaling Pathway<br>PMID 34025437 | Grace Kim: No saffron | Alexander Tai Nguyen: No saffron | – |

Table S2 continued

| Paper | Reviewer 1 | Reviewer 2 | Additional recorded reviews |
|---|---|---|---|
| 1072. Ateba 2021. The Genus Eriosema (Fabaceae): From the Ethnopharmacology to an Evidence-Based Phytotherapeutic Perspective? PMID 34025412 | Grace Kim: No saffron | Alexander Tai Nguyen: No saffron | – |
| 1073. Chen 2021. Anti-inflammatory activities of Gardenia jasminoides extracts in retinal pigment epithelial cells and zebrafish embryos PMID 34007309 | Grace Kim: Not clinical | Alexander Tai Nguyen: No saffron | – |
| 1074. He 2021. Nanotechnology in the Treatment of Diabetic Complications: A Comprehensive Narrative Review PMID 34007847 | Grace Kim: No saffron | Alexander Tai Nguyen: No saffron | – |
| 1075. Arruda 2021. Anthocyanins Recovered from Agri-Food By-Products Using Innovative Processes: Trends, Challenges, and Perspectives for Their Application in Food Systems PMID 33946376 | Grace Kim: No saffron | Alexander Tai Nguyen: No saffron | – |
| 1076. Yaribeygi 2021. Antioxidative Potentials of Incretin-Based Medications: A Review of Molecular Mechanisms PMID 34007411 | Grace Kim: No saffron | Alexander Tai Nguyen: No saffron | – |
| 1077. Zhou 2021. microRNA-363-3p reduces endothelial cell inflammatory responses in coronary heart disease via inactivation of the NOX4-dependent p38 MAPK axis PMID 33744854 | Grace Kim: No saffron | Alexander Tai Nguyen: No saffron | – |
| 1078. van Eenige 2021. Cannabinoid type 1 receptor inverse agonism attenuates dyslipidemia and atherosclerosis in APOE∗3-Leiden.CETP mice PMID 33766515 | Grace Kim: No saffron | Alexander Tai Nguyen: No saffron | – |
| 1079. Costa Dos Santos G Jr 2021. The remodel of the "central dogma": a metabolomics interaction perspective PMID 33969452 | Grace Kim: No saffron | Alexander Tai Nguyen: No saffron | – |
| 1080. Lopresti 2020. Association between Micronutrients and Heart Rate Variability: A Review of Human Studies PMID 31942924 | Grace Kim: No saffron | Alexander Tai Nguyen: No saffron | – |
| 1081. Rosenzweig 2021. Activation of Insulin Signaling by Botanical Products PMID 33919569 | Grace Kim: Not clinical | Alexander Tai Nguyen: Not clinical | – |
| 1082. Wang 2021. Targets identified from exercised heart: killing multiple birds with one stone PMID 33837221 | Grace Kim: No saffron | Alexander Tai Nguyen: No saffron | – |

Table S2 continued

| Paper | Reviewer 1 | Reviewer 2 | Additional recorded reviews |
|---|---|---|---|
| 1083. Alfonso-Muñoz 2021. Role of Oral Antioxidant Supplementation in the Current Management of Diabetic Retinopathy<br>PMID 33924714 | Grace Kim: Not clinical | Alexander Tai Nguyen: No saffron | – |
| 1084. Gregory 2021. Neuroprotective Herbs for the Management of Alzheimer's Disease<br>PMID 33917843 | Grace Kim: Not clinical | Alexander Tai Nguyen: No saffron | – |
| 1085. Nevara 2021. Dietary Fiber: Fractionation, Characterization and Potential Sources from Defatted Oilseeds<br>PMID 33918108 | Grace Kim: No saffron | Alexander Tai Nguyen: No saffron | – |
| 1086. Machado 2021. Association between Dietary Inflammatory Index and Periodontitis: A Cross-Sectional and Mediation Analysis<br>PMID 33916342 | Grace Kim: No saffron | Alexander Tai Nguyen: No saffron | – |
| 1087. Khan 2021. Determinants of Serum Immunoglobulin Levels: A Systematic Review and Meta-Analysis<br>PMID 33897714 | Grace Kim: No saffron | Alexander Tai Nguyen: No saffron | – |
| 1088. Bronczek 2021. Resistance exercise training improves glucose homeostasis by enhancing insulin secretion in C57BL/6 mice<br>PMID 33883630 | Grace Kim: No saffron | Alexander Tai Nguyen: No saffron | – |
| 1089. Gawlik-Kotelnicka 2021. The Influence of Probiotic Supplementation on Depressive Symptoms, Inflammation, and Oxidative Stress Parameters and Fecal Microbiota in Patients with Depression Depending on Metabolic Syndrome Comorbidity-PRO-DEMET Randomized Study Protocol<br>PMID 33804999 | Grace Kim: No saffron | Alexander Tai Nguyen: No saffron | – |
| 1090. Henning 2021. Redox biomarkers in dietary interventions and nutritional observation studies - From new insights to old problems<br>PMID 33756398 | Grace Kim: Not clinical | Alexander Tai Nguyen: Not clinical | – |
| 1091. Saghafi-Asl 2021. The association of dietary patterns with dietary inflammatory index, systemic inflammation, and insulin resistance, in apparently healthy individuals with obesity<br>PMID 33824355 | Grace Kim: No saffron | Alexander Tai Nguyen: Not isolated | – |
| 1092. Marmitt 2021. Compounds of plants with activity against SARS-CoV-2 targets<br>PMID 33706626 | Grace Kim: No saffron | Alexander Tai Nguyen: No saffron | – |

Table S2 continued

| Paper | Reviewer 1 | Reviewer 2 | Additional recorded reviews |
|---|---|---|---|
| 1093. Mueller 2021. Cholesterol Accumulation as a Driver of Hepatic Inflammation Under Translational Dietary Conditions Can Be Attenuated by a Multicomponent Medicine PMID 33815271 | Grace Kim: No saffron | Alexander Tai Nguyen: No saffron | – |
| 1094. Bafadam 2021. Cardioprotective effects of Fenugreek (Trigonella foenum-graceum) seed extract in streptozotocin induced diabetic rats PMID 33815699 | Grace Kim: No saffron | Alexander Tai Nguyen: No saffron | – |
| 1095. Lee 2021. Regulation of miRNAs by Natural Antioxidants in Cardiovascular Diseases: Focus on SIRT1 and eNOS PMID 33802566 | Grace Kim: Not clinical | Alexander Tai Nguyen: No saffron | – |
| 1096. Han 2021. The Oral Administration of Sanguisorba officinalis Extract Improves Physical Performance through LDHA Modulation PMID 33809377 | Grace Kim: No saffron | Alexander Tai Nguyen: No saffron | – |
| 1097. Li 2021. Crocin Improves Endothelial Mitochondrial Dysfunction via GPx1/ROS/KCa3.1 Signal Axis in Diabetes PMID 33777959 | Grace Kim: Not clinical | Alexander Tai Nguyen: Not clinical | – |
| 1098. Lan 2021. Hepatoprotective effect of Qushihuayu formula on non-alcoholic steatohepatitis induced by MCD diet in rat PMID 33726778 | Grace Kim: No saffron | Alexander Tai Nguyen: Not clinical | – |
| 1099. Saeedan 2021. Artemisia judaica L. diminishes diabetes-induced reproductive dysfunction in male rats via activation of Nrf2/HO-1-mediated antioxidant responses PMID 33732055 | Grace Kim: No saffron | Alexander Tai Nguyen: No saffron | – |
| 1100. Al-Kuraishy 2021. Sequential doxycycline and colchicine combination therapy in Covid-19: The salutary effects PMID 33727066 | Grace Kim: No saffron | Alexander Tai Nguyen: No saffron | – |
| 1101. Lucas 2021. Cinnamon and Hop Extracts as Potential Immunomodulators for Severe COVID-19 Cases PMID 33719281 | Grace Kim: No saffron | Alexander Tai Nguyen: No saffron | – |
| 1102. Singh 2021. Pharmacological advances in mitochondrial therapy PMID 33647769 | Grace Kim: No saffron | Alexander Tai Nguyen: No saffron | – |
| 1103. Gao 2021. Applications and challenges of low temperature plasma in pharmaceutical field PMID 33717609 | Grace Kim: Not clinical | Alexander Tai Nguyen: No saffron | – |

Table S2 continued

| Paper | Reviewer 1 | Reviewer 2 | Additional recorded reviews |
|---|---|---|---|
| 1104. Dantas-Medeiros 2021. Antifungal and Antibiofilm Activities of B-Type Oligomeric Procyanidins From Commiphora leptophloeos Used Alone or in Combination With Fluconazole Against Candida spp<br>PMID 33692765 | Grace Kim: No saffron | Alexander Tai Nguyen: No saffron | – |
| 1105. Omidkhoda 2020. Protective Effects of Crocin Against Hepatic Damages in D-galactose Aging Model in Rats<br>PMID 33680043 | Grace Kim: Not clinical | Alexander Tai Nguyen: Not clinical | – |
| 1106. Khodavirdipour 2020. Chromium Supplementation; Negotiation with Diabetes Mellitus, Hyperlipidemia and Depression<br>PMID 32550211 | Grace Kim: No saffron | Alexander Tai Nguyen: No saffron | – |
| 1107. Gaglio 2021. Effect on the Antioxidant, Lipoperoxyl Radical Scavenger Capacity, Nutritional, Sensory and Microbiological Traits of an Ovine Stretched Cheese Produced with Grape Pomace Powder Addition<br>PMID 33671122 | Grace Kim: No saffron | Alexander Tai Nguyen: No saffron | – |
| 1108. Majumdar 2021. Perspectives About Modulating Host Immune System in Targeting SARS-CoV-2 in India<br>PMID 33664772 | Grace Kim: No saffron | Alexander Tai Nguyen: No saffron | – |
| 1109. Ali 2021. Insulin-Mimetic Dihydroxanthyletin-Type Coumarins from Angelica decursiva with Protein Tyrosine Phosphatase 1B and $\alpha$-Glucosidase Inhibitory Activities and Docking Studies of Their Molecular Mechanisms<br>PMID 33672051 | Grace Kim: No saffron | Alexander Tai Nguyen: No saffron | – |
| 1110. Win 2021. Pharmacological and Metabolic Significance of Bile Acids in Retinal Diseases<br>PMID 33669313 | Grace Kim: No saffron | Alexander Tai Nguyen: No saffron | – |
| 1111. Thiem 2020. Deletion of haematopoietic Dectin-2 or CARD9 does not protect from atherosclerosis development under hyperglycaemic conditions<br>PMID 31868000 | Grace Kim: No saffron | Alexander Tai Nguyen: No saffron | – |
| 1112. Poasakate 2021. Genistein Prevents Nitric Oxide Deficiency-Induced Cardiac Dysfunction and Remodeling in Rats<br>PMID 33557258 | Grace Kim: No saffron | Alexander Tai Nguyen: No saffron | – |
| 1113. Tribble 2021. Targeting Diet and Exercise for Neuroprotection and Neurorecovery in Glaucoma<br>PMID 33535578 | Grace Kim: No saffron | Alexander Tai Nguyen: No saffron | – |

Table S2 continued

| Paper | Reviewer 1 | Reviewer 2 | Additional recorded reviews |
|---|---|---|---|
| 1114. Fierascu 2021. Selected Aspects Related to Medicinal and Aromatic Plants as Alternative Sources of Bioactive Compounds<br>PMID 33546333 | Grace Kim: No saffron | Alexander Tai Nguyen: No saffron | – |
| 1115. Saleh 2021. UPLC-MS-Based Metabolomics Profiling for $\alpha$-Glucosidase Inhibiting Property of Parkia speciosa Pods<br>PMID 33499128 | Grace Kim: No saffron | Alexander Tai Nguyen: No saffron | – |
| 1116. Gaderpour 2021. Voluntary exercise improves spermatogenesis and testicular apoptosis in type 2 diabetic rats through alteration in oxidative stress and mir-34a/SIRT1/p53 pathway<br>PMID 33643571 | Grace Kim: No saffron | Alexander Tai Nguyen: No saffron | – |
| 1117. Farhang 2021. An Observational Cohort of First Episode Psychosis in Iran: The Azeri Recent Onset Acute Phase Psychosis Survey (ARAS Cohort) Study Protocol<br>PMID 33643098 | Grace Kim: No saffron | Alexander Tai Nguyen: No saffron | – |
| 1118. Shirzad 2021. Cinnamon effects on blood pressure and metabolic profile: A double-blind, randomized, placebo-controlled trial in patients with stage 1 hypertension<br>PMID 33628723 | Grace Kim: No saffron | Alexander Tai Nguyen: No saffron | – |
| 1119. Sensi 2019. Acute adrenal failure: a potentially fatal consequence of an adulterated herbal remedy<br>PMID 30787026 | Grace Kim: No saffron | Alexander Tai Nguyen: No saffron | – |
| 1120. Abdi Gorabi 2020. The Effects of Ripe Pistachio Hulls Hydroalcoholic Extract and Aerobic Training on Learning and Memory in Streptozotocin-induced Diabetic Male Rats<br>PMID 33613891 | Grace Kim: No saffron | Alexander Tai Nguyen: No saffron | – |
| 1121. Grzych 2020. NASH-related increases in plasma bile acid levels depend on insulin resistance<br>PMID 33615207 | Grace Kim: No saffron | Alexander Tai Nguyen: No saffron | – |
| 1122. Rondanelli 2021. Where to Find Leucine in Food and How to Feed Elderly With Sarcopenia in Order to Counteract Loss of Muscle Mass: Practical Advice<br>PMID 33585538 | Grace Kim: No saffron | Alexander Tai Nguyen: No saffron | – |
| 1123. Zhou 2021. Antioxidant Food Components for the Prevention and Treatment of Cardiovascular Diseases: Effects, Mechanisms, and Clinical Studies<br>PMID 33574978 | Grace Kim: Not clinical | Alexander Tai Nguyen: No saffron | – |

Table S2 continued

| Paper | Reviewer 1 | Reviewer 2 | Additional recorded reviews |
|---|---|---|---|
| 1124. Zhao 2021. Mechanism of long non-coding RNA metastasis-associated lung adenocarcinoma transcript 1 in lipid metabolism and inflammation in heart failure PMID 33448307 | Grace Kim: No saffron | Alexander Tai Nguyen: No saffron | – |
| 1125. Floris 2021. Washingtonia filifera seed extracts inhibit the islet amyloid polypeptide fibrils formations and $\alpha$-amylase and $\alpha$-glucosidase activity PMID 33494628 | Grace Kim: No saffron | Alexander Tai Nguyen: No saffron | – |
| 1126. Jin 2021. Allisartan isoproxil attenuates oxidative stress and inflammation through the SIRT1/Nrf2/NF-$\kappa$B signalling pathway in diabetic cardiomyopathy rats PMID 33495841 | Grace Kim: No saffron | Alexander Tai Nguyen: No saffron | – |
| 1127. Sena-Júnior 2020. Whether or Not the Effects of Curcuma longa Supplementation Are Associated with Physical Exercises in T1DM and T2DM: A Systematic Review PMID 33396291 | Grace Kim: No saffron | Alexander Tai Nguyen: No saffron | – |
| 1128. Merra 2020. Influence of Mediterranean Diet on Human Gut Microbiota PMID 33375042 | Grace Kim: No saffron | Alexander Tai Nguyen: No saffron | – |
| 1129. Li 2021. Herbal formula LLKL ameliorates hyperglycaemia, modulates the gut microbiota and regulates the gut-liver axis in Zucker diabetic fatty rats PMID 33215869 | Grace Kim: Not clinical | Alexander Tai Nguyen: Not clinical | – |
| 1130. Yang 2020. Quercetin: Its Main Pharmacological Activity and Potential Application in Clinical Medicine PMID 33488935 | Grace Kim: No saffron | Alexander Tai Nguyen: No saffron | – |
| 1131. Mohaqiq 2020. Antioxidant Properties of Saffron Stigma and Petals: A Potential Therapeutic Approach for Insulin Resistance through an Insulin-Sensitizing Adipocytokine in High-Calorie Diet Rats PMID 33456740 | Grace Kim: Not clinical | Alexander Tai Nguyen: Not clinical | – |
| 1132. Rashedinia 2020. Neuroprotective Effect of Syringic Acid by Modulation of Oxidative Stress and Mitochondrial Mass in Diabetic Rats PMID 33457416 | Grace Kim: No saffron | Alexander Tai Nguyen: No saffron | – |
| 1133. Ribeiro-Parenti 2021. Bariatric surgery induces a new gastric mucosa phenotype with increased functional glucagon-like peptide-1 expressing cells PMID 33397977 | Grace Kim: No saffron | Alexander Tai Nguyen: No saffron | – |

Table S2 continued

| Paper | Reviewer 1 | Reviewer 2 | Additional recorded reviews |
|---|---|---|---|
| 1134. Lum 2021. Protective effect of mangiferin on memory impairment: A systematic review<br>PMID 33424383 | Grace Kim: No saffron | Alexander Tai Nguyen: No saffron | – |
| 1135. Ruiz-Saavedra 2020. Comparison of Different Dietary Indices as Predictors of Inflammation, Oxidative Stress and Intestinal Microbiota in Middle-Aged and Elderly Subjects<br>PMID 33333806 | Grace Kim: No saffron | Alexander Tai Nguyen: Not isolated | – |
| 1136. Skourtis 2020. Protective antioxidant effects of saffron extract on retinas of streptozotocin-induced diabetic rats<br>PMID 33367177 | Grace Kim: Not clinical | Alexander Tai Nguyen: Not clinical | – |
| 1137. Gál 2020. Bile accelerates carcinogenic processes in pancreatic ductal adenocarcinoma cells through the overexpression of MUC4<br>PMID 33328627 | Grace Kim: No saffron | Alexander Tai Nguyen: No saffron | – |
| 1138. Author not recorded 2020. The 46(th) Annual Meeting of the European Society for Blood and Marrow Transplantation: Physicians Poster Session (P001-P706)<br>PMID 33311612 | Grace Kim: No saffron | Alexander Tai Nguyen: No saffron | – |
| 1139. Rubio 2020. Plant-based and cell-based approaches to meat production<br>PMID 33293564 | Grace Kim: No saffron | Alexander Tai Nguyen: No saffron | – |
| 1140. Schilperoort 2020. Continuous Light Does Not Affect Atherosclerosis in APOE*3-Leiden.CETP Mice<br>PMID 32915671 | Grace Kim: No saffron | Alexander Tai Nguyen: No saffron | – |
| 1141. Emamat 2020. Effect of barberry (Berberis vulgaris) consumption on blood pressure, plasma lipids, and inflammation in patients with hypertension and other cardiovascular risk factors: study protocol for a randomized clinical trial<br>PMID 33246500 | Grace Kim: No saffron | Alexander Tai Nguyen: No saffron | – |
| 1142. Zeka 2020. Activity of Antioxidants from Crocus sativus L. Petals: Potential Preventive Effects towards Cardiovascular System<br>PMID 33182461 | Grace Kim: Not clinical | Alexander Tai Nguyen: Not clinical | – |
| 1143. Sanz-González 2020. Clinical and Molecular-Genetic Insights into the Role of Oxidative Stress in Diabetic Retinopathy: Antioxidant Strategies and Future Avenues<br>PMID 33182408 | Grace Kim: No saffron | Alexander Tai Nguyen: No saffron | – |

Table S2 continued

| Paper | Reviewer 1 | Reviewer 2 | Additional recorded reviews |
|---|---|---|---|
| 1144. Soleti 2020. Apple Supplementation Improves Hemodynamic Parameter and Attenuates Atherosclerosis in High-Fat Diet-Fed Apolipoprotein E-Knockout Mice PMID 33198144 | Grace Kim: No saffron | Alexander Tai Nguyen: No saffron | – |
| 1145. Kakouri 2020. Crocins from Crocus sativus L. in the Management of Hyperglycemia. In Vivo Evidence from Zebrafish PMID 33182581 | Grace Kim: Not clinical | Alexander Tai Nguyen: Not clinical | – |
| 1146. Stasiewicz 2020. Hybrid Dietary-Blood Inflammatory Profiles and Postmenopausal Breast Cancer: A Case-Control Study PMID 33202561 | Grace Kim: No saffron | Alexander Tai Nguyen: No saffron | – |
| 1147. Fakhlaei 2020. The Toxic Impact of Honey Adulteration: A Review PMID 33114468 | Grace Kim: No saffron | Pranav Potluri: No saffron | Alexander Tai Nguyen: No saffron |
| 1148. Jampilek 2020. Potential of Nanonutraceuticals in Increasing Immunity PMID 33182343 | Grace Kim: No saffron | Pranav Potluri: No saffron | Alexander Tai Nguyen: No saffron |
| 1149. Aljehani 2020. Icariin enhances AMP-activated protein kinase and prevents high fructose and high salt-induced metabolic syndrome in rats PMID 33250640 | Grace Kim: No saffron | Pranav Potluri: No saffron | Alexander Tai Nguyen: Not clinical |
| 1150. Liu 2020. Panax ginseng C. A. Meyer as a potential therapeutic agent for organ fibrosis disease PMID 33292321 | Grace Kim: No saffron | Pranav Potluri: No saffron | Alexander Tai Nguyen: No saffron |
| 1151. Zhang 2020. Maternal sitagliptin treatment attenuates offspring glucose metabolism and intestinal proinflammatory cytokines IL-6 and TNF-$\alpha$ expression in male rats PMID 33240638 | Grace Kim: No saffron | Pranav Potluri: No saffron | Alexander Tai Nguyen: No saffron |
| 1152. Mahdian 2020. Effect of Boswellia species on the metabolic syndrome: A review PMID 33235693 | Grace Kim: No saffron | Pranav Potluri: No saffron | Alexander Tai Nguyen: No saffron |
| 1153. Zhang 2020. Association Between Cannabinoid Receptor-1 Gene Polymorphism and the Risk of Diabetic Nephropathy Among Patients with Type 2 Diabetes Mellitus PMID 33209051 | Grace Kim: No saffron | Pranav Potluri: No saffron | Alexander Tai Nguyen: No saffron |
| 1154. Zheng 2020. Dietary inflammatory potential in relation to the gut microbiome: results from a cross-sectional study PMID 32475373 | Grace Kim: No saffron | Pranav Potluri: No saffron | Alexander Tai Nguyen: Not isolated |

Table S2 continued

| Paper | Reviewer 1 | Reviewer 2 | Additional recorded reviews |
|---|---|---|---|
| 1155. Farkhondeh 2020. An Overview of the Role of Adipokines in Cardiometabolic Diseases<br>PMID 33182462 | Grace Kim: No saffron | Pranav Potluri: No saffron | Alexander Tai Nguyen: No saffron |
| 1156. Ee 2020. An integrative collaborative care model for people with mental illness and physical comorbidities<br>PMID 33292354 | Grace Kim: No saffron | Pranav Potluri: No saffron | Alexander Tai Nguyen: No saffron |
| 1157. Cheng 2020. Prothrombotic effects of high uric acid in mice via activation of MEF2C-dependent NF-$\kappa$B pathway by upregulating let-7c<br>PMID 32960786 | Grace Kim: No saffron | Pranav Potluri: No saffron | Alexander Tai Nguyen: No saffron |
| 1158. Chiu 2019. Association between intake of fruits and vegetables by pesticide residue status and coronary heart disease risk<br>PMID 31473415 | Grace Kim: No saffron | Pranav Potluri: No saffron | Alexander Tai Nguyen: No saffron |
| 1159. Zehiroglu 2019. The importance of antioxidants and place in today's scientific and technological studies<br>PMID 31741500 | Grace Kim: No saffron | Alexander Tai Nguyen: No saffron | – |
| 1160. Hashiesh 2020. Therapeutic Potential of $\beta$-Caryophyllene: A Dietary Cannabinoid in Diabetes and Associated Complications<br>PMID 32998300 | Grace Kim: No saffron | Alexander Tai Nguyen: No saffron | – |
| 1161. Reyes-Pavón 2020. Protective Effect of Glycomacropeptide on Food Allergy with Gastrointestinal Manifestations in a Rat Model through Down-Regulation of Type 2 Immune Response<br>PMID 32992996 | Grace Kim: No saffron | Alexander Tai Nguyen: No saffron | – |
| 1162. Karigidi 2019. Antidiabetic activity of corn steep liquor extract of Curculigo pilosa and its solvent fractions in streptozotocin-induced diabetic rats<br>PMID 33134131 | Grace Kim: No saffron | Alexander Tai Nguyen: No saffron | – |
| 1163. Nader 2020. Cross-Species Proteomics Identifies CAPG and SBP1 as Crucial Invasiveness Biomarkers in Rat and Human Malignant Mesothelioma<br>PMID 32867073 | Grace Kim: No saffron | Alexander Tai Nguyen: No saffron | – |
| 1164. Shin 2020. Antiulcer Activity of Steamed Ginger Extract against Ethanol/HCl-Induced Gastric Mucosal Injury in Rats<br>PMID 33066164 | Grace Kim: No saffron | Alexander Tai Nguyen: No saffron | – |
| 1165. Efremov 2020. Differential Regulation of Phytoene Synthase PSY1 During Fruit Carotenogenesis in Cultivated and Wild Tomato Species (Solanum section Lycopersicon)<br>PMID 32916928 | Grace Kim: No saffron | Alexander Tai Nguyen: No saffron | – |

Table S2 continued

| Paper | Reviewer 1 | Reviewer 2 | Additional recorded reviews |
|---|---|---|---|
| 1166. Song 2020. Efficiency and Safety of Chinese Herbal Medicine in the Treatment of Prediabetes: A Systemic Review and Meta-Analysis of Randomized Controlled Trials<br>PMID 33123206 | Grace Kim: No saffron | Alexander Tai Nguyen: No saffron | – |
| 1167. Chota 2020. Potential Treatment of Breast and Lung Cancer Using Dicoma anomala, an African Medicinal Plant<br>PMID 32992537 | Grace Kim: No saffron | Alexander Tai Nguyen: No saffron | – |
| 1168. Ahmed 2020. Nephroprotective Effect of Pleurotus ostreatus and Agaricus bisporus Extracts and Carvedilol on Ethylene Glycol-Induced Urolithiasis: Roles of NF-$\kappa$B, p53, Bcl-2, Bax and Bak<br>PMID 32937925 | Grace Kim: No saffron | Alexander Tai Nguyen: No saffron | – |
| 1169. van den Hoek 2020. A Translational Mouse Model for NASH with Advanced Fibrosis and Atherosclerosis Expressing Key Pathways of Human Pathology<br>PMID 32883049 | Grace Kim: No saffron | Alexander Tai Nguyen: No saffron | – |
| 1170. Carvalho 2020. Effectiveness of ginger in reducing metabolic levels in people with diabetes: a randomized clinical trial<br>PMID 33053078 | Grace Kim: No saffron | Alexander Tai Nguyen: No saffron | – |
| 1171. Malfa 2020. The Antioxidant Activities of Betula etnensis Rafin. Ethanolic Extract Exert Protective and Anti-diabetic Effects on Streptozotocin-Induced Diabetes in Rats<br>PMID 32927638 | Grace Kim: No saffron | Alexander Tai Nguyen: No saffron | – |
| 1172. Diaconeasa 2020. Anthocyanins, Vibrant Color Pigments, and Their Role in Skin Cancer Prevention<br>PMID 32916849 | Grace Kim: No saffron | Alexander Tai Nguyen: No saffron | – |
| 1173. Belosludtsev 2020. Diabetes Mellitus, Mitochondrial Dysfunction and Ca(2+)-Dependent Permeability Transition Pore<br>PMID 32911736 | Grace Kim: No saffron | Alexander Tai Nguyen: No saffron | – |
| 1174. Ariya 2020. Dietary inflammatory index and metabolic syndrome in Iranian population (Fasa Persian Cohort Study)<br>PMID 33028906 | Grace Kim: No saffron | Alexander Tai Nguyen: Not isolated | – |
| 1175. Liu 2020. High follicle-stimulating hormone levels accelerate cartilage damage of knee osteoarthritis in postmenopausal women through the PI3K/AKT/NF-$\kappa$B pathway<br>PMID 32911565 | Grace Kim: No saffron | Alexander Tai Nguyen: No saffron | – |

Table S2 continued

| Paper | Reviewer 1 | Reviewer 2 | Additional recorded reviews |
|---|---|---|---|
| 1176. Alix 2020. Description of the unusual digestive tract of Platax orbicularis and the potential impact of Tenacibaculum maritimum infection<br>PMID 33024633 | Grace Kim: No saffron | Alexander Tai Nguyen: No saffron | – |
| 1177. Do 2020. Tetravalent Bispecific Tandem Antibodies Improve Brain Exposure and Efficacy in an Amyloid Transgenic Mouse Model<br>PMID 33005703 | Grace Kim: No saffron | Alexander Tai Nguyen: No saffron | – |
| 1178. Krishna 2020. Aqueous extract of Tamarindus indica fruit pulp exhibits antihyperglycaemic activity<br>PMID 32995322 | Grace Kim: No saffron | Alexander Tai Nguyen: No saffron | – |
| 1179. Mercader-Barceló 2020. Insights into the Role of Bioactive Food Ingredients and the Microbiome in Idiopathic Pulmonary Fibrosis<br>PMID 32842664 | Grace Kim: No saffron | Alexander Tai Nguyen: No saffron | – |
| 1180. Saraiva 2020. Natural Sweeteners: The Relevance of Food Naturalness for Consumers, Food Security Aspects, Sustainability and Health Impacts<br>PMID 32872325 | Grace Kim: No saffron | Alexander Tai Nguyen: No saffron | – |
| 1181. Ahmad 2021. An updated knowledge of Black seed (Nigella sativa Linn.): Review of phytochemical constituents and pharmacological properties<br>PMID 32983848 | Grace Kim: No saffron | Alexander Tai Nguyen: No saffron | – |
| 1182. Irmisch 2020. Complete Biosynthesis of the Anti-Diabetic Plant Metabolite Montbretin A<br>PMID 32647038 | Grace Kim: No saffron | Alexander Tai Nguyen: No saffron | – |
| 1183. Beccaro 2020. Castanea spp. Agrobiodiversity Conservation: Genotype Influence on Chemical and Sensorial Traits of Cultivars Grown on the Same Clonal Rootstock<br>PMID 32764341 | Grace Kim: No saffron | Alexander Tai Nguyen: No saffron | – |
| 1184. Mendonca 2020. Flavonoids Activation of the Transcription Factor Nrf2 as a Hypothesis Approach for the Prevention and Modulation of SARS-CoV-2 Infection Severity<br>PMID 32722164 | Grace Kim: No saffron | Alexander Tai Nguyen: No saffron | – |
| 1185. El-Sisi 2020. The potential beneficial effects of sildenafil and diosmin in experimentally-induced gastric ulcer in rats<br>PMID 32885082 | Grace Kim: No saffron | Alexander Tai Nguyen: No saffron | – |
| 1186. Khalid 2020. Type of training has a significant influence on the GH/IGF-1 axis but not on regulating miRNAs<br>PMID 32879543 | Grace Kim: No saffron | Alexander Tai Nguyen: No saffron | – |

Table S2 continued

| Paper | Reviewer 1 | Reviewer 2 | Additional recorded reviews |
|---|---|---|---|
| 1187. Lee 2020. Suppressing breast cancer by exercise: consideration to animal models and exercise protocols PMID 32698258 | Grace Kim: No saffron | Alexander Tai Nguyen: No saffron | – |
| 1188. Hossen 2020. A Review on Current Repurposing Drugs for the Treatment of COVID-19: Reality and Challenges PMID 32904710 | Grace Kim: No saffron | Alexander Tai Nguyen: No saffron | – |
| 1189. Qin 2020. Analysis of the analgesic effects of tricyclic antidepressants on neuropathic pain, diabetic neuropathic pain, and fibromyalgia in rat models PMID 32874123 | Grace Kim: No saffron | Alexander Tai Nguyen: No saffron | – |
| 1190. Samaddar 2020. The Enigma of Low COVID-19 Fatality Rate in India PMID 32849833 | Grace Kim: No saffron | Alexander Tai Nguyen: No saffron | – |
| 1191. Barbosa 2020. Repeated Administration of Clinical Doses of Tramadol and Tapentadol Causes Hepato- and Nephrotoxic Effects in Wistar Rats PMID 32664348 | Grace Kim: No saffron | Alexander Tai Nguyen: No saffron | – |
| 1192. Huang 2020. Mitochondrial Dysfunction as a Novel Target for Neuroprotective Nutraceuticals in Ocular Diseases PMID 32629966 | Grace Kim: Not clinical | Alexander Tai Nguyen: No saffron | – |
| 1193. Hu 2020. Regulation effects of total flavonoids in Morus alba L. on hepatic cholesterol disorders in orotic acid induced NAFLD rats PMID 32807146 | Grace Kim: No saffron | Alexander Tai Nguyen: No saffron | – |
| 1194. Kattar 2020. Metformin and Probiotics in the Crosstalk between Colitis-Associated Colorectal Cancer and Diabetes in Mice PMID 32664279 | Grace Kim: No saffron | Alexander Tai Nguyen: No saffron | – |
| 1195. Thivolet 2020. Spectral photon-counting CT imaging of colorectal peritoneal metastases: initial experience in rats PMID 32770125 | Grace Kim: No saffron | Alexander Tai Nguyen: No saffron | – |
| 1196. Cheema 2020. Integrated Datasets of Proteomic and Metabolomic Biomarkers to Predict Its Impacts on Comorbidities of Type 2 Diabetes Mellitus PMID 32753925 | Grace Kim: No saffron | Alexander Tai Nguyen: No saffron | – |
| 1197. Rameshrad 2020. An overview of glucagon-like peptide-1 receptor agonists for the treatment of metabolic syndrome: A drug repositioning PMID 32742592 | Grace Kim: No saffron | Alexander Tai Nguyen: No saffron | – |
| 1198. Makkar 2020. Nutraceuticals in Neurological Disorders PMID 32580329 | Grace Kim: Not clinical | Alexander Tai Nguyen: No saffron | – |

Table S2 continued

| Paper | Reviewer 1 | Reviewer 2 | Additional recorded reviews |
|---|---|---|---|
| 1199. Pérez-Gálvez 2020. Carotenoids and Chlorophylls as Antioxidants<br>PMID 32526968 | Grace Kim: No saffron | Alexander Tai Nguyen: No saffron | – |
| 1200. Dragoş 2020. Mechanisms of Herbal Nephroprotection in diabetes mellitus<br>PMID 32695828 | Grace Kim: Not clinical | Alexander Tai Nguyen: No saffron | – |
| 1201. Xie 2020. Attenuation of Diabetic Nephropathy in Diabetic Mice by Fasudil through Regulation of Macrophage Polarization<br>PMID 32685556 | Grace Kim: No saffron | Alexander Tai Nguyen: No saffron | – |
| 1202. De Caris 2020. Blueberry Counteracts BV-2 Microglia Morphological and Functional Switch after LPS Challenge<br>PMID 32575571 | Grace Kim: No saffron | Alexander Tai Nguyen: No saffron | – |
| 1203. Caradonna 2020. Science and Healthy Meals in the World: Nutritional Epigenomics and Nutrigenetics of the Mediterranean Diet<br>PMID 32545252 | Grace Kim: No saffron | Alexander Tai Nguyen: No saffron | – |
| 1204. Margină 2020. Analysis of the intricate effects of polyunsaturated fatty acids and polyphenols on inflammatory pathways in health and disease<br>PMID 32640331 | Grace Kim: No saffron | Alexander Tai Nguyen: No saffron | – |
| 1205. Saini 2020. Movement Disorders in Inherited Metabolic Diseases in Children<br>PMID 32606521 | Grace Kim: No saffron | Alexander Tai Nguyen: No saffron | – |
| 1206. Rayego-Mateos 2020. Pathogenic Pathways and Therapeutic Approaches Targeting Inflammation in Diabetic Nephropathy<br>PMID 32471207 | Grace Kim: No saffron | Alexander Tai Nguyen: No saffron | – |
| 1207. Fang 2020. Crocin Improves Insulin Sensitivity and Ameliorates Adiposity by Regulating AMPK-CDK5-PPAR$\gamma$ Signaling<br>PMID 32596392 | Grace Kim: Not clinical | Alexander Tai Nguyen: Not clinical | – |
| 1208. Noshahr 2020. Animal Models of Diabetes-Associated Renal Injury<br>PMID 32566684 | Grace Kim: No saffron | Alexander Tai Nguyen: No saffron | – |
| 1209. Wojtunik-Kulesza 2020. Influence of In Vitro Digestion on Composition, Bioaccessibility and Antioxidant Activity of Food Polyphenols-A Non-Systematic Review<br>PMID 32414132 | Grace Kim: No saffron | Alexander Tai Nguyen: No saffron | – |

Table S2 continued

| Paper | Reviewer 1 | Reviewer 2 | Additional recorded reviews |
|---|---|---|---|
| 1210. Sorour 2020. Visual and anatomic outcomes of sustained single agent anti-VEGF treatment versus double anti-VEGF switching in the treatment of persistent diabetic macular edema<br>PMID 32523776 | Grace Kim: No saffron | Alexander Tai Nguyen: No saffron | – |
| 1211. Ntandja Wandji 2020. Combined alcoholic and non-alcoholic steatohepatitis<br>PMID 32514497 | Grace Kim: No saffron | Alexander Tai Nguyen: No saffron | – |
| 1212. Nouri 2020. Nanophytomedicines for the Prevention of Metabolic Syndrome: A Pharmacological and Biopharmaceutical Review<br>PMID 32478050 | Grace Kim: No saffron | Alexander Tai Nguyen: No saffron | – |
| 1213. Chokki 2020. Exploring Antioxidant and Enzymes (A-Amylase and B-Glucosidase) Inhibitory Activity of Morinda lucida and Momordica charantia Leaves from Benin<br>PMID 32260400 | Grace Kim: No saffron | Alexander Tai Nguyen: No saffron | – |
| 1214. Samarghandian 2019. Antidotal or protective effects of honey and one of its major polyphenols, chrysin, against natural and chemical toxicities<br>PMID 31910181 | Grace Kim: No saffron | Alexander Tai Nguyen: No saffron | – |
| 1215. Tousian 2020. Looking for immortality: Review of phytotherapy for stem cell senescence<br>PMID 32405357 | Grace Kim: No saffron | Alexander Tai Nguyen: No saffron | – |
| 1216. Chen 2020. Increased the risk of heart failure and comorbidities in patients with gout treatment: a population-based cohort study<br>PMID 32395506 | Grace Kim: No saffron | Alexander Tai Nguyen: No saffron | – |
| 1217. Esmaily 2018. A Comparison between Decision Tree and Random Forest in Determining the Risk Factors Associated with Type 2 Diabetes<br>PMC7204421 | Grace Kim: No saffron | Alexander Tai Nguyen: No saffron | – |
| 1218. Maitra 2019. Using Drosophila as a platform for drug discovery from natural products in Parkinson's disease<br>PMID 31303984 | Grace Kim: No saffron | Alexander Tai Nguyen: No saffron | – |
| 1219. Sharifi-Rad 2020. Diet, Lifestyle and Cardiovascular Diseases: Linking Pathophysiology to Cardioprotective Effects of Natural Bioactive Compounds<br>PMID 32235611 | Grace Kim: No saffron | Alexander Tai Nguyen: No saffron | – |
| 1220. Rahman 2020. Novel Drug Delivery Systems for Loading of Natural Plant Extracts and Their Biomedical Applications<br>PMID 32346289 | Grace Kim: No saffron | Alexander Tai Nguyen: No saffron | – |

Table S2 continued

| Paper | Reviewer 1 | Reviewer 2 | Additional recorded reviews |
|---|---|---|---|
| 1221. Tan 2020. Effect of pioglitazone on skeletal muscle lipid deposition in the insulin resistance rat model induced by high fructose diet under AMPK signaling pathway<br>PMID 32346341 | Grace Kim: No saffron | Alexander Tai Nguyen: No saffron | – |
| 1222. Shawky 2019. The Biological Impacts of Sitagliptin on the Pancreas of a Rat Model of Type 2 Diabetes Mellitus: Drug Interactions with Metformin<br>PMID 31881657 | Grace Kim: No saffron | Alexander Tai Nguyen: No saffron | – |
| 1223. Shaito 2020. Herbal Medicine for Cardiovascular Diseases: Efficacy, Mechanisms, and Safety<br>PMID 32317975 | Grace Kim: No saffron | Alexander Tai Nguyen: No saffron | – |
| 1224. Author not recorded 2017. Diseases of the Alimentary Tract: Nonruminant<br>PMC7167529 | Grace Kim: No saffron | Alexander Tai Nguyen: No saffron | – |
| 1225. Sivakumar 2010. Dose-dependent effect of galangin on fructose-mediated insulin resistance and oxidative events in rat kidney<br>PMID 21062538 | Grace Kim: No saffron | Alexander Tai Nguyen: No saffron | – |
| 1226. Thompson 2014. Systemic Approach to Differential Diagnosis<br>PMC7156016 | Grace Kim: No saffron | Alexander Tai Nguyen: No saffron | – |
| 1227. Duarte 2020. Phytopharmacological Strategies in the Management of Type 2 Diabetes Mellitus<br>PMID 32131470 | Grace Kim: No saffron | Alexander Tai Nguyen: No saffron | – |
| 1228. Antunes 2020. Effective Synergy of Sorafenib and Nutrient Shortage in Inducing Melanoma Cell Death through Energy Stress<br>PMID 32155825 | Grace Kim: No saffron | Alexander Tai Nguyen: No saffron | – |
| 1229. Lin 2020. Astragaloside IV promotes the eNOS/NO/cGMP pathway and improves left ventricular diastolic function in rats with metabolic syndrome<br>PMID 30843445 | Grace Kim: No saffron | Alexander Tai Nguyen: No saffron | – |
| 1230. Lackie 2013. C<br>PMC7149705 | Grace Kim: No saffron | Alexander Tai Nguyen: No saffron | – |
| 1231. Wynn 2007. Veterinary Herbal Medicine: A Systems-Based Approach<br>PMC7151902 | Grace Kim: No saffron | Alexander Tai Nguyen: No saffron | – |
| 1232. Dewanjee 2020. Plant-Based Antidiabetic Nanoformulations: The Emerging Paradigm for Effective Therapy<br>PMID 32210082 | Grace Kim: No saffron | Alexander Tai Nguyen: No saffron | – |

Table S2 continued

| Paper | Reviewer 1 | Reviewer 2 | Additional recorded reviews |
|---|---|---|---|
| 1233. Alamgir 2018. Biotechnology, In Vitro Production of Natural Bioactive Compounds, Herbal Preparation, and Disease Management (Treatment and Prevention) PMC7123938 | Grace Kim: No saffron | Alexander Tai Nguyen: No saffron | – |
| 1234. Lee 2020. Protective Effect of Hydroxysafflor Yellow A on Nephropathy by Attenuating Oxidative Stress and Inhibiting Apoptosis in Induced Type 2 Diabetes in Rat PMID 32256962 | Grace Kim: No saffron | Alexander Tai Nguyen: No saffron | – |
| 1235. Turner-McGrievy 2019. Impact of a 12-month Inflammation Management Intervention on the Dietary Inflammatory Index, inflammation, and lipids PMID 30904228 | Grace Kim: No saffron | Alexander Tai Nguyen: No saffron | – |
| 1236. Fajri 2020. Protective effects of Equisetum arvense methanolic extract on sperm characteristics and in vitro fertilization potential in experimental diabetic mice: An experimental study PMID 32259003 | Grace Kim: No saffron | Alexander Tai Nguyen: No saffron | – |
| 1237. Bukhari 2020. Cardioprotective Effect of Olive Oil Against Ischemia Reperfusion-induced Cardiac Arrhythmia in Isolated Diabetic Rat Heart PMID 32231891 | Grace Kim: No saffron | Alexander Tai Nguyen: No saffron | – |
| 1238. Yaribeygi 2020. Molecular Mechanisms Linking Oxidative Stress and Diabetes Mellitus PMID 32215179 | Grace Kim: No saffron | Alexander Tai Nguyen: No saffron | – |
| 1239. Author not recorded 2019. Abstracts from the 51(st) European Society of Human Genetics Conference: Posters PMC6778823 | Grace Kim: No saffron | Alexander Tai Nguyen: No saffron | – |
| 1240. Ghorabi 2020. Association between dietary inflammatory index and components of metabolic syndrome PMID 32211135 | Grace Kim: No saffron | Alexander Tai Nguyen: Not isolated | – |
| 1241. Igbokwe 2019. Aluminium toxicosis: a review of toxic actions and effects PMID 32206026 | Grace Kim: No saffron | Alexander Tai Nguyen: No saffron | – |
| 1242. Rahimlou 2019. Effects of ginger supplementation on anthropometric, glycemic and metabolic parameters in subjects with metabolic syndrome: A randomized, double-blind, placebo-controlled study PMID 31275882 | Grace Kim: No saffron | Alexander Tai Nguyen: No saffron | – |
| 1243. Watanabe 2020. Effect of Dose and Timing of Burdock (Arctium lappa) Root Intake on Intestinal Microbiota of Mice PMID 32041173 | Grace Kim: No saffron | Alexander Tai Nguyen: No saffron | – |

Table S2 continued

| Paper | Reviewer 1 | Reviewer 2 | Additional recorded reviews |
|---|---|---|---|
| 1244. Bulboaca 2020. The Effect of Nano-Epigallocatechin-Gallate on Oxidative Stress and Matrix Metalloproteinases in Experimental Diabetes Mellitus<br>PMID 32093214 | Grace Kim: No saffron | Alexander Tai Nguyen: No saffron | – |
| 1245. Rzajew 2020. Calcium-Involved Action of Phytochemicals: Carotenoids and Monoterpenes in the Brain<br>PMID 32093213 | Grace Kim: Not clinical | Alexander Tai Nguyen: Not clinical | – |
| 1246. Kwasek 2020. Nutritional programming improves dietary plant protein utilization in zebrafish Danio rerio<br>PMID 32142555 | Grace Kim: No saffron | Alexander Tai Nguyen: No saffron | – |
| 1247. Margaritis 2020. Effect of crocin on antioxidant gene expression, fibrinolytic parameters, redox status and blood biochemistry in nicotinamide-streptozotocin-induced diabetic rats<br>PMID 32161725 | Grace Kim: Not clinical | Alexander Tai Nguyen: Not clinical | – |
| 1248. Ovadia 2020. Ursodeoxycholic acid enriches intestinal bile salt hydrolase-expressing Bacteroidetes in cholestatic pregnancy<br>PMID 32127609 | Grace Kim: No saffron | Alexander Tai Nguyen: No saffron | – |
| 1249. Remigante 2019. Natural Antioxidants Beneficial Effects on Anion Exchange through Band 3 Protein in Human Erythrocytes<br>PMID 31888111 | Grace Kim: No saffron | Alexander Tai Nguyen: No saffron | – |
| 1250. Unuofin 2020. Antioxidant Effects and Mechanisms of Medicinal Plants and Their Bioactive Compounds for the Prevention and Treatment of Type 2 Diabetes: An Updated Review<br>PMID 32148647 | Grace Kim: No saffron | Alexander Tai Nguyen: No saffron | – |
| 1251. Bresteau 2018. Methylprednisolone-induced acute liver injury in a patient treated for multiple sclerosis relapse<br>PMID 29507031 | Grace Kim: No saffron | Alexander Tai Nguyen: No saffron | – |
| 1252. Weiskirchen 2020. All You Can Feed: Some Comments on Production of Mouse Diets Used in Biomedical Research with Special Emphasis on Non-Alcoholic Fatty Liver Disease Research<br>PMID 31936026 | Grace Kim: No saffron | Alexander Tai Nguyen: No saffron | – |
| 1253. Lawal 2019. Comparative effects of glibenclamide, metformin and insulin on fetal pancreatic histology and maternal blood glucose in pregnant streptozotocin-induced diabetic rats<br>PMID 32127822 | Grace Kim: No saffron | Alexander Tai Nguyen: No saffron | – |

Table S2 continued

| Paper | Reviewer 1 | Reviewer 2 | Additional recorded reviews |
|---|---|---|---|
| 1254. Aubry 2019. A diet enriched with curcumin promotes resilience to chronic social defeat stress<br>PMID 30542090 | Grace Kim: No saffron | Alexander Tai Nguyen: No saffron | – |
| 1255. Cárdenas-Tueme 2020. Neurodegenerative Susceptibility During Maternal Nutritional Programing: Are Central and Peripheral Innate Immune Training Relevant?<br>PMID 32116490 | Grace Kim: No saffron | Alexander Tai Nguyen: No saffron | – |
| 1256. Eid 2020. Public Knowledge, Attitude, and Practice on Herbal Remedies Used During Pregnancy and Lactation in West Bank Palestine<br>PMID 32116721 | Grace Kim: Not clinical | Alexander Tai Nguyen: Not clinical | – |
| 1257. Soetikno 2020. Alpha-Mangostin Improves Cardiac Hypertrophy and Fibrosis and Associated Biochemical Parameters in High-Fat/High-Glucose Diet and Low-Dose Streptozotocin Injection-Induced Type 2 Diabetic Rats<br>PMID 32095085 | Grace Kim: No saffron | Alexander Tai Nguyen: No saffron | – |
| 1258. Almodóvar 2020. Bioaccessibility and Pharmacokinetics of a Commercial Saffron (Crocus sativus L.) Extract<br>PMID 32089715 | Grace Kim: Not clinical | Alexander Tai Nguyen: No saffron | – |
| 1259. Yang 2020. Dietary Inflammatory Index Positively Associated With High-Sensitivity C-Reactive Protein Level in Japanese From NIPPON DATA2010<br>PMID 30745493 | Grace Kim: No saffron | Alexander Tai Nguyen: No saffron | – |
| 1260. Fei 2020. Endotoxin Producers Overgrowing in Human Gut Microbiota as the Causative Agents for Nonalcoholic Fatty Liver Disease<br>PMID 32019793 | Grace Kim: No saffron | Alexander Tai Nguyen: No saffron | – |
| 1261. Samimi 2019. Possible antioxidant mechanism of coenzyme Q10 in diabetes: impact on Sirt1/Nrf2 signaling pathways<br>PMID 32038732 | Grace Kim: No saffron | Alexander Tai Nguyen: No saffron | – |
| 1262. Russo 2020. Lycopene and bone: an in vitro investigation and a pilot prospective clinical study<br>PMID 31996227 | Grace Kim: No saffron | Alexander Tai Nguyen: No saffron | – |
| 1263. Neto 2020. Analysis of the effectiveness of cinnamon (Cinnamomum verum) in the reduction of glycemic and lipidic levels of adults with type 2 diabetes: A study protocol<br>PMID 31895796 | Grace Kim: No saffron | Alexander Tai Nguyen: No saffron | – |
| 1264. Author not recorded 2019. Abstracts of the 3rd Sechenov International Biomedical Summit<br>PMC6985892 | Grace Kim: Not clinical | Alexander Tai Nguyen: Not clinical | – |

Table S2 continued

| Paper | Reviewer 1 | Reviewer 2 | Additional recorded reviews |
|---|---|---|---|
| 1265. Dai 2020. Potential of circulating pro-angiogenic microRNA expressions as biomarkers for rapid angiographic stenotic progression and restenosis risks in coronary artery disease patients underwent percutaneous coronary intervention PMID 31495986 | Grace Kim: No saffron | Alexander Tai Nguyen: No saffron | – |
| 1266. AlMotwaa 2020. Incorporating ifosfamide into salvia oil-based nanoemulsion diminishes its nephrotoxicity in mice inoculated with tumor PMID 31988852 | Grace Kim: No saffron | Alexander Tai Nguyen: No saffron | – |
| 1267. Rameshrad 2019. Pharmacology of dipeptidyl peptidase-4 inhibitors and its use in the management of metabolic syndrome: a comprehensive review on drug repositioning PMID 30674032 | Grace Kim: No saffron | Alexander Tai Nguyen: No saffron | – |
| 1268. Wirth 2020. Impact of a 3-Month Anti-inflammatory Dietary Intervention Focusing on Watermelon on Body Habitus, Inflammation, and Metabolic Markers: A Pilot Study PMID 31975781 | Grace Kim: No saffron | Alexander Tai Nguyen: No saffron | – |
| 1269. Dernoncourt 2019. Myogenic Disease and Metabolic Acidosis: Consider Multiple Acyl-Coenzyme A Dehydrogenase Deficiency PMID 31934457 | Grace Kim: No saffron | Alexander Tai Nguyen: No saffron | – |
| 1270. Zych 2019. Effect of Rosmarinic Acid and Sinapic Acid on Oxidative Stress Parameters in the Cardiac Tissue and Serum of Type 2 Diabetic Female Rats PMID 31771099 | Grace Kim: No saffron | Alexander Tai Nguyen: No saffron | – |
| 1271. Liu 2019. New Insights for Cellular and Molecular Mechanisms of Aging and Aging-Related Diseases: Herbal Medicine as Potential Therapeutic Approach PMID 31915506 | Grace Kim: No saffron | Alexander Tai Nguyen: No saffron | – |
| 1272. Silveira 2019. The Action of Polyphenols in Diabetes Mellitus and Alzheimer's Disease: A Common Agent for Overlapping Pathologies PMID 30081787 | Grace Kim: No saffron | Alexander Tai Nguyen: No saffron | – |
| 1273. Liu 2019. The Bioprotective Effects of Polyphenols on Metabolic Syndrome against Oxidative Stress: Evidences and Perspectives PMID 31885810 | Grace Kim: No saffron | Alexander Tai Nguyen: No saffron | – |

Table S2 continued

| Paper | Reviewer 1 | Reviewer 2 | Additional recorded reviews |
|---|---|---|---|
| 1274. Wang 2019. Simultaneous Use of Stimulatory Agents to Enhance the Production and Hypoglycaemic Activity of Polysaccharides from Inonotus obliquus by Submerged Fermentation<br>PMID 31810277 | Grace Kim: No saffron | Alexander Tai Nguyen: No saffron | – |
| 1275. Schilperoort 2020. Disruption of circadian rhythm by alternating light-dark cycles aggravates atherosclerosis development in APOE*3-Leiden.CETP mice<br>PMID 31599473 | Grace Kim: No saffron | Alexander Tai Nguyen: No saffron | – |
| 1276. Irmisch 2019. Biosynthesis of the anti-diabetic metabolite montbretin A: glucosylation of the central intermediate mini-MbA<br>PMID 31400245 | Grace Kim: No saffron | Alexander Tai Nguyen: No saffron | – |
| 1277. Li 2019. Therapeutic effects of moxibustion simultaneously targeting Nrf2 and NF-$\kappa$B in diabetic peripheral neuropathy<br>PMID 31209719 | Grace Kim: No saffron | Alexander Tai Nguyen: No saffron | – |
| 1278. Pereira 2019. Evaluation of the Anti-Diabetic Activity of Some Common Herbs and Spices: Providing New Insights with Inverse Virtual Screening<br>PMID 31703341 | Grace Kim: Not clinical | Alexander Tai Nguyen: No saffron | – |
| 1279. Arshad 2019. Tamarind: A diet-based strategy against lifestyle maladies<br>PMID 31762991 | Grace Kim: No saffron | Alexander Tai Nguyen: No saffron | – |
| 1280. Al-Attar 2019. Influence of olive leaves extract on hepatorenal injury in streptozotocin diabetic rats<br>PMID 31762669 | Grace Kim: No saffron | Alexander Tai Nguyen: No saffron | – |
| 1281. Elsherbiny 2019. Carbamazepine Alleviates Retinal and Optic Nerve Neural Degeneration in Diabetic Mice via Nerve Growth Factor-Induced PI3K/Akt/mTOR Activation<br>PMID 31736682 | Grace Kim: No saffron | Alexander Tai Nguyen: No saffron | – |
| 1282. Lende 2019. Influence of pre-operative oral carbohydrate loading vs. standard fasting on tumor proliferation and clinical outcome in breast cancer patients – a randomized trial<br>PMID 31703648 | Grace Kim: No saffron | Alexander Tai Nguyen: No saffron | – |
| 1283. Mickle 2019. The Dietary Inflammatory Index® and Alternative Healthy Eating Index 2010 in relation to leucocyte telomere length in postmenopausal women: a cross-sectional study<br>PMID 31723429 | Grace Kim: No saffron | Alexander Tai Nguyen: No saffron | – |

Table S2 continued

| Paper | Reviewer 1 | Reviewer 2 | Additional recorded reviews |
|---|---|---|---|
| 1284. Helal 2019. Nutraceuticals' Novel Formulations: The Good, the Bad, the Unknown and Patents Involved<br>PMID 31577201 | Grace Kim: Not clinical | Alexander Tai Nguyen: No saffron | – |
| 1285. Dlamini 2019. Regulation of alternative splicing in obesity-induced hypertension<br>PMID 31695458 | Grace Kim: No saffron | Alexander Tai Nguyen: No saffron | – |
| 1286. Elbassuoni 2019. Impact of chronic exercise on counteracting chronic stress-induced functional and morphological pancreatic changes in male albino rats<br>PMID 30903523 | Grace Kim: No saffron | Alexander Tai Nguyen: No saffron | – |
| 1287. Hatziagapiou 2018. The Protective Role of Crocus Sativus L. (Saffron) Against Ischemia- Reperfusion Injury, Hyperlipidemia and Atherosclerosis: Nature Opposing Cardiovascular Diseases<br>PMID 29952263 | Grace Kim: Not clinical | Alexander Tai Nguyen: No saffron | – |
| 1288. Faran 2019. Hepatoprotective and Renoprotective Properties of Lovastatin-Loaded Ginger and Garlic Oil Nanoemulsomes: Insights into Serum Biological Parameters<br>PMID 31505863 | Grace Kim: No saffron | Alexander Tai Nguyen: No saffron | – |
| 1289. Stuby 2019. Appetite-Suppressing and Satiety-Increasing Bioactive Phytochemicals: A Systematic Review<br>PMID 31533291 | Grace Kim: Not clinical | Alexander Tai Nguyen: No saffron | – |
| 1290. Author not recorded 2018. UEG Week 2018 Poster Presentations<br>PMC6796246 | Grace Kim: No saffron | Alexander Tai Nguyen: No saffron | – |
| 1291. Naz 2019. Enzyme Inhibitory, Antioxidant And Antibacterial Potentials Of Synthetic Symmetrical And Unsymmetrical Thioureas<br>PMID 31631973 | Grace Kim: No saffron | Alexander Tai Nguyen: No saffron | – |
| 1292. Azizi 2019. Investigating the effect of Crocus sativus L. petal hydroalcoholic extract on inflammatory and enzymatic indices resulting from alcohol use in kidney and liver of male rats<br>PMID 31632125 | Grace Kim: Not clinical | Alexander Tai Nguyen: No saffron | – |
| 1293. Gu 2019. The IRE1$\alpha$ Arm of UPR Regulates Muscle Cells Immune Characters by Restraining p38 MAPK Activation<br>PMID 31607947 | Grace Kim: No saffron | Alexander Tai Nguyen: No saffron | – |

Table S2 continued

| Paper | Reviewer 1 | Reviewer 2 | Additional recorded reviews |
|---|---|---|---|
| 1294. Yang 2019. Design and optimization of crocetin loaded PLGA nanoparticles against diabetic nephropathy via suppression of inflammatory biomarkers: a formulation approach to preclinical study<br>PMID 31524015 | Grace Kim: Not clinical | Alexander Tai Nguyen: No saffron | – |
| 1295. Fan 2019. The Role of Momordica charantia in Resisting Obesity<br>PMID 31487939 | Grace Kim: No saffron | Alexander Tai Nguyen: No saffron | – |
| 1296. Di Sotto 2019. Hypoglycemic, Antiglycation, and Cytoprotective Properties of a Phenol-Rich Extract From Waste Peel of Punica granatum L. var. Dente di Cavallo DC2<br>PMID 31461832 | Grace Kim: No saffron | Alexander Tai Nguyen: No saffron | – |
| 1297. Puspitasari 2019. Broccoli sprouts juice prevents lens protein aggregation in streptozotocin-induced diabetic rat<br>PMID 31544030 | Grace Kim: No saffron | Alexander Tai Nguyen: No saffron | – |
| 1298. Lin 2019. Crocin Improves Cognitive Behavior in Rats with Alzheimer's Disease by Regulating Endoplasmic Reticulum Stress and Apoptosis<br>PMID 31534969 | Grace Kim: Not clinical | Alexander Tai Nguyen: No saffron | – |
| 1299. Shahbazian 2019. Effects of saffron on homocysteine, and antioxidant and inflammatory biomarkers levels in patients with type 2 diabetes mellitus: a randomized double-blind clinical trial<br>PMID 31516857 | Grace Kim: Candidate | Alexander Tai Nguyen: Candidate | – |
| 1300. Koh 2019. Anti-obesity effect of Yangkyuksanwha-tang in high-fat diet-induced obese mice<br>PMID 31488172 | Grace Kim: No saffron | Alexander Tai Nguyen: No saffron | – |
| 1301. Lankatillake 2019. Understanding glycaemic control and current approaches for screening antidiabetic natural products from evidence-based medicinal plants<br>PMID 31516543 | Grace Kim: No saffron | Alexander Tai Nguyen: No saffron | – |
| 1302. Fontes 2019. Antioxidant Versus Pro-Apoptotic Effects of Mushroom-Enriched Diets on Mitochondria in Liver Disease<br>PMID 31426291 | Grace Kim: No saffron | Alexander Tai Nguyen: No saffron | – |
| 1303. Recinella 2019. Protective Effects Induced by Two Polyphenolic Liquid Complexes from Olive (Olea europaea, mainly Cultivar Coratina) Pressing Juice in Rat Isolated Tissues Challenged with LPS<br>PMID 31430921 | Grace Kim: No saffron | Alexander Tai Nguyen: No saffron | – |

Table S2 continued

| Paper | Reviewer 1 | Reviewer 2 | Additional recorded reviews |
|---|---|---|---|
| 1304. Jacobs 2019. Sex-Specific Differences in Fat Storage, Development of Non-Alcoholic Fatty Liver Disease and Brain Structure in Juvenile HFD-Induced Obese Ldlr-/-.Leiden Mice PMID 31405127 | Grace Kim: No saffron | Alexander Tai Nguyen: No saffron | – |
| 1305. Uchio 2019. Hot Water Extract of Curcuma longa L. Improves Serum Inflammatory Markers and General Health in Subjects with Overweight or Prehypertension/Mild Hypertension: A Randomized, Double-Blind, Placebo-Controlled Trial PMID 31394768 | Grace Kim: No saffron | Alexander Tai Nguyen: No saffron | – |
| 1306. Warrior 2019. Olive Oil Intake Associated with Increased Attention Scores in Women Living with HIV: Findings from the Chicago Women's Interagency HIV Study PMID 31370174 | Grace Kim: No saffron | Alexander Tai Nguyen: No saffron | – |
| 1307. Peddada 2015. Role of Curcumin in Common Musculoskeletal Disorders: a Review of Current Laboratory, Translational, and Clinical Data PMID 26311096 | Grace Kim: No saffron | Alexander Tai Nguyen: No saffron | – |
| 1308. Faraone 2019. Antioxidant, Antidiabetic, and Anticholinesterase Activities and Phytochemical Profile of Azorella glabra Wedd PMID 31382601 | Grace Kim: No saffron | Alexander Tai Nguyen: No saffron | – |
| 1309. Guglielmi 2019. Benzo[b]tiophen-3-ol derivatives as effective inhibitors of human monoamine oxidase: design, synthesis, and biological activity PMID 31422706 | Grace Kim: No saffron | Alexander Tai Nguyen: No saffron | – |
| 1310. Gidday 2018. Adaptive Plasticity in the Retina: Protection Against Acute Injury and Neurodegenerative Disease by Conditioning Stimuli PMID 31423482 | Grace Kim: No saffron | Alexander Tai Nguyen: No saffron | – |
| 1311. Ran 2019. Rhizoma coptidis as a Potential Treatment Agent for Type 2 Diabetes Mellitus and the Underlying Mechanisms: A Review PMID 31396083 | Grace Kim: No saffron | Alexander Tai Nguyen: No saffron | – |
| 1312. Barragán-Bonilla 2019. Combined Administration of Streptozotocin and Sucrose Accelerates the Appearance of Type 2 Diabetes Symptoms in Rats PMID 31355292 | Grace Kim: No saffron | Alexander Tai Nguyen: No saffron | – |
| 1313. Collins 2019. Estrogen Regulates the Satellite Cell Compartment in Females PMID 31291574 | Grace Kim: No saffron | Alexander Tai Nguyen: No saffron | – |

Table S2 continued

| Paper | Reviewer 1 | Reviewer 2 | Additional recorded reviews |
|---|---|---|---|
| 1314. Mahmoudi 2019. Effects of Pioglitazone On the Lipid Profile, Serum Antioxidant Capacity, and UCP1 Gene Expression in Mouse Brown Adipose Tissue<br>PMID 31334282 | Grace Kim: No saffron | Alexander Tai Nguyen: No saffron | – |
| 1315. Palozi 2019. Pharmacological safety of Plinia cauliflora (Mart.) Kausel in rabbits<br>PMID 31316897 | Grace Kim: No saffron | Alexander Tai Nguyen: No saffron | – |
| 1316. de Camargo 2019. Is Chickpea a Potential Substitute for Soybean? Phenolic Bioactives and Potential Health Benefits<br>PMID 31146372 | Grace Kim: No saffron | Alexander Tai Nguyen: No saffron | – |
| 1317. Sefidgar 2019. Effect of crocin on biochemical parameters, oxidative/antioxidative profiles, sperm characteristics and testicular histopathology in streptozotocin-induced diabetic rats<br>PMID 31309073 | Grace Kim: Not clinical | Alexander Tai Nguyen: No saffron | – |
| 1319. Varela-Rodríguez 2019. Biological and toxicological evaluation of Rhus trilobata Nutt. (Anacardiaceae) used traditionally in mexico against cancer<br>PMID 31262287 | Grace Kim: No saffron | Alexander Tai Nguyen: No saffron | – |
| 1320. Tewari 2019. Medicinal Plants and Natural Products Used in Cataract Management<br>PMID 31263410 | Grace Kim: No saffron | Alexander Tai Nguyen: No saffron | – |
| 1321. Hashemian 2019. Turmeric, Pepper, Cinnamon, and Saffron Consumption and Mortality (P18-069-19)<br>PMC6574417 | Grace Kim: No saffron | Alexander Tai Nguyen: No saffron | – |
| 1322. Dehghani 2019. The effects of Crataegus pinnatifida (Chinese hawthorn) on metabolic syndrome: A review<br>PMID 31217924 | Grace Kim: No saffron | Alexander Tai Nguyen: No saffron | – |
| 1323. Yimer 2019. Nigella sativa L. (Black Cumin): A Promising Natural Remedy for Wide Range of Illnesses<br>PMID 31214267 | Grace Kim: No saffron | Alexander Tai Nguyen: No saffron | – |
| 1324. Nagappan 2019. Role of Cannabinoid Receptor Type 1 in Insulin Resistance and Its Biological Implications<br>PMID 31035653 | Grace Kim: No saffron | Alexander Tai Nguyen: No saffron | – |
| 1325. Boroushaki 2019. Reno-protective effect of Rheum turkestanicum against gentamicin-induced nephrotoxicity<br>PMID 31156796 | Grace Kim: No saffron | Alexander Tai Nguyen: No saffron | – |
| 1326. Vázquez-Fresno 2019. Herbs and Spices- Biomarkers of Intake Based on Human Intervention Studies - A Systematic Review<br>PMID 31143299 | Grace Kim: Not clinical | Alexander Tai Nguyen: No saffron | – |

Table S2 continued

| Paper | Reviewer 1 | Reviewer 2 | Additional recorded reviews |
|---|---|---|---|
| 1327. Huang 2019. Dietary ginger as a traditional therapy for blood sugar control in patients with type 2 diabetes mellitus: A systematic review and meta-analysis<br>PMID 30921234 | Grace Kim: No saffron | Alexander Tai Nguyen: No saffron | – |
| 1328. Muñoz Balbontín 2019. Herbal Medicinal Product Use During Pregnancy and the Postnatal Period: A Systematic Review<br>PMID 30969204 | Grace Kim: No saffron | Alexander Tai Nguyen: No saffron | – |
| 1329. Broderick 2019. Anti-inflammatory and angiogenic effects of exercise training in cardiac muscle of diabetic mice<br>PMID 31118719 | Grace Kim: No saffron | Alexander Tai Nguyen: No saffron | – |
| 1330. Carvalho 2019. ALK2 inhibitors display beneficial effects in preclinical models of ACVR1 mutant diffuse intrinsic pontine glioma<br>PMID 31098401 | Grace Kim: No saffron | Alexander Tai Nguyen: No saffron | – |
| 1331. Zare 2018. Saffron (adjunct) for people with schizophrenia who have antipsychotic-induced metabolic syndrome<br>PMC6494189 | Grace Kim: Not clinical | Alexander Tai Nguyen: Not clinical | – |
| 1332. Nasim 2019. Relationship between Antioxidant Status and Attention Deficit Hyperactivity Disorder Among Children<br>PMID 31057726 | Grace Kim: No saffron | Alexander Tai Nguyen: No saffron | – |
| 1333. Wu 2019. Inhibition of P53/miR-34a improves diabetic endothelial dysfunction via activation of SIRT1<br>PMID 30793480 | Grace Kim: No saffron | Alexander Tai Nguyen: No saffron | – |
| 1334. Yadav 2018. Antidiabetic activity of mefloquine via GLP-1 receptor modulation against STZ-NA-induced diabetes in albino wistar rats<br>PMID 29744272 | Grace Kim: No saffron | Alexander Tai Nguyen: No saffron | – |
| 1335. Balogh 2019. Efficacy-Based Perspective to Overcome Reduced Opioid Analgesia of Advanced Painful Diabetic Neuropathy in Rats<br>PMID 31024314 | Grace Kim: No saffron | Alexander Tai Nguyen: No saffron | – |
| 1336. Ghyasi 2019. Combination effect of voluntary exercise and garlic (Allium sativum) on oxidative stress, cholesterol level and histopathology of heart tissue in type 1 diabetic rats<br>PMID 31024674 | Grace Kim: No saffron | Alexander Tai Nguyen: No saffron | – |
| 1337. Heitmar 2019. Saffron (Crocus sativus L.) in Ocular Diseases: A Narrative Review of the Existing Evidence from Clinical Studies<br>PMID 30889784 | Grace Kim: Not clinical | Alexander Tai Nguyen: No saffron | – |

Table S2 continued

| Paper | Reviewer 1 | Reviewer 2 | Additional recorded reviews |
|---|---|---|---|
| 1338. Lăcătuşu 2019. The Mediterranean Diet: From an Environment-Driven Food Culture to an Emerging Medical Prescription PMID 30875998 | Grace Kim: No saffron | Alexander Tai Nguyen: No saffron | – |
| 1339. Maiese 2018. The mechanistic target of rapamycin (mTOR) and the silent mating-type information regulation 2 homolog 1 (SIRT1): oversight for neurodegenerative disorders PMID 29523769 | Grace Kim: No saffron | Alexander Tai Nguyen: No saffron | – |
| 1340. Chen 2018. Carbohydrate response element binding protein (ChREBP) modulates the inflammatory response of mesangial cells in response to glucose PMID 30420491 | Grace Kim: No saffron | Alexander Tai Nguyen: No saffron | – |
| 1341. Zhang 2019. Lack of muscle mTOR kinase activity causes early onset myopathy and compromises whole-body homeostasis PMID 30461220 | Grace Kim: No saffron | Alexander Tai Nguyen: No saffron | – |
| 1342. Hardeland 2019. Aging, Melatonin, and the Pro- and Anti-Inflammatory Networks PMID 30862067 | Grace Kim: No saffron | Alexander Tai Nguyen: No saffron | – |
| 1343. Mahajan 2019. Perturbed Biochemical Pathways and Associated Oxidative Stress Lead to Vascular Dysfunctions in Diabetic Retinopathy PMID 30962865 | Grace Kim: No saffron | Alexander Tai Nguyen: No saffron | – |
| 1344. Pouwer 2019. The APOE(∗)3-Leiden Heterozygous Glucokinase Knockout Mouse as Novel Translational Disease Model for Type 2 Diabetes, Dyslipidemia, and Diabetic Atherosclerosis PMID 30949516 | Grace Kim: No saffron | Alexander Tai Nguyen: No saffron | – |
| 1345. Abdollahzadeh Estakhri 2019. Organ toxicity attenuation by nanomicelles containing curcuminoids: Comparing the protective effects on tissues oxidative damage induced by diazinon PMID 30944703 | Grace Kim: No saffron | Alexander Tai Nguyen: No saffron | – |
| 1346. Hirsh 2019. Avoiding holiday seasonal weight gain with nutrient-supported intermittent energy restriction: a pilot study PMID 30931109 | Grace Kim: No saffron | Alexander Tai Nguyen: No saffron | – |

Table S2 continued

| Paper | Reviewer 1 | Reviewer 2 | Additional recorded reviews |
|---|---|---|---|
| 1347. Daneshi-Maskooni 2019. Green cardamom supplementation improves serum irisin, glucose indices, and lipid profiles in overweight or obese non-alcoholic fatty liver disease patients: a double-blind randomized placebo-controlled clinical trial<br>PMID 30871514 | Grace Kim: No saffron | Alexander Tai Nguyen: No saffron | – |
| 1348. Zhao 2019. Inhibition of HDAC3 Ameliorates Cerebral Ischemia Reperfusion Injury in Diabetic Mice In Vivo and In Vitro<br>PMID 30906786 | Grace Kim: No saffron | Alexander Tai Nguyen: No saffron | – |
| 1349. Tayebi 2017. Exercise Training, Neuregulin 4 and Obesity<br>PMID 30899900 | Grace Kim: No saffron | Alexander Tai Nguyen: No saffron | – |
| 1350. Aday 2019. Targeting Residual Inflammatory Risk: A Shifting Paradigm for Atherosclerotic Disease<br>PMID 30873416 | Grace Kim: No saffron | Alexander Tai Nguyen: No saffron | – |
| 1351. Fernando 2019. Heat stability of the in vitro inhibitory effect of spices on lipase, amylase, and glucosidase enzymes<br>PMID 30847119 | Grace Kim: No saffron | Alexander Tai Nguyen: No saffron | – |
| 1352. Zhou 2019. Therapeutic FGF19 promotes HDL biogenesis and transhepatic cholesterol efflux to prevent atherosclerosis<br>PMID 30679232 | Grace Kim: No saffron | Alexander Tai Nguyen: No saffron | – |
| 1353. Zilaee 2019. An evaluation of the effects of saffron supplementation on the asthma clinical symptoms and asthma severity in patients with mild and moderate persistent allergic asthma: a double-blind, randomized placebo-controlled trial<br>PMID 30795753 | Grace Kim: Not clinical | Alexander Tai Nguyen: No outcome | – |
| 1354. Ramezani-Aliakbari 2019. The Effects of Trimetazidine on QT-interval Prolongation and Cardiac Hypertrophy in Diabetic Rats<br>PMID 30570065 | Grace Kim: No saffron | Alexander Tai Nguyen: No saffron | – |
| 1355. Ruiz-Ojeda 2019. Effects of Sweeteners on the Gut Microbiota: A Review of Experimental Studies and Clinical Trials<br>PMID 30721958 | Grace Kim: No saffron | Alexander Tai Nguyen: No saffron | – |
| 1356. Coué 2019. Spirulina Liquid Extract Protects against Fibrosis Related to Non-Alcoholic Steatohepatitis and Increases Ursodeoxycholic Acid<br>PMID 30669332 | Grace Kim: No saffron | Alexander Tai Nguyen: No saffron | – |

Table S2 continued

| Paper | Reviewer 1 | Reviewer 2 | Additional recorded reviews |
|---|---|---|---|
| 1357. Muhammad 2019. Dietary Inflammatory Index Score and Its Association with Body Weight, Blood Pressure, Lipid Profile, and Leptin in Indonesian Adults PMID 30641979 | Grace Kim: No saffron | Alexander Tai Nguyen: No saffron | – |
| 1358. Dasgeb 2018. Colchicine: an ancient drug with novel applications PMID 28832953 | Grace Kim: No saffron | Alexander Tai Nguyen: No saffron | – |
| 1359. Cojean 2018. Chondroblastic osteosarcoma of the middle ear in a guinea pig (Cavia porcellus) PMID 30104775 | Grace Kim: No saffron | Alexander Tai Nguyen: No saffron | – |
| 1360. Memon 2018. Methylglyoxal and insulin resistance in berberine-treated type 2 diabetic patients PMID 30693045 | Grace Kim: No saffron | Alexander Tai Nguyen: No saffron | – |
| 1361. Zhang 2015. Antioxidant Phytochemicals for the Prevention and Treatment of Chronic Diseases PMID 26633317 | Grace Kim: No saffron | Alexander Tai Nguyen: No saffron | – |
| 1362. Harty 2019. Nutritional and Supplementation Strategies to Prevent and Attenuate Exercise-Induced Muscle Damage: a Brief Review PMID 30617517 | Grace Kim: No saffron | Alexander Tai Nguyen: No saffron | – |
| 1363. Gart 2018. Diet-Independent Correlations between Bacteria and Dysfunction of Gut, Adipose Tissue, and Liver: A Comprehensive Microbiota Analysis in Feces and Mucosa of the Ileum and Colon in Obese Mice with NAFLD PMID 30577415 | Grace Kim: No saffron | Alexander Tai Nguyen: No saffron | – |
| 1364. Feng 2018. Effects of sleeve gastrectomy on lipid and energy metabolism in ZDF rats via PI3K/AKT pathway PMID 30662621 | Grace Kim: No saffron | Alexander Tai Nguyen: No saffron | – |
| 1365. Sriraksa 2019. Effect of Azadirachta indica flower extract on functional recovery of sciatic nerve crush injury in rat models of DM PMID 30651834 | Grace Kim: No saffron | Alexander Tai Nguyen: No saffron | – |
| 1366. Zengin 2018. Chromatographic Analyses, In Vitro Biological Activities, and Cytotoxicity of Cannabis sativa L. Essential Oil: A Multidisciplinary Study PMID 30544765 | Grace Kim: No saffron | Bryan Anh Nguyen: No saffron | Alexander Tai Nguyen: No saffron |
| 1367. Soccio 2018. Assessment of Antioxidant Capacity and Putative Healthy Effects of Natural Plant Products Using Soybean Lipoxygenase-Based Methods. An Overview PMID 30544620 | Grace Kim: No saffron | Bryan Anh Nguyen: No saffron | Alexander Tai Nguyen: No saffron |

Table S2 continued

| Paper | Reviewer 1 | Reviewer 2 | Additional recorded reviews |
|---|---|---|---|
| 1368. Ebrahim 2018. Mesenchymal Stem Cell-Derived Exosomes Ameliorated Diabetic Nephropathy by Autophagy Induction through the mTOR Signaling Pathway<br>PMID 30467302 | Grace Kim: No saffron | Bryan Anh Nguyen: No saffron | Alexander Tai Nguyen: No saffron |
| 1369. Stone 2018. Acquired Resilience: An Evolved System of Tissue Protection in Mammals<br>PMID 30627064 | Grace Kim: No saffron | Bryan Anh Nguyen: No saffron | – |
| 1370. Al-Attar 2019. Effect of Olea europaea leaves extract on streptozotocin induced diabetes in male albino rats<br>PMID 30622415 | Grace Kim: No saffron | Bryan Anh Nguyen: No saffron | – |
| 1371. Hua 2018. Regulatory Efficacy of Spirulina platensis Protease Hydrolyzate on Lipid Metabolism and Gut Microbiota in High-Fat Diet-Fed Rats<br>PMID 30551559 | Grace Kim: No saffron | Bryan Anh Nguyen: No saffron | – |
| 1372. Costea 2018. Chemoprevention of Colorectal Cancer by Dietary Compounds<br>PMID 30487390 | Grace Kim: Not clinical | Bryan Anh Nguyen: Not clinical | – |
| 1373. Bahmani 2016. Inhibitory Effect of Crocin(s) on Lens $\alpha$-Crystallin Glycation and Aggregation, Results in the Decrease of the Risk of Diabetic Cataract<br>PMID 26821002 | Grace Kim: Not clinical | Bryan Anh Nguyen: Not clinical | – |
| 1374. Lv 2018. Therapeutic Potential and Effective Components of the Chinese Herb Gardeniae Fructus in the Treatment of Senile Disease<br>PMID 30574425 | Grace Kim: Not clinical | Bryan Anh Nguyen: No saffron | – |
| 1375. de Camargo 2018. Opinion on the Hurdles and Potential Health Benefits in Value-Added Use of Plant Food Processing By-Products as Sources of Phenolic Compounds<br>PMID 30404239 | Grace Kim: No saffron | Bryan Anh Nguyen: No saffron | – |
| 1376. Shojaei Shad 2018. Study of the effect of the essential oil (extract) of rhubarb stem (shoot) on glycosylated hemoglobin and fasting blood glucose levels in patients with type II diabetes<br>PMID 30474605 | Grace Kim: No saffron | Bryan Anh Nguyen: No saffron | – |
| 1377. Morrison 2018. Obeticholic Acid Modulates Serum Metabolites and Gene Signatures Characteristic of Human NASH and Attenuates Inflammation and Fibrosis Progression in Ldlr-/-.Leiden Mice<br>PMID 30556039 | Grace Kim: No saffron | Bryan Anh Nguyen: No saffron | – |

Table S2 continued

| Paper | Reviewer 1 | Reviewer 2 | Additional recorded reviews |
|---|---|---|---|
| 1378. Catanzaro 2018. Immunomodulators Inspired by Nature: A Review on Curcumin and Echinacea<br>PMID 30373170 | Grace Kim: No saffron | Bryan Anh Nguyen: No saffron | – |
| 1379. Ahmad 2018. Protective effects of saffron against zearalenone-induced alterations in reproductive hormones in female mice (Mus musculus)<br>PMID 30538946 | Grace Kim: Not clinical | Bryan Anh Nguyen: Not clinical | – |
| 1380. Rad 2018. Mechanism involved in insulin resistance via accumulation of $\beta$-amyloid and neurofibrillary tangles: link between type 2 diabetes and Alzheimer's disease<br>PMID 30538427 | Grace Kim: No saffron | Bryan Anh Nguyen: No saffron | – |
| 1381. Basnet 2011. Curcumin: an anti-inflammatory molecule from a curry spice on the path to cancer treatment<br>PMID 21642934 | Grace Kim: No saffron | Bryan Anh Nguyen: No saffron | – |
| 1382. Zhang 2011. Astragaloside IV improves metabolic syndrome and endothelium dysfunction in fructose-fed rats<br>PMID 21555978 | Grace Kim: No saffron | Bryan Anh Nguyen: No saffron | – |
| 1383. Tsui 2018. Spices and Atherosclerosis<br>PMID 30423840 | Grace Kim: Not clinical | Bryan Anh Nguyen: No saffron | – |
| 1384. Leonov 2015. Longevity extension by phytochemicals<br>PMID 25871373 | Grace Kim: Not clinical | Bryan Anh Nguyen: No saffron | – |
| 1385. Alam 2018. Gossypol from Cottonseeds Ameliorates Glucose Uptake by Mimicking Insulin Signaling and Improves Glucose Homeostasis in Mice with Streptozotocin-Induced Diabetes<br>PMID 30510623 | Grace Kim: No saffron | Bryan Anh Nguyen: No saffron | – |
| 1386. Rivadeneyra-Domínguez 2018. Alterations of blood chemistry, hepatic and renal function, and blood cytometry in acrylamide-treated rats<br>PMID 30510905 | Grace Kim: No saffron | Bryan Anh Nguyen: No saffron | – |
| 1387. Marín 2018. De novo biosynthesis of myricetin, kaempferol and quercetin in Streptomyces albus and Streptomyces coelicolor<br>PMID 30440014 | Grace Kim: No saffron | Bryan Anh Nguyen: No saffron | – |
| 1388. Gicquel 2017. AAV-mediated transfer of FKRP shows therapeutic efficacy in a murine model but requires control of gene expression<br>PMID 28334834 | Grace Kim: No saffron | Bryan Anh Nguyen: No saffron | – |
| 1389. Hosseini 2018. Saffron (Crocus sativus) petal as a new pharmacological target: a review<br>PMID 30483381 | Grace Kim: Not clinical | Bryan Anh Nguyen: No saffron | – |

Table S2 continued

| Paper | Reviewer 1 | Reviewer 2 | Additional recorded reviews |
|---|---|---|---|
| 1390. Dastgerdi 2018. Effect of Crocin, Exercise, and Crocin-accompanied Exercise on Learning and Memory in Rats under Chronic Unpredictable Stress PMID 30464937 | Grace Kim: Not clinical | Bryan Anh Nguyen: Not clinical | – |
| 1391. Yagi 2018. Phenethyl isothiocyanate activates leptin signaling and decreases food intake PMID 30383868 | Grace Kim: No saffron | Bryan Anh Nguyen: No saffron | – |
| 1392. Ting 2018. Targeting Cellular Stress Mechanisms and Metabolic Homeostasis by Chinese Herbal Drugs for Neuroprotection PMID 29382106 | Grace Kim: Not clinical | Bryan Anh Nguyen: No saffron | – |
| 1393. Hussein 2017. Ginger and Propolis Exert Neuroprotective Effects against Monosodium Glutamate-Induced Neurotoxicity in Rats PMID 29117134 | Grace Kim: No saffron | Bryan Anh Nguyen: No saffron | – |
| 1394. Hashem-Dabaghian 2018. A systematic review on the cardiovascular pharmacology of Emblica officinalis Gaertn PMID 30386531 | Grace Kim: No saffron | Bryan Anh Nguyen: No saffron | – |
| 1395. Zari 2018. Effects of Caralluma russeliana stem extract on some physiological parameters in streptozotocin-induced diabetic male rats PMID 30349340 | Grace Kim: No saffron | Bryan Anh Nguyen: No saffron | – |
| 1396. Yao 2018. Effects of Acute Cold Stress on Liver O-GlcNAcylation and Glycometabolism in Mice PMID 30231545 | Grace Kim: No saffron | Bryan Anh Nguyen: No saffron | – |
| 1397. Shahzad 2018. Activated protein C reverses epigenetically sustained p66(Shc) expression in plaque-associated macrophages in diabetes PMID 30271984 | Grace Kim: No saffron | Bryan Anh Nguyen: No saffron | – |
| 1398. Pifferi 2018. Caloric restriction increases lifespan but affects brain integrity in grey mouse lemur primates PMID 30271916 | Grace Kim: No saffron | Bryan Anh Nguyen: No saffron | – |
| 1399. Author not recorded 2018. Abstracts PMC6153547 | Grace Kim: No saffron | Bryan Anh Nguyen: No saffron | – |
| 1400. Daneshi-Maskooni 2018. Green cardamom increases Sirtuin-1 and reduces inflammation in overweight or obese patients with non-alcoholic fatty liver disease: a double-blind randomized placebo-controlled clinical trial PMID 30263038 | Grace Kim: No saffron | Bryan Anh Nguyen: No saffron | – |

Table S2 continued

| Paper | Reviewer 1 | Reviewer 2 | Additional recorded reviews |
|---|---|---|---|
| 1401. Mala 2018. Safety and Efficacy of Ferula asafoetida in Functional Dyspepsia: A Randomized, Double-Blinded, Placebo-Controlled Study<br>PMID 30224930 | Grace Kim: No saffron | Bryan Anh Nguyen: No saffron | – |
| 1402. Lu 2018. Capillary Electrophoresis Separations of Glycans<br>PMID 29528644 | Grace Kim: No saffron | Bryan Anh Nguyen: No saffron | – |
| 1403. Mahmoudi 2018. Effects of Nigella sativa Extracts on the Lipid Profile and Uncoupling Protein-1 Gene Expression in Brown Adipose Tissue of Mice<br>PMID 30211134 | Grace Kim: No saffron | Bryan Anh Nguyen: No saffron | – |
| 1404. Gómez-Guzmán 2018. Potential Role of Seaweed Polyphenols in Cardiovascular-Associated Disorders<br>PMID 30060542 | Grace Kim: No saffron | Bryan Anh Nguyen: No saffron | – |
| 1405. Esmaeily 2018. Comparing Three Data Mining Algorithms for Identifying the Associated Risk Factors of Type 2 Diabetes<br>PMID 29374085 | Grace Kim: No saffron | Bryan Anh Nguyen: No saffron | – |
| 1406. Vahid 2018. Validation of a Dietary Inflammatory Index (DII) and Association with Risk of Gastric Cancer: a Case-Control Study<br>PMID 29936717 | Grace Kim: No saffron | Bryan Anh Nguyen: No saffron | – |
| 1407. Willis-Owen 2018. COPD is accompanied by co-ordinated transcriptional perturbation in the quadriceps affecting the mitochondria and extracellular matrix<br>PMID 30111857 | Grace Kim: No saffron | Bryan Anh Nguyen: No saffron | – |
| 1408. Just 2018. The gut microbiota drives the impact of bile acids and fat source in diet on mouse metabolism<br>PMID 30071904 | Grace Kim: No saffron | Bryan Anh Nguyen: No saffron | – |
| 1409. Batarseh 2017. Crocus sativus Extract Tightens the Blood-Brain Barrier, Reduces Amyloid $\beta$ Load and Related Toxicity in 5XFAD Mice<br>PMID 28471166 | Grace Kim: Not clinical | Pranav Potluri: Intervention + outcome but no endpoint outcome | – |
| 1410. Hussain 2018. Microencapsulation and the Characterization of Polyherbal Formulation (PHF) Rich in Natural Polyphenolic Compounds<br>PMID 29958444 | Grace Kim: No saffron | Pranav Potluri: No saffron | – |
| 1411. Jiao 2018. DNA barcoding for the efficient and accurate identification of medicinal polygonati rhizoma in China<br>PMID 30021015 | Grace Kim: No saffron | Pranav Potluri: No saffron | – |

Table S2 continued

| Paper | Reviewer 1 | Reviewer 2 | Additional recorded reviews |
|---|---|---|---|
| 1412. DeFilipps 2018. The medicinal plants of Myanmar PMID 30002597 | Grace Kim: No saffron | Pranav Potluri: No saffron | – |
| 1413. Rabiei 2018. Effects of a hydroalcoholic extract of Juglans regia (walnut) leaves on blood glucose and major cardiovascular risk factors in type 2 diabetic patients: a double-blind, placebo-controlled clinical trial PMID 29973195 | Grace Kim: No saffron | Pranav Potluri: No saffron | – |
| 1414. Maiese 2018. Novel Treatment Strategies for the Nervous System: Circadian Clock Genes, Non-coding RNAs, and Forkhead Transcription Factors PMID 29557749 | Grace Kim: No saffron | Pranav Potluri: No saffron | – |
| 1415. Athari Nik Azm 2017. Probiotics improve insulin resistance status in an experimental model of Alzheimer's disease PMID 29951404 | Grace Kim: No saffron | Pranav Potluri: No saffron | – |
| 1416. de Bem 2018. Antidiabetic effect of Euterpe oleracea Mart. (açaí) extract and exercise training on high-fat diet and streptozotocin-induced diabetic rats: A positive interaction PMID 29920546 | Grace Kim: No saffron | Pranav Potluri: No saffron | – |
| 1417. Sanati 2018. A review of the effects of Capsicum annuum L. and its constituent, capsaicin, in metabolic syndrome PMID 29922422 | Grace Kim: No saffron | Pranav Potluri: No saffron | – |
| 1418. Tastekin 2018. Therapeutic Potential of Pterostilbene and Resveratrol on Biomechanic, Biochemical, and Histological Parameters in Streptozotocin-Induced Diabetic Rats PMID 29887910 | Grace Kim: No saffron | Pranav Potluri: No saffron | – |
| 1419. Amini 2018. Selenium, Vitamin C and N-Acetylcysteine do not Reduce the Risk of Acute Kidney Injury after Off-Pump CABG: a Randomized Clinical Trial PMID 29898141 | Grace Kim: No saffron | Pranav Potluri: No saffron | – |
| 1420. Paudel 2018. Protein Tyrosine Phosphatase 1B Inhibition and Glucose Uptake Potentials of Mulberrofuran G, Albanol B, and Kuwanon G from Root Bark of Morus alba L. in Insulin-Resistant HepG2 Cells: An In Vitro and In Silico Study PMID 29786669 | Grace Kim: No saffron | Pranav Potluri: No saffron | – |
| 1421. El Hamrani 2018. Consumption of Alcopops During Brain Maturation Period: Higher Impact of Fructose Than Ethanol on Brain Metabolism PMID 29868598 | Grace Kim: No saffron | Pranav Potluri: No saffron | – |

Table S2 continued

| Paper | Reviewer 1 | Reviewer 2 | Additional recorded reviews |
|---|---|---|---|
| 1422. Wei 2018. Metabolomic Assessment of Acute Cholestatic Injuries Induced by Thioacetamide and by Bile Duct Ligation, and the Protective Effects of Huang-Lian-Jie-Du-Decoction PMID 29867467 | Grace Kim: No saffron | Pranav Potluri: No saffron | – |
| 1423. Kunnumakkara 2017. Curcumin, the golden nutraceutical: multitargeting for multiple chronic diseases PMID 27638428 | Grace Kim: No saffron | Pranav Potluri: No saffron | – |
| 1424. Denova-Gutiérrez 2018. Dietary Inflammatory Index and Type 2 Diabetes Mellitus in Adults: The Diabetes Mellitus Survey of Mexico City PMID 29561774 | Grace Kim: No saffron | Pranav Potluri: No saffron | – |
| 1425. Zheng 2018. Identification of Protein Kinase C Isoforms Involved in Type 1 Diabetic Encephalopathy in Mice PMID 29744369 | Grace Kim: No saffron | Pranav Potluri: No saffron | – |
| 1426. Wojcik 2018. Molecular Mechanisms Underlying Curcumin-Mediated Therapeutic Effects in Type 2 Diabetes and Cancer PMID 29743988 | Grace Kim: No saffron | Pranav Potluri: No saffron | – |
| 1427. Al-Attar 2018. Physiological study on the influence of some plant oils in rats exposed to a sublethal concentration of diazinon PMID 29740245 | Grace Kim: No saffron | Pranav Potluri: No saffron | – |
| 1428. Niwa-Kawakita 2017. PML is a ROS sensor activating p53 upon oxidative stress PMID 28931625 | Grace Kim: No saffron | Pranav Potluri: No saffron | – |
| 1429. Huang 2018. Combined intervention of swimming plus metformin ameliorates the insulin resistance and impaired lipid metabolism in murine gestational diabetes mellitus PMID 29677194 | Grace Kim: No saffron | Pranav Potluri: No saffron | – |
| 1430. Nader 2018. Characterization of increasing stages of invasiveness identifies stromal/cancer cell crosstalk in rat models of mesothelioma PMID 29662647 | Grace Kim: No saffron | Pranav Potluri: No saffron | – |
| 1431. Sellami 2018. Herbal medicine for sports: a review PMID 29568244 | Grace Kim: Not clinical | Pranav Potluri: No saffron | – |
| 1432. Shortt 2018. Systematic review of the effects of the intestinal microbiota on selected nutrients and non-nutrients PMID 29086061 | Grace Kim: No saffron | Pranav Potluri: No saffron | – |
| 1433. Sathyanarayana Rao 2018. Clinical Practice Guidelines for management of Sexual Disorders in Elderly PMID 29535473 | Grace Kim: No saffron | Pranav Potluri: No saffron | – |

Table S2 continued

| Paper | Reviewer 1 | Reviewer 2 | Additional recorded reviews |
|---|---|---|---|
| 1434. Wang 2018. Control of macrophage autophagy by miR-384-5p in the development of diabetic encephalopathy PMID 29511445 | Grace Kim: No saffron | Pranav Potluri: No saffron | – |
| 1435. Samarghandian 2017. Immunomodulatory and antioxidant effects of saffron aqueous extract (Crocus sativus L.) on streptozotocin-induced diabetes in rats PMID 28460761 | Grace Kim: Not clinical | Pranav Potluri: Not clinical | – |
| 1436. Catry 2017. Circumferential Esophageal Replacement by a Tissue-engineered Substitute Using Mesenchymal Stem Cells: An Experimental Study in Mini Pigs PMID 29390879 | Grace Kim: No saffron | Pranav Potluri: No saffron | – |
| 1437. Kermani 2017. The Efficacy of Crocin of Saffron (Crocus sativus L.) on the Components of Metabolic Syndrome: A Randomized Controlled Clinical Trial PMID 29417083 | Grace Kim: Candidate | Bryan Anh Nguyen: Candidate | – |
| 1438. Kunnumakkara 2018. Chronic diseases, inflammation, and spices: how are they linked? PMID 29370858 | Grace Kim: Not clinical | Bryan Anh Nguyen: Not clinical | – |
| 1439. Zimmer 2017. CAT-2003: A novel sterol regulatory element-binding protein inhibitor that reduces steatohepatitis, plasma lipids, and atherosclerosis in apolipoprotein E*3-Leiden mice PMID 29404461 | Grace Kim: No saffron | Bryan Anh Nguyen: Not clinical | – |
| 1440. Abedimanesh 2017. Saffron and crocin improved appetite, dietary intakes and body composition in patients with coronary artery disease PMID 29391933 | Grace Kim: Candidate | Bryan Anh Nguyen: Candidate | – |
| 1441. Nimrouzi 2018. Anorexia: Highlights in Traditional Persian medicine and conventional medicine PMID 29387569 | Grace Kim: No saffron | Bryan Anh Nguyen: Not clinical | – |
| 1442. Dickerson 2018. Cytokine-mediated changes in K(+) channel activity promotes an adaptive Ca(2+) response that sustains $\beta$-cell insulin secretion during inflammation PMID 29348619 | Grace Kim: No saffron | Bryan Anh Nguyen: No saffron | – |
| 1443. Na 2018. Dietary inflammatory index and its relationship with high-sensitivity C-reactive protein in Korean: data from the health examinee cohort PMID 29371758 | Grace Kim: No saffron | Bryan Anh Nguyen: No saffron | – |
| 1444. Chokka 2018. Assessment and management of sexual dysfunction in the context of depression PMID 29344340 | Grace Kim: No saffron | Bryan Anh Nguyen: Not clinical | – |

Table S2 continued

| Paper | Reviewer 1 | Reviewer 2 | Additional recorded reviews |
|---|---|---|---|
| 1445. Nurdiana 2017. Improvement of spatial learning and memory, cortical gyrification patterns and brain oxidative stress markers in diabetic rats treated with Ficus deltoidea leaf extract and vitexin<br>PMID 29322009 | Grace Kim: No saffron | Bryan Anh Nguyen: No saffron | – |
| 1446. Ranasinghe 2017. Evaluation of pharmacodynamic properties and safety of Cinnamomum zeylanicum (Ceylon cinnamon) in healthy adults: a phase I clinical trial<br>PMID 29282046 | Grace Kim: No saffron | Bryan Anh Nguyen: No saffron | – |
| 1447. Tsuda 2017. Corticotropin releasing hormone receptor 2 exacerbates chronic cardiac dysfunction<br>PMID 28550160 | Grace Kim: No saffron | Bryan Anh Nguyen: No saffron | – |
| 1448. Wilson 2017. The Role of Food Antioxidants, Benefits of Functional Foods, and Influence of Feeding Habits on the Health of the Older Person: An Overview<br>PMID 29143759 | Grace Kim: No saffron | Bryan Anh Nguyen: No saffron | – |
| 1449. Tang 2018. Lychee seed extract protects against neuronal injury and improves cognitive function in rats with type II diabetes mellitus with cognitive impairment<br>PMID 29138799 | Grace Kim: No saffron | Bryan Anh Nguyen: No saffron | – |
| 1450. Rodero 2017. Type I interferon-mediated autoinflammation due to DNase II deficiency<br>PMID 29259162 | Grace Kim: No saffron | Bryan Anh Nguyen: No saffron | – |
| 1451. Liu 2017. Herbal Medicine for the Treatment of Obesity: An Overview of Scientific Evidence from 2007 to 2017<br>PMID 29234439 | Grace Kim: Not clinical | Bryan Anh Nguyen: Not isolated | – |
| 1452. #N/A 2017. 11th European Headache Federation Congress jointly with 31st Congress of the Italian Society for the Study of Headaches<br>PMID 29189948 | Grace Kim: Not clinical | Bryan Anh Nguyen: No saffron | – |
| 1453. Vardin 2017. Effects of Cornus mas fruit hydro-methanolic extract on serum antioxidants, lipid profile, and hematologic parameters following cisplatin-induced changes in rats<br>PMID 29204179 | Grace Kim: No saffron | Bryan Anh Nguyen: No saffron | – |
| 1454. Russo 2017. Ketogal: A Derivative Ketorolac Molecule with Minor Ulcerogenic and Renal Toxicity<br>PMID 29163153 | Grace Kim: No saffron | Bryan Anh Nguyen: No saffron | – |

Table S2 continued

| Paper | Reviewer 1 | Reviewer 2 | Additional recorded reviews |
|---|---|---|---|
| 1455. Jovanović 2017. Evaluation of the Antioxidant and Antiglycation Effects of Lactarius deterrimus and Castanea sativa Extracts on Hepatorenal Injury in Streptozotocin-Induced Diabetic Rats<br>PMID 29163175 | Grace Kim: No saffron | Bryan Anh Nguyen: No saffron | – |
| 1456. Cozzolino 2017. Urinary volatile organic compounds in overweight compared to normal-weight children: results from the Italian I.Family cohort<br>PMID 29142292 | Grace Kim: No saffron | Bryan Anh Nguyen: No saffron | – |
| 1457. Abdali 2015. How effective are antioxidant supplements in obesity and diabetes?<br>PMID 25791371 | Grace Kim: No saffron | Bryan Anh Nguyen: No saffron | – |
| 1458. Huang 2017. Elevation of PTPN1 promoter methylation is a significant risk factor of type 2 diabetes in the Chinese population<br>PMID 29042909 | Grace Kim: No saffron | Bryan Anh Nguyen: No saffron | – |
| 1459. Ghorbani 2017. Pharmacological properties of Salvia officinalis and its components<br>PMID 29034191 | Grace Kim: No saffron | Bryan Anh Nguyen: No saffron | – |
| 1460. Zeka 2017. Flavonoids and Their Metabolites: Prevention in Cardiovascular Diseases and Diabetes<br>PMID 32962323 | Grace Kim: No saffron | Bryan Anh Nguyen: No saffron | – |
| 1461. Yashin 2017. Antioxidant Activity of Spices and Their Impact on Human Health: A Review<br>PMID 28914764 | Grace Kim: Not clinical | Bryan Anh Nguyen: Not clinical | – |
| 1462. Nunes-Neto 2017. The Effect of Schinus terebinthifolius Raddi (Anacardiaceae) Bark Extract on Histamine-Induced Paw Edema and Ileum Smooth Muscle Contraction<br>PMID 28928787 | Grace Kim: No saffron | Bryan Anh Nguyen: No saffron | – |
| 1463. Griffiths 2016. Food Antioxidants and Their Anti-Inflammatory Properties: A Potential Role in Cardiovascular Diseases and Cancer Prevention<br>PMID 28933408 | Grace Kim: No saffron | Bryan Anh Nguyen: No saffron | – |
| 1464. Kumar 2017. Picrorhiza kurroa Enhances $\beta$-Cell Mass Proliferation and Insulin Secretion in Streptozotocin Evoked $\beta$-Cell Damage in Rats<br>PMID 28878669 | Grace Kim: No saffron | Bryan Anh Nguyen: No saffron | – |

Table S2 continued

| Paper | Reviewer 1 | Reviewer 2 | Additional recorded reviews |
|---|---|---|---|
| 1465. Christie 2017. A community-based motivational personalised lifestyle intervention to reduce BMI in obese adolescents: results from the Healthy Eating and Lifestyle Programme (HELP) randomised controlled trial PMID 28687677 | Grace Kim: No saffron | Bryan Anh Nguyen: No saffron | – |
| 1466. Ghorbanzadeh 2017. Cardioprotective Effect of Crocin Combined with Voluntary Exercise in Rat: Role of Mir-126 and Mir-210 in Heart Angiogenesis PMID 28678929 | Grace Kim: Not clinical | Bryan Anh Nguyen: Not clinical | – |
| 1467. Schoemaker 2017. A casein hydrolysate based formulation attenuates obesity and associated non-alcoholic fatty liver disease and atherosclerosis in LDLr-/-.Leiden mice PMID 28678821 | Grace Kim: No saffron | Bryan Anh Nguyen: No saffron | – |
| 1468. Aktar 2017. Phytochemistry and Pharmacological Studies of Citrus macroptera: A Medicinal Plant Review PMID 28740540 | Grace Kim: No saffron | Bryan Anh Nguyen: Not clinical | – |
| 1469. Jafari 2016. Evaluation the effect of 50 and 100 mg doses of Cuminum cyminum essential oil on glycemic indices, insulin resistance and serum inflammatory factors on patients with diabetes type II: A double-blind randomized placebo-controlled clinical trial PMID 28725629 | Grace Kim: No saffron | Bryan Anh Nguyen: No saffron | – |
| 1470. Marrocco 2017. Measurement and Clinical Significance of Biomarkers of Oxidative Stress in Humans PMID 28698768 | Grace Kim: No saffron | Bryan Anh Nguyen: No saffron | – |
| 1471. Ortiz 2017. World Congress Integrative Medicine & Health 2017: part three PMID #N/A | Grace Kim: No saffron | Bryan Anh Nguyen: No saffron | – |
| 1472. Ee 2017. World Congress Integrative Medicine & Health 2017: part two PMID #N/A | Grace Kim: No saffron | Bryan Anh Nguyen: No saffron | – |
| 1473. Cascella 2017. Role of Nigella sativa and Its Constituent Thymoquinone on Chemotherapy-Induced Nephrotoxicity: Evidences from Experimental Animal Studies PMID 28629150 | Grace Kim: No saffron | Bryan Anh Nguyen: No saffron | – |
| 1474. Chayarop 2017. Hypoglycaemic activity of Mathurameha, a Thai traditional herbal formula aqueous extract, and its effect on biochemical profiles of streptozotocin-nicotinamide-induced diabetic rats PMID 28662699 | Grace Kim: No saffron | Bryan Anh Nguyen: No saffron | – |

Table S2 continued

| Paper | Reviewer 1 | Reviewer 2 | Additional recorded reviews |
|---|---|---|---|
| 1475. Catarino 2017. Fucaceae: A Source of Bioactive Phlorotannins<br>PMID 28635652 | Grace Kim: No saffron | Bryan Anh Nguyen: No saffron | – |
| 1476. Sotoudeh Anvari 2016. Relationship between calculated total antioxidant status and atherosclerotic coronary artery disease<br>PMID 27488746 | Grace Kim: No saffron | Bryan Anh Nguyen: No saffron | – |
| 1477. Mohammadzadeh 2017. Berberis vulgaris and its constituent berberine as antidotes and protective agents against natural or chemical toxicities<br>PMID 28656089 | Grace Kim: No saffron | Bryan Anh Nguyen: No saffron | – |
| 1478. Zhao 2017. Spatholobus suberectus Exhibits Antidiabetic Activity In Vitro and In Vivo through Activation of AKT-AMPK Pathway<br>PMID 28607575 | Grace Kim: No saffron | Bryan Anh Nguyen: No saffron | – |
| 1479. Suchal 2017. Molecular Pathways Involved in the Amelioration of Myocardial Injury in Diabetic Rats by Kaempferol<br>PMID 28505121 | Grace Kim: No saffron | Bryan Anh Nguyen: No saffron | – |
| 1480. Zhang 2016. A novel protein tyrosine phosphatase 1B inhibitor with therapeutic potential for insulin resistance<br>PMID 26990621 | Grace Kim: No saffron | Bryan Anh Nguyen: No saffron | – |
| 1481. Konstantopoulos 2017. Metabolic effects of Crocus sativus and protective action against non-alcoholic fatty liver disease in diabetic rats<br>PMID 28529733 | Grace Kim: Not clinical | Bryan Anh Nguyen: Not clinical | – |
| 1482. Ahangarpour 2017. Pancreatic protective and hypoglycemic effects of Vitex agnus-castus L. fruit hydroalcoholic extract in D-galactose-induced aging mouse model<br>PMID 28515766 | Grace Kim: No saffron | Bryan Anh Nguyen: No saffron | – |
| 1483. Xiang 2017. Chinese Herbal Medicines Attenuate Acute Pancreatitis: Pharmacological Activities and Mechanisms<br>PMID 28487653 | Grace Kim: No saffron | Bryan Anh Nguyen: No saffron | – |
| 1484. Bhavnani 2016. Mobile technology and the digitization of healthcare<br>PMID 26873093 | Grace Kim: No saffron | Bryan Anh Nguyen: No saffron | – |
| 1485. Gillard 2017. Microbiota Is Involved in Post-resection Adaptation in Humans with Short Bowel Syndrome<br>PMID 28469580 | Grace Kim: No saffron | Bryan Anh Nguyen: No saffron | – |

Table S2 continued

| Paper | Reviewer 1 | Reviewer 2 | Additional recorded reviews |
|---|---|---|---|
| 1486. Duchesne 2017. Bovine and murine models highlight novel roles for SLC25A46 in mitochondrial dynamics and metabolism, with implications for human and animal health PMID 28376083 | Grace Kim: No saffron | Bryan Anh Nguyen: No saffron | – |
| 1487. Bodén 2017. Dietary inflammatory index and risk of first myocardial infarction; a prospective population-based study PMID 28376792 | Grace Kim: No saffron | Bryan Anh Nguyen: No saffron | – |
| 1488. Kure 2017. The Immunomodulatory Effects of Plant Extracts and Plant Secondary Metabolites on Chronic Neuroinflammation and Cognitive Aging: A Mechanistic and Empirical Review PMID 28344556 | Grace Kim: No saffron | Bryan Anh Nguyen: No saffron | – |
| 1489. Kwon 2017. Rosmarinic Acid Potentiates Pentobarbital-Induced Sleep Behaviors and Non-Rapid Eye Movement (NREM) Sleep through the Activation of GABA(A)-ergic Systems PMID 27469144 | Grace Kim: No saffron | Bryan Anh Nguyen: No saffron | – |
| 1490. Menezes 2017. Impact of Flavonols on Cardiometabolic Biomarkers: A Meta-Analysis of Randomized Controlled Human Trials to Explore the Role of Inter-Individual Variability PMID 28208791 | Grace Kim: No saffron | Bryan Anh Nguyen: No saffron | – |
| 1491. Li 2017. Therapeutic mechanism of Yīn-Chén-Hāo decoction in hepatic diseases PMID 28275293 | Grace Kim: No saffron | Bryan Anh Nguyen: Not isolated | – |
| 1492. Saldanha 2016. Culinary Spice Plants in Dietary Supplement Products and Tested in Clinical Trials PMID 26980817 | Grace Kim: No saffron | Bryan Anh Nguyen: Not clinical | – |
| 1493. Mirmosayyeb 2017. Possible Role of Common Spices as a Preventive and Therapeutic Agent for Alzheimer's Disease PMID 28250905 | Grace Kim: Not clinical | Bryan Anh Nguyen: Not isolated | – |
| 1494. Samarghandian 2017. Catechin Treatment Ameliorates Diabetes and Its Complications in Streptozotocin-Induced Diabetic Rats PMID 28228702 | Grace Kim: No saffron | Bryan Anh Nguyen: No saffron | – |
| 1495. Mulder 2017. The CCR2 Inhibitor Propagermanium Attenuates Diet-Induced Insulin Resistance, Adipose Tissue Inflammation and Non-Alcoholic Steatohepatitis PMID 28076416 | Grace Kim: No saffron | Bryan Anh Nguyen: No saffron | – |

Table S2 continued

| Paper | Reviewer 1 | Reviewer 2 | Additional recorded reviews |
|---|---|---|---|
| 1496. Dehghan 2016. Purslane (Portulaca oleracea) Seed Consumption And Aerobic Training Improves Biomarkers Associated with Atherosclerosis in Women with Type 2 Diabetes (T2D)<br>PMID 27917862 | Grace Kim: No saffron | Bryan Anh Nguyen: No saffron | – |
| 1497. Sayah 2017. Antioxidant Activity and Inhibitory Potential of Cistus salviifolius (L.) and Cistus monspeliensis (L.) Aerial Parts Extracts against Key Enzymes Linked to Hyperglycemia<br>PMID 28116307 | Grace Kim: No saffron | Bryan Anh Nguyen: No saffron | – |
| 1498. Tiwary 2017. Radical Scavenging Activities of Lagerstroemia speciosa (L.) Pers. Petal Extracts and its hepato-protection in CCl(4)-intoxicated mice<br>PMID 28100224 | Grace Kim: No saffron | Bryan Anh Nguyen: No saffron | – |
| 1499. Taylor 2017. The Effectiveness of Pharmacological and Non-Pharmacological Interventions for Improving Glycaemic Control in Adults with Severe Mental Illness: A Systematic Review and Meta-Analysis<br>PMID 28056018 | Grace Kim: Not clinical | Bryan Anh Nguyen: No saffron | – |
| 1500. Mollazadeh 2016. Cinnamon effects on metabolic syndrome: a review based on its mechanisms<br>PMID 28096957 | Grace Kim: No saffron | Bryan Anh Nguyen: No saffron | – |
| 1501. Farshid 2016. The effects of crocin, insulin and their co-administration on the heart function and pathology in streptozotocin-induced diabetic rats<br>PMID 28078246 | Grace Kim: Not clinical | Bryan Anh Nguyen: Not clinical | – |
| 1502. Chodari 2016. Testosterone and Voluntary Exercise, Alone or Together Increase Cardiac Activation of AKT and ERK1/2 in Diabetic Rats<br>PMID 28558078 | Grace Kim: No saffron | Bryan Anh Nguyen: No saffron | – |
| 1503. Andrade 2016. Exercise and Beta-Glucan Consumption (Saccharomyces cerevisiae) Improve the Metabolic Profile and Reduce the Atherogenic Index in Type 2 Diabetic Rats (HFD/STZ)<br>PMID 27999319 | Grace Kim: No saffron | Bryan Anh Nguyen: No saffron | – |
| 1504. Habibi 2016. Involvement of microRNA-133 and -29 in cardiac disturbances in diabetic ovariectomized rats<br>PMID 27917273 | Grace Kim: No saffron | Bryan Anh Nguyen: No saffron | – |

Table S2 continued

| Paper | Reviewer 1 | Reviewer 2 | Additional recorded reviews |
|---|---|---|---|
| 1505. Fontés 2015. The ΔF508 Mutation in the Cystic Fibrosis Transmembrane Conductance Regulator Is Associated With Progressive Insulin Resistance and Decreased Functional $\beta$-Cell Mass in Mice<br>PMID 26283735 | Grace Kim: No saffron | Bryan Anh Nguyen: No saffron | – |
| 1506. Block 2015. Designing a broad-spectrum integrative approach for cancer prevention and treatment<br>PMID 26590477 | Grace Kim: No saffron | Bryan Anh Nguyen: No saffron | – |
| 1507. Liu 2016. Catalpol ameliorates diabetic atherosclerosis in diabetic rabbits<br>PMID 27830011 | Grace Kim: No saffron | Bryan Anh Nguyen: No saffron | – |
| 1508. Zhang 2015. Oxidative stress response and Nrf2 signaling in aging<br>PMID 26066302 | Grace Kim: No saffron | Bryan Anh Nguyen: No saffron | – |
| 1509. Maughan 2016. Improved adipose tissue function with initiation of protease inhibitor-only ART<br>PMID 27516476 | Grace Kim: No saffron | Bryan Anh Nguyen: No saffron | – |
| 1510. Toop 2016. Fructose Beverage Consumption Induces a Metabolic Syndrome Phenotype in the Rat: A Systematic Review and Meta-Analysis<br>PMID 27657120 | Grace Kim: No saffron | Bryan Anh Nguyen: No saffron | – |
| 1511. Islam 2016. Dietary Phytochemicals: Natural Swords Combating Inflammation and Oxidation-Mediated Degenerative Diseases<br>PMID 27721914 | Grace Kim: Not clinical | Bryan Anh Nguyen: Not clinical | – |
| 1512. Xu 2016. Development of a model based on biochemical, real-time tissue elastography and ultrasound data for the staging of liver fibrosis and cirrhosis in patients with chronic hepatitis B<br>PMID 27573619 | Grace Kim: No saffron | Bryan Anh Nguyen: No saffron | – |
| 1513. Morrison 2016. Intervention with a caspase-1 inhibitor reduces obesity-associated hyperinsulinemia, non-alcoholic steatohepatitis and hepatic fibrosis in LDLR-/-.Leiden mice<br>PMID 27121255 | Grace Kim: No saffron | Bryan Anh Nguyen: No saffron | – |
| 1514. Guo 2016. Objectively measured sedentary time, physical activity and kidney function in people with recently diagnosed Type 2 diabetes: a prospective cohort analysis<br>PMID 26282583 | Grace Kim: No saffron | Bryan Anh Nguyen: No saffron | – |

Table S2 continued

| Paper | Reviewer 1 | Reviewer 2 | Additional recorded reviews |
|---|---|---|---|
| 1515. Liu 2016. Photoreceptor Cells Influence Retinal Vascular Degeneration in Mouse Models of Retinal Degeneration and Diabetes PMID 27548901 | Grace Kim: No saffron | Bryan Anh Nguyen: No saffron | – |
| 1516. Schneck 2016. Roux-En Y Gastric Bypass Results in Long-Term Remission of Hepatocyte Apoptosis and Hepatic Histological Features of Non-alcoholic Steatohepatitis PMID 27594839 | Grace Kim: No saffron | Bryan Anh Nguyen: No saffron | – |
| 1517. Willhite 2014. Systematic review of potential health risks posed by pharmaceutical, occupational and consumer exposures to metallic and nanoscale aluminum, aluminum oxides, aluminum hydroxide and its soluble salts PMID 25233067 | Grace Kim: No saffron | Bryan Anh Nguyen: No saffron | – |
| 1518. Temprano 2016. Redundant roles of the phosphatidate phosphatase family in triacylglycerol synthesis in human adipocytes PMID 27344312 | Grace Kim: No saffron | Bryan Anh Nguyen: No saffron | – |
| 1519. Arun 2016. Chronic restraint stress induces sperm acrosome reaction and changes in testicular tyrosine phosphorylated proteins in rats PMID 27525328 | Grace Kim: No saffron | Bryan Anh Nguyen: No saffron | – |
| 1520. Borrelli 2016. Molecular Approaches to Genetically Improve the Accumulation of Health-Promoting Secondary Metabolites in Staple Crops-A Case Study: The Lipoxygenase-B1 Genes and Regulation of the Carotenoid Content in Pasta Products PMID 27455242 | Grace Kim: No saffron | Bryan Anh Nguyen: No saffron | – |
| 1521. Shay 2016. Abstracts from the 3rd International Genomic Medicine Conference (3rd IGMC 2015) : Jeddah, Kingdom of Saudi Arabia. 30 November - 3 December 2015 PMID 27454254 | Grace Kim: No saffron | Bryan Anh Nguyen: No saffron | – |
| 1522. Gray 2016. Engagement, Retention, and Progression to Type 2 Diabetes: A Retrospective Analysis of the Cluster-Randomised "Let's Prevent Diabetes" Trial PMID 27404094 | Grace Kim: No saffron | Bryan Anh Nguyen: No saffron | – |
| 1523. Lee 2016. Effect of a fermented dietary supplement containing chromium and zinc on metabolic control in patients with type 2 diabetes: a randomized, placebo-controlled, double-blind cross-over study PMID 27343205 | Grace Kim: No saffron | Bryan Anh Nguyen: Not isolated | – |

Table S2 continued

| Paper | Reviewer 1 | Reviewer 2 | Additional recorded reviews |
|---|---|---|---|
| 1524. Nagata 2016. Total dosage of gardenia fruit used by patients with mesenteric phlebosclerosis<br>PMID 27402114 | Grace Kim: No saffron | Bryan Anh Nguyen: No saffron | – |
| 1525. Khorasany 2016. Therapeutic effects of saffron (<br>PMID 27403251 | Grace Kim: Not clinical | Bryan Anh Nguyen: Not clinical | – |
| 1526. Mazidi 2016. Simvastatin Treatment Does Not Affect Serum Vitamin D Concentrations in Patients with Dyslipidemia: A Randomized Double-blind Placebo-controlled Cross-over Trial<br>PMID 27330686 | Grace Kim: No saffron | Bryan Anh Nguyen: No saffron | – |
| 1527. Cui 2016. Polyketides from the Mangrove-Derived Endophytic Fungus Nectria sp. HN001 and Their $\alpha$-Glucosidase Inhibitory Activity<br>PMID 27136568 | Grace Kim: No saffron | Bryan Anh Nguyen: No saffron | – |
| 1528. Bonnefont-Rousselot 2016. Resveratrol and Cardiovascular Diseases<br>PMID 27144581 | Grace Kim: No saffron | Bryan Anh Nguyen: No saffron | – |
| 1529. Yang 2016. The combination of Artemisia princeps Pamp, Leonurus japonicas Houtt, and Gardenia jasminoides Ellis fruit attenuates the exacerbation of energy, lipid, and glucose by increasing hepatic PGC-1$\alpha$ expression in estrogen-deficient rats<br>PMID 27216600 | Grace Kim: No saffron | Bryan Anh Nguyen: No saffron | – |
| 1530. Xiao 2016. APT070 (mirococept), a membrane-localizing C3 convertase inhibitor, attenuates early human islet allograft damage in vitro and in vivo in a humanized mouse model<br>PMID 26565566 | Grace Kim: No saffron | Bryan Anh Nguyen: No saffron | – |
| 1531. Mulder 2016. Surgical removal of inflamed epididymal white adipose tissue attenuates the development of non-alcoholic steatohepatitis in obesity<br>PMID 26499443 | Grace Kim: No saffron | Bryan Anh Nguyen: No saffron | – |
| 1532. Butt 2013. Anti-oncogenic perspectives of spices/herbs: A comprehensive review<br>PMID 27092039 | Grace Kim: Not clinical | Bryan Anh Nguyen: Not clinical | – |
| 1533. Anwar 2016. Anti-Hypertensive Herbs and Their Mechanisms of Action: Part II<br>PMID 27014064 | Grace Kim: No saffron | Bryan Anh Nguyen: No saffron | – |

Table S2 continued

| Paper | Reviewer 1 | Reviewer 2 | Additional recorded reviews |
|---|---|---|---|
| 1534. Ranjbari 2016. In vivo and in vitro evaluation of the effects of Urtica dioica and swimming activity on diabetic factors and pancreatic beta cells<br>PMID 26980377 | Grace Kim: No saffron | Bryan Anh Nguyen: No saffron | – |
| 1535. #N/A #N/A. POSTER SESSION ABSTRACTS<br>PMID #N/A | Grace Kim: No saffron | Bryan Anh Nguyen: No saffron | – |
| 1536. Nasri 2015. Diabetes mellitus and renal failure: Prevention and management<br>PMID 26941817 | Grace Kim: No saffron | Bryan Anh Nguyen: No saffron | – |
| 1537. Velena 2016. 1,4-Dihydropyridine Derivatives: Dihydronicotinamide Analogues-Model Compounds Targeting Oxidative Stress<br>PMID 26881016 | Grace Kim: No saffron | Bryan Anh Nguyen: No saffron | – |
| 1538. Mollazadeh 2015. Razi's Al-Hawi and saffron (<br>PMID 26877844 | Grace Kim: Not clinical | Bryan Anh Nguyen: Not clinical | – |
| 1539. Dambha-Miller 2016. Patient-centred care, health behaviours and cardiovascular risk factor levels in people with recently diagnosed type 2 diabetes: 5-year follow-up of the ADDITION-Plus trial cohort<br>PMID 26739725 | Grace Kim: No saffron | Bryan Anh Nguyen: No saffron | – |
| 1540. Weiskirchen 2016. Hepatoprotective and Anti-fibrotic Agents: It's Time to Take the Next Step<br>PMID 26779021 | Grace Kim: No saffron | Bryan Anh Nguyen: No saffron | – |
| 1541. Kim 2016. Black Ginseng Extract Counteracts Streptozotocin-Induced Diabetes in Mice<br>PMID 26751692 | Grace Kim: No saffron | Bryan Anh Nguyen: No saffron | – |
| 1542. Mashmoul 2013. Saffron: A Natural Potent Antioxidant as a Promising Anti-Obesity Drug<br>PMID 26784466 | Grace Kim: Not clinical | Bryan Anh Nguyen: Not clinical | – |
| 1543. Lamb 2016. Prospective associations between sedentary time, physical activity, fitness and cardiometabolic risk factors in people with type 2 diabetes<br>PMID 26518682 | Grace Kim: No saffron | Bryan Anh Nguyen: No saffron | – |
| 1544. Yao 2015. Geniposide promotes beta-cell regeneration and survival through regulating $\beta$-catenin/TCF7L2 pathway<br>PMID 25950476 | Grace Kim: No saffron | Bryan Anh Nguyen: No saffron | – |
| 1545. Alkerwi 2014. No significant independent relationships with cardiometabolic biomarkers were detected in the Observation of Cardiovascular Risk Factors in Luxembourg study population<br>PMID 25190219 | Grace Kim: No saffron | Bryan Anh Nguyen: No saffron | – |

Table S2 continued

| Paper | Reviewer 1 | Reviewer 2 | Additional recorded reviews |
|---|---|---|---|
| 1546. Chen 2015. Acute and subchronic toxicity as well as evaluation of safety pharmacology of traditional Chinese medicine “Huhezi”<br>PMID 26550447 | Grace Kim: No saffron | Bryan Anh Nguyen: No saffron | – |
| 1547. Nasri 2015. Antioxidant plants and diabetes mellitus<br>PMID 26487879 | Grace Kim: No saffron | Bryan Anh Nguyen: No saffron | – |
| 1549. Hoffman 2015. Food Processing and the Mediterranean Diet<br>PMID 26393643 | Grace Kim: Not clinical | Bryan Anh Nguyen: No saffron | – |
| 1550. Morrison 2015. Replacement of Dietary Saturated Fat by PUFA-Rich Pumpkin Seed Oil Attenuates Non-Alcoholic Fatty Liver Disease and Atherosclerosis Development, with Additional Health Effects of Virgin over Refined Oil<br>PMID 26405765 | Grace Kim: No saffron | Bryan Anh Nguyen: No saffron | – |
| 1551. Babatunde 2015. Hepatoprotective and Pancreatoprotective Properties of the Ethanolic Extract of Nigerian Propolis<br>PMID 26401394 | Grace Kim: No saffron | Bryan Anh Nguyen: No saffron | – |
| 1552. Chen 2015. HMGB1 is activated in type 2 diabetes mellitus patients and in mesangial cells in response to high glucose<br>PMID 26261550 | Grace Kim: No saffron | Bryan Anh Nguyen: No saffron | – |
| 1553. Takitani 2015. Altered retinol status and expression of retinol-related proteins in streptozotocin-induced type 1 diabetic model rats<br>PMID 26060349 | Grace Kim: No saffron | Bryan Anh Nguyen: No saffron | – |
| 1554. Candeias 2015. Gut-brain connection: The neuroprotective effects of the anti-diabetic drug liraglutide<br>PMID 26131323 | Grace Kim: No saffron | Bryan Anh Nguyen: No saffron | – |
| 1555. Sun 2015. Magnolia officinalis Extract Contains Potent Inhibitors against PTP1B and Attenuates Hyperglycemia in db/db Mice<br>PMID 26064877 | Grace Kim: No saffron | Bryan Anh Nguyen: No saffron | – |
| 1556. Al-Attar 2015. Effect of grapeseed oil on diazinon-induced physiological and histopathological alterations in rats<br>PMID 25972749 | Grace Kim: No saffron | Bryan Anh Nguyen: No saffron | – |
| 1557. Lee 2015. Neurons in the hippocampal CA1 region, but not the dentate gyrus, are susceptible to oxidative stress in rats with streptozotocin-induced type 1 diabetes<br>PMID 25878595 | Grace Kim: No saffron | Bryan Anh Nguyen: No saffron | – |

Table S2 continued

| Paper | Reviewer 1 | Reviewer 2 | Additional recorded reviews |
|---|---|---|---|
| 1558. Shafiq 2014. Cardio-protective and anti-cancer therapeutic potential of<br>PMID 25859300 | Grace Kim: No saffron | Bryan Anh Nguyen: No saffron | – |
| 1559. Linjawi 2015. Evaluation of the protective effect of Nigella sativa extract and its primary active component thymoquinone against DMBA-induced breast cancer in female rats<br>PMID 25861310 | Grace Kim: No saffron | Bryan Anh Nguyen: No saffron | – |
| 1560. Farrokhfall 2015. Comparison of inducible nitric oxide synthase activity in pancreatic islets of young and aged rats<br>PMID 25810884 | Grace Kim: No saffron | Bryan Anh Nguyen: No saffron | – |
| 1561. Sharma 2015. Caulerpa lentillifera extract ameliorates insulin resistance and regulates glucose metabolism in C57BL/KsJ-db/db mice via PI3K/AKT signaling pathway in myocytes<br>PMID 25889508 | Grace Kim: No saffron | Bryan Anh Nguyen: No saffron | – |
| 1562. Rajaei 2015. Antihyperglycemic and antihyperlipidemic effects of hydroalcoholic extract of Securigera securidaca seeds in streptozotocin-induced diabetic rats<br>PMID 25709998 | Grace Kim: No saffron | Bryan Anh Nguyen: No saffron | – |
| 1563. Park 2014. Daumone fed late in life improves survival and reduces hepatic inflammation and fibrosis in mice<br>PMID 24796965 | Grace Kim: No saffron | Bryan Anh Nguyen: No saffron | – |
| 1564. Wang 2014. Exendin-4 decreases liver inflammation and atherosclerosis development simultaneously by reducing macrophage infiltration<br>PMID 24490861 | Grace Kim: No saffron | Bryan Anh Nguyen: No saffron | – |
| 1565. #N/A #N/A. Abstracts from ATTD 2014<br>PMID #N/A | Grace Kim: Not clinical | Bryan Anh Nguyen: No saffron | – |
| 1566. Chandra 2014. Consensus statement on management of dyslipidemia in Indian subjects<br>PMID 25595144 | Grace Kim: No saffron | Bryan Anh Nguyen: No saffron | – |
| 1567. Gulati 2014. Sugar intake, obesity, and diabetes in India<br>PMID 25533007 | Grace Kim: No saffron | Bryan Anh Nguyen: No saffron | – |
| 1568. Shin 2014. Effect of intake of gardenia fruits and combined exercise of middle-aged obese women on hormones regulating energy metabolism<br>PMID 25566438 | Grace Kim: Not clinical | Bryan Anh Nguyen: No saffron | – |

Table S2 continued

| Paper | Reviewer 1 | Reviewer 2 | Additional recorded reviews |
|---|---|---|---|
| 1569. Lecarpentier 2014. Circadian rhythms, Wnt/beta-catenin pathway and PPAR alpha/gamma profiles in diseases with primary or secondary cardiac dysfunction PMID 25414671 | Grace Kim: No saffron | Bryan Anh Nguyen: No saffron | – |
| 1570. Mohan 2014. In vitro protection of biological macromolecules against oxidative stress and in vivo toxicity evaluation of Acacia nilotica (L.) and ethyl gallate in rats PMID 25043389 | Grace Kim: No saffron | Bryan Anh Nguyen: No saffron | – |
| 1571. Bibak 2014. Effects of melatonin on biochemical factors and food and water consumption in diabetic rats PMID 25250287 | Grace Kim: No saffron | Bryan Anh Nguyen: No saffron | – |
| 1572. Regnault 2014. Impaired liver function in Xenopus tropicalis exposed to benzo[a]pyrene: transcriptomic and metabolic evidence PMID 25103525 | Grace Kim: No saffron | Bryan Anh Nguyen: No saffron | – |
| 1573. Zhao 2014. Antidiabetic and antioxidative effect of jiang tang xiao ke granule in high-fat diet and low-dose streptozotocin induced diabetic rats PMID 25089145 | Grace Kim: No saffron | Bryan Anh Nguyen: No saffron | – |
| 1574. Toczek 2014. Periaortic brown adipose tissue as a major determinant of [$^{18}$F]-fluorodeoxyglucose vascular uptake in atherosclerosis-prone, apoE-/- mice PMID 25054923 | Grace Kim: No saffron | Bryan Anh Nguyen: No saffron | – |
| 1575. Eid 2014. Lingonberry (Vaccinium vitis-idaea L.) Exhibits Antidiabetic Activities in a Mouse Model of Diet-Induced Obesity PMID 25013446 | Grace Kim: No saffron | Bryan Anh Nguyen: No saffron | – |
| 1576. Duivenvoorden 2014. A statin-loaded reconstituted high-density lipoprotein nanoparticle inhibits atherosclerotic plaque inflammation PMID 24445279 | Grace Kim: No saffron | Bryan Anh Nguyen: No saffron | – |
| 1577. Verschuren 2014. A systems biology approach to understand the pathophysiological mechanisms of cardiac pathological hypertrophy associated with rosiglitazone PMID 24938300 | Grace Kim: No saffron | Bryan Anh Nguyen: No saffron | – |
| 1578. Griffin 2014. Multiple behaviour change intervention and outcomes in recently diagnosed type 2 diabetes: the ADDITION-Plus randomised controlled trial PMID 24759957 | Grace Kim: No saffron | Bryan Anh Nguyen: No saffron | – |

Table S2 continued

| Paper | Reviewer 1 | Reviewer 2 | Additional recorded reviews |
|---|---|---|---|
| 1579. Yapca 2014. Bilateral ovarian ischemia/reperfusion injury and treatment options in rats with an induced model of diabetes<br>PMID 24904723 | Grace Kim: No saffron | Bryan Anh Nguyen: No saffron | – |
| 1580. Ziaee 2014. Saffron reduced toxic effects of its constituent, safranal, in acute and subacute toxicities in rats<br>PMID 24644432 | Grace Kim: Not clinical | Bryan Anh Nguyen: Not clinical | – |
| 1581. Nasrolahi 2013. Honey and metformin ameliorated diabetes-induced damages in testes of rat; correlation with hormonal changes<br>PMID 24639728 | Grace Kim: No saffron | Bryan Anh Nguyen: No saffron | – |
| 1582. Chhatre 2014. Phytopharmacological overview of Tribulus terrestris<br>PMID 24600195 | Grace Kim: No saffron | Bryan Anh Nguyen: No saffron | – |
| 1583. Posadzki 2013. Herb-drug interactions: an overview of systematic reviews<br>PMID 22670731 | Grace Kim: Not clinical | Bryan Anh Nguyen: Not clinical | – |
| 1584. Hosseinzadeh 2014. Proteomic screening of molecular targets of crocin<br>PMID 24393168 | Grace Kim: No saffron | Bryan Anh Nguyen: Not clinical | – |
| 1585. Hasani-Ranjbar 2013. A systematic review of anti-obesity medicinal plants - an update<br>PMID 23777875 | Grace Kim: No saffron | Bryan Anh Nguyen: Not clinical | – |
| 1586. Chen 2014. Peroxisome proliferator-activated receptor activating hypoglycemic effect of Gardenia jasminoides Ellis aqueous extract and improvement of insulin sensitivity in steroid induced insulin resistant rats<br>PMID 24438349 | Grace Kim: No saffron | Bryan Anh Nguyen: No saffron | – |
| 1587. Samoylenko 2013. Nutritional countermeasures targeting reactive oxygen species in cancer: from mechanisms to biomarkers and clinical evidence<br>PMID 23458328 | Grace Kim: No saffron | Bryan Anh Nguyen: No saffron | – |
| 1588. Cooper 2014. Association between objectively assessed sedentary time and physical activity with metabolic risk factors among people with recently diagnosed type 2 diabetes<br>PMID 24196189 | Grace Kim: No saffron | Bryan Anh Nguyen: No saffron | – |
| 1589. Khedoe 2013. The effect of PPE-induced emphysema and chronic LPS-induced pulmonary inflammation on atherosclerosis development in APOE*3-LEIDEN mice<br>PMID 24303000 | Grace Kim: No saffron | Bryan Anh Nguyen: No saffron | – |

Table S2 continued

| Paper | Reviewer 1 | Reviewer 2 | Additional recorded reviews |
|---|---|---|---|
| 1590. Kim 2012. Single-dose oral toxicity of fermented scutellariae radix extract in rats and dogs<br>PMID 24278619 | Grace Kim: No saffron | Bryan Anh Nguyen: No saffron | – |
| 1591. McBane 2013. Evaluation of a collagen-chitosan hydrogel for potential use as a pro-angiogenic site for islet transplantation<br>PMID 24204863 | Grace Kim: No saffron | Bryan Anh Nguyen: No saffron | – |
| 1592. Li 2013. Gardenia oil increases estradiol levels and bone material density by a mechanism associated with upregulation of COX-2 expression in an ovariectomized rat model<br>PMID 24137227 | Grace Kim: No saffron | Bryan Anh Nguyen: No saffron | – |
| 1593. Ergin 2013. Carbonyl stress in aging process: role of vitamins and phytochemicals as redox regulators<br>PMID 24124633 | Grace Kim: No saffron | Bryan Anh Nguyen: No saffron | – |
| 1594. Pallauf 2013. Nutrition and healthy ageing: calorie restriction or polyphenol-rich "MediterrAsian" diet?<br>PMID 24069505 | Grace Kim: No saffron | Bryan Anh Nguyen: No saffron | – |
| 1595. Jung 2012. Syzygium aromaticum ethanol extract reduces high-fat diet-induced obesity in mice through downregulation of adipogenic and lipogenic gene expression<br>PMID 23181109 | Grace Kim: No saffron | Bryan Anh Nguyen: No saffron | – |
| 1596. #N/A #N/A. #N/A<br>PMID #N/A | Grace Kim: No saffron | Bryan Anh Nguyen: No saffron | – |
| 1597. #N/A 2012. The 26th Annual Symposium of The Protein Society: Abstracts<br>PMID #N/A | Grace Kim: No saffron | Bryan Anh Nguyen: No saffron | – |
| 1598. Gonzalez 2013. Hepatic Steatosis, Carbohydrate Intake, and Food Quotient in Patients with NAFLD<br>PMID 23737773 | Grace Kim: No saffron | Bryan Anh Nguyen: No saffron | – |
| 1599. Auvinen 2013. Both transient and continuous corticosterone excess inhibit atherosclerotic plaque formation in APOE*3-leiden.CETP mice<br>PMID 23717502 | Grace Kim: No saffron | Bryan Anh Nguyen: No saffron | – |
| 1600. Tamaddonfard 2013. Crocin Improved Learning and Memory Impairments in Streptozotocin-Induced Diabetic Rats<br>PMID 23638297 | Grace Kim: Not clinical | Bryan Anh Nguyen: Not clinical | – |
| 1602. Rezaee 2013. Safranal: From an Aromatic Natural Product to a Rewarding Pharmacological Agent<br>PMID 23638289 | Grace Kim: Not clinical | Bryan Anh Nguyen: Not clinical | – |

Table S2 continued

| Paper | Reviewer 1 | Reviewer 2 | Additional recorded reviews |
|---|---|---|---|
| 1603. Radonjic 2013. Differential effects of drug interventions and dietary lifestyle in developing type 2 diabetes and complications: a systems biology analysis in LDLr-/- mice PMID 23457508 | Grace Kim: No saffron | Bryan Anh Nguyen: No saffron | – |
| 1604. Atalar 2012. The role of mediastinal adipose tissue 11$\beta$-hydroxysteroid d ehydrogenase type 1 and glucocorticoid expression in the development of coronary atherosclerosis in obese patients with ischemic heart disease PMID 23009206 | Grace Kim: No saffron | Bryan Anh Nguyen: No saffron | – |
| 1605. Nilius 2011. Tasty and healthy TR(i)Ps. The human quest for culinary pungency PMID 21979815 | Grace Kim: No saffron | Bryan Anh Nguyen: No saffron | – |
| 1606. Macciò 2012. Management of anemia of inflammation in the elderly PMID 23091709 | Grace Kim: No saffron | Bryan Anh Nguyen: No saffron | – |
| 1607. Salim 2010. Cancer chemopreventive potential of volatile oil from black cumin seeds, Nigella sativa L., in a rat multi-organ carcinogenesis bioassay PMID 22966405 | Grace Kim: No saffron | Bryan Anh Nguyen: No saffron | – |
| 1608. #N/A #N/A. Abstracts of 28th Annual Congress of Iranian Psychiatric Association PMID 22952544 | Grace Kim: Not clinical | Bryan Anh Nguyen: No saffron | – |
| 1609. Shojaii 2012. Review of Pharmacological Properties and Chemical Constituents of Pimpinella anisum PMID 22848853 | Grace Kim: No saffron | Bryan Anh Nguyen: No saffron | – |
| 1610. Kandpal 2012. Transcriptome analysis using next generation sequencing reveals molecular signatures of diabetic retinopathy and efficacy of candidate drugs PMID 22605924 | Grace Kim: No saffron | Bryan Anh Nguyen: No saffron | – |
| 1611. Patouraux 2012. The osteopontin level in liver, adipose tissue and serum is correlated with fibrosis in patients with alcoholic liver disease PMID 22530059 | Grace Kim: No saffron | Bryan Anh Nguyen: No saffron | – |
| 1612. Díaz 2012. Copy number variation affecting the Photoperiod-B1 and Vernalization-A1 genes is associated with altered flowering time in wheat (Triticum aestivum) PMID 22457747 | Grace Kim: No saffron | Bryan Anh Nguyen: No saffron | – |
| 1613. Bahmani 2012. Glycine therapy inhibits the progression of cataract in streptozotocin-induced diabetic rats PMID 22355255 | Grace Kim: No saffron | Bryan Anh Nguyen: No saffron | – |

Table S2 continued

| Paper | Reviewer 1 | Reviewer 2 | Additional recorded reviews |
|---|---|---|---|
| 1614. Christie 2011. Assessing the efficacy of the Healthy Eating and Lifestyle Programme (HELP) compared with enhanced standard care of the obese adolescent in the community: study protocol for a randomized controlled trial PMID 22088133 | Grace Kim: No saffron | Bryan Anh Nguyen: No saffron | – |
| 1615. Zaid 2012. Greco-arab and islamic herbal-derived anticancer modalities: from tradition to molecular mechanisms PMID 22203868 | Grace Kim: Not clinical | Bryan Anh Nguyen: Not clinical | – |
| 1616. Johri 2011. Cuminum cyminum and Carum carvi: An update PMID 22096320 | Grace Kim: No saffron | Bryan Anh Nguyen: Not clinical | – |
| 1617. Haddad 2011. NCX 1000 Alone or in Combination with Vitamin E Reverses Experimental Nonalcoholic Steatohepatitis in the Rat Similarly to UDCA PMID 22013536 | Grace Kim: No saffron | Bryan Anh Nguyen: No saffron | – |
| 1618. Aggarwal 2011. Identification of novel anti-inflammatory agents from Ayurvedic medicine for prevention of chronic diseases: "reverse pharmacology" and "bedside to bench" approach PMID 21561421 | Grace Kim: Not clinical | Bryan Anh Nguyen: Not clinical | – |
| 1619. Pasinetti 2011. Caloric intake, dietary lifestyles, macronutrient composition, and alzheimer’ disease dementia PMID 21808725 | Grace Kim: Not clinical | Bryan Anh Nguyen: No saffron | – |
| 1620. Haddad 2011. Antioxidant and hepatoprotective effects of silibinin in a rat model of nonalcoholic steatohepatitis PMID 19884114 | Grace Kim: No saffron | Bryan Anh Nguyen: No saffron | – |
| 1621. Dubchak 2010. New and improved strategies for the treatment of gout PMID 21694941 | Grace Kim: No saffron | Bryan Anh Nguyen: No saffron | – |
| 1622. Griffin 2011. Protocol for the ADDITION-Plus study: a randomised controlled trial of an individually-tailored behaviour change intervention among people with recently diagnosed type 2 diabetes under intensive UK general practice care PMID 21463520 | Grace Kim: No saffron | Bryan Anh Nguyen: No saffron | – |
| 1623. Wielinga 2011. Beneficial effects of alternate dietary regimen on liver inflammation, atherosclerosis and renal activation PMID 21483792 | Grace Kim: No saffron | Bryan Anh Nguyen: No saffron | – |

Table S2 continued

| Paper | Reviewer 1 | Reviewer 2 | Additional recorded reviews |
|---|---|---|---|
| 1624. Zeng 2010. Analysis of the adverse reactions induced by natural product-derived drugs<br>PMID 20233209 | Grace Kim: No saffron | Bryan Anh Nguyen: No saffron | – |
| 1625. Tucci 2010. The role of lipid and carbohydrate digestive enzyme inhibitors in the management of obesity: a review of current and emerging therapeutic agents<br>PMID 21437083 | Grace Kim: Not clinical | Bryan Anh Nguyen: Not clinical | – |
| 1626. Gezen-Ak 2011. The effects of vitamin D receptor silencing on the expression of LVSCC-A1C and LVSCC-A1D and the release of NGF in cortical neurons<br>PMID 21408608 | Grace Kim: No saffron | Bryan Anh Nguyen: Not clinical | – |
| 1627. Cavicchia 2009. A new dietary inflammatory index predicts interval changes in serum high-sensitivity C-reactive protein<br>PMID 19864399 | Grace Kim: No saffron | Bryan Anh Nguyen: No saffron | – |
| 1628. Lionis 2010. Bio-psychosocial determinants of cardiovascular disease in a rural population on Crete, Greece: formulating a hypothesis and designing the SPILI-III study<br>PMID 20937097 | Grace Kim: No saffron | Bryan Anh Nguyen: No saffron | – |
| 1629. de la Garza-Rodea 2010. Anomer-Equilibrated Streptozotocin Solution for the Induction of Experimental Diabetes in Mice (<br>PMID 20122315 | Grace Kim: No saffron | Bryan Anh Nguyen: No saffron | – |
| 1630. Picon 2010. Randomized clinical trial of a phytotherapic compound containing Pimpinella anisum, Foeniculum vulgare, Sambucus nigra, and Cassia augustifolia for chronic constipation<br>PMID 20433751 | Grace Kim: No saffron | Bryan Anh Nguyen: No saffron | – |
| 1631. van der Hoorn 2009. The dual PPARalpha/gamma agonist tesaglitazar blocks progression of pre-existing atherosclerosis in APOE*3Leiden.CETP transgenic mice<br>PMID 19220285 | Grace Kim: No saffron | Bryan Anh Nguyen: No saffron | – |
| 1632. Jain 2010. Biofield therapies: helpful or full of hype? A best evidence synthesis<br>PMID 19856109 | Grace Kim: No saffron | Bryan Anh Nguyen: No saffron | – |
| 1633. Kleemann 2010. Time-resolved and tissue-specific systems analysis of the pathogenesis of insulin resistance<br>PMID 20098690 | Grace Kim: No saffron | Bryan Anh Nguyen: No saffron | – |
| 1634. Kaefer 2008. The role of herbs and spices in cancer prevention<br>PMID 18499033 | Grace Kim: Not clinical | Bryan Anh Nguyen: No saffron | – |

Table S2 continued

| Paper | Reviewer 1 | Reviewer 2 | Additional recorded reviews |
|---|---|---|---|
| 1635. LEMON 1960. Hypercitricemia in human cancer. Factors concerned in pathogenesis and treatment PMID 13760761 | Grace Kim: No saffron | Bryan Anh Nguyen: No saffron | – |
| 1636. Bhardwaj 2008. Comparative redox status in alcoholic liver disease and nonalcoholic fatty liver disease PMID 19669305 | Grace Kim: No saffron | Bryan Anh Nguyen: No saffron | – |
| 1637. Sheng 2008. Crocetin improves the insulin resistance induced by high-fat diet in rats PMID 18469847 | Grace Kim: No saffron | Bryan Anh Nguyen: Not clinical | – |
| 1638. #N/A #N/A. Association Intelligence PMID #N/A | Grace Kim: No saffron | Bryan Anh Nguyen: No saffron | – |
| 1639. Fraser #N/A. Bacterial expression and membrane targeting of the rat complement regulator Crry: A new model anticomplement therapeutic PMID 12237472 | Grace Kim: No saffron | Bryan Anh Nguyen: No saffron | – |
| 1640. Kleemann 2007. Atherosclerosis and liver inflammation induced by increased dietary cholesterol intake: a combined transcriptomics and metabolomics analysis PMID 17892536 | Grace Kim: No saffron | Bryan Anh Nguyen: No saffron | – |
| 1641. Berry 1962. EFFECTS OF BACTERIAL ENDOTOXINS ON METABOLISM : V. THE HYPERREACTIVITY OF MICE INFECTED WITH MYCOBACTERIUM TUBERCULOSIS, STRAIN BCG PMID 19867219 | Grace Kim: No saffron | Bryan Anh Nguyen: No saffron | – |
| 1642. BAAR 1947. Dysontogenetic pituitary cysts (pituitary cachexia in childhood) PMID 20252742 | Grace Kim: No saffron | Bryan Anh Nguyen: No saffron | – |
| 1643. Vaidya 2007. Current status of herbal drugs in India: an overview PMID 18392106 | Grace Kim: Not clinical | Bryan Anh Nguyen: No saffron | – |
| 1644. Stephens 2002. Resolution of P-glycoprotein and non-P-glycoprotein effects on drug permeability using intestinal tissues from mdr1a (-/-) mice PMID 11959808 | Grace Kim: No saffron | Bryan Anh Nguyen: No saffron | – |
| 1645. Sullivan 1905. Synthetic Culture Media and the Biochemistry of bacterial Pigments PMID 19971689 | Grace Kim: No saffron | Bryan Anh Nguyen: No saffron | – |
| 1646. Gatfield 1968. Hyperpipecolatemia: A new metabolic disorder associated with neuropathy and hepatomegaly: A case study. PMID 5700850 | Grace Kim: No saffron | Bryan Anh Nguyen: No saffron | – |

Table S2 continued

| Paper | Reviewer 1 | Reviewer 2 | Additional recorded reviews |
|---|---|---|---|
| 1647. Calandra 1970. Hormonal and Metabolic Studies in Pheochromocytoma<br>PMID 5422431 | Grace Kim: No saffron | Bryan Anh Nguyen: No saffron | – |
| 1648. HogenEsch #N/A. A spontaneous mutation characterized by chronic proliferative dermatitis in C57BL mice.<br>PMID 8362989 | Grace Kim: No saffron | Bryan Anh Nguyen: No saffron | – |
| 1649. Seemayer 1986. Obliterative segmental sclerosis of pancreatic islets. A possible consequence of hypotensive shock in young BB rats.<br>PMID 3538891 | Grace Kim: No saffron | Bryan Anh Nguyen: No saffron | – |
| 1650. Piche 2002. Fatigue is associated with high circulating leptin levels in chronic hepatitis C<br>PMID 12171970 | Grace Kim: No saffron | Bryan Anh Nguyen: No saffron | – |
| 1651. Ekundayo 1995. Gastrin and the growth of the gastrointestinal tract<br>PMID 7883218 | Grace Kim: No saffron | Bryan Anh Nguyen: No saffron | – |
| 1652. Issa-Chergui 1988. Major histocompatibility complex gene product expression on pancreatic beta cells in acutely diabetic BB rats.<br>PMID 3276208 | Grace Kim: No saffron | Bryan Anh Nguyen: No saffron | – |
| 1653. ELLIOTT 1961. Whipple's Disease Associated with Benign Thymoma<br>PMID 13889946 | Grace Kim: No saffron | Bryan Anh Nguyen: No saffron | – |
| 1654. O'Connor 2006. Development of baroreflex function and hind limb vascular reactivity in the horse fetus<br>PMID 16469779 | Grace Kim: No saffron | Bryan Anh Nguyen: No saffron | – |
| 1655. #N/A #N/A. Opinion<br>PMID #N/A | Grace Kim: No saffron | Bryan Anh Nguyen: No saffron | – |
| 1656. AUGER 1952. Lipomelanotic Hyperplasia of Lymph Nodes<br>PMID 14905380 | Grace Kim: No saffron | Bryan Anh Nguyen: No saffron | – |
| 1657. Porter 1981. Origins and genetic nonvariability of the proteins which diffuse from maize pollen<br>PMID 7460885 | Grace Kim: No saffron | Bryan Anh Nguyen: No saffron | – |
| 1658. Giussani 2005. Development of cardiovascular function in the horse fetus<br>PMID 15790668 | Grace Kim: No saffron | Bryan Anh Nguyen: No saffron | – |

Table S2 continued

| Paper | Reviewer 1 | Reviewer 2 | Additional recorded reviews |
|---|---|---|---|
| 1659. #N/A #N/A. Proceedings of the Physiological Society, 16-17 December 1983, Royal Veterinary College Meeting: Communications<br>PMID #N/A | Grace Kim: No saffron | Bryan Anh Nguyen: No saffron | – |
| 1660. Jacobson 1977. COMMUNICATIONS<br>PMID 894544 | Grace Kim: No saffron | Bryan Anh Nguyen: No saffron | – |
| 1661. Green 1976. COMMUNICATIONS<br>PMID 789858 | Grace Kim: No saffron | Bryan Anh Nguyen: No saffron | – |
| 1662. Merrett 1986. Characterization of the binding of the anti-sickling compound, BW12C, to haemoglobin<br>PMID 3814079 | Grace Kim: No saffron | Bryan Anh Nguyen: No saffron | – |
| 1663. Farooqi 2000. Dominant and recessive inheritance of morbid obesity associated with melanocortin 4 receptor deficiency<br>PMID 10903343 | Grace Kim: No saffron | Bryan Anh Nguyen: No saffron | – |
| 1664. Chaikoff 1934. THE BLOOD LIPIDS IN THE POSTABSORPTIVE STATE AND AFTER THE INGESTION OF FAT IN NORMAL HUMAN SUBJECTS AND IN A CASE OF DISSEMINATED CUTANEOUS XANTHOMATA<br>PMID 16694194 | Grace Kim: No saffron | Bryan Anh Nguyen: No saffron | – |
| 1665. Thorner 1982. Somatotroph hyperplasia. Successful treatment of acromegaly by removal of a pancreatic islet tumor secreting a growth hormone-releasing factor<br>PMID 6290540 | Grace Kim: No saffron | Bryan Anh Nguyen: No saffron | – |
| 1666. Nedenskov-Sørensen 1991. Sampling efficiency in the diagnosis of Helicobacter pylori infection and chronic active gastritis<br>PMID 1890167 | Grace Kim: No saffron | Bryan Anh Nguyen: No saffron | – |
| 1667. Kampa 2002. A new automated method for the determination of the Total Antioxidant Capacity (TAC) of human plasma, based on the crocin bleaching assay<br>PMID 12197944 | Grace Kim: Not clinical | Bryan Anh Nguyen: No saffron | – |
| 1668. Thomas-Vaslin 1997. Abnormal T cell selection on nod thymic epithelium is sufficient to induce autoimmune manifestations in C57BL/6 athymic nude mice<br>PMID 9114036 | Grace Kim: No saffron | Bryan Anh Nguyen: No saffron | – |
